# Integrative Genomic, Transcriptomic, and Microbiome Profiles of Colon Cancer by Ancestry Provide Insights into Molecular Distinctions

**DOI:** 10.64898/2026.03.25.26349226

**Authors:** Melissa Kramer, Pascal Belleau, Sofia C. Tortora, Astrid Deschênes, Kyriaki Founta, Carino Gurjao, Brian Yueh, Sara Goodwin, Devin Gee, Santhilal Subhash, Mali Barbi, Charlie Chung, Kadir Ozler, Onur Eskiocak, Ben Izar, Heather Geiger, Timothy R. Chu, Zoe Goldstein, Lara Winkerton, Andrew Araneo, Richard L Whelan, David Rivadeneira, Sharon Fox, Arjun Kandel, Fatih Ozay, Desiree Joy Anne Talabong, Olalekan Lanipekun, Henry Talus, Jianying Zeng, Arvind Rishi, Nyasha Chambwe, Nicolas Robine, Jeff Boyd, Alex Krasnitz, Semir Beyaz, W. Richard McCombie, Laura A. Martello

## Abstract

Colorectal cancer (CRC) incidence, tumor biology, and clinical outcomes differ by patient ancestry, yet African ancestry (AFR) populations remain underrepresented in genomic and microbiome studies. Here, we comprehensively characterized genomic, transcriptomic and microbiome features of AFR and European ancestry (EUR) colon cancer patients residing in New York City and Long Island. While confirming known drivers from other large CRC studies, our AFR to EUR comparison of somatic variation also revealed a possible enrichment of functional KRAS variants in AFR tumors. Colon cancer genomes in patients in this study also exhibit distinct patterns of DNA copy number variation, correlating with consensus molecular subtypes. *Fusobacterium nucleatum-*positive tumors were enriched for co-occurring oral taxa, suggesting an organized oral microbial structure within the tumor microenvironment. Our findings highlight ancestry-associated differences in somatic mutation, copy number variation, and tumor microbiome composition, underscoring the urgent need to expand AFR representation in genomic studies to uncover population-specific determinants of CRC risk and to develop treatment strategies that reflect the full diversity of patients affected by this disease.

## Introduction

There is significant epidemiological evidence that the incidence, severity at diagnosis, and mortality of colorectal cancer (CRC) occur substantially above population-wide averages in African Americans (AAs) (American Cancer Society, 2023; American Cancer Society, 2025). The limited availability of data exploring the genomic, transcriptomic, and microbiome features of CRC in patients of African ancestry has impeded progress in addressing the health disparities in this population. While there are ongoing efforts to better characterize CRC in AAs, many of the larger studies have been performed with sequencing of specific gene panels rather than whole genomes or exomes. In fact, many of the genes included in these clinical panels were based on earlier cancer discovery studies that did not fully represent population diversity. Without comprehensive data, it is challenging to develop targeted interventions or treatment strategies that could effectively reduce these disparities and improve outcomes for these patients.

An important aspect that has been underutilized in studies is the use of genetic ancestry rather than relying solely on self-reported race. Self-reported race can be a poor proxy for genetic ancestry, as it does not account for the complex and varied genetic backgrounds within racial groups (Brown *et al*., 2022; Wang *et al*., 2024; Gouveia *et al*., 2025). AAs share diverse African ancestry from many disparate regions on the African continent as well as European, Native American, and other ancestry, making this population group ill-defined by the catch-all term “race” (Lewis *et al*., 2022), which may bias interpretations related to differences in biology or outcomes. Identifying driver mutations in populations with a high degree of background genetic variability (and within this, geographic biases) is a challenging task that needs to be addressed to more rigorously identify oncogenic events across genetic ancestries (M. S. Lawrence *et al*., 2013).

Here, we have performed genomic, transcriptomic, and microbiome analyses on colon cancer cohorts of individuals with AFR and EUR genetic ancestry, which provided insights into differences and similarities across the spectrum and frequency of cancer mutations, transcriptional differences, and contributions of gut microbiome to disease. Our multi-faceted assays of AFR colon cancer may be a first step in larger efforts to more comprehensively characterize the complex factors underlying CRC etiology and outcomes.

## Results

Our study is comprised of two colon adenocarcinoma (COAD) cohorts: the first included WGS and transcriptome samples from the Polyethnic-1000 (P-1000) consortium (Robine and Varmus, 2022), which focuses on the study of cancer in individuals of diverse ancestry, and the second cohort included retrospective FFPE samples from a I CSHL/Downstate/Northwell exome sequencing pilot project on racial disparity in cancer (see **Figure 1A** and sample characteristics in **Table S1**). For this study, the P-1000 and FFPE cohorts included those samples which had greater than or equal to 70% African (deemed group AFRg) or European (deemed group EURg) ancestral fraction determined by genetic ancestry analysis (Alexander and Lange, 2011). Of the 52 patients in the P-1000 cohort that fit these criteria, 81% of the patients were of African genetic ancestry and 19% were of European genetic ancestry (**Table 1**). No individuals within this cohort self-identified as Hispanic or Latino and 54% of this cohort was female (**Table 1 and Notes S1**).

**Figure 1.**
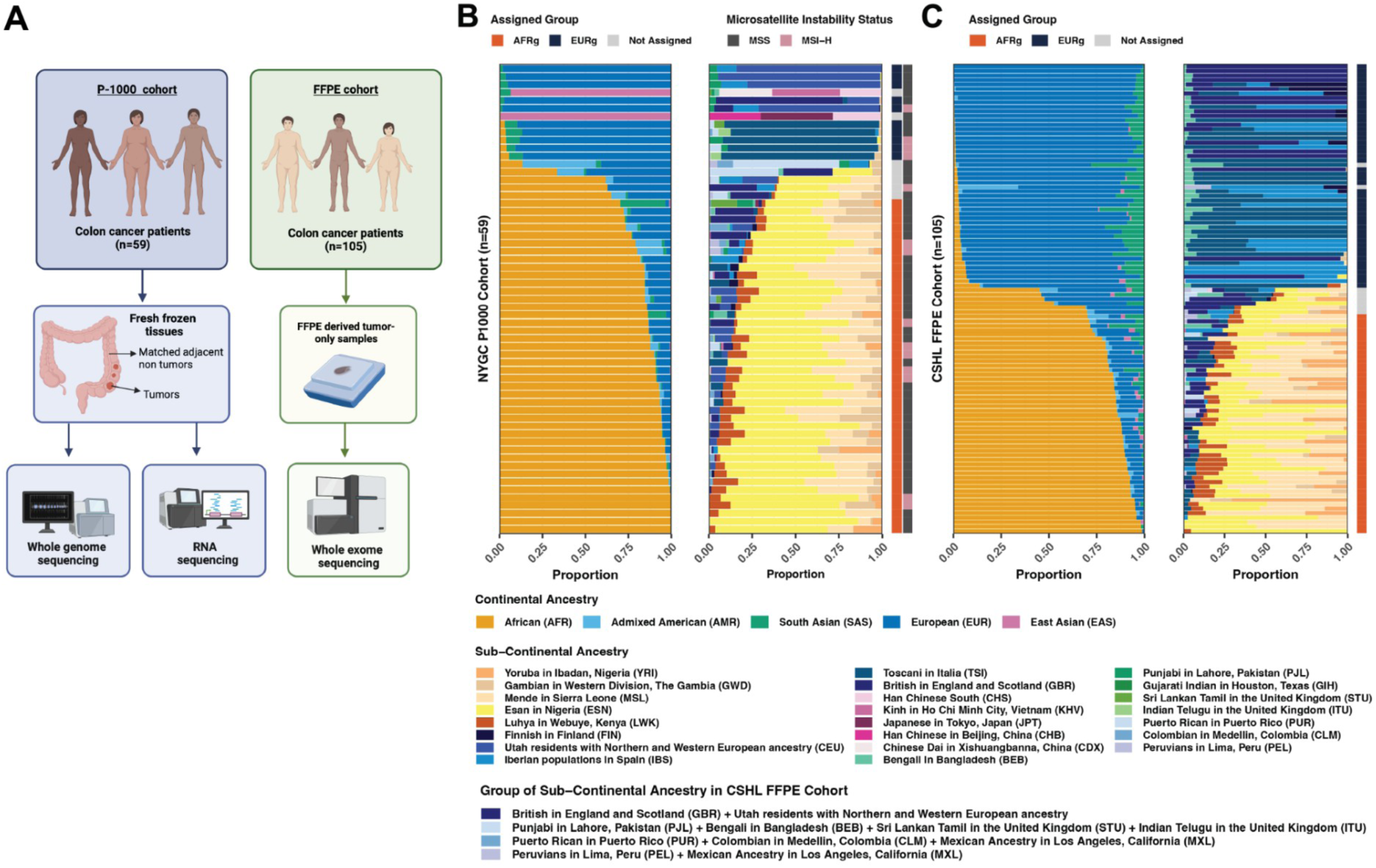
Genetic ancestry of the subjects in the study cohorts. **A)** Schematic of study designs for genetic ancestry analysis with two independent cohorts: the P-1000 cohort, with fresh frozen colon tumors and matched adjacent non tumors analyzed by whole genome sequencing (WGS) and RNA-sequencing (RNA-seq), and FFPE cohort, with FFPE-derived tumors profiled by whole exome sequencing (WES). The continental and subcontinental ancestry distribution of the subjects in the NYGC P-1000 Cohort **(B)** and the CSHL FFPE Cohort **(C)**. The profiles are respectively assigned in AFRg and EURg groups, when at least 70% of the continental ancestry fraction is African (AFR) or European (EUR). Otherwise, the profiles are not assigned to a group. In the NYGC P-1000 cohort, the information about the microsatellite instability (MSI) status of the tumor has been added.

**Table 1.** P-1000 sample characteristics and inclusion criteria.

| Characteristic |  | Ancestry |  | P-value |
| --- | --- | --- | --- | --- |
|  |  | African (N = 42) | European (N = 10) |  |
| Age at Diagnosis |  | 67 (46-86) | 79 (65-89) | 0.002 |
| Sex | Female | 22 (52%) | 6 (60%) | 0.7 |
|  | Male | 20 (48%) | 4 (40%) |  |
| Race | Black/African American | 42 (100%) | 0 (0%) |  |
|  | White | 0 (0%) | 10 (100%) |  |
| Ethnicity | Non-Hispanic | 42 (100%) | 10 (100%) |  |
| Histology | Adenocarcinoma NOS | 42 (100%) | 6 (60%) | <0.001 |
|  | Mucinous adenocarcinoma | 0 (0%) | 4 (40%) |  |
| Grade | G1 | 5 (12%) | 0 (0%) | 0.025 |
|  | G2 | 32 (76%) | 5 (50%) |  |
|  | G2-G3 | 1 (2.4%) | 0 (0%) |  |
|  | G3 | 4 (9.5%) | 5 (50%) |  |
| Stage | Stage I | 7 (17%) | 2 (20%) | 0.7 |
|  | Stage II | 22 (52%) | 4 (40%) |  |
|  | Stage III | 13 (31%) | 4 (40%) |  |
|  | Stage IV | 0 (0%) | 0 (0%) |  |
|  | Cannot be assessed | 0 (0%) | 0 (0%) |  |
| Specimen Collection Site | Ascending colon | 14 (33%) | 1 (10%) | 0.2 |
|  | Cecum | 3 (7.1%) | 1 (10%) |  |
|  | Colon, unspecified | 9 (21%) | 5 (50%) |  |
|  | Descending colon | 3 (7.1%) | 0 (0%) |  |
|  | Hepatic flexure | 1 (2.4%) | 0 (0%) |  |
|  | Rectosigmoid junction | 1 (2.4%) | 2 (20%) |  |
|  | Sigmoid colon | 9 (21%) | 1 (10%) |  |
|  | Transverse colon | 2 (4.8%) | 0 (0%) |  |
| Specimen Laterality | Left | 16 (38%) | 4 (40%) | >0.9 |
|  | Right | 26 (62%) | 6 (60%) |  |

The full P-1000 colon cancer cohort is comprised of a total of 59 individual samples, and data for all sequenced individuals will be shared as a public resource for further studies (see Resource Availability). We performed analysis of whole-genome sequencing (WGS) of fresh-frozen colon tumors and matched adjacent non-tumor from the P-1000 cohort, for a total of 42 patients assigned by the previously described genetic ancestry pipeline as AFRg and 10 individuals assigned as EURg. The tumors were sequenced to ∼80-100X coverage with matched adjacent non-tumor samples sequenced to ∼40-50X depth. In addition, we performed exome sequencing of FFPE derived colon tumor-only tissue (without matched adjacent non-tumor) samples for 52 individuals assigned by our genetic ancestry pipeline as AFRg and 51 individuals assigned as EURg (see sample characteristics **Table S1**). These samples were sequenced to an average depth of ∼150X coverage. These data were further compared with exome data derived from Dana Farber Cancer Institute (DFCI) colorectal cancer study (*dbGaP Study*, 2022) and The Cancer Genome Atlas (TCGA) (Cancer Genome Atlas Network, 2012) cohorts to increase the numbers of samples compared for each ancestry group.

### Genetic ancestry and self-reported identity

We performed genetic ancestry analysis using WGS data from matched adjacent non-tumor tissue from the P-1000 consortium and from FFPE derived tumor samples from the exome cohort (**Figure 1, Tables S4-7**). In both the P-1000 data set and the FFPE data set, most individuals’ self-reported racial identity aligned with their assigned ancestry group (**Figure S1A and B**). A subset of people (n=5 in the P-1000 cohort, and n=8 in the CSHL FFPE cohort) in the self-reported Black or African American (AA), or the White or European American (EA) racial groups had less than 70% ancestry fraction shared with the AFR and EUR 1000 Genomes superpopulations, thus they were not assigned to a single genetic ancestry group or included in downstream cohort comparison analyses. This threshold was selected in order to maximize sample inclusion to increase power with respect to the sample size. The level of EUR ancestry in the AFRg assigned patients is larger than the level of AFR ancestry in the EURg assigned patients (Wilcoxon rank test: p-value=0.02 for P-1000 cohort and p-value=4×10-10 for FFPE cohort) (**Figures S1C-D**). These results are similar to what is observed in other studies (Gouveia *et al*., 2025). When sub-continental ancestry was assessed most individuals who self-report as AA have alleles in common with the Esan in Nigeria (ESN) population, a finding also found in the TCGA African American dataset (**Figure S2**).

### Mutation analyses of somatic and germline variants

Mutational analysis (see **Methods**) was performed for the P-1000 WGS data on somatic and germline variants (**Figure 2**). We used the P-1000 dataset which had matched adjacent non-tumor for filtering as the initial discovery cohort and used the FFPE exome samples as a secondary validation cohort (**Figure S3**) due to less definitive determination of germline versus somatic variants from lack of matched adjacent non-tumor. We prioritized variants using an in-house pipeline to identify potential drivers/high impact variants (see **Methods**).

**Figure 2.**
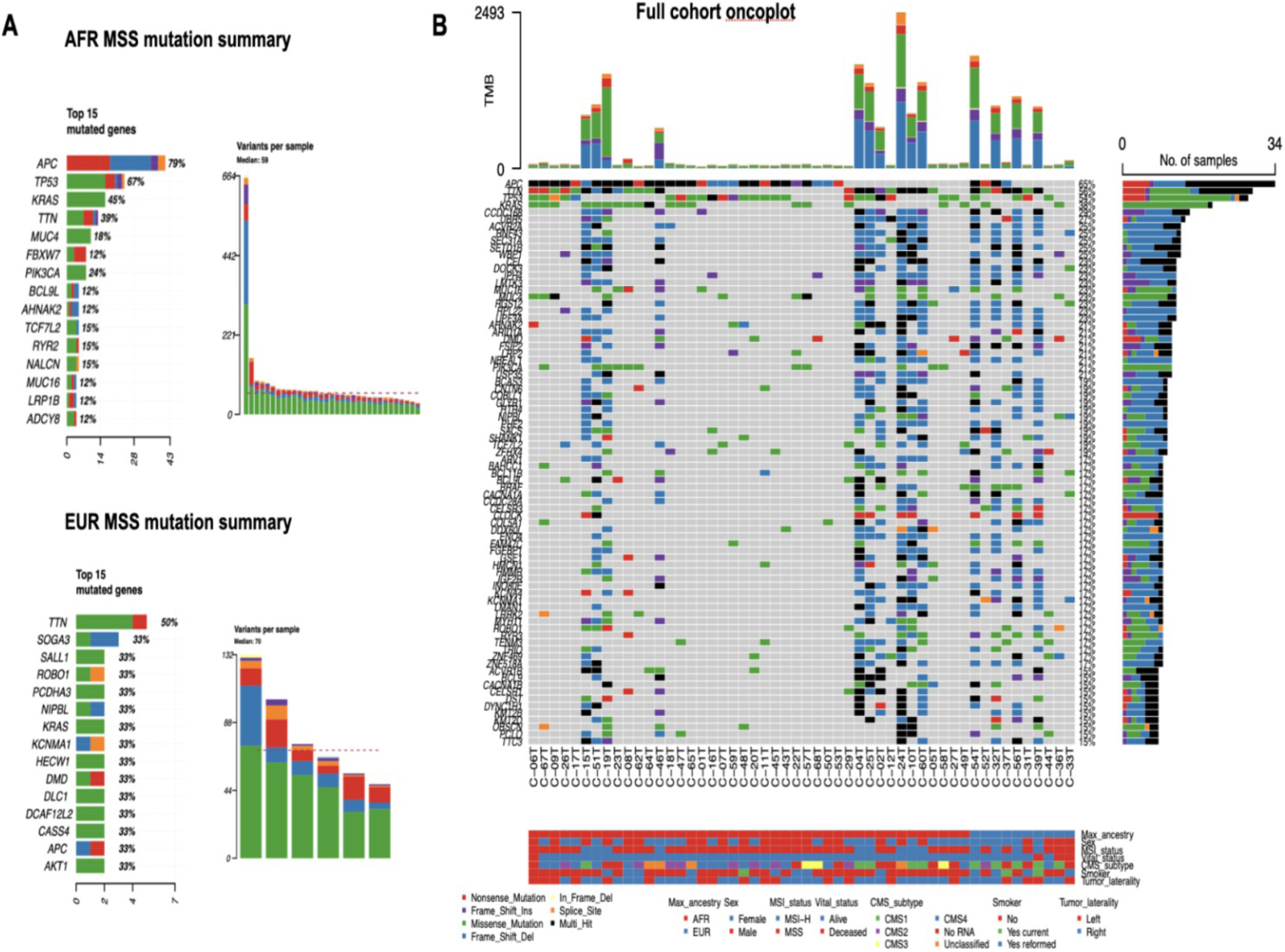
Somatic mutation overview of study cohort **A)** Top mutated genes and variants per sample for the AFR and EUR MSS groups used for cohort comparisons. **B)** Oncoplot of the entire cohort including both MSI-H and MSS samples showing frequency of top mutated genes across the sample set.

Our cohort included both microsatellite stable (MSS) tumors, and tumors with high microsatellite instability (MSI-H), with 9 (21%) of the AFRg and 4 (40%) of EURg samples being MSI-H (see **Note S2**). We note a similar range of tumor mutation burden (TMB) in our samples compared to the TCGA COAD cohorts (**Figure S4**). For the comparative analyses we stratified the MSI-H tumors which have differential mutational load (**Figure 2**).

Our data show mutations dominated by C->T transitions (**Figure S3**), indicative of 5mC deamination as has been reported for CRC (Hsu *et al*., 2017; Myer *et al*., 2022). As expected, we detected known CRC drivers from previous studies (He *et al*., 2022) as highly mutated genes in our cohorts, particularly *APC*, *KRAS*, *TP53*, *PIK3CA*, *FBXW7*, and *TCF7L2*. Importantly, we also detected mutations in genes noted as potential new drivers in recently published large studies of 1000-2000 individuals of predominantly European ancestry (Cornish *et al*., 2024; Nunes *et al*., 2024), such as *SETD5*, *FLCN*, *RPL22*, *TPTE*, *ROBO2*, *MBD6*. Several of these genes had not been previously identified to have a correlation with CRC or, in some cases, with any cancer. Their presence in our African ancestry-enriched cohort demonstrates the potential of diverse cohorts to refine the catalog of CRC drivers by identifying mutations that may be of low frequency in European ancestry patients and revealing population-associated variation that may inform CRC studies.

### Cohort enrichment analyses

We compared variants to identify any differing patterns between cohorts (see **Methods**), both at the variant frequency and gene network/pathway levels (**Figure 3** and **Figure S5A**). When the collected P-1000 WGS and FFPE exome data is compared to publicly available data from EUR individuals, other potential driver targets are identified. Although our cohort represents a small number of AFRg individuals, it is of similar size to the publicly available TCGA AFR colon cancer data (49 individuals).

**Figure 3.**
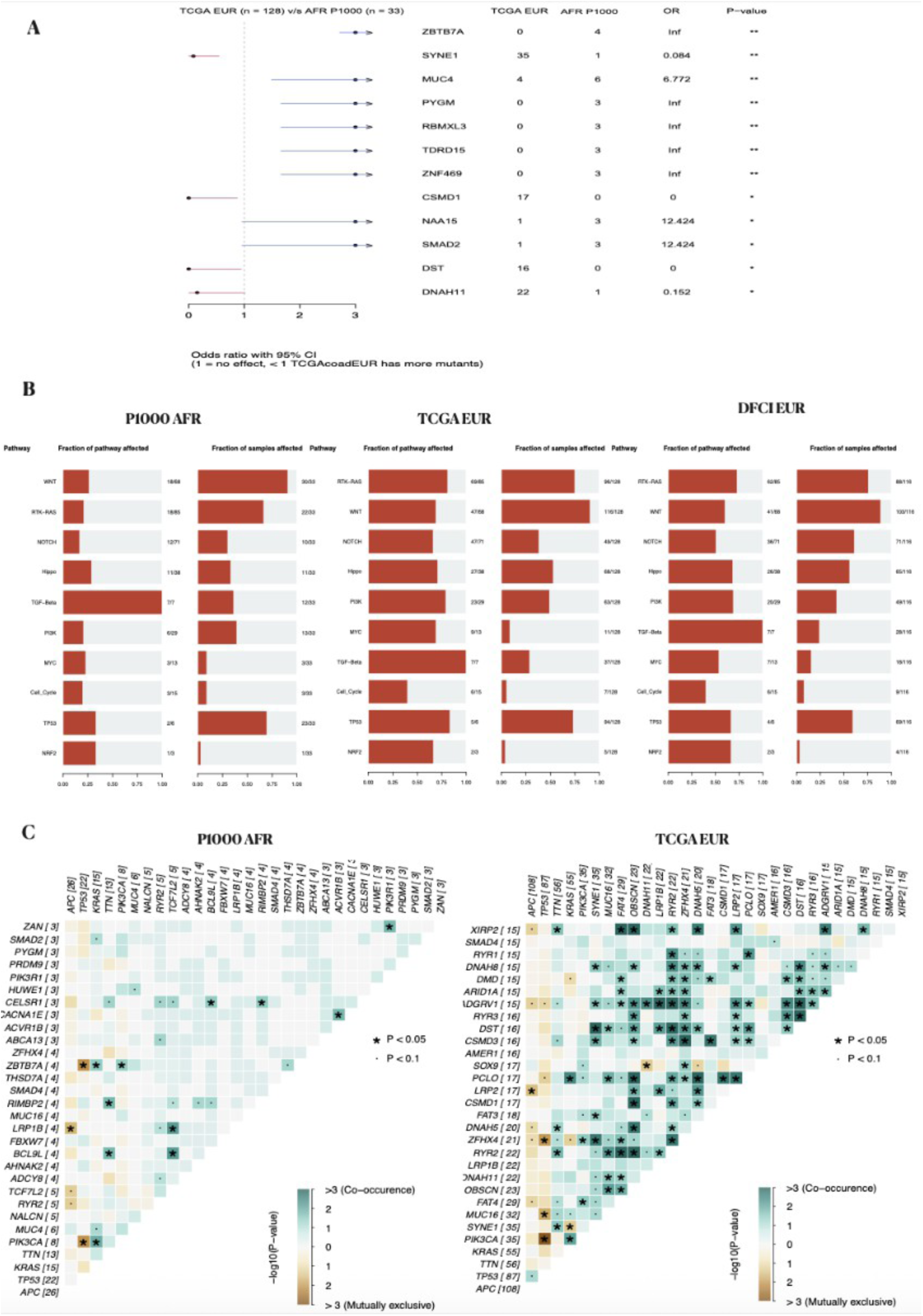
**A)** Forest plot with odds ratio of P-1000 AFRg versus TCGA EUR cohort comparison, with blue arrows denoting potentially enriched mutation frequency in the AFRg cohort. Listed p-value of less than 0.05 (*) or 0.01 (**) is unadjusted and used for ranking as no genes met the p<0.05 criteria after multiple testing correction. **B)** Oncogenic pathways showing number of genes and number of samples mutated in the P-1000 AFRg, TCGA EUR and DFCI EUR cohorts. **C)** Somatic interactions noting co-occurring and mutually exclusive mutations in the P-1000 AFRg and TCGA EUR cohorts.

In each dataset *KRAS* was identified as being more frequently mutated in AFRg than EURg (50% AFRg vs 34% EURg in the FFPE cohort, 45% AFRg vs 33% EURg in the P-1000 cohort, 45% P1000 AFRg vs 34% DFCI EURg, **Table S2**), a finding that has been observed in other studies (Staudacher *et al*., 2017; Myer *et al*., 2022; Innocenti *et al*., 2024). We do not, however, see a significant enrichment of *APC* (79% AFRg vs 84% TCGA-COAD EUR) or *PIK3CA* (24% AFRg vs 27% TCGA-COAD EUR) mutations nor a significant depletion of *BRAF* mutations (6% AFRg vs 8% TCGA-COAD EUR) in our P-1000 AFRg cohort compared to TCGA EUR, findings which have been seen in other studies (Staudacher *et al*., 2017; Myer *et al*., 2022; Innocenti *et al*., 2024; Lawler, Parlato and Warren Andersen, 2024). However, we note a potential increased AFRg frequency of another Wnt pathway regulator, *BCL9L* (**Table S2**), which has a complex dynamic of potentially oncogenic roles (Deka *et al*., 2010; Gay *et al*., 2019) and is noted as a CRC driver (Nunes *et al*., 2024). We also note reduced frequency of *CSMD1* in our P-1000 AFRg cohort (**Figure 3A**) compared to TCGA-COAD EUR (0/33 0% vs 17/128 13%) and DFCI EUR (15/116 13%). *CSMD1* is a known tumor suppressor in CRC (Zhang *et al*., 2014), however this finding may be due to sampling of our much smaller cohort, since it does not show depletion when compared in the AFRg FFPE study or the TCGA AFR cohort (**Table S2**).

We explored whether variants fell into differing oncogenic pathways for our P-1000 AFRg versus the larger TCGA EUR COAD (Kirk *et al*., 2016) and DFCI EUR cohorts. In most pathways, we see small differences likely derived from sampling the larger dataset. However, in the P-1000 AFRg MSS stable tumors, we see fewer affected genes, and fewer samples with mutations in the HIPPO pathway (11/33 33%) compared to TCGA EUR (68/128 53%) (**Figure 3B**). This also holds when compared to DFCI EUR (65/116 56%) (**Figure 3B**). The HIPPO pathway has been shown to have complex interactions with other pathways in CRC such as Wnt and TGF-beta (Mohammadpour *et al*., 2024). Other hits may include methylation changes which we did not assess, or hits in genes that otherwise interact with Hippo signaling.

We also assessed somatic interactions (**Figure 3C**), and we note that several interactions seem consistent between our P-1000 AFRg cohort and the TCGA-COAD EUR cohort. For example, both show co-occurrence of *PIK3CA* and *KRAS*, mutual exclusivity of *PIK3CA* and *TP53*, and mutual exclusivity of *APC* and *LRP1B*. However, we do note some potentially interesting interactions in our P-1000 AFRg cohort. *ZBTB7A* shows significant co-occurrence with *KRAS* and *PIK3CA* in our AFRg cohort but is mutually exclusive with *TP53*. *BCL9L* shows co-occurrence with both *TCF7L2*, another Wnt Pathway regulator, and with *CELSR1*, which has been shown to be upregulated in CRC (Liu *et al*., 2020).

We also note the importance of grouping tumors by microsatellite stability status when assessing cohort differences (see **Note S2**). **Figure 2B** oncoplot shows the MSI-H samples (9 AFRg and 4 EURg) harboring many more mutations than MSS samples, however, one sample deemed MSS in the AFRg cohort shows higher mutational load (**Figure 2A; Note S2**) exemplifying the complexity of classification of this tumor. Genes identified in the MSI-H samples in the P-1000 and FFPE data sets included *TCF7L2*, *ACVR2A*, *RNF43* and *BRAF*. These genes have been identified in previous studies including the recent UKBiobank CRC analysis (Cornish *et al*., 2024), as CRC drivers. When microsatellite instability was not accounted for in the analysis, several genes such as *SETD1B*, *SEC31A*, *ENO4* and *UPF3A* appeared to be more enriched in the AFR cohort compared to large EUR datasets such as TCGA and DFCI. In addition to ancestry based comparisons, when we compared mutations in left and right sided tumors (using all samples) (**Figure S7**), similar to previous studies, we noted an increased frequency of *SMAD4* mutations in left sided tumors (Ciepiela *et al*., 2024). However, several of the genes noted as more frequently mutated in right-sided tumors, such as *ACVR2A*, *ENO4* and *UPF3A*, are more prevalent in the MSI-H tumors.

### Germline variants

When applying the driver prioritization pipeline to select potentially deleterious germline alterations, *AHNAK2* showed increased mutation frequency in the AFRg cohort compared to EURg cohorts (37% vs 10% FFPE cohort, 75% vs 25% P-1000 cohort, 75% vs 62% for P-1000 AFRg vs DFCI EURg; **Table S2**). We note that in the FFPE exome data, since *AHNAK2* is also known to have somatic mutations in CRC, as seen in our cohort, the frequency may be affected by the inherent difficulty in completely stratifying somatic versus germline mutations without matched adjacent non-tumor tissue. High *AHNAK2* expression has been linked to poor survival in CRC and pancreatic cancer, and knockdown of *AHNAK2* inhibits tumor progression and metastasis of adenocarcinomas (Xu *et al*., 2022).

### Mutational signatures

Contribution of mutational signatures was assessed for the P-1000 WGS AFRg and EURg samples using deconstructSigs compared to the COSMIC v3 SBS, DBS and ID mutation types (**Figure S8A-C**). Significant proportions, defined as >=4.5% of total signature contribution, of SBS1 (deamination of 5mC clock-like signature) and SBS5 (clock-like) signatures, which are associated with aging, were noted (SBS1 40/42 AFRg and 9/10 EURg; SBS5 3/42 AFRg and 3/10 EURg). The bulk of the P-1000 cohort is over age 60, however, we did not see a clear increase in SBS1 proportion with age (**Figure S8D**). This is not unusual since SBS1 mutations are known to accumulate frequently in colon adenomas and are associated with replication stress (Crisafulli, 2024). Signatures associated with defective DNA mismatch repair, SBS6 (6/42 AFRg and 1/10 EURg), SBS15 (18/42 AFRg and 4/10 EURg), SBS26 (9/42 AFRg and 3/10 EURg), and SBS44 (8/42 AFRg and 5/10 EURg), DBS7 (20/42 AFRg and 7/10 EURg) and ID7 (9/42 AFRg and 5/10 EURg), were found across both AFRg and EURg samples. All MSI-H samples in the cohort showed high contributions of at least one of these signatures, along with contributions across one or more additional MMR signatures (**Figure S8E**). SBS10c (13/42 AFRg and 1/10 EURg) and SBS20 (7/42 AFRg and 3/10 EURg), defective *POLD1* signatures, were also noted. This correlates with previous reports regarding high incidence of MMR and polymerase deficiency in colorectal cancer (Pandey *et al*., 2019; Degasperi *et al*., 2022). The SBS18 signature (33/42 AFRg and 4/10 EURg) is associated with damage by reactive oxygen which can be incurred via pollutants, highly processed foods, tobacco or alcohol use, or gut inflammation, and contributes to carcinogenesis via inflammation and DNA damage, promoting proliferation and tumor growth (Catalano *et al*., 2025). SBS36 denotes defective base excision *MUTYH* mutations, and the one individual in the cohort with high levels of this signature does indeed carry a missense damaging *MUTYH* mutation.

Additionally, we note the presence of SBS93 in multiple samples (9/42 AFRg and 3/10 EURg), which has previously been detected in MSS CRC tumors and associated with gastric cancer (Crisafulli, 2024), and was noted to be significant in a study of over 2,000 colorectal cancers by Cornish et al. (Cornish *et al*., 2024). DBS13 was found in several samples (16/42 AFRg and 3/10 EURg), indicating homologous recombination deficiency which studies have shown may make CRC tumors sensitive to PARP inhibitors (Corti *et al*., 2024). We also note the DBS20 (27/42 AFRg and 5/10 EURg) doublet signature in aristolochic acid exposure, possibly indicating environmental toxin exposure. SBS5 and DBS2 (21/42 AFRg and 7/10 EURg) are also associated with tobacco and exogenous mutagens, however this did not seem to correlate with self-reported tobacco use so it may also indicate environmental exposure. The main indel signatures were ID1 (41/42 AFRg and 10/10 EURg) and ID2 (/42 AFRg and 10/10 EURg), indicative of slippage during DNA replication, and ID7 (9/42 AFRg and 5/10 EURg) pointing to defective DNA mismatch repair. Again, we note ID23 which associates with aristolochic acid exposure (42/42 AFRg and /10 EURg). There were also significant proportions of multiple signatures of unknown etiology (SBS8, SBS16, SBS17a and 17b, SBS39, SBS40a, SBS89, DBS15, DBS16, DBS17 DBS18, ID4, ID5, ID9 and ID14) across samples.

### Somatic Non-coding variants

Non-coding variants for our WGS P-1000 cohort were analyzed with MutEnricher (Soltis *et al*., 2020) to identify likely regulatory impact or transcription modifiers (see **Methods**) by overlap with ENCODE regions such as promoters, enhancers, DNAse hypersensitivity sites, histone marks and transcription factor binding sites (TFBS) (**Tables S13-S14**). Our AFRg showed enrichment of regulatory variants across samples in 2 genes, *NPRL3* and *ZFHX3*. Multiple samples in the AFRg cohort showed non-coding variants in a locus control region of *NPRL3*. Downregulation of NPRL3 has been potentially linked to cancer proliferation by reducing function of the GATOR1 complex, thereby overstimulating the mTORC pathway (Wang *et al*., 2022). *ZFHX3* has been identified as a tumor suppressor in multiple cancers (Hu *et al*., 2019) and the AFRg cohort showed noncoding variants in multiple samples which overlapped enhancers. Since these variants may impact regulation and expression of genes contributing to poorer outcomes in African Americans, we investigated the relationship between ancestry and expression of these genes within our cohort as well as in TCGA. A trend toward reduced *NPRL3* expression in AFRg was shown for both P-1000 AFRg and TCGA AFR compared to EUR (**Figure S9**), however the trend was less clear for *ZFHX3*. In addition, a subset of both the AFRg and EURg samples showed enrichment of noncoding variants in TFBS for SETDB1, MGA and MAX within the *APC* gene, and enrichment of variants within TFBS for RBFOX2, POLR2A/G and AGO2 within the *TP53* gene. Further exploration of these noncoding changes would be necessary in a larger cohort.

### Analysis of preneoplastic polyps

In order to further assess cancer development and the mutations underlying transformation in AFR and EUR colon cancer, we sequenced exomes of preneoplastic polyps from additional individuals and determined their genetic ancestry. In total, we sequenced 40 AFRg polyps and 37 EURg polyps. The polyps consisted of two subtypes: tubular adenomas (26 AFRg and 23 EURg) and serrated polyps (14 AFRg and 14 EURg). We identified potential drivers in these samples (see **Methods; Figure S5B**), and although these polyps are not patient-matched with the tumors, we assessed whether we could determine progression between the polyp and tumor samples by comparing mutation frequencies across genes (**Figure 4**).

**Figure 4.**
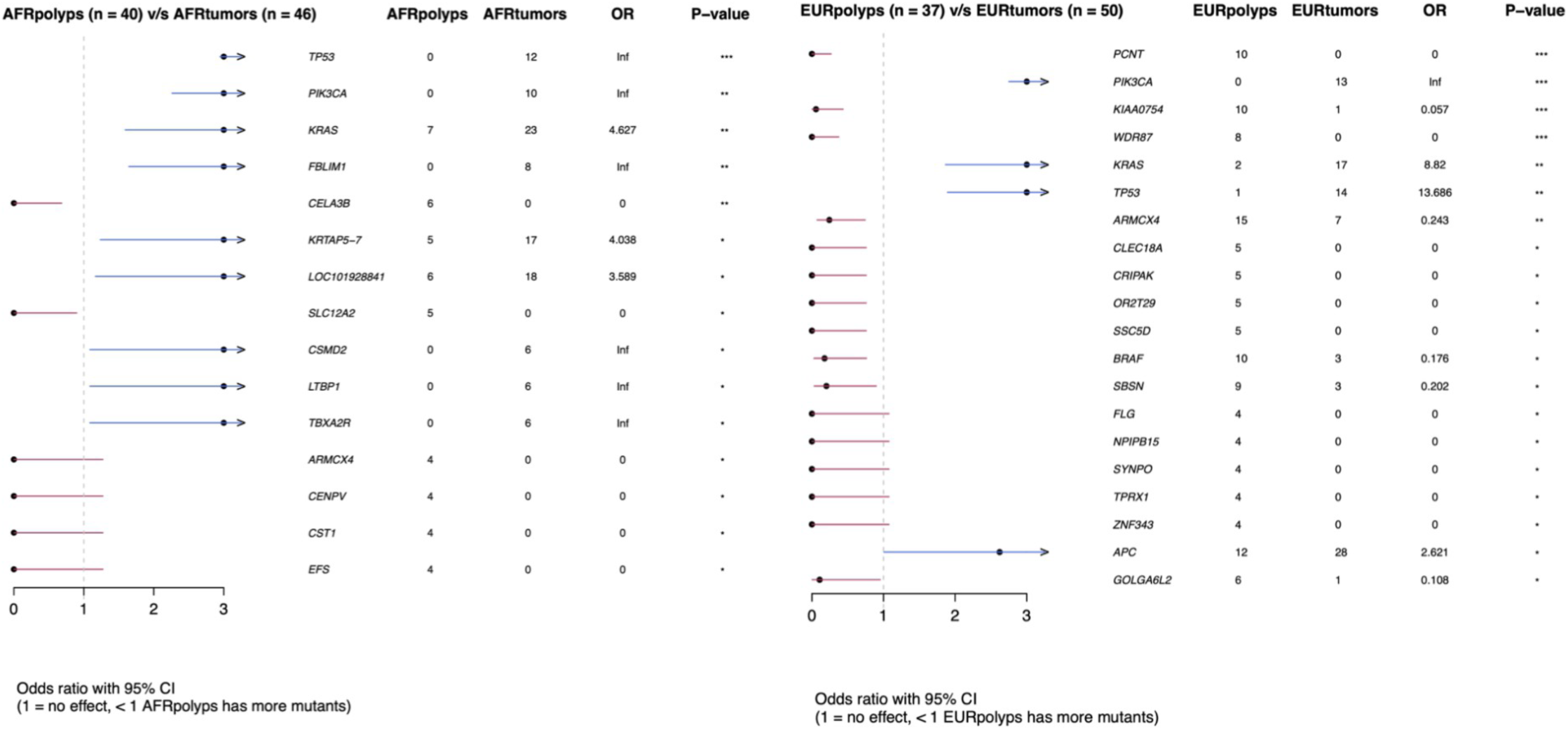
Forest plots of tumor vs polyp mutations for the AFR (left) and EUR (right) FFPE exome cohort, with blue arrows denoting potentially enriched gene mutation frequency in the tumors. Sample counts per cohort, odds ratio and unadjusted p-value less than 0.05 (*), 0.01 (**) or 0.001 (***) are shown.

We noted mutations in *TP53* (AFRg 0/40 0% polyps vs 12/46 26% tumors; EURg 1/37 3% polyps vs 14/50 28% tumors), *KRAS* (AFRg 7/40 18% polyps vs 23/46 50% tumors; EURg 2/37 5% polyps vs 17/50 34% tumors) and *PIK3CA* (AFRg 0/40 0% polyps vs 10/46 22% tumors; EURg 0/37 0% polyps vs 13/50 26% tumors) were more enriched in tumors compared to the precancerous adenomas indicating later drivers. However, the polyps did display *APC* mutations (AFRg 21/40 polyps vs 26/46 tumors; EURg 12/37 polyps vs 28/50 tumors), confirming *APC* as a “gatekeeper” gene and early driver (Klos and Dharmarajan, 2016). In particular, the serrated polyps in the EURg cohort showed multiple *BRAF* mutations (EURg 10/37 polyps vs 3/50 tumors; AFRg 4/40 polyps vs 2/46 tumors) as the early event rather than *APC* loss, as has been shown in other studies (Klos and Dharmarajan, 2016).

### Gene Expression Based Consensus Molecular Subtyping

We predicted consensus molecular subtypes (CMSs) (Guinney *et al*., 2015; Crisafulli, 2024) based on bulk gene expression profiling (**Figure 5A**; Figure S10; Figure S11A) to place our P-1000 cohort in context with previously published work supporting distinct molecular markers and different disease outcomes by subtype (Thanki *et al*., 2017) (n=43/52 after filtering; see **Methods**). 10 of our cohort samples were classified as CMS1, 11 as CMS2, 3 as CMS3 and 14 as CMS4, while 5 samples were unclassifiable (**Figure 5A**). Unclassifiable samples may represent an intermediate subtype, due to tumor heterogeneity that may be better defined by single cell analysis (Joanito *et al*., 2022). Nine (75%) of the 12 MSI-H samples in the cohort were assigned to CMS1, confirming the known association of that subtype with microsatellite instability (**Figure 5A**). Pathway activation scoring also confirmed the link between CMS1 and JAK-STAT, as well as MAPK pathway activity, while CMS4 samples showed hyperactivation of WNT and TGFB signaling when compared to the rest of the samples (**Figure 5B**).

**Figure 5.**
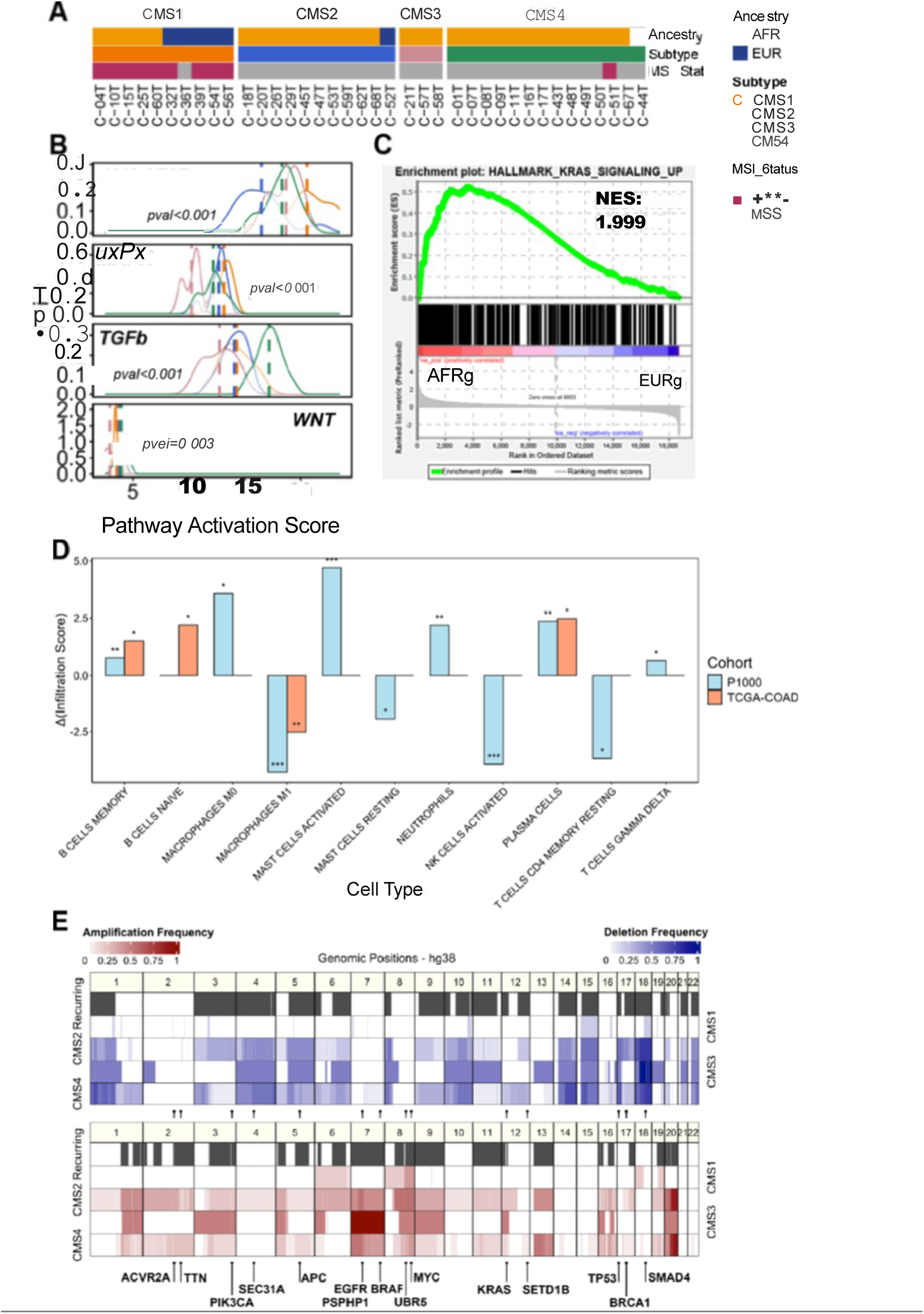
Gene Expression-Based Molecular Subtypes and Expression Signatures by Ancestry. **A)** Predicted CMS classifications (column annotations), as well as MSI status annotation for each sample in the cohort. Unclassifiable samples are not shown. **B)** Distribution of activation score per CMS for the JAK-STAT, MAPK, TGF-beta and WNT pathways. Average activation score per CMS is shown (dashed colored line), while significant pathway activation differences between different CMS tumors are also indicated (one-way ANOVA *p*-value). **C)** GSEA results showing upregulated *KRAS* signaling. Enrichment was determined using ranked results from the differential expression analysis. **D)** Differential immune cell infiltration between patients of African and European ancestry in the P-1000 and TCGA-COAD cohorts. The y-axis shows the difference in infiltration scores (Δ(Infiltration Score): µ(AFR) - µ(EUR)) for all cell types identified as significantly differentially infiltrating between the two population groups. Significance of differential infiltration is indicated above each bar (pval < 0.001 is indicated with “***”, pval < 0.01 is indicated with “**”, pval < 0.05 is indicated with “*”). **E)** Copy number prevalence (amplifications and deletions) for the AFRg WGS profiles (P-1000 cohort) correlate with CMS subtyping. Regions identified as having recurring copy number events (amplifications and deletions) are also shown in dark gray.

Molecular subtype was associated with genetic ancestry (Fisher’s exact test p-value 0.045). 5/7 (71%) EUR samples were classified as CMS1, whereas CMS2, CMS3 and CMS4 predominantly or exclusively consisted of AFR ancestry individuals (**Figure 5A, Figure S11A**). This relationship did not hold in the larger TCGA cohort (Fisher’s exact test p-value 0.9). However, proportional distribution of different ancestries within each CMS group in the TCGA-COAD cohort was significantly different than our cohort (**Figure S11B**). In our cohort, 25 samples fall into the more aggressive CMS2 and CMS4 subtypes, suggesting lower immune reactivity and likely treatment resistance.

### Gene Expression Signatures Associated with African Ancestry

We conducted a differential gene expression analysis comparing colon tumors from individuals of African and European ancestry in the P-1000 cohort, while adjusting for potential confounders including tumor purity, CMS subtype, age, and sex. This analysis identified three differentially expressed pseudogenes (Benjamini-Hochberg adjusted *p* < 0.05; effect size ≥ 0.1), *PSPHP1*, *RP11-208G20.2* and *RP11-208G20.3*, all of which were overexpressed in individuals of African ancestry (Figure S11C). Among them, *PSPHP1* exhibited the strongest association with ancestry. Notably, *PSPHP1* is a pseudogene of *PSPH* (phosphoserine phosphatase), previously reported to be overexpressed in colorectal (Jovov *et al*., 2012) and breast (Field *et al*., 2012) tumors from African-American patients. Similarly, overexpression of *PSPHL* (phosphoserine phosphatase homolog) has been observed in African-American prostate cancer patients (Wallace *et al*., 2008).

Expression levels of all three pseudogenes increased with higher African admixture in our cohort (**Figure S11C**). However, we observed substantial inter-patient variability at lower admixture levels (0–35% and 35–70%), suggesting that additional factors may modulate expression in this range. In particular, *PSPHP1* displayed a bimodal expression pattern among individuals with low African ancestry, indicating potential interaction between African admixture and some other clinical or molecular variable.

We next used gene set enrichment analysis (GSEA) to query the Hallmark gene sets related to ancestry (**Table S7**). Gene sets enriched in the African ancestry group include “KRAS signaling up” (200 genes upregulated by *KRAS* activation (Liberzon *et al*., 2015) (**Figure 5C**). In order to assess whether this finding correlated with the increased *KRAS* somatic mutation frequency in our AFRg cohort, we assessed the AFRg patients with microsatellite stable (MSS) tumors (**Figure S12A-B**). When the AFRg patients are split by subtyping, a significant difference is identified in the CMS4 subtype where *KRAS* mutation is associated with a higher “KRAS signaling up” score (4 KRAS mutated samples versus 8 *KRAS-*wild-type samples, Wilcoxon rank sum exact test p-value = 0.0040) (**Figure S12B**). This analysis is only conducted in the AFRg CMS2 and CMS4 subtypes (respectively 10 and 12 samples) due to a lack of *KRAS* wild-type samples in the other subtypes (respectively 0 and 3 samples in CMS1 and CMS3).

Previous studies have shown differential immune cell infiltration by ancestry in colon cancer (Paredes *et al*., 2020). Here, we used computational deconvolution approaches (Newman *et al*., 2015) on bulk transcriptomic sequencing generated on our cohort to infer the relative proportions of different cell types in our samples. We identified significant differences in immune cell type proportions between African and European tumors across various immune cell types. Suggestive differences were detected for memory B cells, Plasma cells, CD4 memory resting T cells, γδ T cells, activated Natural Killer cells, both M0 and M1 Macrophages, resting and activated Mast cells, and Neutrophils **Figure 5D**). Due to the small sample size, we then investigated the larger TCGA COAD cohort using the same analysis and found overlap in the enrichment of 3 cell types, including memory B cells, plasma cells, and M1 macrophages.

### Copy Number Analysis

We also used copy number variation (CNV) profiles of our AFRg set of the P-1000 cohort (41 profiles) to assess whether copy number could infer and stratify subtypes corresponding to the CMS typing, as well as to investigate whether subtypes differed by ancestry. Similarity between the profiles of the P-1000 cohort (only AFRg and EURg patients included) was assessed on the copy-number amplifications and deletions separately (see **Note S3 and Figures S14A-B**). For both analyses, the clustering generated a stratification by CMS subtyping rather than by ancestry. The CMS1 subtype is known to have low prevalence of somatic copy number alterations, while CMS2 to 4 display higher chromosomal instability (Guinney *et al*., 2015), as seen in the AFRg group in **Figure 5E**. This distinctive feature seems to be driving the copy-number similarity clustering.

We evaluated the presence of large regions with recurrent copy number events in the subset of AFRg of the P-1000 cohort with MSS tumors. A total of 103 regions (53 with recurring deletions and 50 with recurring amplifications) were identified (**Figure 5E**, **Table S11**). Significant difference in gene expression was confirmed for 92 of those regions when comparing expression in patients with the corresponding event versus without the event (Benjamini-Hochberg adjusted p < 0.05) (**Table S12**). Most of the regions (89 out of 103 regions) overlap with genes associated with cancer in the COSMIC Cancer Gene Census database (**Table S11**). Two regions overlap the *KRAS* gene: one region with recurring amplification events (chr12.p12.1−p11.21) and one region with recurring deletion events (chr12.p13.33−p11.1). Amplifications of chromosomes 8q and 20q have been associated with the transition from adenoma to carcinoma and cancer progression (Carvalho *et al*., 2009; Lee *et al*., 2015). We identified four amplified recurrent regions in chromosome 8q (regions c43 to c46 in **Table S11**). Those regions include multiple genes associated with cancer, such as *RAD21* and *MYC*. A total of eight amplified regions were identified in our cohort on chromosome 20q (regions c91, c93 to c99 in **Table S11**). A study of the TCGA CRC collection noted that the amplification of 7q increased expression of the potential oncogene *GTF2IRD1* (Nambara *et al*., 2020). This gene is included in the three amplified regions we identified in our cohort on chromosome 7q (regions c36 to c38 in **Table S11**). Two of those regions overlap the oncogene *BRAF*. We also found losses of 17p in CMS2, and losses of 18q in CMS2, CMS3, and CMS4. These losses had been identified as prevalent in a large proportion of CRCs by Nunes et al. (Nunes *et al*., 2024). In our cohort, the chr17:q12-q25.3 region has been identified as recurrently deleted (c77 in **Table S11**). This region overlaps with multiple genes associated with cancer, such as *BRCA1* and *ERBB2*. We also identified five deleted regions in chromosome 18q (regions c77 to c83 in **Table S11**).

The copy number variation signatures (COSMIC CN) were extracted for the AFRg and EURg patients in the P-1000 and TCGA COAD cohorts (**Figure S14A-B**). In both cohorts, the signatures of diploid (CN1) and tetraploid genomes (CN2) were the two most abundant signatures. The CN1 signature was detected in 25 of the 51 patients in the P-1000 cohort (49%) and in 131 of 215 patients in the TCGA COAD cohort (61%). The CN2 signature was detected in 26 of the 51 patients in the P-1000 cohort (51%) and in 104 of 215 patients in the TCGA COAD cohort (48%). The CN1 and CN2 signatures did not show any significant difference in frequency by ancestry (AFRg vs EURg comparison). However, we identified a significant association between the presence of the CN2 signature and the LOH status of mutated *TP53* in AFRg and EURg patients with MSS tumors (Fisher’s exact test, p-value=2.268e-05) (**Figure S14C**).

In the P-1000 cohort, the signature for chromosomal-scale LOH (CN13 signature) is detected at a higher frequency (14/51, 27%) than in the TCGA COAD cohort (9/215, 4%) (only AFRg and EURg patients retained, Fisher’s exact test, p-value=3.833e-06). No significant difference in frequency of CN13 was detected when comparing AFRg to EURg by ancestry in either cohort. However, we did identify a significant association between the presence of the CN13 signature and the deletion of the chr18:q12.3-q23 region (region c81 in Table S11) in the AFRg and EURg patients with MSS tumors (Fisher’s exact test, p-value=0.004) (**Figure S14C**). The region overlaps the *DCC*, *SMAD2* and *SMAD4* genes. The allelic loss of chromosome 18q has been associated with poorer survival in a subset of colorectal cancer patients (Jen *et al*., 1994).

### Microbiome Analysis

Microbiome community and diversity analyses were conducted through 16S rRNA gene sequencing on matched tumor and adjacent non-tumor samples from a subset of AFRg patients in the P-1000 cohort. Alpha diversity was assessed using observed amplicon sequence variants (ASVs) and the Shannon index. Tumor tissues exhibited significantly lower microbial diversity than non-tumor tissues (**Figure 6A**). Notably, right-sided tumors displayed reduced diversity and approximately half the observed ASVs relative to left-sided tumors (**Figure 6A**). Among the most abundant genera identified in right-sided tumors were *Streptococcus, Fusobacterium*, and *Pseudomonas*, whereas left-sided tumors showed increased relative abundances of *Prevotella, Lactobacillus*, and *Bacteroides* (**Table S8**). No significant differences were observed for colon tumor samples based on location or cancer stage for beta-diversity analysis (**Figures S17A–C**). Differential abundance analysis revealed oral-origin bacteria as statistically significant ASVs. *Treponema socranskii* and *Prevotella_7* were enriched in left-sided tumors, while *F. necrophorum* and *F. nucleatum* were more abundant in right-sided tumors. *Veillonella spp.* was enriched in non-tumor tissues (**Figure 6B**). Additionally, the phylum *Fusobacteriota* was exclusively detected in tumor tissues (**Figure 6C**). To explore bacterial co-occurrence patterns with *F. nucleatum* presence, we categorized tumors as *F. nucleatum* (Fn)-positive or -negative using qPCR. By grouping tumors based on their Fn-status, we found that Fn-positive tumors showed a 4-fold increase 14 in oral-origin genera, including *Fusobacterium* (10-fold, p>0.001), *Leptotrichia* (168-fold, p=0.201)*, Campylobacter* (78-fold, p=0.024*), Prevotella* (4-fold, p=0.001), *Treponema* (50-fold, p=0.0081), *and Porphyromonas* (10-fold, p=0.003) which accounted for 31.1% of the tumor microbiome vs. 7.8% in Fn-negative tumors (**Figure 6D**).

**Figure 6.**
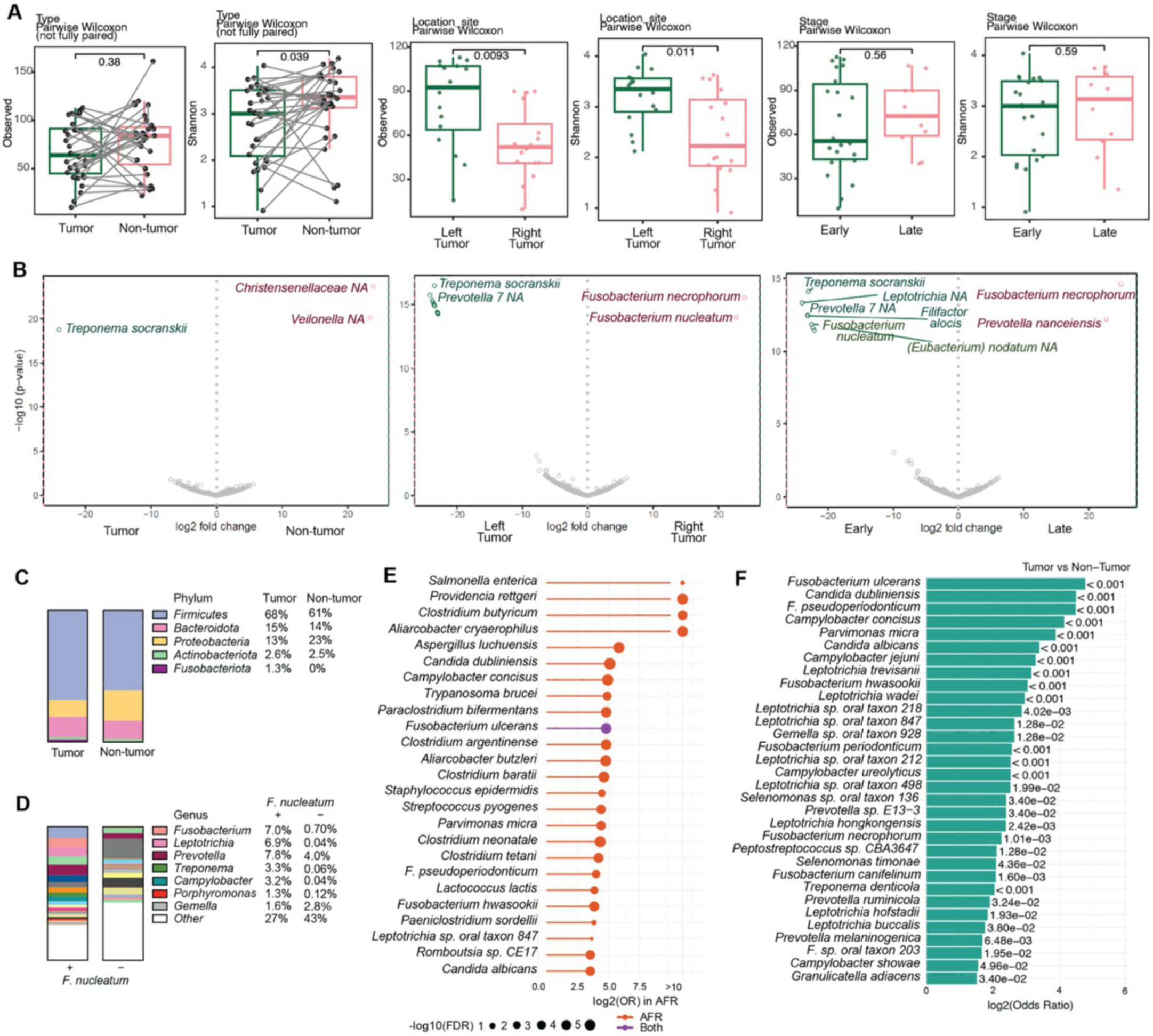
Microbial diversity and tumor-enriched taxa in colon cancer tissues. **A)** Alpha diversity (Observed richness, Shannon index) in tumor vs. non-tumor samples (n = 32, 31), and tumors stratified by location (right, n = 16; left, n = 16) or stage (early, n = 22; late, n = 10). **B)** Differential abundance analysis at the species level comparing tumor vs non-tumor (left), right vs left tumor (middle), and early vs late stage tumor (right). Volcano plots highlight significantly enriched taxa (FDR < 0.05). **C)** Phylum-level taxonomic composition in tumor and non-tumor tissues, showing enrichment of *Fusobacteriota* in tumors. **D)** Genus-level composition of tumors stratified by *Fusobacterium nucleatum* detection by qPCR (positive vs. negative). **E)** Top species enriched in tumors from African (AFRg) and European (EURg) ancestry patients (n = 68, WGS); dot size reflects significance (–log10 FDR); colors denote ancestry specificity (red = AFR only, purple = shared). **F)** Oral-origin taxa significantly enriched in tumors based on presence/absence data and annotation using the Human Oral Microbiome Database (HOMD v16.01). Bar length indicates log2 odds ratio of presence in tumor vs non-tumor (Fisher’s exact test). 16S rRNA sequencing was performed in a sub-cohort of 32 patients (A–D), and WGS in 68 patients (E–F) from the P-1000 cohort.

We also performed metagenomic analysis based on WGS of tumor and matched non-tumor tissues (P-1000 cohort), similarly to another AFRg CRC cohort (Gurjao *et al*., 2024). Briefly, we used Kraken2 (Wood, Lu and Langmead, 2019)(Lu *et al*., 2022) to assign P1000 reads to bacterial species, and the outputted abundances were re-estimated using Bracken (Lu *et al*., 2017). A binary heatmap of taxa significantly enriched in tumors (Fisher’s exact test, FDR < 0.05, OR > 1) revealed heterogeneous microbial patterns across samples. Taxa were clustered hierarchically, and samples were grouped by clinical features including laterality, stage, CMS, MSI status, and ancestry (**Figure S16**). In ancestry-stratified analyses, 83 taxa were significantly enriched in AFRg tumors (FDR < 0.05, OR > 1), while no taxa reached significance only in EURg tumors (**Table S9**). The top 25 AFR-enriched taxa ranked by log₂(OR) included multiple oral-associated species, such as *Parvimonas micra*, *Fusobacterium nucleatum*, and *Treponema denticola*. Of these, *Fusobacterium ulcerans* was the only species also significantly enriched within AFR and EUR tumors (**Figure 6E**).

In addition, leveraging the WGS-derived metagenomic results, we performed a co-abundance network analysis using SparCC (Jen *et al*., 1994; Friedman and Alm, 2012) in order to detect correlation networks (**Figure S17**). We strikingly find five well defined co-abundance clusters, including two that pointed to an enrichment in oral-origin bacterial species (Clusters 2 and 4) (**Figure S18**). Cluster 2 contained hallmark oral anaerobes, including *Fusobacterium nucleatum* and *F. periodonticum, Porphyromonas gingivalis*, *Prevotella* spp. *(intermedia, nigrescens, oris, denticola), Selenomonas sputigena,* and *Dialister pneumosintes*. Cluster 4 similarly aggregated an oral consortium comprising *Parvimonas micra, Filifactor alocis, Tannerella forsythia, Porphyromonas endodontalis, Fusobacterium* spp. *(hwasookii, pseudoperiodonticum), Streptococcus (anginosus, constellatus, oralis), Gemella morbillorum,* and multiple oral Prevotella (**Table S10**). These oral species commonly co-assemble in biofilm and function together in the oral cavity, and are significantly enriched in a chronic oral disease known as periodontitis (Colombo *et al*., 2009). To further identify oral-origin microbes enriched in tumor tissues, species-level annotations were matched to genera listed in the Human Oral Microbiome Database (HOMD v16.01). Of the 115 tumor-enriched species, 32 (28%) were classified as oral bacteria. Prominent examples include *Fusobacterium ulcerans*, *Candida dubliniensis*, *Parvimonas micra*, *Leptotrichia wadei*, and several *Fusobacterium* and *Leptotrichia* oral taxa (**Figure 6F**). These findings suggest that oral microbial consortia may contribute to tumor microbiome organization.

## Discussion

In this study, we present a multi-omic characterization of colon cancer in AFR and EUR ancestry patients, identifying ancestry-associated differences in somatic mutation profiles, microsatellite instability, and tumor microbiome composition. We noted an increased mutation frequency of *KRAS* among our P-1000 and FFPE cohorts of African genetic ancestry compared to individuals of European ancestry. We also saw a higher level of MSI-H tumors in our P-1000 AFRg group (9/42, ∼21%) compared to other studies, several of which note a lower frequency of MSI-H tumors in African Americans (Myer *et al*., 2022; Matejcic *et al*., 2025). This may be partly due to the high proportion of right sided tumors in the P-1000 AFRg (62%), since MSI-H tumors tend to be more frequently located in the right colon (Baran *et al*., 2018). Several genes of possible interest showed suggestive enrichment in the AFRg compared to European samples, albeit with small sample numbers. In the P-1000 WGS cohort, we note *ZNF469*, *RBMXL3, ZBTB7A*, *SMAD2*, and *NAA15* (**Figure 3A** and **Figure S19)**. *NAA15* is involved in the regulation of cell proliferation and metastasis and has increased expression in colon cancer (Koufaris and Kirmizis, 2020; Zhu *et al*., 2024). *ZNF469* is a zinc finger protein, a gene family that has been increasingly studied in CRC (Yu *et al*., 2025). *ZBTB7A* has been linked to poor prognosis and increased tumor progression in CRC (Wang *et al*., 2020). *RBMXL3* was found to be a late mutation in progression from adenoma to carcinoma (Wolff *et al*., 2018). *SMAD2* has been implicated as a tumor suppressor in CRC, and mutations in *SMAD2* contribute to inactivation of the TGF-β pathway (Fleming *et al*., 2013). *SMAD2* mutation have also been correlated with lower overall survival (Amani and Peymani, 2024), and association of higher *SMAD2* frequency in individuals of AFR ancestry was noted in a recent large cohort study (Matejcic *et al*., 2025). The Exome FFPE AFRg samples showed a slight indication of increased frequency of mutations than the EURg cohort in *FBLIM1*, *LRP1B*, *TBXA2R*, *LTBP1* and *CSMD2* (**Figure S5**). *FBLIM1* has been shown to be hypermethylated in CRC (He *et al*., 2023), *TBXA2R* has been shown to be overexpressed in CRC (Li *et al*., 2015) and *LTBP1* has been shown to be overexpressed and correlated with poor prognosis in gastric cancers (Li *et al*., 2015; Jiang *et al*., 2024). *LRP1B* and *CSMD2* are likely tumor suppressors previously shown to be down regulated in CRC (Zhang and Song, 2014; Wang *et al*., 2017). Though larger cohorts would be necessary to corroborate these findings, this study has identified potentially interesting differences to explore in larger datasets, and particularly in cohorts which utilize whole exome or whole genome sequencing rather than panels which may not assess the full spectrum of variants contributing to disease.

Gut microbiome composition contributes to CRC risk, with race-associated differences increasingly identified in intratumoral microbial profiles (Ahn *et al*., 2013; Feng *et al*., 2015; Flemer *et al*., 2018; Luo *et al*., 2022; Tortora *et al*., 2022). Using 16S rRNA gene sequencing on 66 matched tumor and adjacent non-tumor samples from AFRg colon cancer patients, we found reduced microbial diversity in tumor tissues, with *Fusobacteriota* detected exclusively in tumors, consistent with prior reports of higher pro-inflammatory bacterial abundance in AA compared to White Americans (Farhana *et al*., 2018). The underrepresentation of AFR patients in microbiome studies means that potentially disease-relevant microbial taxa unique to this population remain uncharacterized in the context of CRC (Tričković *et al*., 2025). These observations suggest the need for expanded inclusion of African-ancestry cohorts to reduce bias and enhance generalizability. Consistent with this, our AFRg tumor dataset includes several less-reported taxa, broadening taxonomic coverage.

To our knowledge, this is the first study to evaluate tumor location-associated microbial differences using fresh-frozen tissue from AFRg patients. Right-sided tumors exhibited significantly lower alpha diversity, with oral-associated genera, including *Fusobacterium, Prevotella, Treponema, Leptotrichia,* and *Porphyromonas*, comprising ∼17% of the right-sided tumor microbiota. The co-enrichment of these oral taxa suggests coordinated translocation from the oral cavity to the proximal colon, potentially preserving organized biofilm-like consortia analogous to oral plaque structure (Koliarakis *et al*., 2019). Periodontitis, a condition marked by oral dysbiosis and chronic inflammation, has been linked to increased CRC risk (Momen-Heravi *et al*., 2017; Hoare *et al*., 2019) and is more prevalent among AAs (Eke, Borgnakke and Genco, 2020). Additionally, *Fusobacterium* presence increases CRC risk (Ahn *et al*., 2013). In our study, several *Fusobacterium* strains were enriched in tumors, which underscores its potential role in CRC pathogenesis, particularly in populations with a high burden of periodontal disease, such as AAs.

There are many genes of interest in CRC that have not been targeted in the MSK impact panel nor med oncoGS. Panel studies have been an attractive option for large sequencing projects due to their low cost, however, relying on the use of panels rather than exome or whole genome sequencing may miss important contributions to disease. Today lower sequencing costs make whole genome projects more tenable and alleviates the possibility of missing potential drivers not found on panels or covered by exomes. Furthermore, long read whole genome sequencing, which has been shown to provide a much more comprehensive view of the genome than short reads, may be an important application to elucidate undiscovered genomic changes that may underlie racial disparities in carcinogenesis. Compared to short reads, long reads uncover thousands of structural variants as well as improving accessibility of complex genomic regions including medically relevant genes and regions of regulatory impact (Sudmant *et al*., 2015; Nattestad *et al*., 2018; Ebbert *et al*., 2019; Abel *et al*., 2020; Aganezov *et al*., 2020; Miller *et al*., 2021; Gulsuner *et al*., 2024; Mahmoud *et al*., 2024).

We acknowledge technical heterogeneity across cohorts, including differences in sample source (fresh tumor vs. FFPE), sequencing platform (WGS vs. exome capture), and control type (matched adjacent non-tumor vs. unmatched public controls). Despite these limitations, these datasets constitute a valuable and rare resource for CRC health disparities research. Critically, current KRAS-targeted therapies focus on the G12C mutation (Hoyek *et al*., 2024) yet G12D, G12V or G13D variants, more commonly observed in AFR, remain without approved targeted agents (Sylvester *et al*., 2012; Myer *et al*., 2022) (**Figure S6**). Addressing this gap requires both expanding AFR representation in genomic databases and broadening the scope of therapeutic development to reflect the mutational landscape of all patient populations. Beyond tumor biology, area-level factors such as neighborhood environment, socioeconomic status, and healthcare access likely contribute to the CRC outcome disparities observed between AFR and EUR populations and warrant investigation in future studies. Ultimately, increasing AFR representation in genomic databases and clinical cohorts is essential to identifying ancestry-specific drug targets and informing more equitable prevention and treatment strategies.

## Supplemental information

Document S1. Supplemental Figures, Figures S1–S20, and Tables S1–S2

Document S2. Supplemental Notes, Notes S1-S5

Table S3. Continental admixture proportion for the profiles in the NYGC P-1000 cohort

Table S4. Continental admixture proportion for the profiles in the CSHL FFPE cohort

Table S5. Sub-continental admixture proportion for the profiles in the NYGC P-1000 cohort

Table S6. Sub-continental admixture proportion for the profiles in the CSHL FFPE cohort

Table S7. The up and down-regulated GSEA results for the HALLMARK geneset analysis

Table S8. Microbiome relative abundance at the genus-level

Table S9. Tumor species enrichment by ancestry

Table S10. Species–cluster assignments from tumor and matched non-tumor WGS

Table S11. Regions of recurring large copy number events in the NYGC AFRg patients with MSS tumors.

Table S12. Difference, in the median expression, between samples having an event vs samples without event, in the regions of recurring large copy number events in the NYGC AFRg patients with MSS tumors.

Table S13. Noncoding variant enrichment

## Methods

### Participant Recruitment and Data Collection

This cohort was recruited in the context of the New York Genome Center (NYGC) Polyethnic-1000 project (P-1000) (Robine and Varmus, 2022). Specifically, study participants were recruited from the State University of New York (SUNY) Downstate Medical Center and Northwell Health under the respective institutional IRB approved protocols. Study eligibility criteria included treatment-naive patients diagnosed with colon adenocarcinoma (COAD) with [age of diagnosis at least 50]. We excluded individuals who had treatment prior to sample collection, did not self-identify as “White” or “Black or African American”, who identified as “Hispanic or Latino” and whose tumor had advanced to stage IV. Additional clinical and pathological metadata was collected at each recruitment site, harmonized, captured in a study-specific RedCap database and de-identified cohort metadata exported for analysis.

### Patient Cohort

#### Polyethnic-1000 consortium

Patients aged ≥45 years with histologically confirmed colon adenocarcinoma were enrolled at SUNY Downstate Medical Center and Kings County Hospital under IRB-approved protocol #312509. Eligibility was limited to individuals self-identifying as African American or Caribbean American who provided informed consent. Exclusion criteria included HIV, Hepatitis B/C, inflammatory bowel disease (Crohn’s or ulcerative colitis), immunosuppressive or antibiotic treatment at the time of enrollment, prior neoadjuvant therapy, history of other malignancies, rectal cancer, and hereditary colorectal cancer syndromes (Lynch syndrome or familial adenomatous polyposis).

### Clinical Metadata

Demographic and clinical variables were collected, including age, sex, race/ethnicity, BMI, diabetes, smoking status, and alcohol use. Pathology reports were used to confirm tumor stage, volume, histological differentiation, TNM classification, microsatellite instability (MSI) status, and mismatch repair (MMR) protein expression. Follow-up data included treatment history, recurrence, and survival.

### NYGC sequencing of WGS samples

#### Nucleic Acid extraction from Fresh Frozen Tissue

Purified genomic DNA and total RNA were extracted from fresh frozen tissue following the Qiagen Allprep DNA/RNA (Qiagen,80204) procedure. Fresh frozen tissue was added to a mixture of Buffer RLT/B-mercaptoethanol for homogenization with bead mill, Tissuelyser LT (Qiagen, 85600). DNA was bound to the DNA spin-column while the RNA portion is in the flow-through and processed further with the RNeasy MinElute spin column. Both DNA and RNA separately followed a series of washes for sample purification and final elution.

#### WGS library preparation and sequencing, Truseq PCR-free (450bp)

Whole genome sequencing (WGS) libraries were prepared using the Truseq DNA PCR-free Library Preparation Kit (Illumina) in accordance with the manufacturer’s instructions. Briefly, 1ug of DNA was sheared using a Covaris LE220 sonicator (adaptive focused acoustics). DNA fragments underwent bead-based size selection and were subsequently end-repaired, adenylated, and ligated to Illumina sequencing adapters. Final libraries were quantified using the QuantStudio5 (Life Technologies) and Fragment Analyzer (Advanced Analytical) or Agilent 2100 BioAnalyzer. Libraries were sequenced on an Illumina Novaseq6000 sequencer using 2×150bp cycles.

Alignment of reads to the human reference, alignment processing and variant calling (SNVs, indels, SVs and CNVs) were performed using the NYGC somatic WGS pipeline, as described in Arora et al. (Arora *et al*., 2019) For full documentation also see documentation from NYGC (NYGC, 2019).

#### RNA library preparation and sequencing, KAPA RNA HyperPrep Kit with RiboErase (HMR)

RNA sequencing libraries were prepared using the KAPA RNA HyperPrep Kit with RiboErase (HMR) (Roche 08098140702) in accordance with the manufacturer’s instructions. Briefly, samples were normalized to 100ng and underwent rRNA depletion through oligo hybridization and RNase H treatment. DNase treatment removed DNA oligos. Samples were fragmented to 350bp inserts and underwent cDNA generation. Second strand synthesis and A-tailing were combined to generate dscDNA. Samples were barcoded with Illumina sequencing adapters and amplified. Final libraries were quantified using the Qubit Fluorometer (Life Technologies) or Spectramax M2 (Molecular Devices) and Fragment Analyzer (Agilent). Libraries were sequenced on an Illumina Novaseq6000 sequencer using 2×100bp cycles.

Reads were aligned with STAR (version 2.5.2a) (RRID:SCR_004463) (Dobin *et al*., 2013), and annotated genes were quantified with featureCounts (v1.4.3-p1) (RRID:SCR_012919) (Liao, Smyth and Shi, 2014). The GRCh38 genome canonical chromosomes (chr1-22, X, Y, and M) were used for alignment utilizing GENCODE v25 annotations (RRID:SCR_014966)(Harrow *et al*., 2012).

### CSHL sequencing of FFPE samples

#### Exome capture, sequencing and variant calling

IDT Xgen probes designed for reference hg38 were used to target ∼34Mb of the human exome. FFPE blocks for each sample were cut and ten curls of 10uM thickness were selected for each sample. DNA extraction was performed using QIAamp FFPE tissue kit. DNA was assessed for quality and purity using BioAnalyzer and Qubit. In the event that DNA extraction yield was low, additional curls were cut for the sample to obtain adequate yield.

Extracted DNA was size selected using Ampure beads to target fragments in the ∼450bp range, though some FFPE material was fragmented resulting in smaller sizes. Barcoded Illumina adapters were ligated to the size selected fragments. The resulting libraries were then pooled in sets of 6. These 6plex pools were hybridized with the capture probes for 72 hours, followed by washing and PCR enrichment. The captures were then pooled into sets of 18 samples and sequenced on an Illumina NexSeqSeq2000 instrument to achieve ∼150-200X coverage. Demultiplexing samples into separate FASTQ files by barcode was performed with bcl2fastq v2.20.0.422 (RRID:SCR_015058).

We then employed an analysis pipeline to prioritize identification of driver mutations. The sequencing reads were aligned to the hg38 reference genome using BWA v0.7.17 (Li, 2013) (RRID:SCR_010910), followed by conversion to BAM format and sorting with SAMTOOLS v1.9 (Li *et al*., 2009) (RRID:SCR_002105), removal of PCR duplicates with Picard v1.88 (*Picard*, 2019) (RRID:SCR_006525), and filtering with Bamtools v2.5.1 (*Picard*, 2019; Barnett *et al*., 2011) (RRID:SCR_015987) for mapping quality and proper read pairing.

Coverage of the target regions was assessed for breadth and depth using Picardv1.88 (RRID:SCR_006525) HSmetrics to ensure adequate coverage for confident variant detection of low frequency somatic variants. SNP and small indel variants are then called using VarScan2 v2.3.9 (Koboldt *et al*., 2012) (RRID:SCR_006849) with filters for minimum sequence depth of 10X and minimum alternate allele frequency of 10%, along with strand bias filters.

#### Variant prioritization

Variant calls from all samples were then annotated with ANNOVAR (Wang, Li and Hakonarson, 2010) (RRID:SCR_012821) to cover a broad range of variant assessment tools including prediction of deleteriousness (dbNSFP v41a (RRID:SCR_005178), SIFT (Lowe, 1999) (RRID:SCR_012813), Polyphen (*Object recognition from local scale-invariant features*, no date; Adzhubei, Jordan and Sunyaev, 2013) (RRID:SCR_013189), MutationTaster (Schwarz *et al*., 2010) (RRID:SCR_010777), etc.) and conservation scores (CADD(Rentzsch *et al*., 2018) (RRID:SCR_018393), GERP (RRID:SCR_000563), DANN (Quang, Chen and Xie, 2014; Rentzsch *et al*., 2018), etc.). We then selected rare loss of function variants (nonsense, frameshift, splice site) with frequency less than 1% in the gnomAD (Karczewski *et al*., 2020) (RRID:SCR_014964), ExAc (Karczewski *et al*., 2016) (RRID:SCR_004068), and 1000 Genomes (RRID:SCR_006828) databases. Missense and in-frame indel variants were selected if they were noted as pathogenic by ClinVar 20240917 (Landrum *et al*., 2015) (RRID:SCR_006169), or if they are both rare and annotated as pathogenic by COSMIC v96 (Karczewski *et al*., 2016; Tate *et al*., 2019) (RRID:SCR_002260), or if they are both rare and found to be present in the TCGA (*The Cancer Genome Atlas Program (TCGA)*, 2022) (RRID:SCR_003193) or ICGC(Zhang *et al*., 2019) (RRID:SCR_021722) cohorts.

Several measures were taken to select for putative somatic variants in the FFPE exomes which lacked matched normals. Variants were filtered against the 1000 genomes pool of normals variant panel as well as our 6 platform matched HapMap controls. In addition, for the FFPE exomes we requested access for DFCI tumor-normal pairs to model alternate allele frequency of germline versus somatic variants when running our pipeline with matched normals. We then applied a filter for alternate allele frequency of less than 40% to our dataset to remove likely germline events.

Further evaluation of these candidate variants was performed using Maftools v2.14 (Mayakonda *et al*., 2018) (RRID:SCR_024519). Variant frequency per gene across samples was assessed and variant summaries and oncoplots were generated. AA vs EA cohorts were compared using the Maftools function mafCompare, performing a fisher’s exact test to assess differentially mutated genes in each sample set. Variants had to present in a minimum of 3 samples per cohort to avoid bias of mutations in a single sample. Additional analysis was performed with maftools oncopathways, somatic interactions, clinicalEnrichment, lollipopPlots, and tcgaCompare. TCGA COAD (Kirk *et al*., 2016) MSI status was derived from (Vaughan-Shaw *et al*., 2021) and DFCI cohort was stratified by mutations in MMR genes and high TMB.

Non-coding variants were analyzed with MutEnricher v1.3.3 (Soltis *et al*., 2020) to identify likely regulatory impact or transcription modifiers by overlap with ENCODE (ENCODE Project Consortium, 2012) (RRID:SCR_006793) regions such as promoters, enhancers, DNAse hypersensitivity sites, histone marks and TFBS, as well as conserved sequence.

#### NYGC cohort genetic ancestry inference

The continental and sub-continental ancestral admixtures of the P-1000 patients, relative to a reference population set based on 1000 Genomes (1KG) project (Fairley *et al*., 2020), were both inferred with ADMIXTURE (Alexander, Novembre and Lange, 2009) software version 1.3.0 (RRID:SCR_001263) using the matching non-tumor genome sequencing. The reference population set was prepared by first genotyping the markers as previously described (Martini *et al*., 2022). The patients with dominant European (EUR) and African (AFR) continental admixture ancestries (70% or more) were respectively assigned in the EURg and AFRg groups.

#### CSHL cohort genetic ancestry inference

First, the base quality scores of the aligned tumor exomes were recalibrated to remove systematic errors made by the sequencing machine (BaseRecalibrator and ApplyBQSR programs) using GATK (Van der Auwera and O’Connor, 2020) software version 4.2.5.0 (RRID:SCR_001876). Allele-specific read counts at genome positions with high single-nucleotide variant frequencies in the 1000 Genomes population reference were extracted using the snp-pileup program present in FACETS (Shen and Seshan, 2016; Van der Auwera and O’Connor, 2020) software version 0.6.1 (RRID:SCR_026264). The continental and sub-continental ancestral admixtures of the patients were inferred using an in-house version of RAIDS (Belleau *et al*., 2023, 2025) software (RRID:SCR_027265) invoking ADMIXTURE (Alexander and Lange, 2011) software version 1.3.0 in supervised mode (RRID:SCR_001263) as a component. The final results include the proportional breakdown into 5 continental and 18 subcontinental populations for each CSHL patient. Patients with dominant European (EUR) and African (AFR) continental ancestries (70% or more) were assigned to the EURg and AFRg groups, respectively.

For both the NYGC and CSHL cohorts, the continental and sub-continental ancestry admixture graphs were generated with CRAN gglot2 (Wickham, 2016) package version 3.5.1 (RRID:SCR_014601) and CRAN cowplot (Wilke, 2015; Wickham, 2016) package version 1.1.3 (RRID:SCR_018081). The Sankey graphs showing the relation between race and ancestry have been generated with the CRAN networkD3 package version 0.4 (Allaire, 2025) (RRID:SCR_027266).

#### NYGC cohort copy number inference

The ASCAT (Van Loo *et al*., 2010; Yan, 2023) software version 3.1.2 (RRID:SCR_016868) was used to call copy numbers, as well as tumor purity and ploidy (genome-wide average of copy number) for each tumor sample. The cut-off for the detection of the amplification and deletion events were respectively set to 0.2 and -0.235 in difference between copy number and ploidy. The AFRg profile C-45T failed the ASCAT convergence step for ploidy and purity estimation and was removed from following analyses.

The AFRg patients were retained for the frequency plot of the amplification and deletion events, as obtained with ASCAT, using the Bioconductor gtrellis (Gu, Eils and Schlesner, 2016b) package version 1.36.0 (RRID:SCR_027267).

Similarity between copy number profiles of the EURg and AFRg cohort has been calculated with Bioconductor CNVMetics (Wickham, 2016; Wilke, 2015; Deschênes *et al*., 2022) package version 1.6.0 (RRID:SCR_027263). The BIC-seq copy number profiles (see above for CNV calling methods), deletion and amplification in two separated sets, were used as input. The similarity metric retained was the Szymkiewicz–Simpson coefficient (M.k and K, 2016), also known as the overlap coefficient, which is calculated by dividing the size of the intersection of the copy number events (in base paired) by the smaller size of the two sets. Significance of the observed metrics has been assessed, in comparison to the null distribution, using simulated profiles generated by the CNVMetrics package. For each pair of profiles, a minimum of 147 and up to 500 synthetic profiles were generated for one profile of the pair (when a metric could not be successfully calculated with a synthetic profile, the synthetic profile was discarded). The similarity heatmaps were generated with Bioconductor ComplexHeatmap package version 2.17.0 (Gu, Eils and Schlesner, 2016a) (RRID:SCR_017270) using the Euclidean distance and the complete method for clustering. The Venn diagram was generated with the CRAN ggvenn (Yan, 2023) package version 0.1.10 (RRID:SCR_025300).

#### NYGC cohort copy number variations signatures

The copy number variations signatures (COSMIC CN) were extracted, for the AFRg and EURg patients from the P-1000, via the SigProfilerAssignment tool v0.2.6 (Díaz-Gay *et al*., 2023) (RRID:SCR_023121). The CNVs from ASCAT were used as input (41 AFRg profiles available). The copy number variations signatures were also extracted for the samples in the TCGA COAD (Kirk *et al*., 2016) using an identical pipeline. For this cohort, the ASCAT3 copy number variants, as made available by the NCI Genomic Data Commons (GDC) (Heath *et al*., 2021), were used as input. In both cohorts, the signatures are considered detected in a patient when at least 10 mutations are present in the signature.

The heatmaps of the copy number variations signatures for both the P-1000 and TCGA COAD cohort, using only AFRg and EURg assigned patients, were generated with Bioconductor ComplexHeatmap package v2.22.0 (Gu, Eils and Schlesner, 2016a) (RRID:SCR_017270). The consensus molecular subtyping of the TCGA COAD patient is described later. When two different subtypes were associated to the same patient (in presence of multiple samples from the same patient), the patient was removed from the final graph.

#### NYGC regions of recurring large copy number events

The AFRg patients with MSS tumors were retained for this analysis. Recurring copy number amplification and deletion events were identified with CRAN CORE (Krasnitz *et al*., 2013) package v3.2 (RRID:SCR_027419) using Core I setting for larger event detection. The copy numbers, as obtained from JaBbA, were used as input with the exclusion of the HLA region (chr6:28600000-33200000) and as well as the backlisted regions. A total of 103 regions were significantly detected on chromosomes 1 to 22 (<u>Table S11</u>).

Genes present in each region were identified using the Biconductor TxDb.Hsapiens.UCSC.hg38.knownGene v3.20.0 and GenomicRanges (M. Lawrence *et al*., 2013) (RRID:SCR_000025) v1.58.0 packages. Genes associated with cancer were identified using the COSMIC Cancer Gene Census database v102 (Sondka *et al*., 2024) (RRID:SCR_002260).

When a region contained at least 5 genes with associated expression, the region was tested to see if the events were associated with a difference in expression (Wilcoxon Rank Sum test) using only patients with either neutral event or event corresponding to the one identified in the region (amplification or deletion). The *p*-values were adjusted for multiple testing using the Benjamini–Hochberg false discovery rate (FDR) procedure (T<u>able S12</u>).

#### NYGC cohort HLA typing

The HLA typing was performed on the cancer and the matching non-tumor genome sequencing with the help of Kourami (Lee and Kingsford, 2018) version 0.9.6 (RRID:SCR_022280). Differences between the cancer and matching non-tumor types were then assessed. First, the fields 1 and 2 were extracted for the comparison. Loss of heterozygosity was assigned when at least two different types were detected in the matching non-tumor, but only one remained in the cancer. A difference in typing was assigned when at least one of the types differed between the matching non-tumor and the cancer (see <u>Note S</u>4 and <u>Figure S15A</u>). The HLA typing graph has been generated with the Bioconductor ComplexHeatmap (Gu, Eils and Schlesner, 2016a) package v2.22.0 (RRID:SCR_017270).

The HLA-A, B, C, DQB1 and DRB1 allele frequencies were calculated specifically in the subsection of the NYGC cohort that was assigned African ancestry. The ranking of the most frequent alleles was then compared to the National Marrow Donor Program (NMDP) registry haplotype frequencies for the African Americans (Gragert *et al*., 2013). The NMDP registry has created group annotations for some alleles. In the case that a specific allele was not found, the allele was looked up against the NMDP allele groups (see <u>Note S</u>5 and <u>Figures S15B-F</u>). The HLA frequency graphs were generated with the ggplot2 (Wickham, 2016; Gragert *et al*., 2013) v3.5.2 (RRID:SCR_014601).

### Transcriptome analyses

#### RNA Quality control analysis and data preprocessing

Sample quality was assessed using common quality control metrics, specifically read distribution across different genomics regions (intergenic regions, intronic regions, untranslated regions, and protein coding regions), mean GC% content, read mappability and mean inner distance between paired reads. These metrics were calculated using Picard v2.25.4 (*Picard*, 2019; Wickham, 2016; Gragert *et al*., 2013) (RRID:SCR_006525) and RSeQC v2.6.2 (Wang, Wang and Li, 2012) (RRID:SCR_005275). Two samples (C-05T, C-06T) were excluded due to the high proportion of unmapped reads while one sample was marked for exclusion due to the high value of mean inner distance between read pairs (C-22T).

Read counts across the two batches were preprocessed together using the *DESeq2* R package (version 1.46.0) (RRID:SCR_015687). After exclusion of the 3 bad quality, 3 metastatic and 3 rectal samples, genes with zero counts in more than 20% of the samples were removed and data were normalized for library size. Variance stabilizing transformation was further applied on the data. Batch correction was performed using limma::removeBatchEffect() R software (version 3.62.2) (RRID:SCR_010943), while accounting for the different levels of continental admixture proportions across samples.

#### Consensus molecular subtyping

We performed subtyping analysis in primary colorectal tumors from African and European ancestry patients (n=43) using the R package CMScaller (Wang, Wang and Li, 2012; Eide *et al*., 2017) (version 2.0.1) (RRID:SCR_027275). CMScaller classifies colorectal cancer samples into one of four subtypes based on consensus molecular subtypes (CMS) first described in Guinney et al. CMScaller uses the Nearest Template Prediction (NTP) algorithm to categorize the samples into the four biologically distinct CMS while expanding the expression signatures used with additional cancer cell-intrinsic subtype-enriched markers to achieve higher classification performance, especially in identifying CMS4 samples. We assessed the association between gene expression based molecular subtype and inferred genetic ancestry group using Fisher’s exact test. CMScaller predictions were generated for the TCGA-COAD cohort following the same process.

#### Differential gene expression analysis

To identify gene expression patterns associated with African (AFR) ancestry in primary colorectal tumors, we employed a multivariate linear regression model using the Bioconductor *limma (Wu, 2017)* package (version 3.62.2) (RRID:SCR_010943). The model included CMS subtype, tumor purity, age, and sex as covariates. We compared tumor samples from individuals with predominantly African (AFRg n = 35) and European (EURg n = 7) ancestry. Genes were considered significantly associated with AFR ancestry at a Benjamini-Hochberg adjusted p-value < 0.05 and an absolute effect size ≥ 0.1.

Differential gene expression analysis between early- and late-stage COAD tumors in the TCGA cohort was conducted using *limma*, controlling for CMS subtype, tumor purity, age, sex and genetic ancestry (categorical variable). Genetic ancestry assignments for TCGA samples are based on consensus calls derived from five independent tools, as reported by Carrot-Zhang et al. (Carrot-Zhang *et al*., 2020)

#### Pathway Activation Analysis

Pathway activation scores for the transcriptomic cohort were calculated using the PROGENy model within the *decoupleR* R package (version 2.12.0). Significant pathway activation differences by CMS were calculated using one-way ANOVA.

#### Gene set enrichment analysis

A gene set enrichment analysis was performed on the log2 fold change rank-ordered gene list from the ancestry based differential gene expression analysis using the GseaPreranked method [set_min=15, set_max=500, nperm=1000, scoring_scheme=weighted, rnd_seed=22, chip=Human_Gene_Symbol_with_Remapping_MSigDB.v2025.1.Hs.chip] in the GSEA (Subramanian *et al*., 2005) software version 4.3.3 (RRID:SCR_003199) on the hallmark gene sets (h.all.v2025.1.Hs.symbols.gmt) from the Molecular Signatures Database (MSigDB) (Liberzon *et al*., 2015) (RRID:SCR_016863).

#### Gene set variation analysis

The variation of the hallmark KRAS signaling up gene set (from h.all.v2025.1.Hs.symbols.gmt) was calculated on the AFRg samples with MSS tumors, and available RNA. The analysis was performed using the GSVA (Hänzelmann, Castelo and Guinney, 2013) package v2.0.4 (RRID:SCR_021058). The batch-corrected variance stabilizing transformed (vst) expression from library-size adjusted read counts was used as input (see RNA Quality control analysis and data preprocessing section). The comparison of the enrichment score distribution between the KRAS mutated and KRAS wild-type groups was done with a Wilcoxon rank sum exact test. The associated graphs were generated with CRAN ggplot2 (Wickham, 2016; Hänzelmann, Castelo and Guinney, 2013) package v4.0.0 (RRID:SCR_014601) and CRAN cowplot (Wilke, 2015) package v1.1.3 (RRID:SCR_018081).

#### Cell Type Deconvolution Analysis

Immune cell type abundance in colorectal tumor samples was inferred using CIBERSORT (Newman *et al*., 2015) (version 1.04) (RRID:SCR_016955). We used the CIBERSORT LM22 gene signature matrix, which contains 547 genes that distinguish 22 human hematopoietic cell phenotypes as the reference gene expression matrix. Significant differences in immune cell type proportions between tumors derived from African and European patients were calculated using Fisher’s exact test.

### Microbiome Analyses

#### Microbiome DNA isolation and collection

We only used fresh frozen colon tissues for 16S rRNA sequencing, analysis was done at the Microbiome Core at Columbia University Irving Medical Center. A total of 33 paired tumor and adjacent non-tumors tissue samples, each not exceeding 25 mg were used. Each sample underwent two washes with Phosphate-Buffered Saline (PBS) to remove any residue. After the PBS was discarded, the samples were processed according to the manufacturer’s recommended protocol using the QIAamp® 96 DNA QIAcube® HT system (Qiagen) for pretreatment. DNA was extracted using the QIAamp® 96 Virus QIAcube® HT kit (Qiagen), following the manufacturer’s instructions. The DNA was then eluted in 100 µl of RNase/DNase-free buffer, and the DNA yield was determined using the Qubit 2.0 Fluorometer (Invitrogen).

#### 16S rRNA gene amplification and analysis

Considering the presumed high host DNA content, all samples were treated as having low bacterial DNA content. Consequently, we employed the Zymo Quick-16S/V3V4 Plus NGS Library Prep Kit, a qPCR-based library preparation technique, to optimize bacterial 16S rRNA gene amplification and identify the most suitable region for this study. PCR amplifications for the target region was performed using the CFX Opus 96 Real-Time PCR System (Bio-rad). After amplification, the resulting 16S libraries were pooled by adding an equal volume to each, and the final library concentration was quantified using the Qubit 2.0 Fluorometer (Invitrogen). Following equimolar pooling based on these quantifications, sequencing was conducted on the Illumina MiSeq platform. A loading concentration of 8pM, supplemented with 20% PhiX, was used along with the paired-end 300 cycle MiSeq Reagent Kit V3 (Illumina).

16S rRNA sequences from targeted (V3V4) were processed and applied using the DADA2 (Callahan *et al*., 2016) (DADA2 1.12.1) pipeline (RRID:SCR_023519) in conjunction with R v4.1.0 (RRID:SCR_001905). DADA2 was employed for quality filtering, trimming, error correction, exact sequence inference, chimera removal, and generation of the amplicon sequence variant table (ASV) with a minimum count cutoff set at 2000. Based on the quality score profiles of the sequencing reads, forward reads were truncated at 240 bp, and reverse reads were truncated at 240 bp before merging. Ambiguities in the overlap region were not permitted, and default parameters were utilized in the R DADA2 package’s filterAndTrim() function [truncLen=c(240,240); trimLeft=c(5,5), maxN = 0, maxEE=c(2,2), truncQ = 2]. After the dereplication and merging of reads, chimeric reads were identified by consensus across samples using the DADA2 function removeBimeraDenovo(). All samples passed the set threshold of 2000 reads post-quality filtering for inclusion in the analysis. The MAFFT and FastTree modules in QIIME2 (Bolyen *et al*., 2019) (RRID:SCR_021258) facilitated the generation of a phylogenetic tree of all ASV sequences. Taxonomic classification was undertaken using a native naïve RDP Bayesian classifier aligned against the SILVA (Quast *et al*., 2013) version 138 database (RRID:SCR_006423).

Descriptive statistics were utilized to summarize the demographic information for the entire cohort. These data were integrated into R (version 4.1.0) (RRID:SCR_001905) to carry out analyses on alpha-diversity metrics using Shannon, and Principal Coordinates Analysis (PCoA) based on the weight and unweight Unifrac distance matrix. These analyses were conducted using the phyloseq (McMurdie and Holmes, 2013) package (v1.36.0) (RRID:SCR_013080) in R. Differences in alpha-diversity between groups were ascertained using the non-parametric Wilcoxon Rank Sum test. In assessing beta-diversity, permutational multivariate analysis of variance (PERMANOVA) was employed. PERMANOVA is a non-parametric form of multivariate ANOVA that highlights variances in sample centroids. Differential abundance (DA) analysis for bacterial species level was performed using the DESeq2 (McMurdie and Holmes, 2013; Love, Huber and Anders, 2014) (RRID:SCR_015687) and ANCOM -BC (Lin and Peddada, 2020) package adjusting for multiple comparisons (RRID:SCR_024901). To account for multiple testing, p-values were adjusted utilizing the Benjamini-Hochberg procedure, which controls the false discovery rate. To assess the differences in relative abundance at the genus-level between tumor and non-tumor tissue samples, we employed the nonparametric Wilcoxon matched-pairs signed-rank test. Statistical analyses were conducted using GraphPad Prism 10 (GraphPad Inc., San Diego, CA, USA) (RRID:SCR_002798) and R. P values of < 0.05 were considered statistically significant.

#### Microbiome analysis by whole-genome sequencing (WGS)

Computational classification of P-1000 samples at the taxonomic level was performed using Kraken2 (Wood, Lu and Langmead, 2019; Lin and Peddada, 2020) (RRID:SCR_026838), identically to a previous study (Gurjao *et al*., 2024). Briefly, particular attention was given to the removal of false positive bacterial reads by leveraging a Kraken2 database containing complete bacterial genomes and human DNA sequences (including GRCh38 and T2T-CHM13 versions). Co-abundance network analysis was conducted with SPARCC with the SPIECEASI (Kurtz *et al*., 2015) R package version 1.1.1 (RRID:SCR_022712) with default parameters.

Microbial presence/absence data were derived from whole-genome sequencing (WGS) of paired tumor and non-tumor colon tissue samples. Species prevalence matrices were generated by binarizing species abundance values, assigning presence (1) or absence (0) for each species per sample. A species was considered present if detected in ≥10% of samples and supported by more than five sequencing reads. Data was manually filtered and curated to remove potential contaminants.

Tumor and non-tumor microbiome profiles were analyzed separately for AFR and EUR ancestry patients, as determined from genomic data. For each ancestry group, taxa detected in at least two samples were tested for enrichment in tumors versus paired non-tumor tissues using Fisher’s exact test on presence/absence data. Odds ratios (OR) and associated 95% confidence intervals were calculated, and *p*-values were adjusted for multiple testing using the Benjamini–Hochberg false discovery rate (FDR) procedure. Taxa with FDR < 0.05 and OR > 1 were considered significantly enriched in tumors. For visualization, taxa were ranked by log₂(OR), and the top 25 AFR-enriched taxa were plotted. To identify taxa enriched across ancestries, the same statistical workflow was applied to AFR and EUR samples combined (“BOTH” analysis), and overlaps with ancestry-specific results were recorded. To assess the contribution of oral bacteria to tumor-enriched communities, significant taxa were compared against the Human Oral Microbiome Database (HOMD) reference catalog (v16.01) (RRID:SCR_025964). Species-level annotations from the enrichment analysis were matched to genera present in HOMD. A binary heatmap was generated using the ComplexHeatmap (v 3.21) package in R, with rows (taxa) clustered by Euclidean distance and complete linkage to highlight taxonomic co-occurrence patterns. Columns (samples) were ordered by clinical annotations, including maximum ancestry, tumor laterality, MSI status, and CMS subtype.

Statistical analyses were conducted using R version 4.4.2 (R Core Team, 2024) (RRID:SCR_001905) within RStudio version 1.4.1717. Key R packages used included dplyr (v1.x) (RRID:SCR_016708), ggplot2 (v3.x), ggpubr (v0.x) (RRID:SCR_021139), vegan (v2.x) (RRID:SCR_011950), and ape (v5.x) (RRID:SCR_017343).

#### Detection of *Fusobacterium nucleatum* in colon tissues

Primer pairs and probes custom TaqMan Gene Expression Assays (FAM) were designed to amplify human genomic sequences of the PGT gene and microbial genomic sequences of *F. nucleatum* were sourced from existing literature. The specific primer sequences (5′-3′) for the nusG gene of *F. nucleatum* are Forward primer: CAACCATTACTTTAACTCTACCATGTTCA, Reverse primer: GTTGACTTTACAGAAGGAGATTATGTAAAAATC, and FAM probe: TCAGCAACTTGTCCTTCTTGATCTTTAAATGAACC. For PGT gene sequences (5′-3′) were Forward primer: ATCCCCAAAGCACCTGGTTT, Reverse primer: AGAGGCCAAGATAGTCCTGGTAA, and FAM probe: CCATCCATGTCCTCATCTC. DNA was quantified using a Nanodrop spectrophotometer. Each reaction for qPCR contained 40ng of DNA, 1X final concentration TaqMan Universal PCR Master Mix (Applied Biosystems; Thermo Fisher Scientific), 18 mM each primer, and 5 mM probe in a total volume of 10 µL in a 384-well optical PCR plate. Cycling conditions were as follows: 2 min at 50°C, 10 min at 95°C, and 40 cycles of 15 sec at 95°C and 1 min at 60°C. Cycle thresholding was calculated using the automated settings for SDS 2.2 (Applied Biosystems). For absolute quantification, microbial genomic DNA of *F. nucleatum subsp. nucleatum* strain VPI 4355 (ATCC # 25586D-5) was used to prepare the standard curve and as a positive control; the negative control was sterile H_2_O. 10X serial dilutions were prepared of each DNA sample starting from 10 ng/µL. A standard curve using the cycle threshold (Ct) values of the standard dilutions was plotted on the x-axis and the logarithm of the initial DNA concentration of the standard dilutions on the y-axis. We then determined the efficiency of the amplification reaction using the slope of the standard curve, R values for *F. nucleatum was* 0.995.

## Resource Availability

Sequence data for the FFPE exome cohort, including aligned BAMs, and VCF files for genomic SNV/indel variants will be accessible under a dbGap study (accession in progress). Data will be controlled access as allowed by the IRB. The P-1000 WGS data including clinical metadata, aligned BAMs, and VCF files for genomic variants (SNV/indel, SV and CNV), as well as aligned reads and expression counts for the RNA-seq will be available through ICGC-ARGO(*ICGC ARGO, 2025*). Microbiome sequencing data have been submitted to the NCBI Sequence Read Archive under BioProject accession number PRJNA1336699.

## Data Availability

Sequence data for the FFPE exome cohort, including aligned BAMs, and VCF files for genomic SNV/indel
variants will be accessible under a dbGap study (accession in progress). Data will be controlled access as allowed by the IRB. The P-1000 WGS data including clinical metadata, aligned BAMs, and VCF files for genomic variants (SNV/indel, SV and CNV), as well as aligned reads and expression counts for the RNA-seq will be available through ICGC-ARGO(ICGC ARGO, 2025). Microbiome sequencing data have been submitted to the NCBI Sequence Read Archive under BioProject accession number PRJNA1336699.

## Acknowledgements

We would like to acknowledge funding support from Cold Spring Harbor Laboratory and Northwell Health Affiliation and we thank the Northwell Health Biospecimen Repository and the SUNY Downstate GI Research Laboratory for providing colon tissue samples. We thank the CSHL Cancer Center DNA Sequencing Shared Resource for providing the Illumina sequence data for the FFPE exome project. The Sequencing Shared Resource is supported by the Cold Spring Harbor Cancer Center Grant (5P30CA045508). S.G. was supported by the National Institutes of Health (5R50CA243890). W.R.M is the Davis Family Professor of Human Genetics at CSHL. This work was performed with assistance from the US National Institutes of Health Grant S10OD028632-01. A.K. acknowledges support by the National Institutes of Health (U01CA289357) and by the Simons Foundation. L.A.M. acknowledges support by the National Institutes of Health (1P20CA192994) and the 2021 Downstate Health Sciences University Seed Grant (S.C.T.). S.B was supported by the National Cancer Institute (R37CA292807), Oliver S. and Jennie R. Donaldson Charitable Trust, the Mark Foundation for Cancer Research (20-028-EDV), the Cold Spring Harbor Laboratory and Northwell Health Affiliation and Swim Across America. This work was conducted as part of New York Genome Center’s Polyethnic-1000 initiative. Funding and other external support for Polytechnic-1000 were provided in part by Illumina, Inc. Sample procurement, next-generation sequencing, and clinical data harmonization was performed by the New York Genome Center in collaboration with SUNY Downstate, Northwell Health and Cold Spring Harbor Laboratory. We thank the Broad Institute for generating high-quality sequence data supported by NHGRI funds (grant # U54 HG003067) with Eric Lander as PI. The datasets used in this manuscript were obtained from dbGaP at http://www.ncbi.nlm.nih.gov/gap through dbGaP accession number phs000722. We also thank Nurses’ Health Study (NHS) and Health Professionals Follow-up Study (HPFS) for the samples collection.

## Author Contributions

Melissa Kramer: Formal analysis, Visualization, Writing original draft, Writing review & editing

Pascal Belleau: Data curation, Formal analysis, Methodology, Software, Visualization, Writing review & editing

Sofia C. Tortora: Conceptualization, Data curation, Formal analysis, Investigation, Visualization, Writing original draft, Writing review & editing

Astrid Deschênes: Formal analysis, Software, Visualization, Writing original draft, Writing review & editing

Kyriaki Founta: Data curation, Formal analysis, Investigation, Methodology, Visualization, Writing original draft, Writing review & editing

Carino Gurjao: Data curation, Formal analysis, Investigation, Methodology, Software, Visualization, Writing original draft, Writing review & editing

Brian Yueh: Data curation, Investigation

Sara Goodwin: Data curation, Writing review & editing

Devin Gee: Data curation, Formal analysis, Investigation, Visualization, Writing original draft, Writing review & editing

Santhilal Subhash: Formal analysis, Investigation, Methodology, Software, Visualization, Writing original draft, Writing review & editing

Mali Barbi: Data curation, Supervision, Validation

Charlie Chung: Investigation

Kadir Ozler: Data curation, Investigation

Onur Eskiocak: Investigation

Benjamin Izar: Writing review & editing

Heather Geiger: Data curation, Investigation

Timothy R. Chu: Data curation, Investigation

Zoe Goldstein: Data curation, Investigation

Lara Winkerton: Data curation, Project administration

Andrew L. Araneo: Formal analysis, Investigation, Writing original draft

Richard L Whelan: Investigation

David Rivadeneira: Investigation

Sharon Fox: Project administration

Arjun Kandel: Data curation, Writing review & editing

Fatih Ozay: Data curation, Writing review & editing

Desiree Joy Anne Talabong: Data curation, Writing review & editing

Olalekan Lanipekun: Data curation, Writing review & editing

Henry Talus: Resources, Writing review & editing

Jianying Zeng: Investigation, Resources, Writing review & editing

Arvind Rishi: Investigation, Writing review & editing

Nicolas Robine: Conceptualization, Data curation, Methodology, Investigation, Project administration, Supervision, Writing review & editing

Nyasha Chambwe: Conceptualization, Methodology, Project administration, Supervision, Writing review & editing

Jeff Boyd: Conceptualization, Methodology, Project administration, Supervision, Writing review & editing

Alexander Krasnitz: Conceptualization, Formal analysis, Funding acquisition, Methodology, Software, Writing review & editing

Semir Beyaz: Conceptualization, Data curation, Funding acquisition, Methodology, Project administration, Resources, Supervision, Writing review & editing

W. Richard McCombie: Conceptualization, Data Curation, Funding acquisition, Methodology, Project administration, Resources, Supervision, Writing review & editing

Laura A. Martello: Conceptualization, Data curation, Funding acquisition, Project administration, Resources, Supervision, Writing review & editing

All authors reviewed and approved the manuscript.

## References

1. Abel, H.J. et al. (2020) ‘Mapping and characterization of structural variation in 17,795 human genomes’, Nature, 583(7814), pp. 83–89.

2. Adzhubei, I., Jordan, D.M. and Sunyaev, S.R. (2013) ‘Predicting functional effect of human missense mutations using PolyPhen-2’, Current protocols in human genetics, Chapter 7, p. Unit7.20.

3. Aganezov, S. et al. (2020) ‘Comprehensive analysis of structural variants in breast cancer genomes using single-molecule sequencing’, Genome research, 30(9), pp. 1258–1273.

4. Ahn, J. et al. (2013) ‘Human gut microbiome and risk for colorectal cancer’, Journal of the National Cancer Institute, 105(24), pp. 1907–1911.

5. Alexander, D.H. and Lange, K. (2011) ‘Enhancements to the ADMIXTURE algorithm for individual ancestry estimation’, BMC bioinformatics, 12, p. 246.

6. Alexander, D.H., Novembre, J. and Lange, K. (2009) ‘Fast model-based estimation of ancestry in unrelated individuals’, Genome research, 19(9), pp. 1655–1664.

7. Allaire, J.J. et al. (2025) ‘networkD3’ https://archive.linux.duke.edu/cran/web/packages/networkD3/networkD3.pdf (Accessed: 4 November 2025).

8. Amani, M.S. and Peymani, M. (2024) ‘Investigating the impact of SMAD2 and SMAD4 downregulation in colorectal cancer and their correlation with immune markers, prognosis, and drug resistance and sensitivity’, Molecular biology reports, 51(1), p. 831.

9. American Cancer Society. Cancer Facts & Figures 2025. Atlanta: American Cancer Society; 2025.

10. American Cancer Society. Colorectal Cancer Facts & Figures 2023-2025. Atlanta: American Cancer Society; 2023.

11. Arora, K. et al. (2019) ‘Deep whole-genome sequencing of 3 cancer cell lines on 2 sequencing platforms’, Scientific reports, 9(1), p. 19123.

12. Baran, B. et al. (2018) ‘Difference Between Left-Sided and Right-Sided Colorectal Cancer: A Focused Review of Literature’, Gastroenterology research, 11(4), pp. 264–273.

13. Barnett, D.W. et al. (2011) ‘BamTools: a C++ API and toolkit for analyzing and managing BAM files’, Bioinformatics, 27(12), pp. 1691–1692.

14. Belleau, P. et al. (2023) ‘Genetic Ancestry Inference from Cancer-Derived Molecular Data across Genomic and Transcriptomic Platforms’, Cancer research, 83(1), pp. 49–58.

15. Belleau, P. et al. (2025) ‘Inference of Genetic Ancestry from Cancer-Derived Molecular Data with RAIDS’, Methods in molecular biology (Clifton, N.J.), 2932, pp. 153–176.

16. Bolyen, E. et al. (2019) ‘Reproducible, interactive, scalable and extensible microbiome data science using QIIME 2’, Nature biotechnology, 37(8), pp. 852–857.

17. Brown, D.W. et al. (2022) ‘Abstract PO-192: Comparing the association of self-reported race-ethnicity and genetic ancestry with all-cause mortality: A pan-cancer survivor analysis in the PLCO Screening Trial’, in Epidemiology, Lifestyle, and Genetics: Race, Admixture, and Ethnicity. Abstracts: AACR Virtual Conference: 14th AACR Conference on the Science of Cancer Health Disparities in Racial/Ethnic Minorities and the Medically Underserved; October 6-8, 2021, American Association for Cancer Research. Available at: 10.1158/1538-7755.disp21-po-192.

18. Callahan, B.J. et al. (2016) ‘DADA2: High-resolution sample inference from Illumina amplicon data’, Nature methods, 13(7), pp. 581–583.

19. Cancer Genome Atlas Network (2012) ‘Comprehensive molecular characterization of human colon and rectal cancer’, Nature, 487(7407), pp. 330–337.

20. Carrot-Zhang, J. et al. (2020) ‘Comprehensive analysis of genetic ancestry and its molecular correlates in cancer’, Cancer cell, 37(5), pp. 639–654.e6.

21. Carvalho, B. et al. (2009) ‘Multiple putative oncogenes at the chromosome 20q amplicon contribute to colorectal adenoma to carcinoma progression’, Gut, 58(1), pp. 79–89.

22. Catalano, T. et al. (2025) ‘The Role of Reactive Oxygen Species in Colorectal Cancer Initiation and Progression: Perspectives on Theranostic Approaches’, Cancers, 17(5). Available at: 10.3390/cancers17050752.

23. Ciepiela, I. et al. (2024) ‘Tumor location matters, next generation sequencing mutation profiling of left-sided, rectal, and right-sided colorectal tumors in 552 patients’, Scientific reports, 14(1), p. 4619.

24. Colombo, A.P.V. et al. (2009) ‘Comparisons of subgingival microbial profiles of refractory periodontitis, severe periodontitis, and periodontal health using the human oral microbe identification microarray’, Journal of periodontology, 80(9), pp. 1421–1432.

25. Cornish, A.J. et al. (2024) ‘The genomic landscape of 2,023 colorectal cancers’, Nature, 633(8028), pp. 127–136.

26. Corti, G. et al. (2024) ‘Prediction of homologous recombination deficiency identifies colorectal tumors sensitive to PARP inhibition’, NPJ precision oncology, 8(1), p. 231.

27. Crisafulli, G. (2024) ‘Mutational Signatures in Colorectal Cancer: Translational Insights, Clinical Applications, and Limitations’, Cancers, 16(17). Available at: 10.3390/cancers16172956.

28. *dbGaP Study* (2022). Available at: https://www.ncbi.nlm.nih.gov/projects/gap/cgi-bin/study.cgi?study_id=phs000722 (Accessed: 23 September 2025).

29. Degasperi, A. et al. (2022) ‘Substitution mutational signatures in whole-genome-sequenced cancers in the UK population’, Science (New York, N.Y.), 376(6591). Available at: 10.1126/science.abl9283.

30. Deka, J. et al. (2010) ‘Bcl9/Bcl9l are critical for Wnt-mediated regulation of stem cell traits in colon epithelium and adenocarcinomas’, Cancer research, 70(16), pp. 6619–6628.

31. Deschênes, A. et al. (2022) ‘Quantifying similarity between copy number profiles with CNVMetrics package’, F1000Research, 11. Available at: 10.7490/f1000research.1119043.1.

32. Díaz-Gay, M. et al. (2023) ‘Assigning mutational signatures to individual samples and individual somatic mutations with SigProfilerAssignment’, *Bioinformatics (Oxford*, England), 39(12). Available at: 10.1093/bioinformatics/btad756.

33. Dobin, A. et al. (2013) ‘STAR: ultrafast universal RNA-seq aligner’, *Bioinformatics (Oxford*, England), 29(1), pp. 15–21.

34. Ebbert, M.T.W. et al. (2019) ‘Systematic analysis of dark and camouflaged genes reveals disease-relevant genes hiding in plain sight’, Genome biology, 20(1), p. 97.

35. Eide, P.W. et al. (2017) ‘CMScaller: an R package for consensus molecular subtyping of colorectal cancer pre-clinical models’, Scientific reports, 7(1), p. 16618.

36. Eke, P.I., Borgnakke, W.S. and Genco, R.J. (2020) ‘Recent epidemiologic trends in periodontitis in the USA’, Periodontology 2000, 82(1), pp. 257–267.

37. ENCODE Project Consortium (2012) ‘An integrated encyclopedia of DNA elements in the human genome’, Nature, 489(7414), pp. 57–74.

38. Fairley, S. et al. (2020) ‘The International Genome Sample Resource (IGSR) collection of open human genomic variation resources’, Nucleic acids research, 48(D1), pp. D941–D947.

39. Farhana, L. et al. (2018) ‘Gut microbiome profiling and colorectal cancer in African Americans and Caucasian Americans’, World journal of gastrointestinal pathophysiology, 9(2), pp. 47–58.

40. Feng, Q. et al. (2015) ‘Gut microbiome development along the colorectal adenoma-carcinoma sequence’, Nature communications, 6, p. 6528.

41. Field, L.A. et al. (2012) ‘Identification of differentially expressed genes in breast tumors from African American compared with Caucasian women’, Cancer, 118(5), pp. 1334–1344.

42. Flemer, B. et al. (2018) ‘The oral microbiota in colorectal cancer is distinctive and predictive’, Gut, 67(8), pp. 1454–1463.

43. Fleming, N.I. et al. (2013) ‘SMAD2, SMAD3 and SMAD4 mutations in colorectal cancer’, Cancer research, 73(2), pp. 725–735.

44. Friedman, J. and Alm, E.J. (2012) ‘Inferring correlation networks from genomic survey data’, PLoS computational biology, 8(9), p. e1002687.

45. Gay, D.M. et al. (2019) ‘Loss of BCL9/9l suppresses Wnt driven tumourigenesis in models that recapitulate human cancer’, Nature communications, 10(1), p. 723.

46. Gouveia, M.H. et al. (2025) ‘Subcontinental genetic variation in the All of Us Research Program: Implications for biomedical research’, American journal of human genetics, 112(6), pp. 1286–1301.

47. Gragert, L. et al. (2013) ‘Six-locus high resolution HLA haplotype frequencies derived from mixed-resolution DNA typing for the entire US donor registry’, Human immunology, 74(10), pp. 1313–1320.

48. Guinney, J. et al. (2015) ‘The consensus molecular subtypes of colorectal cancer’, Nature medicine, 21(11), pp. 1350–1356.

49. Gulsuner, S. et al. (2024) ‘Long-read DNA and cDNA sequencing identify cancer-predisposing deep intronic variation in tumor-suppressor genes’, Genome research, 34(11), pp. 1825–1831.

50. Gurjao, C. et al. (2024) ‘Abstract 994: Single analyte profiling of the mutational, immune and microbiome landscape in african american colon cancer patients’, Cancer research, 84(6_Supplement), pp. 994–994.

51. Gu, Z., Eils, R. and Schlesner, M. (2016a) ‘Complex heatmaps reveal patterns and correlations in multidimensional genomic data’, Bioinformatics (Oxford, England), 32(18), pp. 2847–2849.

52. Gu, Z., Eils, R. and Schlesner, M. (2016b) ‘gtrellis: an R/Bioconductor package for making genome-level Trellis graphics’, BMC bioinformatics, 17, p. 169.

53. Hänzelmann, S., Castelo, R. and Guinney, J. (2013) ‘GSVA: gene set variation analysis for microarray and RNA-seq data’, BMC bioinformatics, 14, p. 7.

54. Harrow, J. et al. (2012) ‘GENCODE: the reference human genome annotation for The ENCODE Project’, Genome research, 22(9), pp. 1760–1774.

55. Heath, A.P. et al. (2021) ‘The NCI Genomic Data Commons’, Nature genetics, 53(3), pp. 257–262.

56. He, C. et al. (2023) ‘Screening and identifying of biomarkers in early colorectal cancer and adenoma based on genome-wide methylation profiles’, World journal of surgical oncology, 21(1), p. 312.

57. He, Y. et al. (2022) ‘Comprehensive Analysis of Genomic and Expression Data Identified Potential Markers for Predicting Prognosis and Immune Response in CRC’, Genetics research, 2022, p. 1831211.

58. Hoare, A. et al. (2019) ‘Chronic Inflammation as a Link between Periodontitis and Carcinogenesis’, Mediators of inflammation, 2019, p. 1029857.

59. Hoyek, C. et al. (2024) ‘Advances in targeting KRAS G12C in gastrointestinal malignancies: Focus on KRAS G12C inhibitors’, Touch reviews in oncology & haematology, 20(2), pp. 25–33.

60. Hsu, C.W. et al. (2017) ‘How does inflammation drive mutagenesis in colorectal cancer?’, Trends in cancer research, 12, pp. 111–132.

61. Hu, Q. et al. (2019) ‘ZFHX3 is indispensable for ERβ to inhibit cell proliferation via MYC downregulation in prostate cancer cells’, Oncogenesis, 8(4), p. 28.

62. *ICGC ARGO* (2025). Available at: https://platform.icgc-argo.org/ (Accessed: 4 November 2025).

63. Innocenti, F. et al. (2024) ‘DNA Mutational Profiling in Patients With Colorectal Cancer Treated With Standard of Care Reveals Differences in Outcome and Racial Distribution of Mutations’, Journal of clinical oncology : official journal of the American Society of Clinical Oncology, 42(4), pp. 399–409.

64. Jen, J. et al. (1994) ‘Allelic loss of chromosome 18q and prognosis in colorectal cancer’, The New England journal of medicine, 331(4), pp. 213–221.

65. Jiang, X. et al. (2024) ‘A prognostic marker LTBP1 is associated with epithelial mesenchymal transition and can promote the progression of gastric cancer’, Functional & integrative genomics, 24(1), p. 30.

66. Joanito, I. et al. (2022) ‘Single-cell and bulk transcriptome sequencing identifies two epithelial tumor cell states and refines the consensus molecular classification of colorectal cancer’, Nature genetics, 54(7), pp. 963–975.

67. Jovov, B. et al. (2012) ‘Differential gene expression between African American and European American colorectal cancer patients’, PloS one, 7(1), p. e30168.

68. Karczewski, K.J. et al. (2016) ‘The ExAC browser: displaying reference data information from over 60 000 exomes’, Nucleic Acids Research, 45(D1), pp. D840–D845.

69. Karczewski, K.J. et al. (2020) ‘The mutational constraint spectrum quantified from variation in 141,456 humans’, Nature, 581(7809), pp. 434–443.

70. Kirk, S. et al. (2016) ‘The Cancer Genome Atlas Colon Adenocarcinoma Collection (TCGA-COAD)’. The Cancer Imaging Archive. Available at: 10.7937/K9/TCIA.2016.HJJHBOXZ.

71. Klos, C.L. and Dharmarajan, S. (2016) ‘Polyp Genetics’, Clinics in colon and rectal surgery, 29(4), pp. 289–295.

72. Koboldt, D.C. et al. (2012) ‘VarScan 2: Somatic mutation and copy number alteration discovery in cancer by exome sequencing’, Genome Research, 22(3), pp. 568–576.

73. Koliarakis, I. et al. (2019) ‘Oral Bacteria and Intestinal Dysbiosis in Colorectal Cancer’, International journal of molecular sciences, 20(17). Available at: 10.3390/ijms20174146.

74. Koufaris, C. and Kirmizis, A. (2020) ‘N-Terminal Acetyltransferases Are Cancer-Essential Genes Prevalently Upregulated in Tumours’, Cancers, 12(9). Available at: 10.3390/cancers12092631.

75. Krasnitz, A. et al. (2013) ‘Target inference from collections of genomic intervals’, Proceedings of the National Academy of Sciences of the United States of America, 110(25), pp. E2271–8.

76. Kurtz, Z.D. et al. (2015) ‘Sparse and compositionally robust inference of microbial ecological networks’, PLoS computational biology, 11(5), p. e1004226.

77. Landrum, M.J. et al. (2015) ‘ClinVar: public archive of interpretations of clinically relevant variants’, Nucleic Acids Research, 44(D1), pp. D862–D868.

78. Lawler, T., Parlato, L. and Warren Andersen, S. (2024) ‘Racial disparities in colorectal cancer clinicopathological and molecular tumor characteristics: a systematic review’, Cancer causes & control : CCC, 35(2), pp. 223–239.

79. Lawrence, M. et al. (2013) ‘Software for computing and annotating genomic ranges’, PLoS computational biology, 9(8), p. e1003118.

80. Lawrence, M.S. et al. (2013) ‘Mutational heterogeneity in cancer and the search for new cancer-associated genes’, Nature, 499(7457), pp. 214–218.

81. Lee, H. and Kingsford, C. (2018) ‘Kourami: graph-guided assembly for novel human leukocyte antigen allele discovery’, Genome biology, 19(1), p. 16.

82. Lee, K.S. et al. (2015) ‘c-MYC Copy-Number Gain Is an Independent Prognostic Factor in Patients with Colorectal Cancer’, PloS one, 10(10), p. e0139727.

83. Lewis, A.C.F. et al. (2022) ‘Getting genetic ancestry right for science and society’, Science (New York, N.Y.), 376(6590), pp. 250–252.

84. Liao, Y., Smyth, G.K. and Shi, W. (2014) ‘featureCounts: an efficient general purpose program for assigning sequence reads to genomic features’, *Bioinformatics (Oxford*, England), 30(7), pp. 923–930.

85. Liberzon, A. et al. (2015) ‘The Molecular Signatures Database (MSigDB) hallmark gene set collection’, Cell systems, 1(6), pp. 417–425.

86. Li, H. et al. (2009) ‘The Sequence Alignment/Map format and SAMtools’, Bioinformatics, 25(16), pp. 2078–2079.

87. Li, H. (2013) ‘Aligning sequence reads, clone sequences and assembly contigs with BWA-MEM’. Available at: http://arxiv.org/abs/1303.3997 (Accessed: 25 September 2025).

88. Li, H. et al. (2015) ‘Circulating Prostaglandin Biosynthesis in Colorectal Cancer and Potential Clinical Significance’, EBioMedicine, 2(2), pp. 165–171.

89. Lin, H. and Peddada, S.D. (2020) ‘Analysis of compositions of microbiomes with bias correction’, Nature communications, 11(1), p. 3514.

90. Liu, Y. et al. (2020) ‘Identification and verification of three key genes associated with survival and prognosis of COAD patients via integrated bioinformatics analysis’, Bioscience reports, 40(9). Available at: 10.1042/BSR20200141.

91. Love, M.I., Huber, W. and Anders, S. (2014) ‘Moderated estimation of fold change and dispersion for RNA-seq data with DESeq2’, Genome biology, 15(12), p. 550.

92. Lowe, D.G. (1999) ‘Object recognition from local scale-invariant features’, Proceedings of the Seventh IEEE International Conference on Computer Vision, pp. 1150–1157 vol.2, doi: 10.1109/ICCV.1999.790410.

93. Lu, J. et al. (2017) ‘Bracken: estimating species abundance in metagenomics data’, PeerJ. Computer science, 3. Available at: 10.7717/peerj-cs.104.

94. Lu, J. et al. (2022) ‘Metagenome analysis using the Kraken software suite’, Nature protocols, 17(12), pp. 2815–2839.

95. Luo, M. et al. (2022) ‘Race is a key determinant of the human intratumor microbiome’, Cancer cell, 40(9), pp. 901–902.

96. Mahmoud, M. et al. (2024) ‘Utility of long-read sequencing for All of Us’, Nature Communications, 15(1), pp. 1–13.

97. Martini, R. et al. (2022) ‘African Ancestry-Associated Gene Expression Profiles in Triple-Negative Breast Cancer Underlie Altered Tumor Biology and Clinical Outcome in Women of African Descent’, Cancer discovery, 12(11), pp. 2530–2551.

98. Matejcic, M. et al. (2025) ‘Colorectal Tumors in Diverse Patient Populations Feature a Spectrum of Somatic Mutational Profiles’, Cancer research, 85(10), pp. 1928–1944.

99. Mayakonda, A. et al. (2018) ‘Maftools: efficient and comprehensive analysis of somatic variants in cancer’, Genome research, 28(11), pp. 1747–1756.

100. McMurdie, P.J. and Holmes, S. (2013) ‘phyloseq: an R package for reproducible interactive analysis and graphics of microbiome census data’, PloS one, 8(4), p. e61217.

101. Miller, D.E. et al. (2021) ‘Targeted long-read sequencing identifies missing disease-causing variation’, American journal of human genetics, 108(8), pp. 1436–1449.

102. M.k, V. and K, K. (2016) ‘A survey on similarity measures in text mining’, Machine Learning and Applications An International Journal, 3(1), pp. 19–28.

103. Mohammadpour, S. et al. (2024) ‘Hippo Signaling Pathway in Colorectal Cancer: Modulation by Various Signals and Therapeutic Potential’, Analytical cellular pathology (Amsterdam), 2024, p. 5767535.

104. Momen-Heravi, F. et al. (2017) ‘Periodontal disease, tooth loss and colorectal cancer risk: Results from the Nurses’ Health Study’, International journal of cancer, 140(3), pp. 646–652.

105. Myer, P.A. et al. (2022) ‘The Genomics of Colorectal Cancer in Populations with African and European Ancestry’, Cancer discovery, 12(5), pp. 1282–1293.

106. Nambara, S. et al. (2020) ‘GTF2IRD1 on chromosome 7 is a novel oncogene regulating the tumor-suppressor gene TGFβR2 in colorectal cancer’, Cancer science, 111(2), pp. 343–355.

107. Nattestad, M. et al. (2018) ‘Complex rearrangements and oncogene amplifications revealed by long-read DNA and RNA sequencing of a breast cancer cell line’, Genome research, 28(8), pp. 1126–1135.

108. Newman, A.M. et al. (2015) ‘Robust enumeration of cell subsets from tissue expression profiles’, Nature methods, 12(5), pp. 453–457.

109. NYGC (2019) Available at: https://bioinformatics.nygenome.org/wp-content/uploads/2019/06/SomaticPipeline_v6.0_Human_WGS.pdf (Accessed: 25 September 2025).

110. Nunes, L. et al. (2024) ‘Prognostic genome and transcriptome signatures in colorectal cancers’, Nature, 633(8028), pp. 137–146.

111. Pandey, P. et al. (2019) ‘Mutational signatures in colon cancer’, BMC research notes, 12(1), p. 788.

112. Paredes, J. et al. (2020) ‘Immune-Related Gene Expression and Cytokine Secretion Is Reduced Among African American Colon Cancer Patients’, Frontiers in oncology, 10, p. 1498.

113. *Picard toolkit* (2019). Available at: https://broadinstitute.github.io/picard/ (Accessed: 25 September 2025)

114. Quang, D., Chen, Y. and Xie, X. (2014) ‘DANN: a deep learning approach for annotating the pathogenicity of genetic variants’, Bioinformatics, 31(5), pp. 761–763.

115. Quast, C. et al. (2013) ‘The SILVA ribosomal RNA gene database project: improved data processing and web-based tools’, Nucleic acids research, 41(Database issue), pp. D590–6.

116. Rentzsch, P. et al. (2018) ‘CADD: predicting the deleteriousness of variants throughout the human genome’, Nucleic Acids Research, 47(D1), pp. D886–D894.

117. Robine, N. and Varmus, H. (2022) ‘New York’s Polyethnic-1000: a regional initiative to understand how diverse ancestries influence the risk, progression, and treatment of cancers’, Trends in cancer, 8(4), pp. 269–272.

118. Schwarz, J.M. et al. (2010) ‘MutationTaster evaluates disease-causing potential of sequence alterations’, Nature methods, 7(8), pp. 575–576.

119. Shen, R. and Seshan, V.E. (2016) ‘FACETS: allele-specific copy number and clonal heterogeneity analysis tool for high-throughput DNA sequencing’, Nucleic acids research, 44(16), p. e131.

120. Soltis, A.R. et al. (2020) ‘MutEnricher: a flexible toolset for somatic mutation enrichment analysis of tumor whole genomes’, BMC bioinformatics, 21(1), p. 338.

121. Sondka, Z. et al. (2024) ‘COSMIC: a curated database of somatic variants and clinical data for cancer’, Nucleic acids research, 52(D1), pp. D1210–D1217.

122. Staudacher, J.J. et al. (2017) ‘Increased Frequency of KRAS Mutations in African Americans Compared with Caucasians in Sporadic Colorectal Cancer’, Clinical and translational gastroenterology, 8(10), p. e124.

123. Subramanian, A. et al. (2005) ‘Gene set enrichment analysis: a knowledge-based approach for interpreting genome-wide expression profiles’, Proceedings of the National Academy of Sciences of the United States of America, 102(43), pp. 15545–15550.

124. Sudmant, P.H. et al. (2015) ‘An integrated map of structural variation in 2,504 human genomes’, Nature, 526(7571), pp. 75–81.

125. Sylvester, B.E. et al. (2012) ‘Molecular analysis of colorectal tumors within a diverse patient cohort at a single institution’, Clinical cancer research : an official journal of the American Association for Cancer Research, 18(2), pp. 350–359.

126. Tate, J.G. et al. (2019) ‘COSMIC: the Catalogue Of Somatic Mutations In Cancer’, Nucleic acids research, 47(D1). Available at: 10.1093/nar/gky1015.

127. Thanki, K. et al. (2017) ‘Consensus Molecular Subtypes of Colorectal Cancer and their Clinical Implications’, International biological and biomedical journal, 3(3), pp. 105–111.

128. *The Cancer Genome Atlas Program (TCGA)* (2022). Available at: https://www.cancer.gov/ccg/research/genome-sequencing/tcga (Accessed: 25 September 2025).

129. Tortora, S.C. et al. (2022) ‘Microbiome and colorectal carcinogenesis: Linked mechanisms and racial differences’, World journal of gastrointestinal oncology, 14(2), pp. 375–395.

130. Tričković, M. et al. (2025) ‘Subspecies of the human gut microbiota carry implicit information for in-depth microbiome research’, Cell host & microbe, 33(8), pp. 1446–1458.e4.

131. Van der Auwera, G.A. and O’Connor, B.D. (2020) *Genomics in the Cloud: Using Docker, GATK, and WDL in Terra*. ‘O’Reilly Media, Inc.’

132. Van Loo, P. et al. (2010) ‘Allele-specific copy number analysis of tumors’, Proceedings of the National Academy of Sciences of the United States of America, 107(39), pp. 16910–16915.

133. Vaughan-Shaw, P.G. et al. (2021) ‘Differential genetic influences over colorectal cancer risk and gene expression in large bowel mucosa’, International journal of cancer, 149(5), pp. 1100–1108.

134. Wallace, T.A. et al. (2008) ‘Tumor immunobiological differences in prostate cancer between African-American and European-American men’, Cancer research, 68(3), pp. 927–936.

135. Wang, K., Li, M. and Hakonarson, H. (2010) ‘ANNOVAR: functional annotation of genetic variants from high-throughput sequencing data’, Nucleic acids research, 38(16), p. e164.

136. Wang, L. et al. (2020) ‘ZBTB7A functioned as an oncogene in colorectal cancer’, BMC gastroenterology, 20(1), p. 370.

137. Wang, L., Wang, S. and Li, W. (2012) ‘RSeQC: quality control of RNA-seq experiments’, *Bioinformatics (Oxford*, England), 28(16), pp. 2184–2185.

138. Wang, X. et al. (2024) ‘Additional impact of genetic ancestry over race/ethnicity to prevalence of KRAS mutations and allele-specific subtypes in non-small cell lung cancer’, HGG advances, 5(3), p. 100320.

139. Wang, Y.-C. et al. (2022) ‘NPRL2 down-regulation facilitates the growth of hepatocellular carcinoma via the mTOR pathway and autophagy suppression’, Hepatology communications, 6(12), pp. 3563–3577.

140. Wang, Z. et al. (2017) ‘Down-regulation of LRP1B in colon cancer promoted the growth and migration of cancer cells’, Experimental cell research, 357(1), pp. 1–8.

141. Wickham, H. (2016). ggplot2: Elegant Graphics for Data Analysis. Springer-Verlag New York. ISBN 978-3-319-24277-4 Springer International Publishing.

142. Wilke, C.O. (2015) ‘cowplot: Streamlined Plot Theme and Plot Annotations for “ggplot2”’, CRAN: Contributed Packages [Preprint]. Available at: 10.32614/cran.package.cowplot.

143. Wolff, R.K. et al. (2018) ‘Mutation analysis of adenomas and carcinomas of the colon: Early and late drivers’, Genes, chromosomes & cancer, 57(7), pp. 366–376.

144. Wood, D.E., Lu, J. and Langmead, B. (2019) ‘Improved metagenomic analysis with Kraken 2’, Genome biology, 20(1), p. 257.

145. Wu, D. et al. (2017) limma. Bioconductor. Available at: 10.18129/B9.BIOC.LIMMA.

146. Xu, M. et al. (2022) ‘AHNAK2 is a biomarker and a potential therapeutic target of adenocarcinomas’, Acta biochimica et biophysica Sinica, 54(11), pp. 1708–1719.

147. Yan, L. (2023) ‘Draw Venn Diagram by “ggplot2” [R package ggvenn version 0.1.10]’. Available at: https://CRAN.R-project.org/package=ggvenn (Accessed: 25 September 2025).

148. Yu, T. et al. (2025) ‘Research Progress on the Role of Zinc Finger Protein in Colorectal Cancer’, Cancer reports (Hoboken, N.J.), 8(3), p. e70123.

149. Zhang, J. et al. (2019) ‘The International Cancer Genome Consortium Data Portal’, Nature Biotechnology, 37(4), pp. 367–369.

150. Zhang, R. and Song, C. (2014) ‘Loss of CSMD1 or 2 may contribute to the poor prognosis of colorectal cancer patients’, Tumour biology: the journal of the International Society for Oncodevelopmental Biology and Medicine, 35(5), pp. 4419–4423.

151. Zhu, R. et al. (2024) ‘The role of N-acetyltransferases in cancers’, Gene, 892, p. 147866.

