## Supplemental Figures and Tables for "Integrative Genomic, Transcriptomic, and Microbiome Profiles of Colon Cancer by Ancestry Provide Insights into Molecular Distinctions"

**Supplemental Materials**

**Figure S1.** Relation between race and ancestry in the NYGC P-1000 cohort (**A**) and the CSHL FFPE cohort (**B**) when a cut-off of 70% is used to assign continental ancestry. The number of profiles (n) is indicated for each class. Race/ethnicity is on the left side of the graph, while assigned ancestry based on genetic ancestry is on the right side. For the assigned continental ancestry: AFR (African), EUR (European), EAS (East Asian), and Admixed (less than 70% continental ancestry). Distribution of the ratio of the AFR and EUR ancestry in the patients assigned to the EURg and AFRg groups in the NYGC P-1000 cohort (**C**) and the CSHL FFPE cohort (**D**).

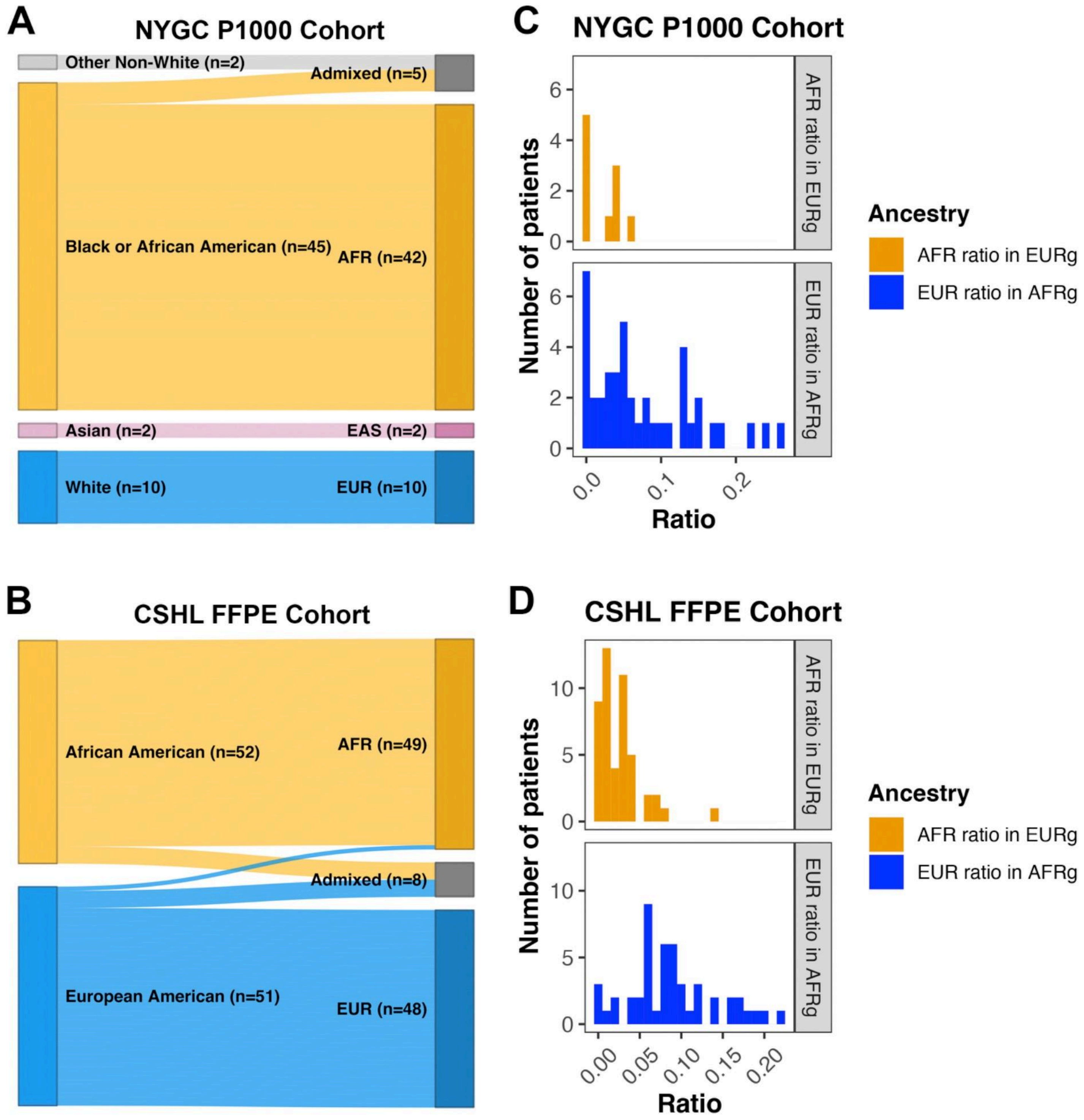

**Figure S2.** Density distribution of some sub-continental ancestries related to African continental ancestry for the African-related profiles (at least 70% African continental ancestry) in the CSHL FFPE cohort (**A**), in the P-1000 cohort (**B**), and in TGCA (**C**). The sub-continental populations represented in the graph are Nigeria (ESN), Gambian in Western Division, The Gambia—Mandinka (GWD), Luhya in Webuye, Kenya (LWK), Sierra Leone (MSL), and Yoruba in Ibadan, Nigeria (YRI).

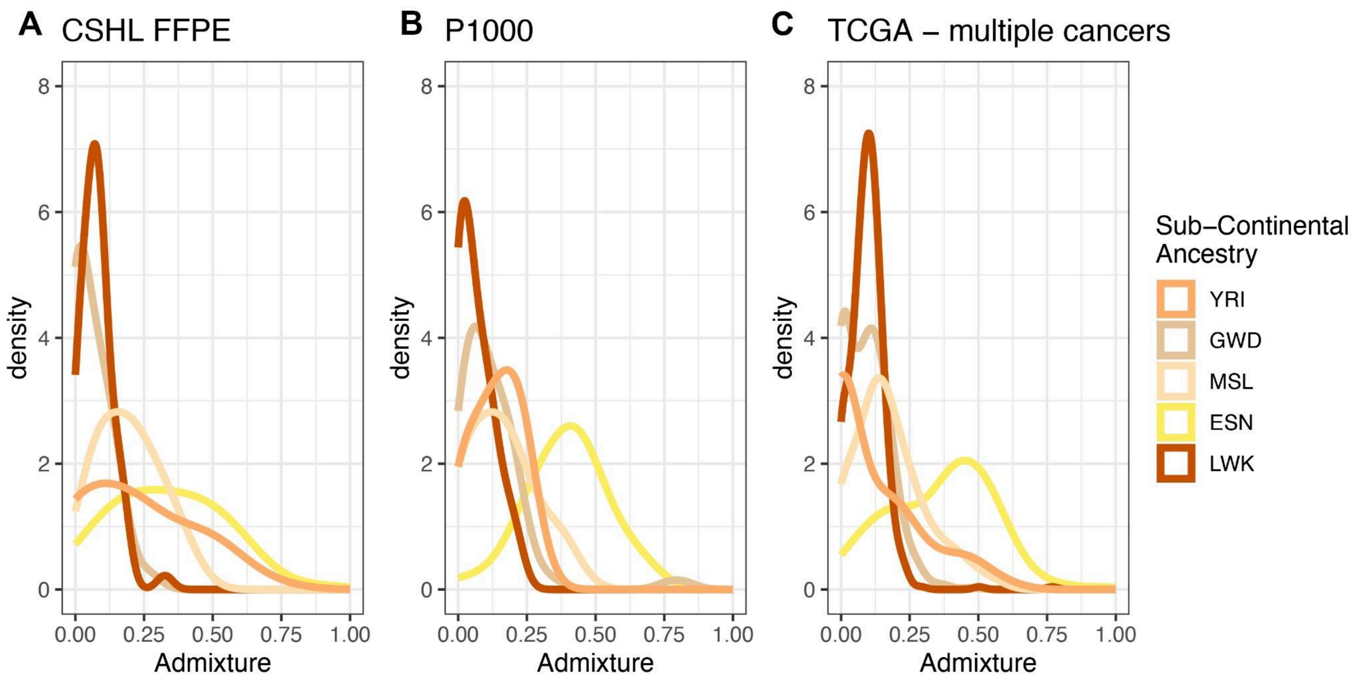

**Figure S3. A)** Summary of putative somatic variants in the FFPE exome cohort samples (MSS only) as determined by our prioritization pipeline, including variant classifications, counts per sample, and top mutated genes. **B)** Summary of putative somatic variants in the P-1000 exome cohort samples (including both MSS and MSI-H) as determined by our prioritization pipeline, including variant classifications, counts per sample, and top mutated genes.

**A**

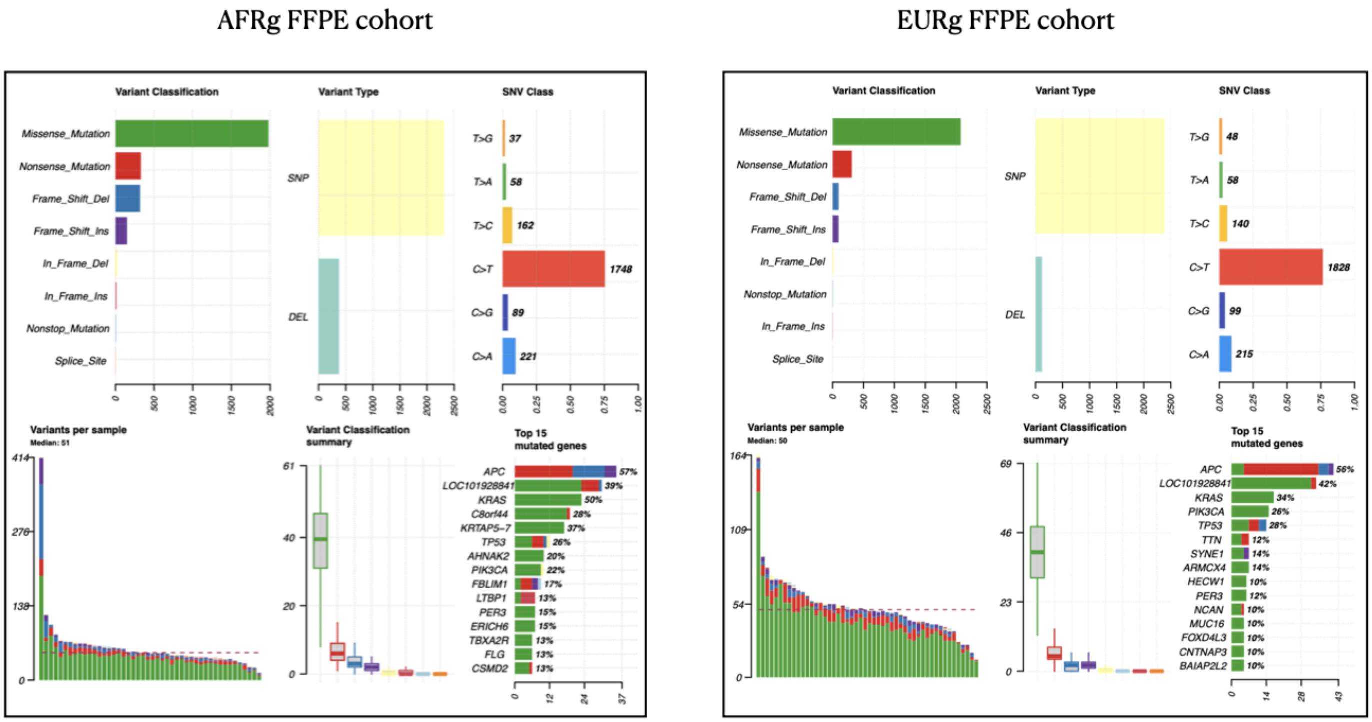

**B**

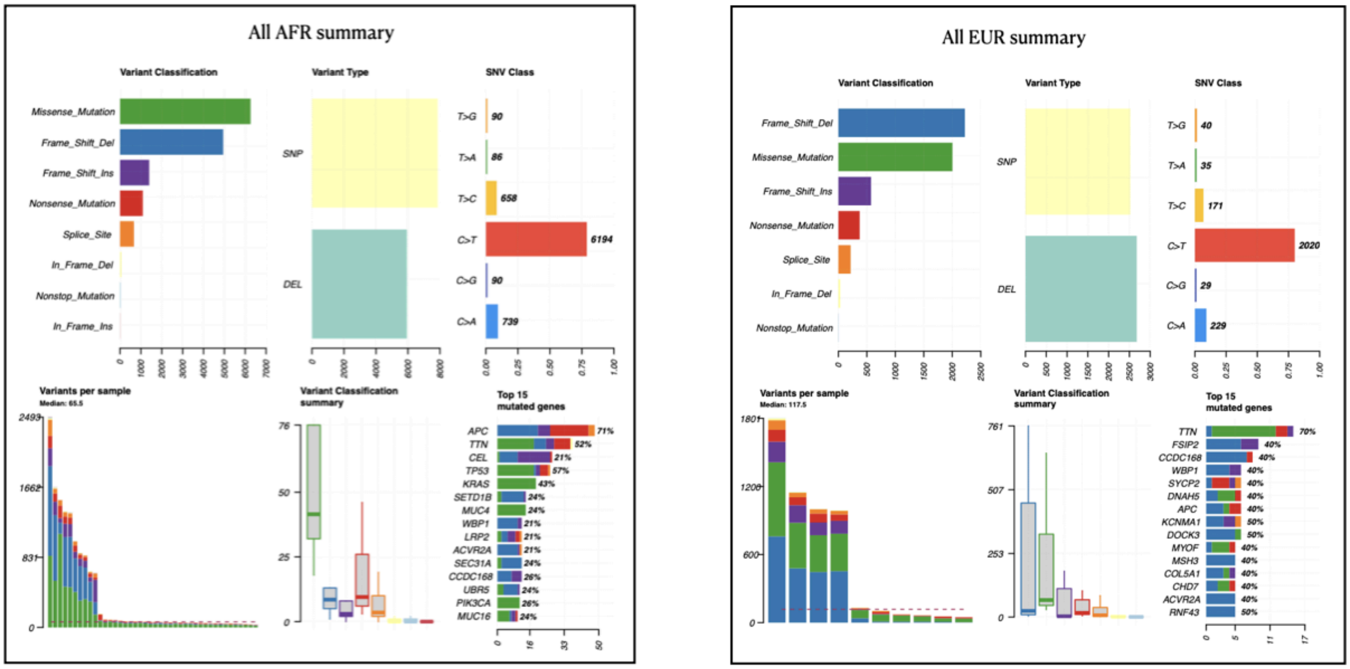

**Figure S4.** P-1000 WGS tumor mutational load (TMB) per megabase compared to TCGA cohorts.

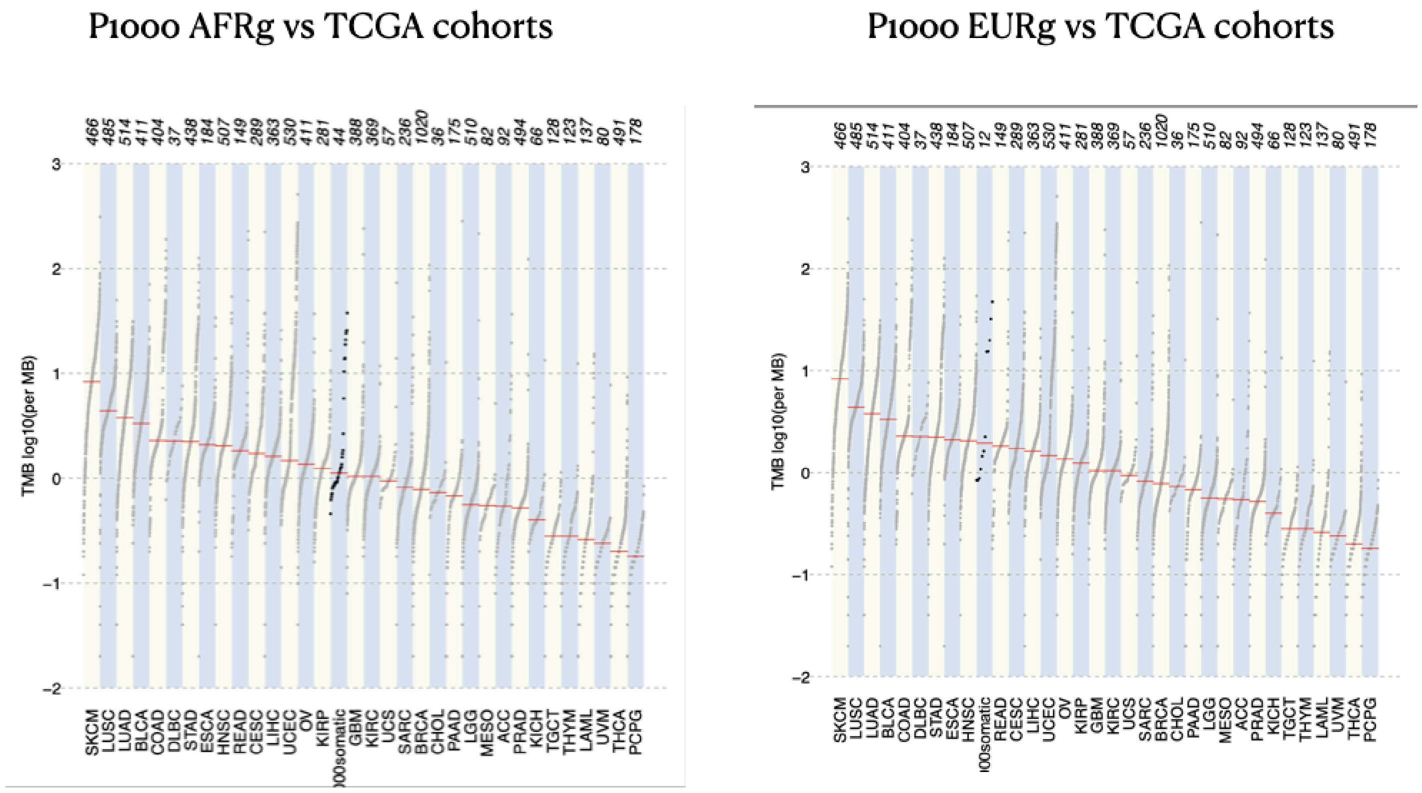

**Figure S5. A)** FFPE exome cohort tumors somatic variant comparison for AFRg vs EURg groups showing suggestive enrichment of mutation frequency (number of individuals per cohort with mutations in each gene) between cohorts in genes with uncorrected p-value less than 0.05 (*), 0.01 (**), or 0.001(***). **B**) Oncoplots for somatic mutations in AFRg and EURg polyps, showing tubular adenomas on the left and sessile serrated polyps on the right in each plot.

**A**

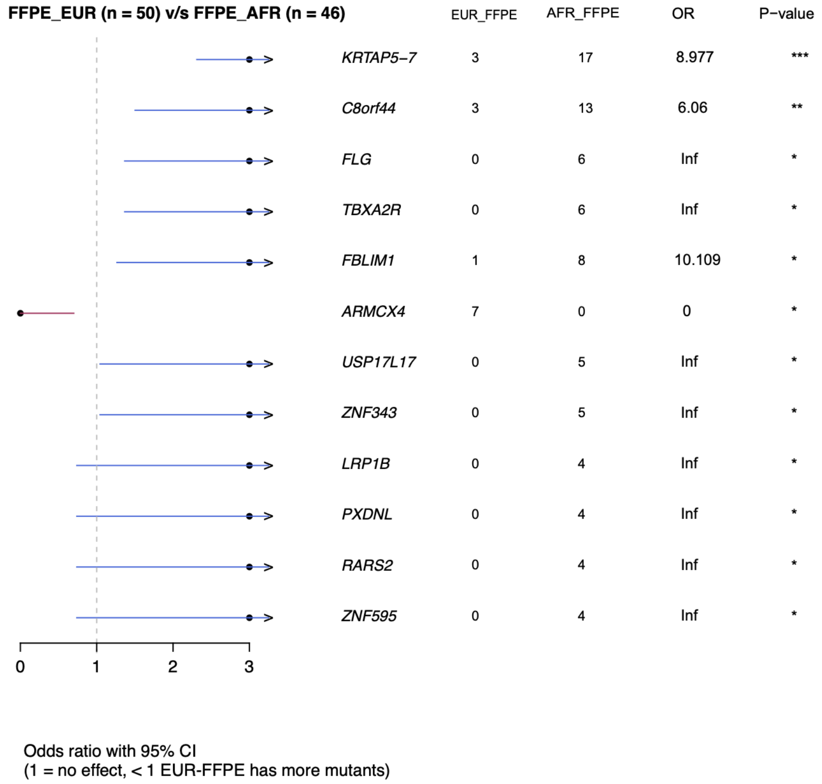

**B**

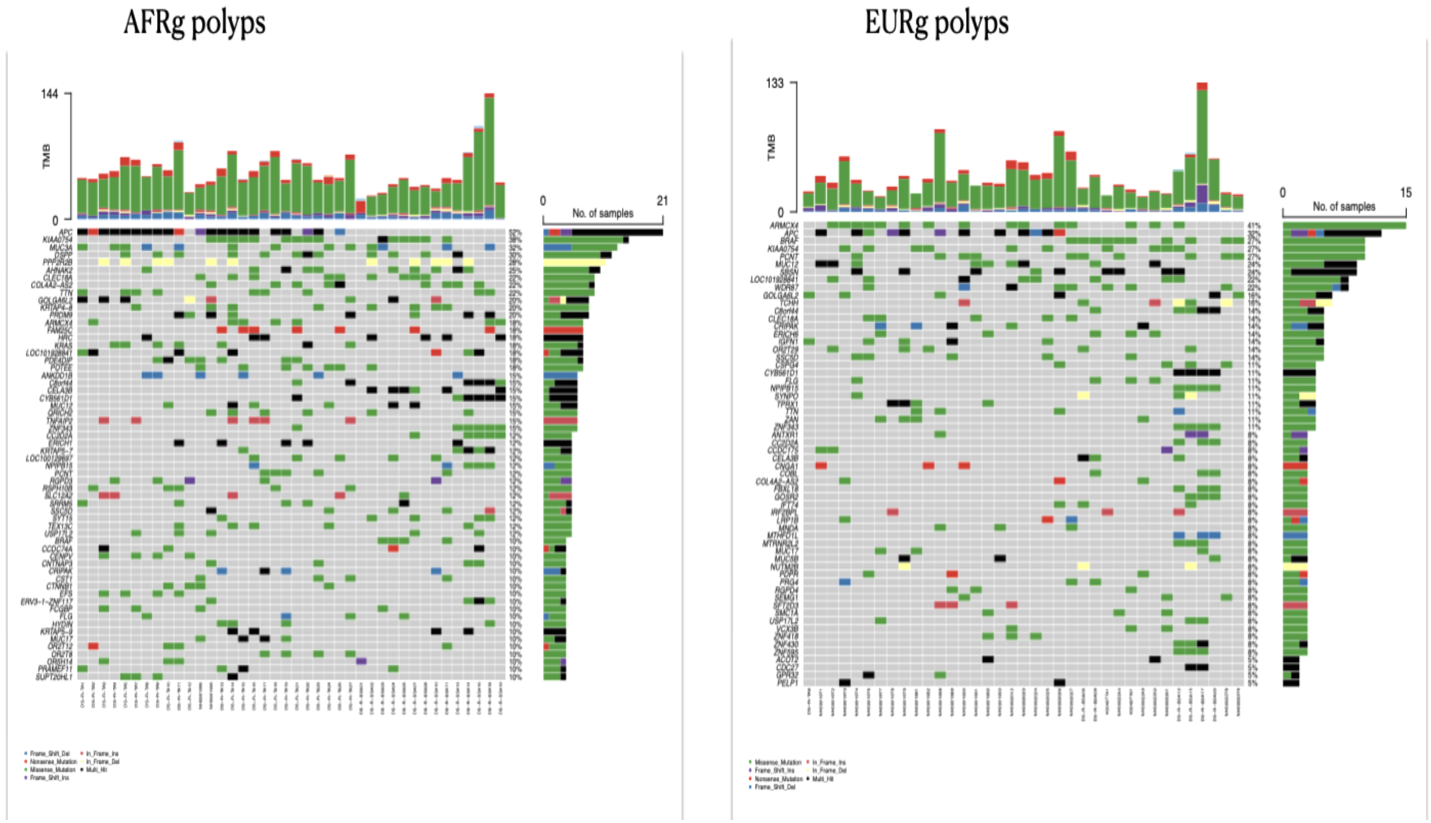

**Figure S6.** Lollipop plots showing the spectrum of KRAS mutations found in the P-1000 and FFPE exome microsatellite stable AFRg and EURg cohorts.

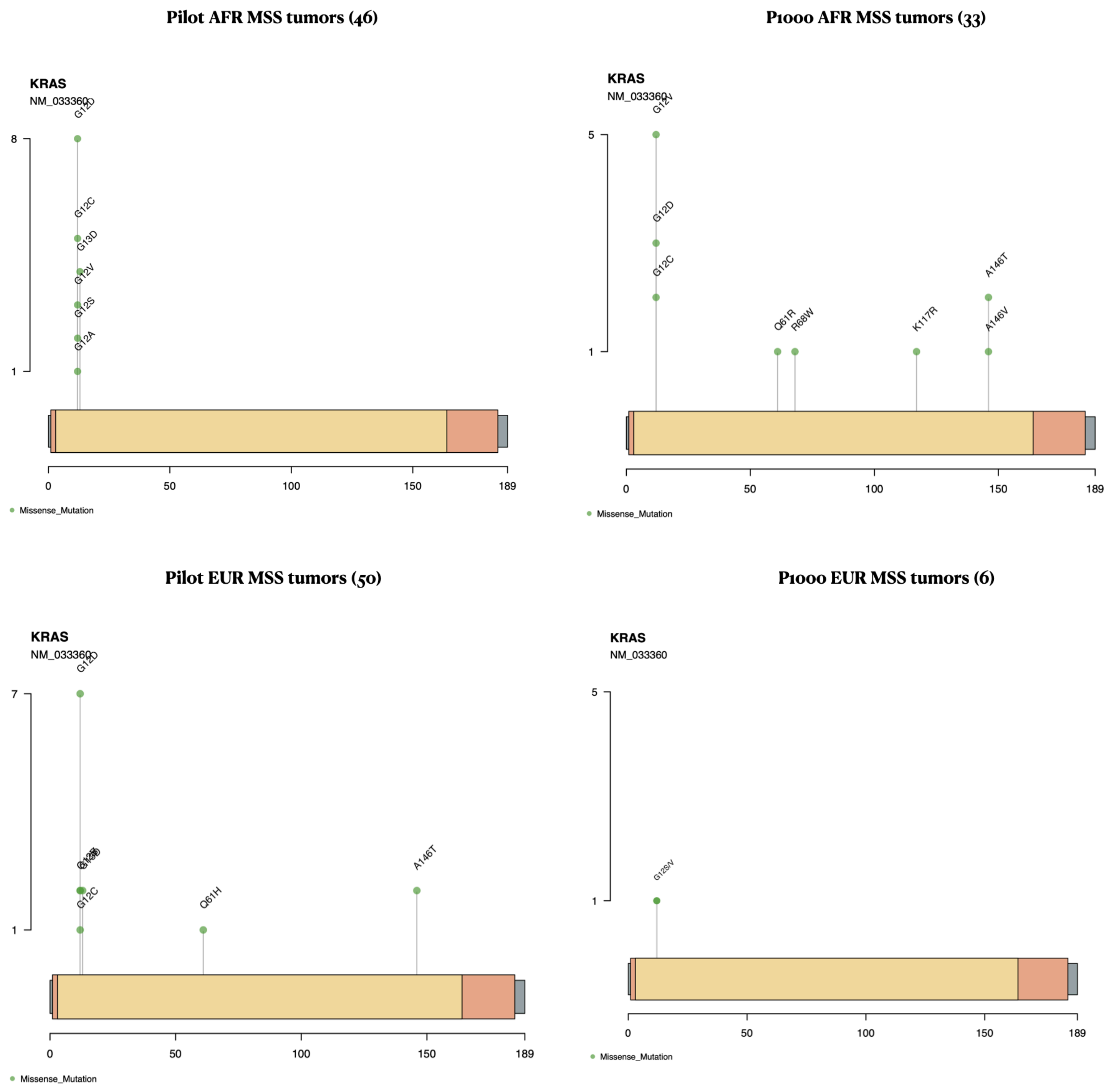

**Figure S7.** P-1000 WGS full cohort comparisons (both MSS and MSI-H, not stratified by ancestry) for tumor laterality (left vs right), smoking status (current, reformed, or never), and sex (male vs female). Plots show counts of mutations in genes reaching a p-value of 0.05.

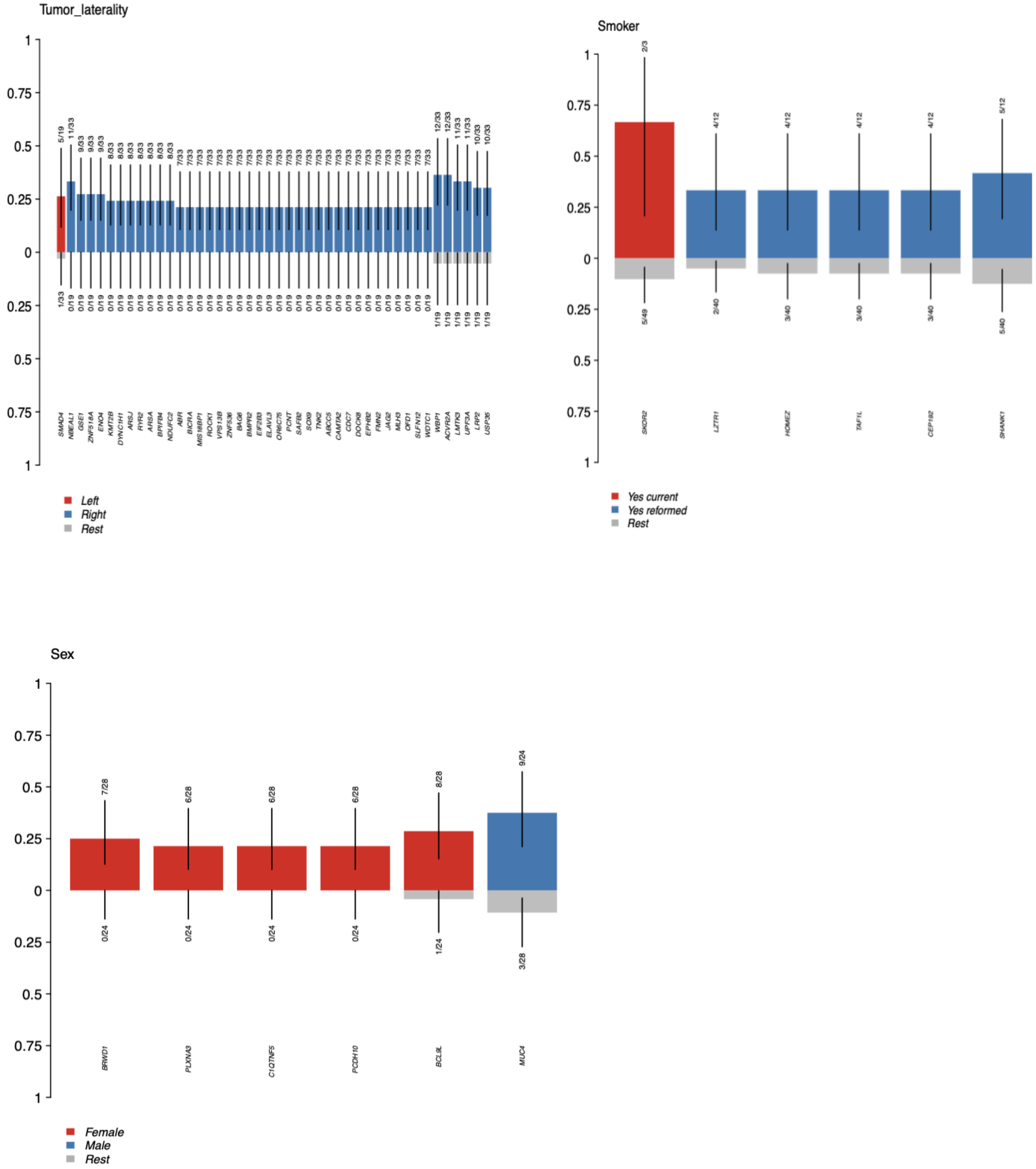

**Figure S8.** Mutational signatures of the P-1000 cohort, corresponding to known COSMIC signatures, split by ancestry, including **A)** single base substitutions (SBS), **B)** doublet base substitutions (DBS) and **C)** small insertions and deletions (ID). **D)** Boxplots for signature SBS1 versus age at diagnosis. **E)** Boxplots for MMR deficiency signatures by tumor microsatellite stability.

**A**

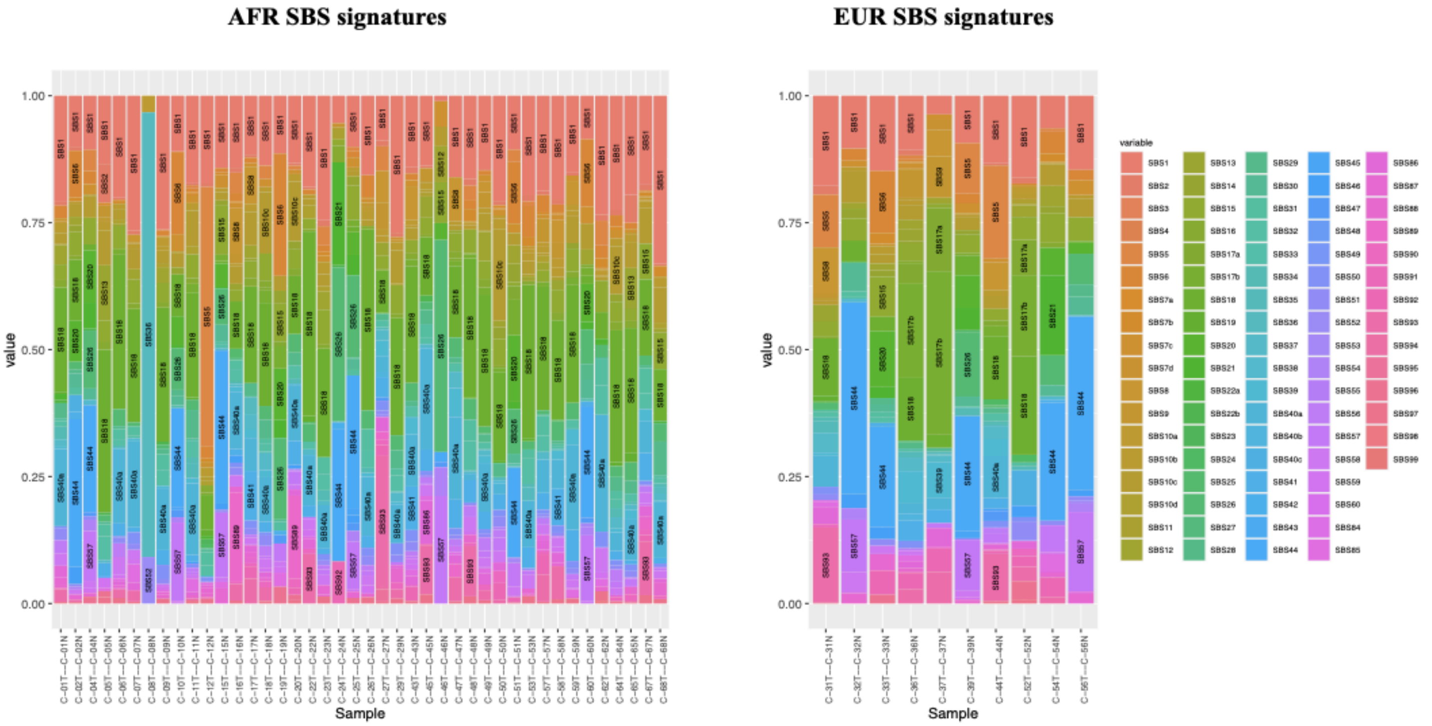

**B**

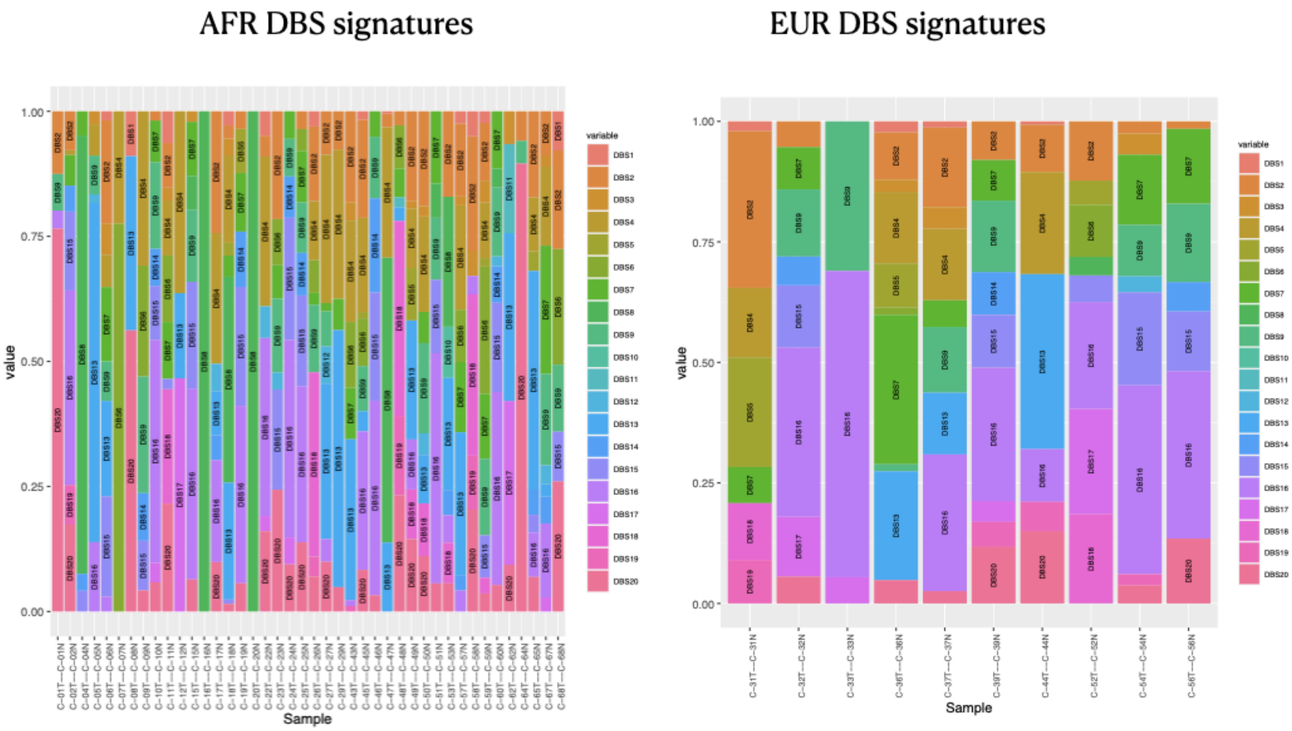

AFR DBS signatures
EUR DBS signatures
value
Sample
1.00
0.75
0.50
0.25
0.00
variable
DBS1
DBS2
DBS3
DBS4
DBS5
DBS6
DBS7
DBS8
DBS9
DBS10
DBS11
DBS12
DBS13
DBS14
DBS15
DBS16
DBS17
DBS18
DBS19
DBS20

**C**

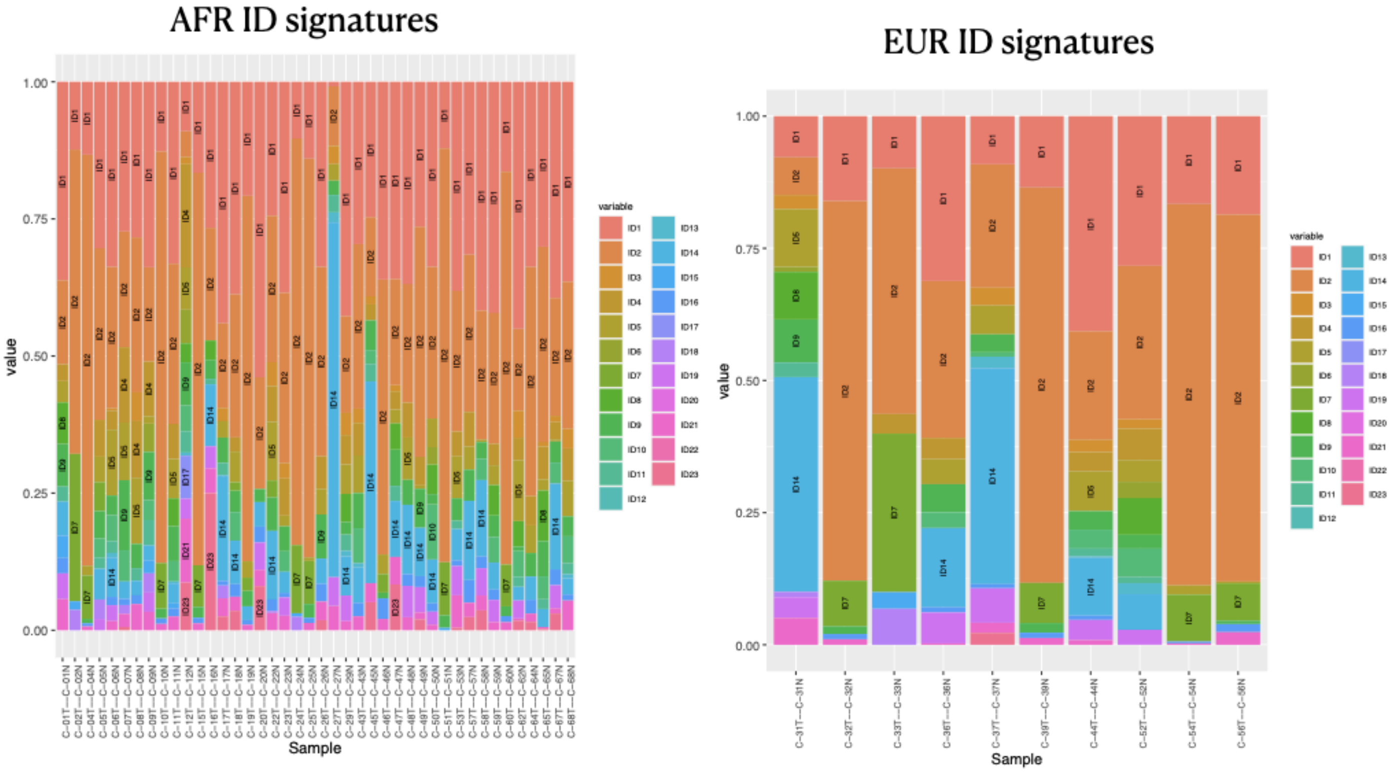

AFR ID signatures
EUR ID signatures
value
Sample
1.00
0.75
0.50
0.25
0.00
variable
ID1
ID2
ID3
ID4
ID5
ID6
ID7
ID8
ID9
ID10
ID11
ID12
ID13
ID14
ID15
ID16
ID17
ID18
ID19
ID20
ID21
ID22
ID23

**D**

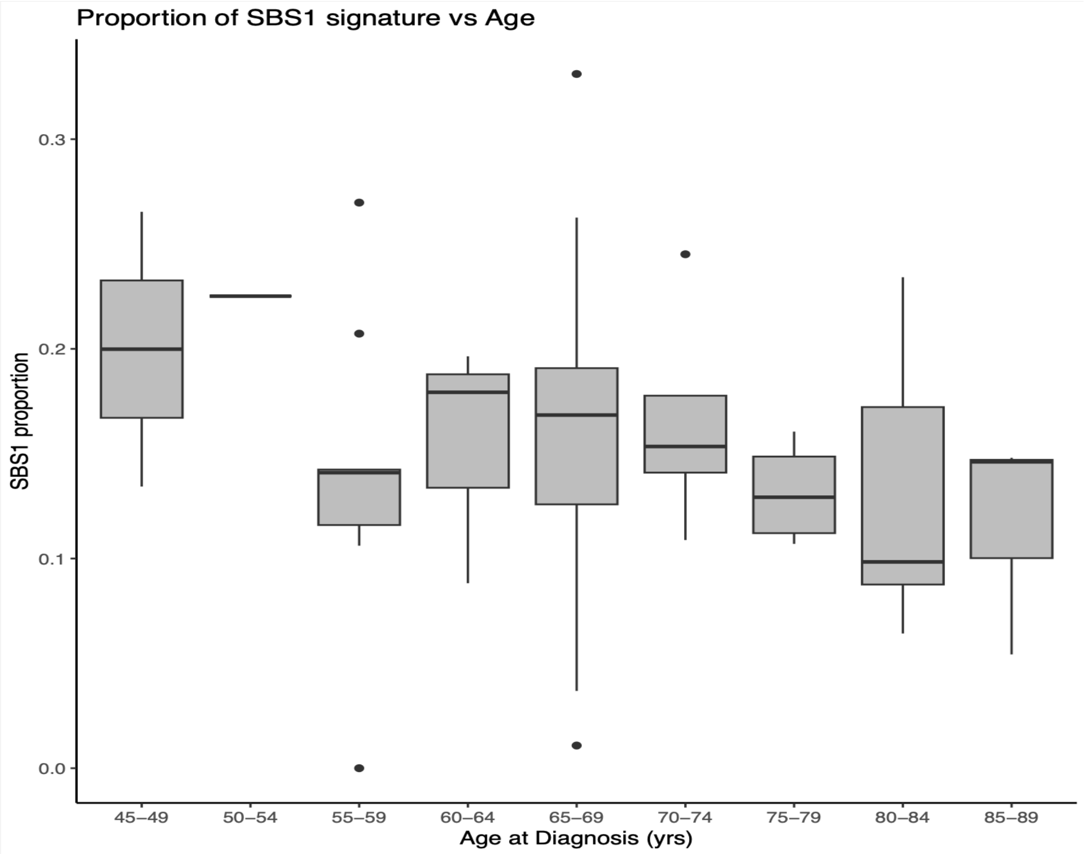

Proportion of SBS1 signature vs Age
SBS1 proportion
0.0
0.1
0.2
0.3
45–49
50–54
55–59
60–64
65–69
70–74
75–79
80–84
85–89
Age at Diagnosis (yrs)

**E**

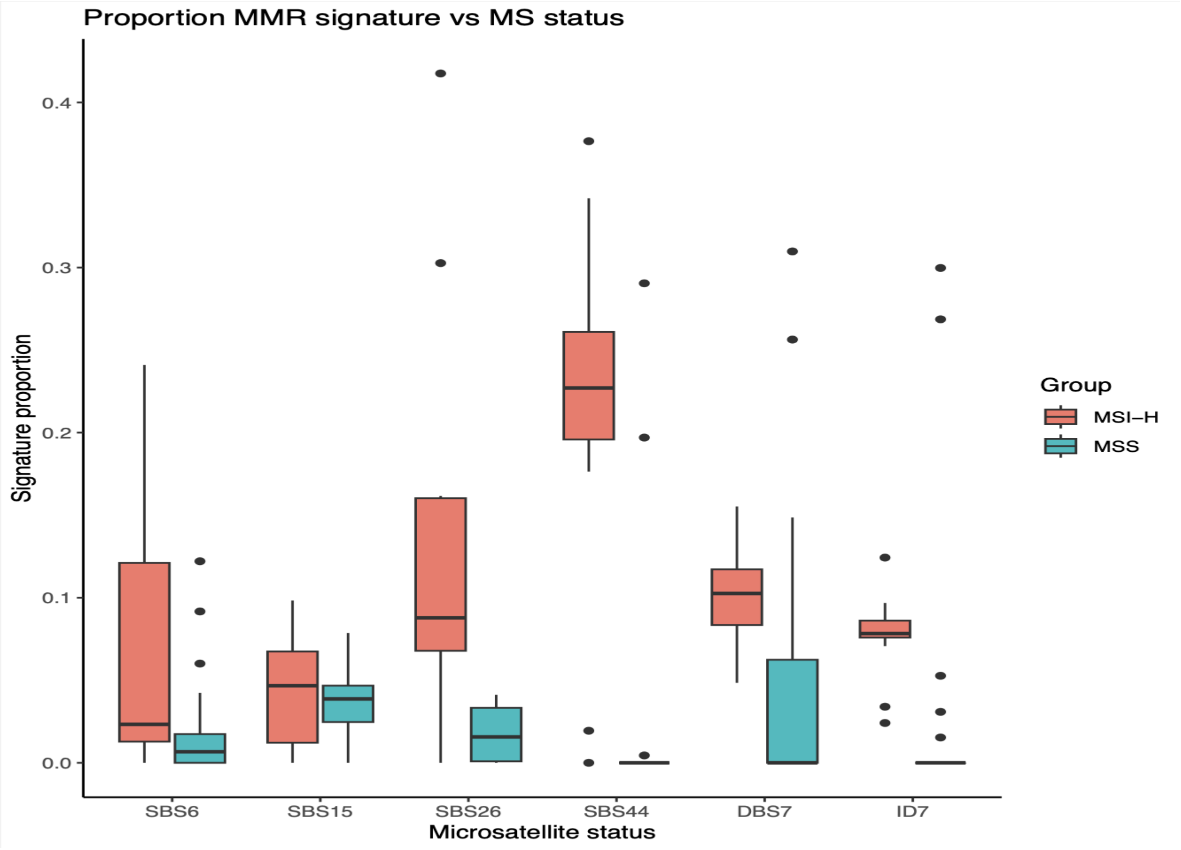

Proportion MMR signature vs MS status
Signature proportion
0.0
0.1
0.2
0.3
0.4
SBS6
SBS15
SBS26
SBS44
DBS7
ID7
Microsatellite status
Group
MSI–H
MSS

**Figure S9.** Expression counts by ancestral African fraction (AFR %) for gene NPRL3 in (**A**) the P-1000 cohort and in (**B**) TCGA-COAD.

**A**

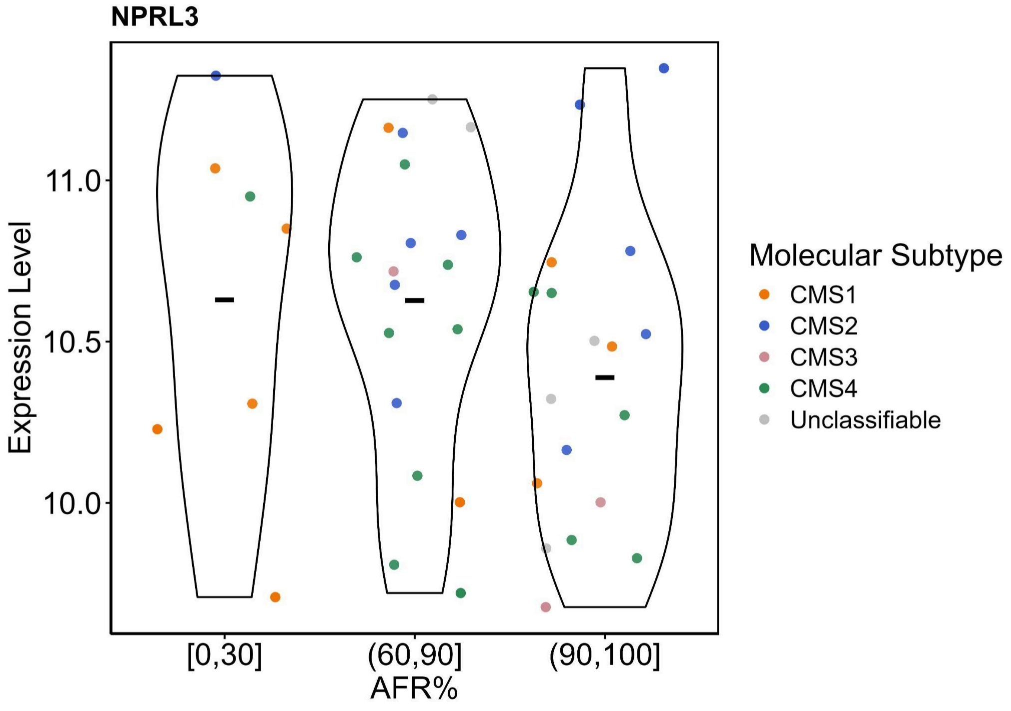

**B**

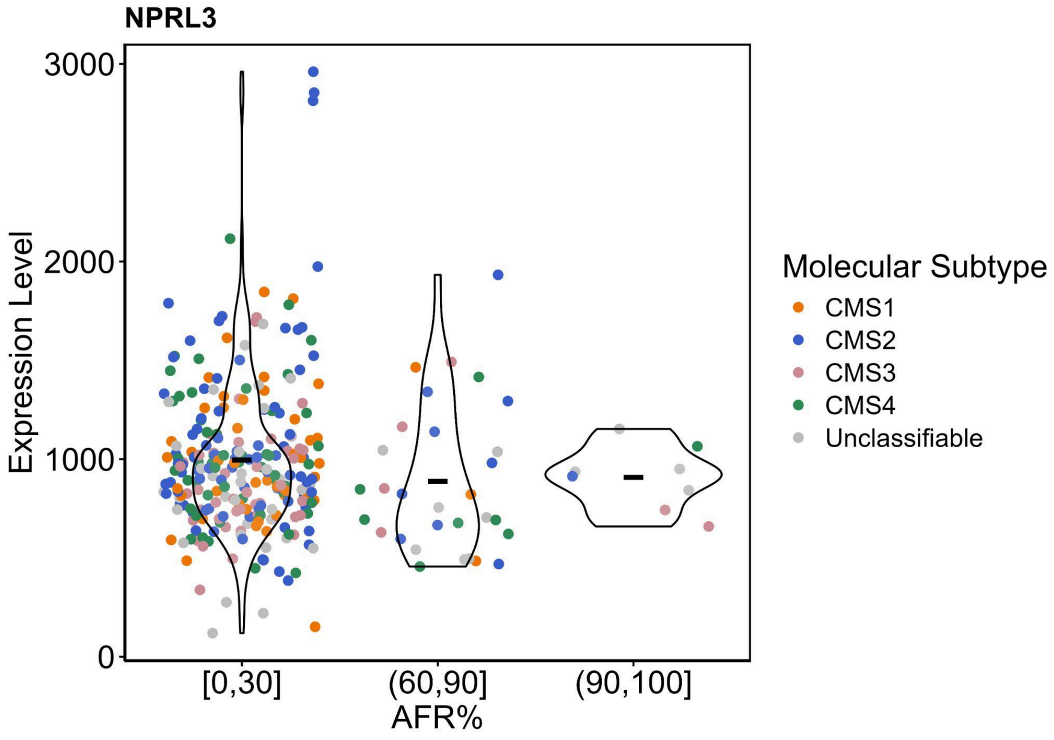

**Figure S10.** RNAseq Quality Control Metrics. **A**, Average GC content across all reads per sample. Samples are grouped by genetic ancestry (dot color). **B,** Mean inner distance between read pairs (calculated as the distance from the end of Read 1 to the start of Read 2) per sample. Samples are grouped by genetic ancestry (dot color). **C,** Distribution of mapped reads across genomic regions for each sample. **D,** Mapping efficiency across samples, showing the proportion of reads that uniquely map to one genomic locus (UQ_map), map to multiple loci (Mult_map), or remain unmapped (Unmapped).

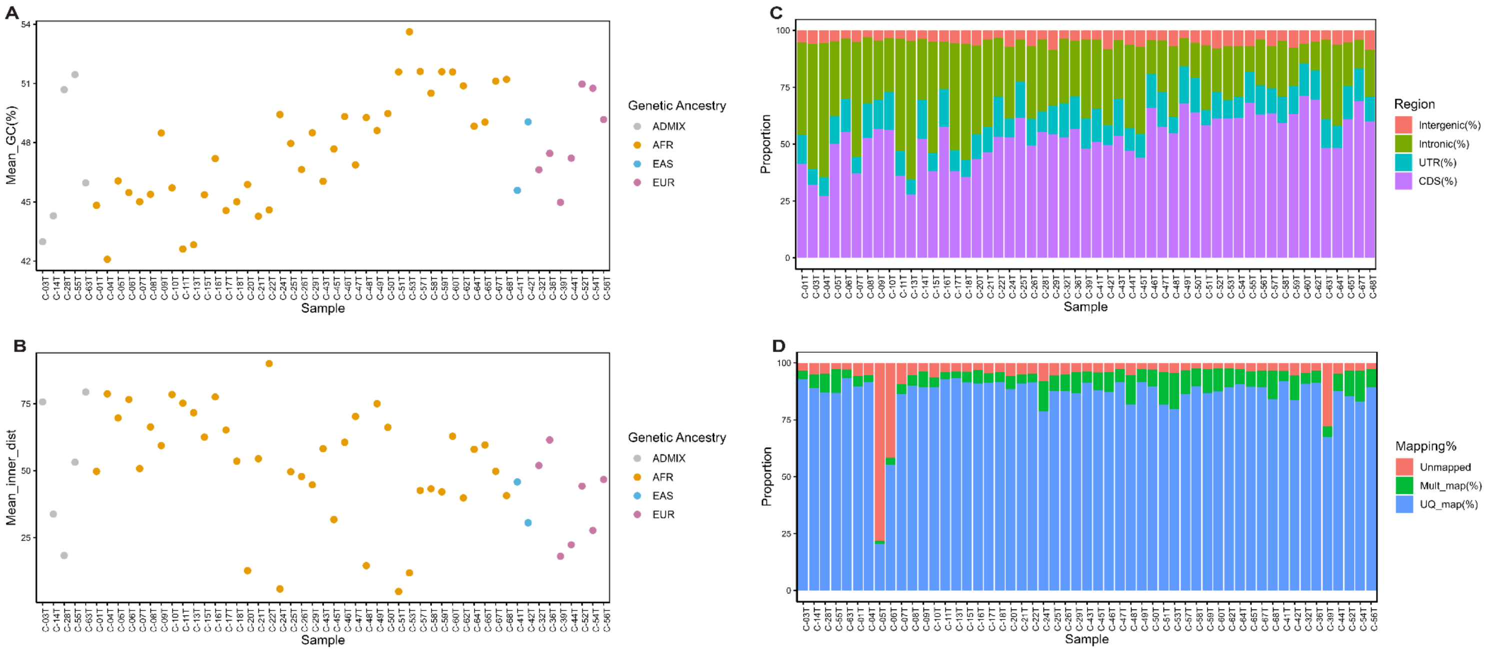

**Figure S11. A,** Heatmap visualizing z-score transformed expression levels for all samples in the transcriptomic cohort across CMS marker genes (row annotations). Predicted CMS classifications, as well as genetic ancestry information and MSI status per sample, are also shown (column annotations). Unclassifiable samples are not displayed. **B,** Proportional distribution of different ancestries within CMS groups in the P1000 and TCGA-COAD cohorts. Significant differences between ancestry group distribution by subtype between the two cohorts are indicated above each comparison (Chi-Square p-value; pval < 0.01 is indicated with "***", pval < 0.03 is indicated with "**", pval < 0.05 is indicated with "*"). **C,** Differential gene expression between individuals of African and European ancestry. Expression levels of ancestry-associated genes are displayed across bins of African genetic admixture. Each point represents a sample, with shape indicating molecular subtype and color representing the proportion of European ancestry.

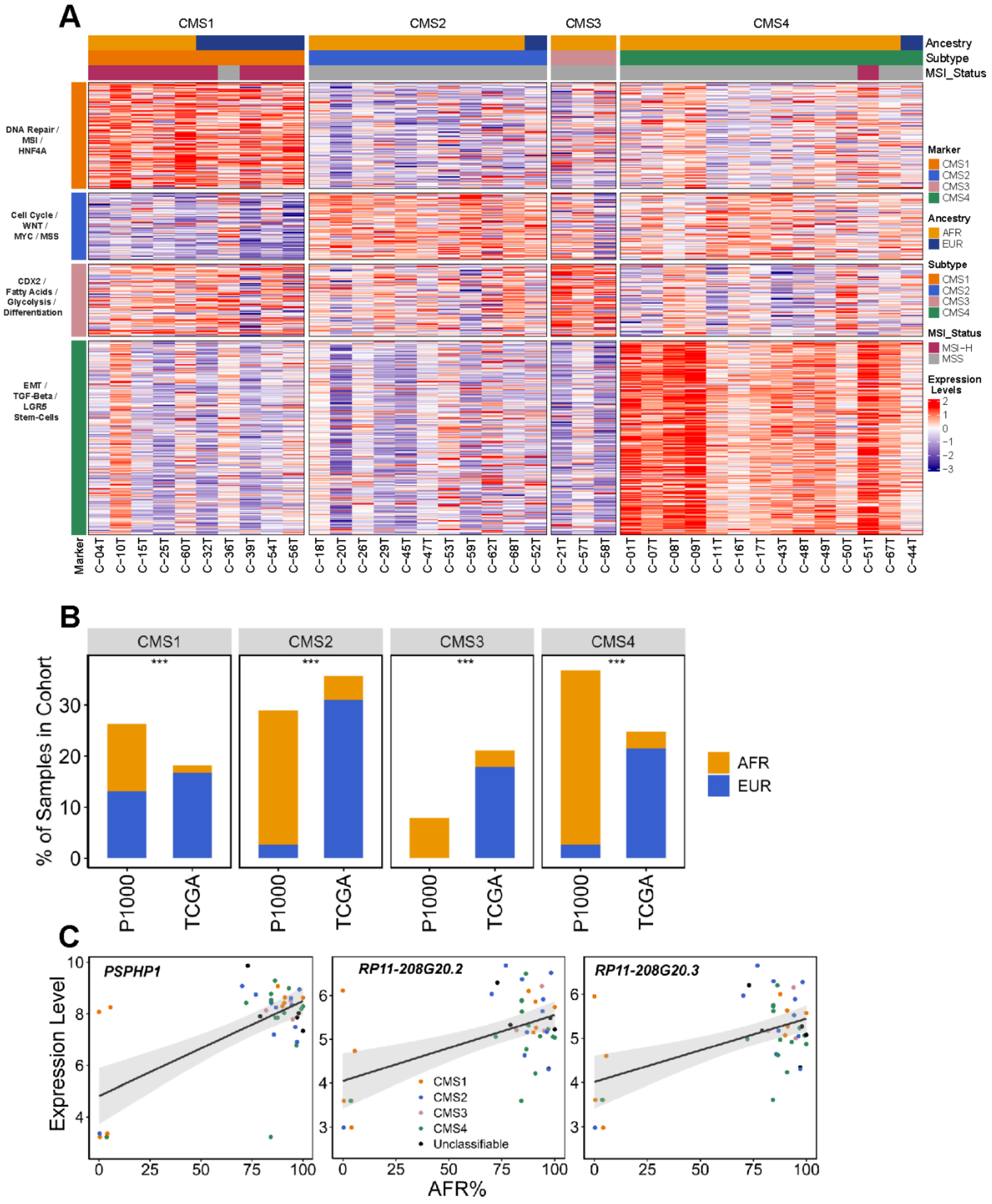

**Figure S12.** Distribution of the Hallmark "KRAS signaling up", as obtained by gene set variation analysis (GSVA), in the AFRg patients with MMS tumors from the P-1000 cohort. **A)** The AFRg patients are divided by the KRAS mutation status. **B**) The AFRg patients are divided by KRAS mutation status and CMS subtypes. The p-value of the Wilcoxon rank sum exact test done on KRAS status (mutated vs wild-type) subgroups is shown. A dashed line indicates the median of each subset.

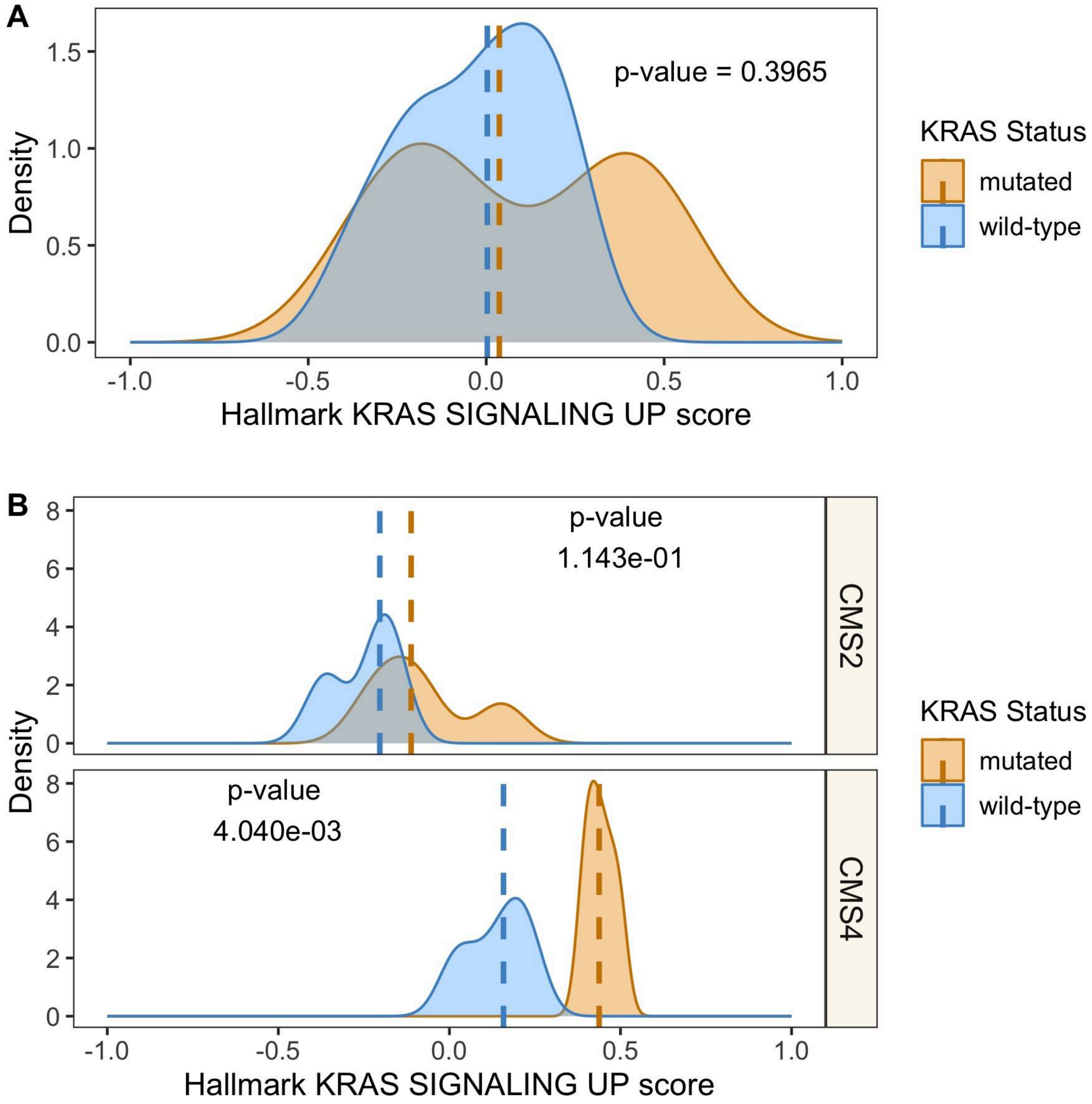

**Figure S13.** Similarity between copy number profiles of the EURg and AFRg sections of the NYGC P-1000 cohort. The Szymkiewicz–Simpson coefficient was calculated separately for the amplification (**A**) and deletion (**B**) profiles. A star is present when the coefficient between a pair of profiles is in the 5% highest values when compared to simulated profiles created from one sample of the pair. **C**) The distribution of the profiles between the two clusters obtained for the amplification and the deletion analyses.

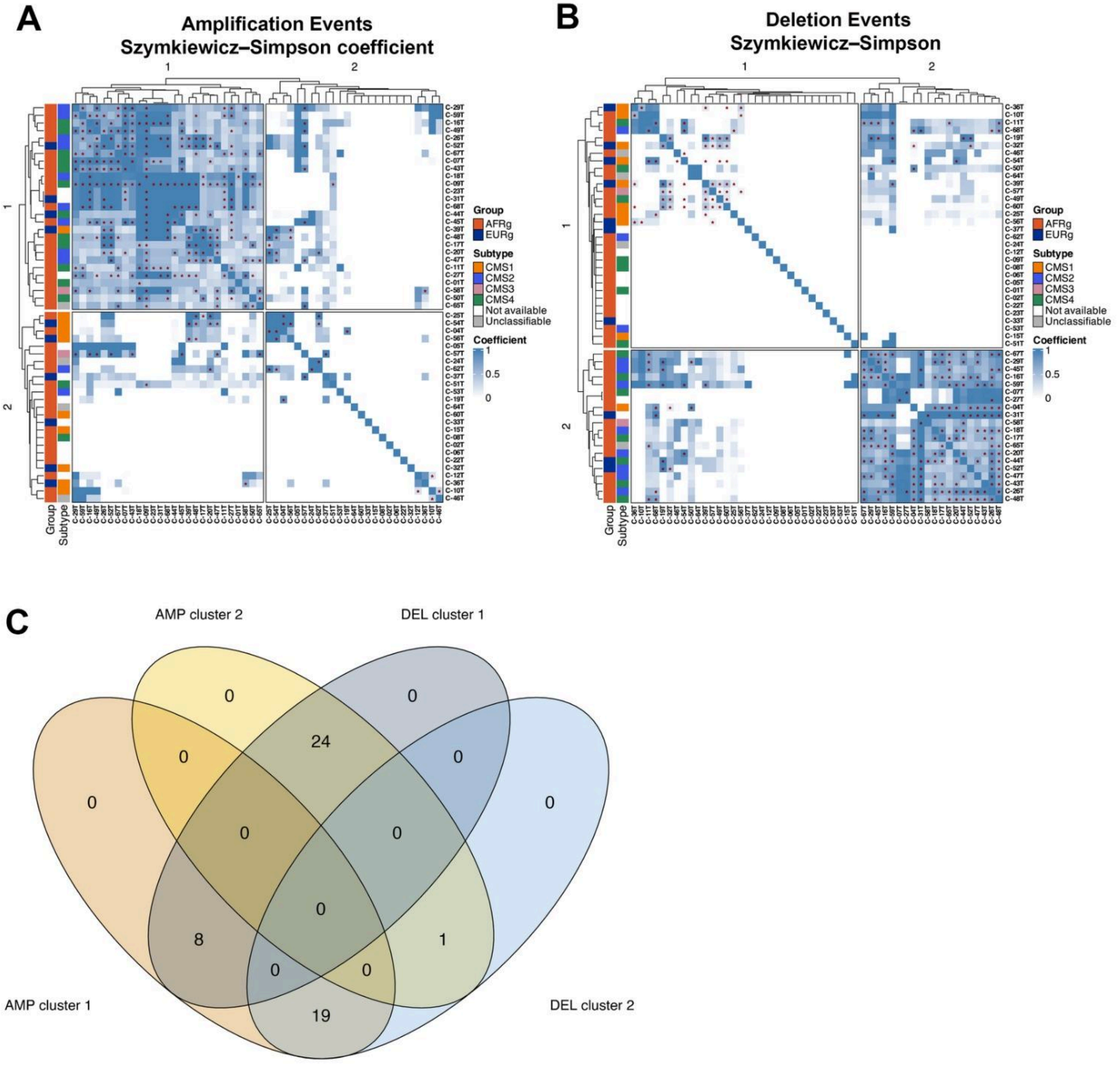

**Figure S14.** The copy number variation signatures (COSMIC CN) for the AFRg and EURg patients in both P-1000 (**A**) and TCGA COAD (**B**) cohorts. A signature is considered detected in a patient when at least 10 mutations are present in the signature. **C)** The associations between the presence of the CN1, CN2, and CN13 signatures and 1) TP53 mutation (TP53m) status, 2) TP53m with LOH status, and 3) chr18q deletion status are shown in P-1000 MSS patients. The chr18q region includes the SMAD4 and SMAD2 genes. TP53 LOH indicates patients with a TP53 mutation + LOH.

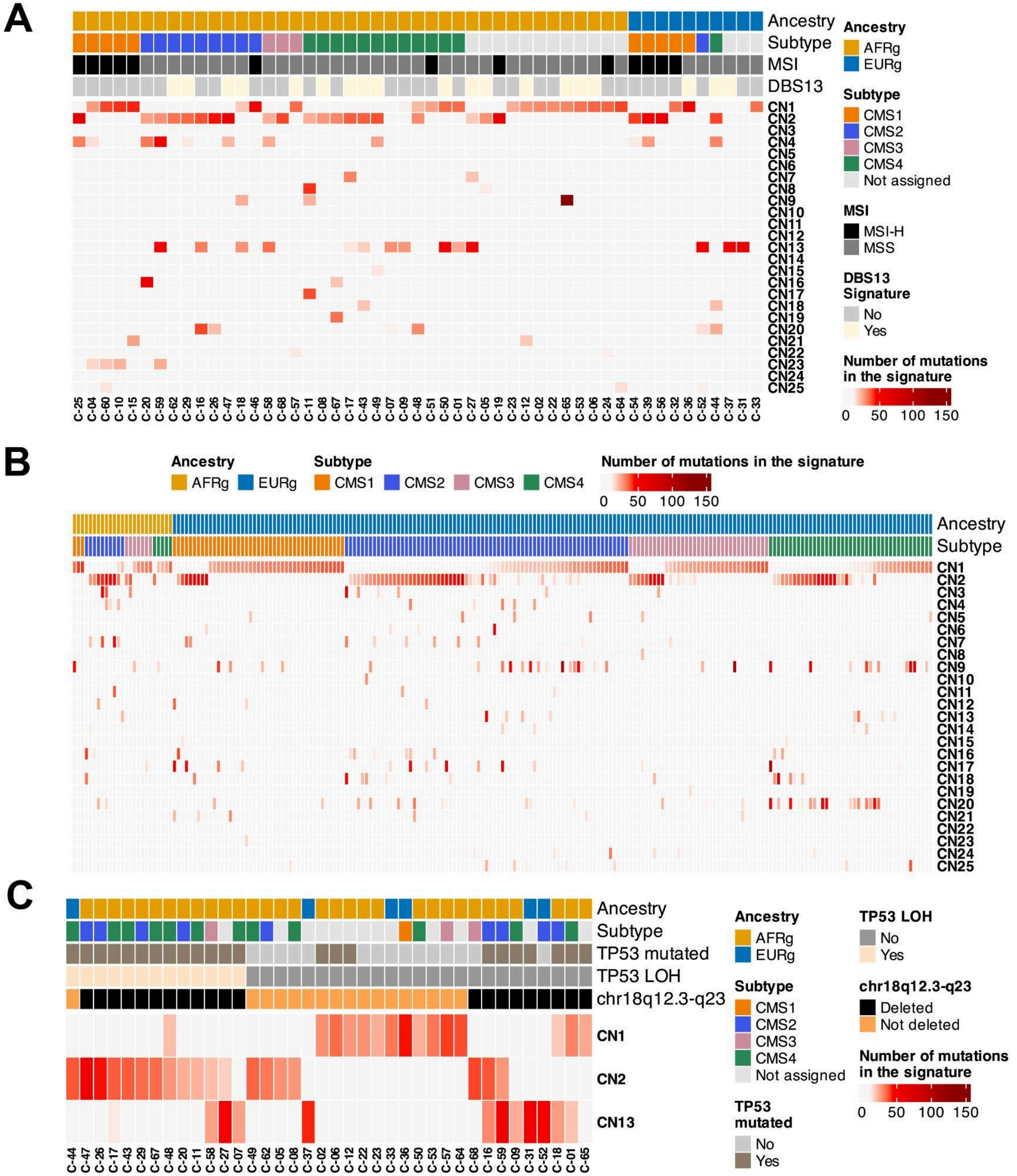

**Figure S15.** HLA typing of the NYGC P-1000 AFRg and EURg patients. **A**) The comparison between HLA typing in cancer and matching adjacent non-tumor typing reveals loss of heterozygosity in a few patients. The comparison between the HLA allele rankings in the P-1000 AFRg cohort, for three HLA Class I and two HLA Class II genes, with the rankings in the African American National Marrow Donor Program (NMDP) database: **B**) HLA-A; **C**) HLA-B; **D**) HLA-C; **E**) HLA-DQB1; and **F**) HLA-DRB1.

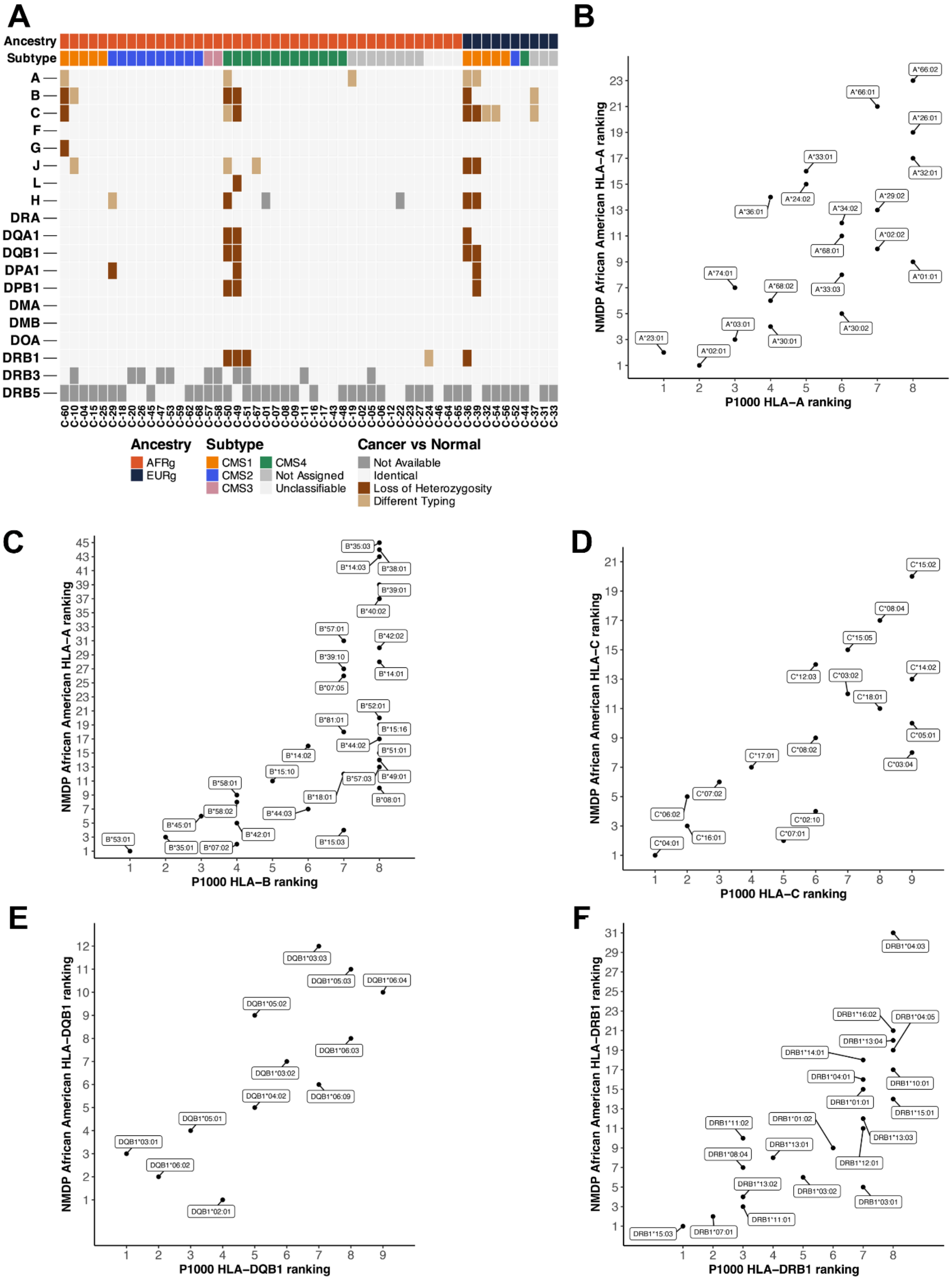

**Figure S16.** Beta diversity of 16S rRNA gene profiles (**A–D**). Principal coordinates analysis (PCoA) of unweighted UniFrac (left) and weighted UniFrac (right) distances. Points are individual samples (labeled by ID), and dashed ellipses denote 95% confidence regions for each group. Comparisons shown: (**A**) tissue type (tumor, n=32; non-tumor, n=31), (**B**) sex (male, n=14; female, n=18), (**C**) tumor location (left, n=16; right, n=16), and (**D**) stage (early, n=22; late, n=10). PERMANOVA $R^2$ and p values are displayed on each plot; no comparison reached statistical significance (all $p > 0.05$).

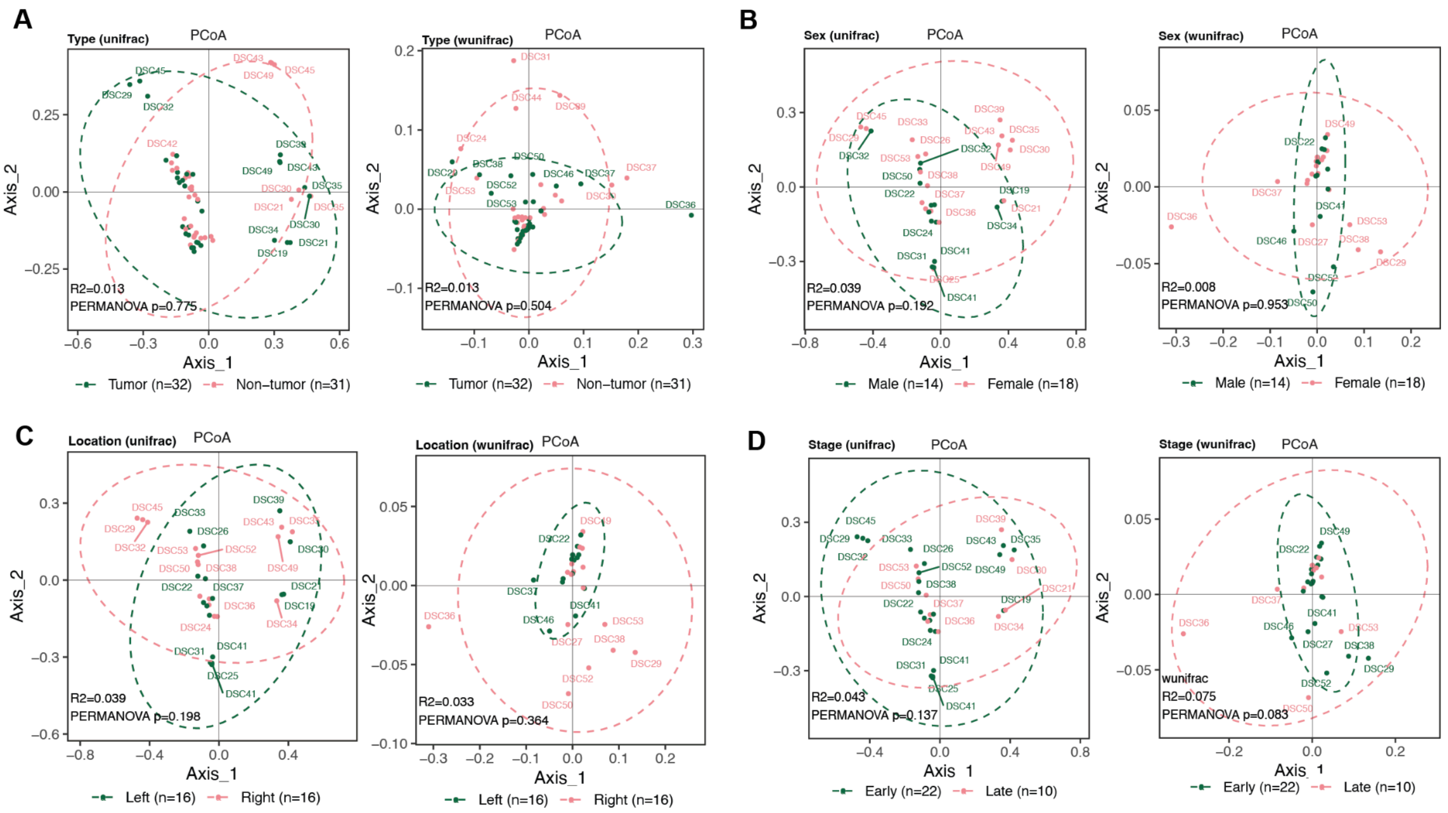

**Figure S17.** Visualization of Differentially Prevalent Taxa and Clinical Metadata

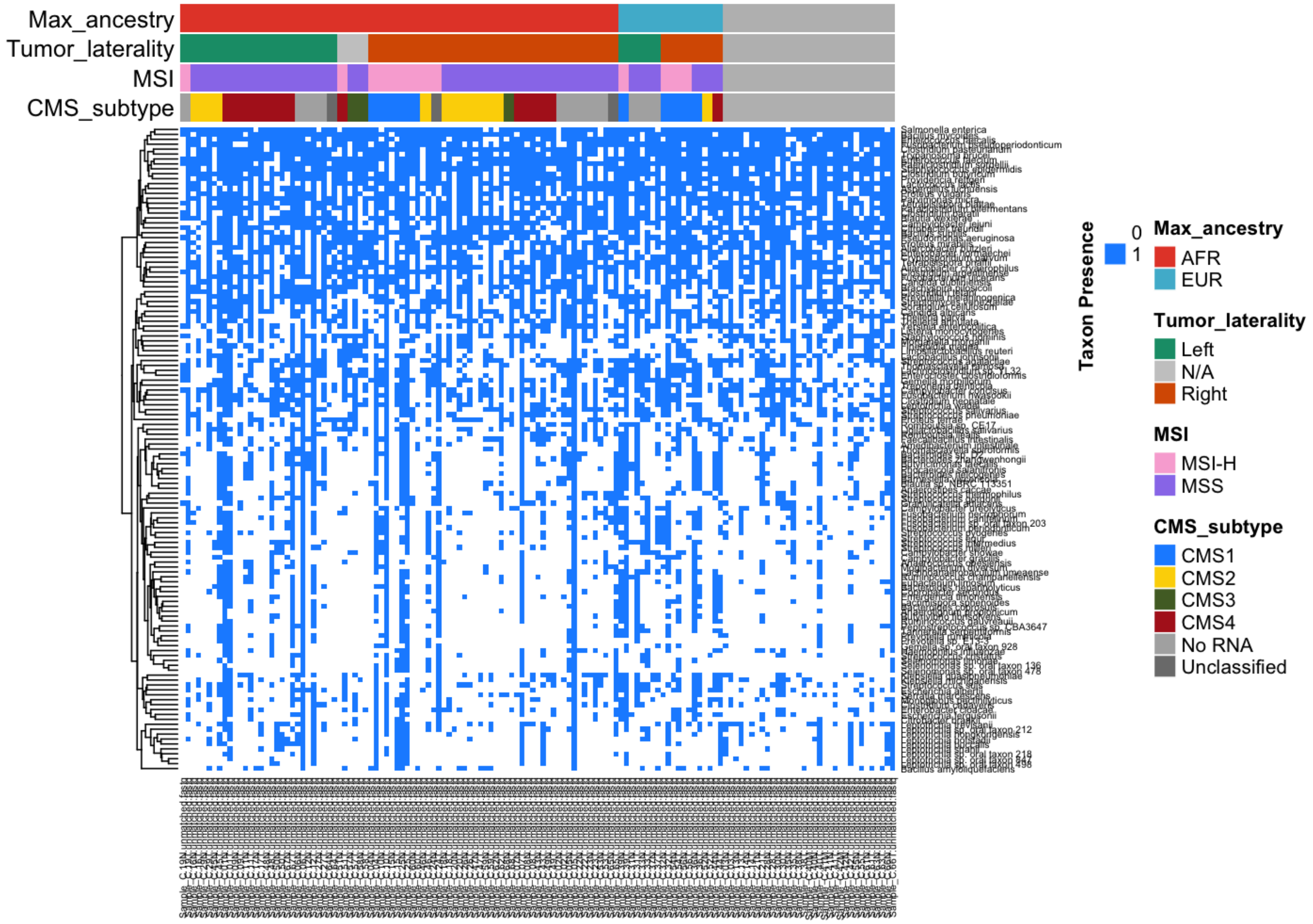

**Figure S18.** Cluster of oral-origin bacteria in tumor and adjacent non-tumor colon tissues

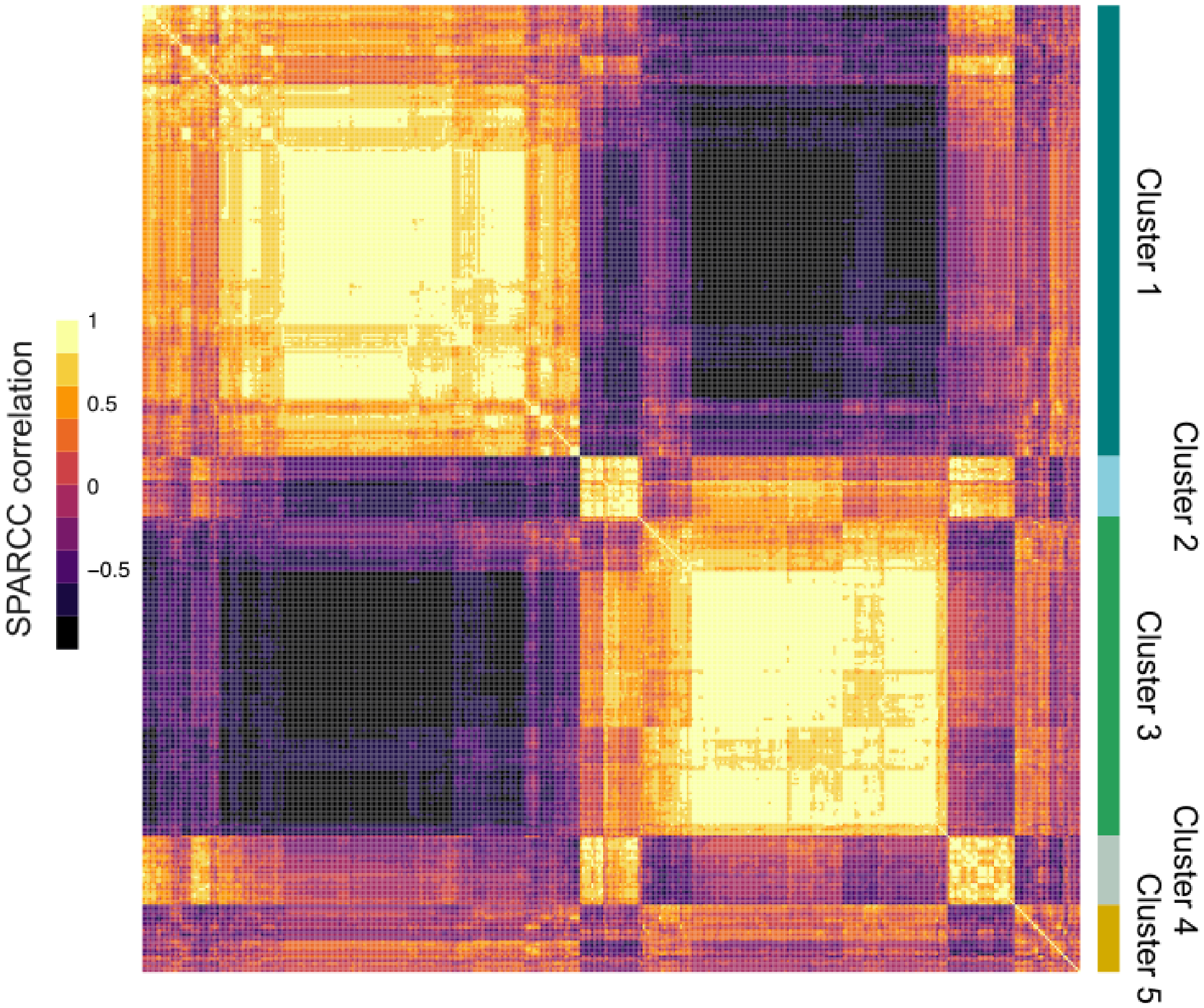

**Figure S19.** Dana Farber Cancer Institute (DFCI) exome cohort somatic variant comparison for P-1000 AFRg vs DFCI EURg groups showing suggestive enrichment of mutation frequency (number of individuals per cohort with mutations in each gene) between cohorts in genes with uncorrected p-value less than 0.05 (*), 0.01 (**), or 0.001 (***).

**DFCIEUR (n = 116) v/s P1000AFR (n = 33)**

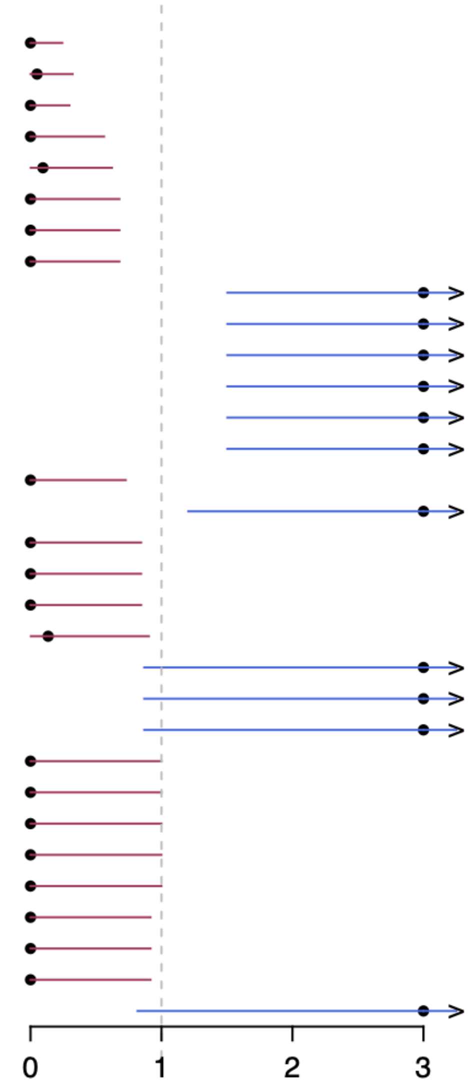

| Gene | DFCIEUR | P1000AFR | OR | P−value |
|---|---|---|---|---|
| *USP17L24* | 40 | 0 | 0 | *** |
| *MUC6* | 45 | 1 | 0.05 | *** |
| *ZNF729* | 35 | 0 | 0 | *** |
| *GOLGA8H* | 22 | 0 | 0 | ** |
| *ZNF208* | 29 | 1 | 0.095 | ** |
| *HLA−DRB1* | 19 | 0 | 0 | ** |
| *PABPC1* | 19 | 0 | 0 | ** |
| *ZNF714* | 19 | 0 | 0 | ** |
| *GYS2* | 0 | 3 | Inf | * |
| *NAA15* | 0 | 3 | Inf | * |
| *RBMXL3* | 0 | 3 | Inf | * |
| *TAF2* | 0 | 3 | Inf | * |
| *TDRD15* | 0 | 3 | Inf | * |
| *ZNF469* | 0 | 3 | Inf | * |
| *USP17L12* | 18 | 0 | 0 | * |
| *NALCN* | 3 | 5 | 6.609 | * |
| *CNTNAP3* | 16 | 0 | 0 | * |
| *FRG1* | 16 | 0 | 0 | * |
| *LRRC37A* | 16 | 0 | 0 | * |
| *MUC3A* | 22 | 1 | 0.135 | * |
| *COL5A1* | 1 | 3 | 11.253 | * |
| *PYGM* | 1 | 3 | 11.253 | * |
| *TSHZ3* | 1 | 3 | 11.253 | * |
| *GOLGA6L3* | 14 | 0 | 0 | * |
| *HYDIN* | 14 | 0 | 0 | * |
| *IGSF3* | 14 | 0 | 0 | * |
| *KRTAP4−7* | 14 | 0 | 0 | * |
| *ZNF98* | 14 | 0 | 0 | * |
| *CSMD1* | 15 | 0 | 0 | * |
| *ZNF107* | 15 | 0 | 0 | * |
| *ZNF90* | 15 | 0 | 0 | * |
| *ZBTB7A* | 3 | 4 | 5.119 | * |

Odds ratio with 95% CI
(1 = no effect, < 1 DFCIEUR has more mutants)

**Table S1:** FFPE exome cohort sample characteristics

<table>
<tr><th>Race</th><th>Sex</th><th>Age Range</th><th>Average Age</th><th colspan="2">Tumor Stage/<br>Location*</th></tr>
<tr><td rowspan="2">African Americans</td><td>Female<br>37</td><td rowspan="2">44 - 94</td><td rowspan="2">67.7</td><td colspan="2" rowspan="2">T1 - 2<br>T2 - 6<br>T3 - 30<br>T4 - 14</td></tr>
<tr><td>Male<br>15</td></tr>
<tr><td colspan="4">N of AA participants = 52</td><td>S - 12<br>D - 6<br>T - 3</td><td>A - 14<br>C - 16<br>H - 1</td></tr>
<tr><td rowspan="2">European Americans</td><td>Female<br>21</td><td rowspan="2">47 - 97</td><td rowspan="2">69.1</td><td colspan="2" rowspan="2">T1 - 4<br>T2 - 5<br>T3 - 28<br>T4 - 14</td></tr>
<tr><td>Male<br>30</td></tr>
<tr><td colspan="4">N of EA participants = 51</td><td>S - 18<br>D - 7<br>T - 5</td><td>A - 9<br>C - 12<br>H - 0</td></tr>
<tr><td colspan="6">*Tumor Location: S= sigmoid, D= descending, T= transverse, A= ascending, C= cecum, H= hepatic flexure</td></tr>
</table>

**Table S2**. Frequency of mutations in selected genes across cohorts

| Somatic Variant Gene | AFR FFPE (52) | EUR FFPE (51) | AFR P-1000 (42) | EUR P-1000 (10) | EUR DFCI (177) | AFR TCGA COAD (49) | EUR TCGA COAD (175) |
|---|---|---|---|---|---|---|---|
| **KRAS** | 24 (46%) | 18 (35%) | 18 (43%) | 2 (20%) | 54 (31%) | 26 (53%) | 77 (44%) |
| **CSMD2** | 9 (17%) | 1 (2%) | 6 (14%) | 0 | 17 (10%) | 9 (18%) | 22 (13%) |
| | **AFR FFPE MSS (46)** | **EUR FFPE MSS (50)** | **AFR P-1000 MSS (33)** | **EUR P-1000 MSS (6)** | **EUR DFCI MSS* (116)** | **AFR TCGA COAD MSS (43)** | **EUR TCGA COAD MSS (128)** |
| **BRAF** | 2 (4%) | 3 (6%) | 2 (6%) | 1 | 15 (13%) | 2 (5%) | 10 (8%) |
| **KRAS** | 23 (50%) | 17 (34%) | 15 (45%) | 2 (33%) | 41 (35%) | 22 (51%) | 55 (43%) |
| **CSMD2** | 6 (13%) | 1 (2%) | 1 (3%) | 0 | 9 (8%) | 4 (9%) | 10 (8%) |
| **BCL9L** | 2 (4%) | 1 (2%) | 4 (12%) | 0 | 4 (3%) | 1 (2%) | 8 (6%) |
| **FBLIM1** | 8 (17%) | 1 (2%) | 0 | 0 | 1 (0.9%) | 0 | 0 |
| **TBXA2R** | 6 (13%) | 0 | 1 (3%) | 0 | 0 | 0 | 0 |
| **LTBP1** | 6 (13%) | 1 (2%) | 1 (3%) | 0 | 1 | 1 (2%) | 2 (2%) |
| **LRP1B** | 4 (9%) | 0 | 4 (12%) | 0 | 17 (15%) | 7 (16%) | 22 (17%) |
| **CSMD1** | 3 (7%) | 3 (6%) | 0 | 0 | 15 (13%) | 6 (14%) | 17 (13%) |
| **TCF7L2** | 2 (4%) | 4 (8%) | 5 (15%) | 0 | 10 (9%) | 2 (5%) | 13 (10%) |
| **SMAD2** | 1 (2%) | 0 | 3 (9%) | 2 (33%) | 3 (3%) | 3 (7%) | 1 (0.8%) |
| **ZBTB7A** | 1 (2%) | 0 | 4 (12%) | 1 (17%) | 3 (3%) | 1 (2%) | 0 |
| **ZNF469** | 1 (2%) | 2 (4%) | 3 (9%) | 2 (33%) | 0 | 0 | 0 |
| **RBMXL3** | 1 (2%) | 0 | 3 (9%) | 0 | 0 | 1 (2%) | 0 |
| **Germline Variant Gene** | **AFR FFPE (52)** | **EUR FFPE (51)** | **AFR P-1000 (42)** | **EUR P-1000 (10)** | **EUR DFCI (187)** | **AFR TCGA COAD (49)** | **AFR TCGA multi cancer (157)** |
| **AHNAK2** | 19 (37%) | 5 (10%) | 33 (75%) | 3 (25%) | 116 (62%) | 37 (70%) | 113 (72%) |

Table S3. Continental admixture proportion for the profiles in the NYGC P-1000 cohort

| The columns are: | |
|---|---|
| PatientID | The identifiant of the patient whom continental ancestry has been infered. |
| SAS | The proportion of ancestry associated to the South Asian super population (SAS) |
| EAS | The proportion of ancestry associated to the East Asian super population (EAS) |
| EUR | The proportion of ancestry associated to the European super population (EUR) |
| AMR | The proportion of ancestry associated to the Ad Mixed American super population (AMR) |
| AFR | The proportion of ancestry associated to the African super population (AFR) |
| Assigned Group | The group assigned to this patient for the analysis purpose. |

| PatientID | SAS | EAS | EUR | AMR | AFR | Assigned Group |
|---|---|---|---|---|---|---|
| C-01 | 1,00E-05 | 0,010694 | 1,00E-05 | 0,021057 | 0,968229 | AFRg |
| C-02 | 0,005553 | 1,00E-05 | 0,03189 | 1,00E-05 | 0,962537 | AFRg |
| C-04 | 0,006056 | 1,00E-05 | 0,082354 | 1,00E-05 | 0,91157 | AFRg |
| C-05 | 1,00E-05 | 1,00E-05 | 1,00E-05 | 1,00E-05 | 0,99996 | AFRg |
| C-06 | 1,00E-05 | 1,00E-05 | 0,054921 | 0,013543 | 0,931516 | AFRg |
| C-07 | 0,004383 | 1,00E-05 | 0,087232 | 1,00E-05 | 0,908365 | AFRg |
| C-08 | 1,00E-05 | 1,00E-05 | 1,00E-05 | 1,00E-05 | 0,99996 | AFRg |
| C-09 | 0,000504 | 1,00E-05 | 0,149119 | 0,007973 | 0,842394 | AFRg |
| C-10 | 0,011623 | 1,00E-05 | 0,083489 | 0,005383 | 0,899495 | AFRg |
| C-11 | 1,00E-05 | 1,00E-05 | 0,043771 | 0,025331 | 0,930878 | AFRg |
| C-12 | 0,004144 | 1,00E-05 | 0,049725 | 1,00E-05 | 0,94611 | AFRg |
| C-14 | 0,012823 | 1,00E-05 | 0,25298 | 0,08374 | 0,650447 | Not assigned |
| C-15 | 0,011131 | 0,008731 | 0,071286 | 1,00E-05 | 0,908842 | AFRg |
| C-16 | 0,002914 | 1,00E-05 | 0,152999 | 1,00E-05 | 0,844067 | AFRg |
| C-17 | 0,001132 | 1,50E-05 | 0,007171 | 1,00E-05 | 0,991672 | AFRg |

| | | | | | | |
|---|---|---|---|---|---|---|
| C-18 | 1,00E-05 | 1,00E-05 | 0,142381 | 1,00E-05 | 0,857589 | AFRg |
| C-19 | 0,019222 | 1,00E-05 | 0,029865 | 0,150441 | 0,800462 | AFRg |
| C-20 | 0,005757 | 0,002262 | 0,223176 | 1,00E-05 | 0,768795 | AFRg |
| C-22 | 0,018125 | 1,00E-05 | 0,180346 | 0,066625 | 0,734894 | AFRg |
| C-23 | 1,00E-05 | 1,00E-05 | 1,00E-05 | 1,00E-05 | 0,99996 | AFRg |
| C-24 | 1,00E-05 | 1,00E-05 | 1,00E-05 | 1,00E-05 | 0,99996 | AFRg |
| C-25 | 1,00E-05 | 1,00E-05 | 1,00E-05 | 1,00E-05 | 0,99996 | AFRg |
| C-26 | 1,00E-05 | 0,003074 | 0,128093 | 1,00E-05 | 0,868813 | AFRg |
| C-27 | 0,00784 | 0,000615 | 0,053396 | 0,009694 | 0,928456 | AFRg |
| C-28 | 1,00E-05 | 1,00E-05 | 0,372437 | 1,00E-05 | 0,627533 | Not assigned |
| C-29 | 1,00E-05 | 1,00E-05 | 0,053045 | 0,000943 | 0,945992 | AFRg |
| C-31 | 0,044859 | 1,00E-05 | 0,951388 | 1,00E-05 | 0,003733 | EURg |
| C-32 | 0,011868 | 1,00E-05 | 0,983427 | 1,00E-05 | 0,004686 | EURg |
| C-33 | 0,073036 | 1,00E-05 | 0,896168 | 1,00E-05 | 0,030775 | EURg |
| C-36 | 0,020904 | 1,00E-05 | 0,979066 | 1,00E-05 | 1,00E-05 | EURg |
| C-37 | 0,023186 | 1,00E-05 | 0,972167 | 1,00E-05 | 0,004627 | EURg |

| | | | | | | |
|---|---|---|---|---|---|---|
| C-38 | 0,030996 | 1,00E-05 | 0,409237 | 0,425197 | 0,13456 | Not assigned |
| C-39 | 0,078619 | 1,00E-05 | 0,882042 | 1,00E-05 | 0,039318 | EURg |
| C-41 | 0,061111 | 0,934684 | 1,00E-05 | 1,00E-05 | 0,004185 | Not assigned |
| C-42 | 1,00E-05 | 0,993609 | 1,00E-05 | 1,00E-05 | 0,006361 | Not assigned |
| C-43 | 0,007194 | 1,00E-05 | 0,105269 | 0,011196 | 0,876331 | AFRg |
| C-44 | 0,095105 | 1,00E-05 | 0,868194 | 1,00E-05 | 0,036681 | EURg |
| C-45 | 0,001794 | 1,00E-05 | 0,00408 | 0,011558 | 0,982558 | AFRg |
| C-46 | 0,015343 | 1,00E-05 | 0,040498 | 0,153419 | 0,79073 | AFRg |
| C-47 | 0,26248 | 0,013281 | 0,02224 | 1,00E-05 | 0,701988 | AFRg |
| C-48 | 0,105617 | 1,00E-05 | 0,031151 | 1,00E-05 | 0,863211 | AFRg |
| C-49 | 0,021115 | 1,00E-05 | 0,235721 | 0,021636 | 0,721518 | AFRg |
| C-50 | 3,10E-05 | 0,003028 | 0,131561 | 0,020716 | 0,844664 | AFRg |
| C-51 | 0,000482 | 1,00E-05 | 0,131583 | 0,002375 | 0,86555 | AFRg |
| C-52 | 0,034592 | 1,00E-05 | 0,962272 | 1,00E-05 | 0,003115 | EURg |
| C-53 | 1,00E-05 | 1,00E-05 | 0,037241 | 1,00E-05 | 0,962729 | AFRg |
| C-54 | 0,050339 | 1,00E-05 | 0,908344 | 1,00E-05 | 0,041297 | EURg |

| | | | | | | |
|---|---|---|---|---|---|---|
| **C-55** | **0,013309** | **1,00E-05** | **0,495986** | **0,155513** | **0,335182** | **Not assigned** |
| **C-56** | **0,080072** | **1,00E-05** | **0,864186** | **1,00E-05** | **0,055721** | **EURg** |
| **C-57** | **0,000623** | **1,00E-05** | **0,05774** | **0,002416** | **0,93921** | **AFRg** |
| **C-58** | **0,008992** | **1,00E-05** | **0,165148** | **0,006756** | **0,819094** | **AFRg** |
| **C-59** | **1,00E-05** | **0,005276** | **0,04966** | **0,0033** | **0,941754** | **AFRg** |
| **C-60** | **0,008595** | **0,010412** | **0,056864** | **0,047229** | **0,876901** | **AFRg** |
| **C-62** | **0,023845** | **1,00E-05** | **0,100337** | **0,030576** | **0,845233** | **AFRg** |
| **C-63** | **1,00E-05** | **1,00E-05** | **0,338951** | **0,043643** | **0,617387** | **Not assigned** |
| **C-64** | **1,00E-05** | **1,00E-05** | **0,012964** | **0,007568** | **0,979448** | **AFRg** |
| **C-65** | **0,010487** | **1,00E-05** | **0,260196** | **1,00E-05** | **0,729297** | **AFRg** |
| **C-67** | **1,00E-05** | **0,007021** | **0,129624** | **0,019546** | **0,843799** | **AFRg** |
| **C-68** | **0,013423** | **1,00E-05** | **0,015243** | **1,00E-05** | **0,971314** | **AFRg** |

Table S4. Continental admixture proportion for the profiles in the CSHL FFPE cohort

| The columns are: | |
|---|---|
| PatientID | The identifiant of the patient whom continental ancestry has been infered. |
| SAS | The proportion of ancestry associated to the South Asian super population (SAS) |
| EAS | The proportion of ancestry associated to the East Asian super population (EAS) |
| EUR | The proportion of ancestry associated to the European super population (EUR) |
| AMR | The proportion of ancestry associated to the Ad Mixed American super |

| | |
|---|---|
| | population (AMR) |
| AFR | The proportion of ancestry associated to the African super population (AFR) |
| Assigned Group | The group assigned to this patient for the analysis purpose. |

| PatientID | SAS | EAS | EUR | AMR | AFR | Assigned Group |
|---|---|---|---|---|---|---|
| **DS-R-C1** | **0,186815** | **0,007741** | **0,089821** | **1,00E-05** | **0,715612** | **AFRg** |
| **DS-R-C10** | **1,00E-05** | **0,007056** | **0,019258** | **0,018182** | **0,955494** | **AFRg** |
| **DS-R-C11** | **0,019514** | **1,00E-05** | **1,00E-05** | **1,00E-05** | **0,980456** | **AFRg** |
| **DS-R-C12** | **1,00E-05** | **0,010434** | **0,124395** | **0,013708** | **0,851453** | **AFRg** |
| **DS-R-C13** | **0,001379** | **1,00E-05** | **0,057888** | **0,025737** | **0,914986** | **AFRg** |
| **DS-R-C14** | **1,00E-05** | **0,007487** | **0,09661** | **0,008074** | **0,887819** | **AFRg** |
| **DS-R-C15** | **0,035037** | **0,015193** | **0,07622** | **1,00E-05** | **0,873539** | **AFRg** |
| **DS-R-C16** | **0,049682** | **1,00E-05** | **0,203533** | **0,035739** | **0,711035** | **AFRg** |
| **DS-R-C17** | **1,00E-05** | **0,033612** | **0,107963** | **0,004143** | **0,854272** | **AFRg** |
| **DS-R-C18** | **0,028115** | **1,00E-05** | **0,158236** | **0,018219** | **0,795419** | **AFRg** |
| **DS-R-C19** | **0,007658** | **1,00E-05** | **0,059575** | **0,005667** | **0,92709** | **AFRg** |
| **DS-R-C2** | **0,100611** | **1,00E-05** | **0,833547** | **1,00E-05** | **0,065822** | **EURg** |
| **DS-R-C20** | **0,011586** | **1,00E-05** | **0,063691** | **0,018049** | **0,906664** | **AFRg** |

| | | | | | | |
|---|---|---|---|---|---|---|
| DS-R-C21 | 0,023158 | 0,005537 | 0,021167 | 1,00E-05 | 0,950129 | AFRg |
| DS-R-C22 | 1,00E-05 | 0,005172 | 0,100798 | 1,00E-05 | 0,89401 | AFRg |
| DS-R-C23 | 0,064863 | 0,030998 | 0,358896 | 0,094921 | 0,450321 | Not assigned |
| DS-R-C24 | 0,00458 | 0,022419 | 0,076232 | 0,010411 | 0,886358 | AFRg |
| DS-R-C25 | 1,00E-05 | 0,012349 | 0,055422 | 0,009828 | 0,922391 | AFRg |
| DS-R-C26 | 0,020523 | 0,025054 | 0,080372 | 0,024098 | 0,849953 | AFRg |
| DS-R-C27 | 0,063463 | 0,00594 | 0,072898 | 0,018706 | 0,838993 | AFRg |
| DS-R-C28 | 0,157176 | 0,022469 | 0,361718 | 1,00E-05 | 0,458627 | Not assigned |
| DS-R-C29 | 0,094099 | 0,000187 | 0,358114 | 0,022209 | 0,525392 | Not assigned |
| DS-R-C3 | 1,00E-05 | 1,00E-05 | 0,077039 | 1,00E-05 | 0,922931 | AFRg |
| DS-R-C30 | 0,071393 | 0,03673 | 0,093733 | 0,020435 | 0,777709 | AFRg |
| DS-R-C31 | 0,09887 | 0,034164 | 0,783896 | 0,003763 | 0,079306 | EURg |
| DS-R-C32 | 1,00E-05 | 0,010028 | 0,160252 | 0,033052 | 0,796658 | AFRg |
| DS-R-C33 | 0,011351 | 0,011533 | 0,064005 | 0,024518 | 0,888594 | AFRg |
| DS-R-C34 | 0,026453 | 0,004725 | 0,104352 | 0,029132 | 0,835338 | AFRg |
| DS-R-C35 | 0,023853 | 0,006706 | 0,088652 | 1,00E-05 | 0,880779 | AFRg |

| | | | | | | |
|---|---|---|---|---|---|---|
| **DS-R-C36** | **1,00E-05** | **1,00E-05** | **0,053702** | **0,020303** | **0,925975** | **AFRg** |
| **DS-R-C37** | **0,005169** | **0,003151** | **0,035491** | **1,00E-05** | **0,956179** | **AFRg** |
| **DS-R-C38** | **0,033139** | **0,026388** | **0,092896** | **0,01128** | **0,836298** | **AFRg** |
| **DS-R-C39** | **1,00E-05** | **0,02153** | **1,00E-05** | **1,00E-05** | **0,97844** | **AFRg** |
| **DS-R-C4** | **0,028328** | **1,00E-05** | **0,003075** | **0,109344** | **0,859243** | **AFRg** |
| **DS-R-C40** | **0,05434** | **0,01312** | **0,116704** | **0,011316** | **0,80452** | **AFRg** |
| **DS-R-C41** | **0,122165** | **0,018476** | **0,795341** | **1,00E-05** | **0,064008** | **EURg** |
| **DS-R-C42** | **0,108506** | **0,0016** | **0,059127** | **0,015351** | **0,815416** | **AFRg** |
| **DS-R-C43** | **0,034687** | **0,027189** | **0,057512** | **1,00E-05** | **0,880601** | **AFRg** |
| **DS-R-C44** | **0,050684** | **0,031829** | **0,167496** | **0,023405** | **0,726586** | **AFRg** |
| **DS-R-C45** | **1,00E-05** | **1,00E-05** | **0,186591** | **1,00E-05** | **0,813379** | **AFRg** |
| **DS-R-C46** | **1,00E-05** | **1,00E-05** | **0,089922** | **1,00E-05** | **0,910048** | **AFRg** |
| **DS-R-C5** | **0,017582** | **1,00E-05** | **0,086133** | **1,00E-05** | **0,896266** | **AFRg** |
| **DS-R-C6** | **0,034146** | **0,018882** | **0,083621** | **0,017275** | **0,846076** | **AFRg** |
| **DS-R-C7** | **1,00E-05** | **0,007297** | **0,167979** | **0,017981** | **0,806733** | **AFRg** |
| **DS-R-C8** | **0,102998** | **0,020989** | **0,064962** | **0,108423** | **0,702628** | **AFRg** |

| | | | | | | |
|---|---|---|---|---|---|---|
| **DS-R-C9** | **1,00E-05** | **0,009353** | **0,139778** | **1,00E-05** | **0,850849** | **AFRg** |
| **NH0000513** | **0,00513** | **1,80E-05** | **0,983931** | **1,00E-05** | **0,010911** | **EURg** |
| **NH0000580** | **0,160642** | **1,00E-05** | **0,804785** | **1,00E-05** | **0,034552** | **EURg** |
| **NH0001224** | **0,104829** | **1,00E-05** | **0,889341** | **1,00E-05** | **0,00581** | **EURg** |
| **NH0001391** | **0,124087** | **0,000883** | **0,851615** | **1,00E-05** | **0,023405** | **EURg** |
| **NH0001540** | **0,086222** | **0,007773** | **0,876216** | **1,00E-05** | **0,029779** | **EURg** |
| **NH0001541** | **0,235948** | **0,007439** | **0,716304** | **0,010923** | **0,029386** | **EURg** |
| **NH0001542** | **0,042587** | **1,00E-05** | **0,937983** | **1,00E-05** | **0,019409** | **EURg** |
| **NH0001543** | **0,022948** | **1,00E-05** | **0,135066** | **1,00E-05** | **0,841966** | **AFRg** |
| **NH0001544** | **0,005027** | **1,00E-05** | **0,043105** | **1,00E-05** | **0,951848** | **AFRg** |
| **NH0001545** | **1,00E-05** | **0,006631** | **0,084214** | **0,00652** | **0,902626** | **AFRg** |
| **NH0001669** | **1,00E-05** | **1,00E-05** | **0,0561** | **0,020785** | **0,923096** | **AFRg** |
| **NH0001670** | **1,00E-05** | **1,00E-05** | **0,012669** | **1,00E-05** | **0,987301** | **AFRg** |
| **NH0001671** | **1,00E-05** | **1,00E-05** | **0,262558** | **0,038538** | **0,698884** | **Not assigned** |
| **NH0001672** | **1,00E-05** | **1,00E-05** | **0,0529** | **1,00E-05** | **0,94707** | **AFRg** |
| **NH0001673** | **1,00E-05** | **0,010658** | **0,183279** | **1,00E-05** | **0,806043** | **AFRg** |

| | | | | | | |
|---|---|---|---|---|---|---|
| NH0001674 | 0,024721 | 1,00E-05 | 0,222574 | 0,012694 | 0,740001 | AFRg |
| NH0001675 | 0,044651 | 1,00E-05 | 0,955319 | 1,00E-05 | 1,00E-05 | EURg |
| NH0001676 | 0,014646 | 1,00E-05 | 0,940552 | 1,00E-05 | 0,044783 | EURg |
| NH0001679 | 0,115583 | 1,60E-05 | 0,848025 | 1,00E-05 | 0,036366 | EURg |
| NH0001680 | 0,091027 | 1,00E-05 | 0,877002 | 1,00E-05 | 0,031951 | EURg |
| NH0001681 | 0,105678 | 1,00E-05 | 0,85181 | 0,002068 | 0,040435 | EURg |
| NH0001682 | 0,083741 | 1,00E-05 | 0,848876 | 0,026362 | 0,041011 | EURg |
| NH0001719 | 0,09246 | 0,009484 | 0,874964 | 1,00E-05 | 0,023082 | EURg |
| NH0001798 | 0,167525 | 1,00E-05 | 0,787831 | 1,00E-05 | 0,044624 | EURg |
| NH0001799_set6 | 0,100873 | 1,00E-05 | 0,879428 | 1,00E-05 | 0,019679 | EURg |
| NH0001800_set6 | 0,068769 | 0,009262 | 0,891009 | 1,00E-05 | 0,030949 | EURg |
| NH0001801 | 0,042316 | 1,00E-05 | 0,947299 | 1,00E-05 | 0,010365 | EURg |
| NH0001802 | 0,057761 | 1,00E-05 | 0,914755 | 1,00E-05 | 0,027464 | EURg |
| NH0001803 | 0,048514 | 1,00E-05 | 0,941065 | 0,010401 | 1,00E-05 | EURg |
| NH0001804 | 0,150626 | 1,00E-05 | 0,822979 | 1,00E-05 | 0,026375 | EURg |
| NH0001805 | 0,044622 | 1,00E-05 | 0,951978 | 0,003381 | 1,00E-05 | EURg |

| | | | | | | |
|---|---|---|---|---|---|---|
| **NH0001806** | **0,009471** | **1,00E-05** | **0,990499** | **1,00E-05** | **1,00E-05** | **EURg** |
| **NH0001807** | **0,084778** | **1,00E-05** | **0,906713** | **1,00E-05** | **0,008489** | **EURg** |
| **NH0001808** | **1,00E-05** | **1,00E-05** | **0,523556** | **0,002067** | **0,474357** | **Not assigned** |
| **NH0001867** | **0,101456** | **0,002424** | **0,832766** | **0,003912** | **0,059443** | **EURg** |
| **NH0001868** | **0,106156** | **0,019606** | **0,827429** | **0,020974** | **0,025836** | **EURg** |
| **NH0001869** | **0,036679** | **1,00E-05** | **0,940451** | **0,021141** | **0,00172** | **EURg** |
| **NH0001870** | **0,065086** | **1,00E-05** | **0,928755** | **1,00E-05** | **0,006139** | **EURg** |
| **NH0001871** | **0,081406** | **0,00509** | **0,88459** | **1,00E-05** | **0,028905** | **EURg** |
| **NH0001872** | **0,207328** | **1,00E-05** | **0,751812** | **0,006389** | **0,034461** | **EURg** |
| **NH0001873** | **0,050041** | **1,00E-05** | **0,947893** | **1,00E-05** | **0,002045** | **EURg** |
| **NH0001874** | **0,074734** | **0,010134** | **0,90537** | **0,001315** | **0,008447** | **EURg** |
| **NH0001875** | **0,025197** | **1,20E-05** | **0,905483** | **1,00E-05** | **0,069298** | **EURg** |
| **NH0001876** | **0,061474** | **1,00E-05** | **0,938496** | **1,00E-05** | **1,00E-05** | **EURg** |
| **NH0001877** | **0,062734** | **1,00E-05** | **0,927992** | **1,00E-05** | **0,009254** | **EURg** |
| **NH0001878** | **0,019726** | **1,00E-05** | **0,980244** | **1,00E-05** | **1,00E-05** | **EURg** |
| **NH0001879** | **0,035472** | **1,80E-05** | **0,957858** | **1,00E-05** | **0,006642** | **EURg** |

| | | | | | | |
|---|---|---|---|---|---|---|
| **NH0001880** | **0,013986** | **1,00E-05** | **0,9762** | **1,00E-05** | **0,009794** | **EURg** |
| **NH0001881** | **0,103017** | **1,00E-05** | **0,889373** | **1,00E-05** | **0,007591** | **EURg** |
| **NH0001882** | **0,060286** | **1,00E-05** | **0,928917** | **1,00E-05** | **0,010777** | **EURg** |
| **NH0001883** | **0,08178** | **1,00E-05** | **0,910421** | **1,00E-05** | **0,007779** | **EURg** |
| **NH0001884** | **0,026802** | **1,00E-05** | **0,634963** | **0,313236** | **0,02499** | **Not assigned** |
| **NH0001885** | **0,088452** | **1,00E-05** | **0,900444** | **1,00E-05** | **0,011084** | **EURg** |
| **NH0001886** | **0,049421** | **1,00E-05** | **0,944392** | **0,006167** | **1,00E-05** | **EURg** |
| **NH0001887** | **0,280352** | **1,00E-05** | **0,692127** | **0,014457** | **0,013054** | **Not assigned** |
| **NH0001888** | **1,00E-05** | **0,005623** | **0,116589** | **1,00E-05** | **0,877768** | **AFRg** |
| **NH0001889** | **1,00E-05** | **1,00E-05** | **0,302691** | **0,007798** | **0,689491** | **Not assigned** |
| **NH0002026** | **0,099572** | **1,00E-05** | **0,746487** | **0,01657** | **0,137361** | **EURg** |
| **NH0002027** | **0,119607** | **1,00E-05** | **0,845822** | **1,00E-05** | **0,034551** | **EURg** |

Table S5. Sub-continental admixture proportion for the profiles in the NYGC P-1000 cohort

| The columns are: | |
|---|---|
| PatientID | The identifiant of the patient whom sub-continental ancestry has been infered. |
| BEB | The proportion of ancestry associated to Bengali in Bangladesh (BEB) population |
| CDX | The proportion of ancestry associated to Chinese Dai in Xishuangbanna, China (CDX) population |
| CEU | The proportion of ancestry associated to Utah residents with Northern and Western European ancestry (CEU) population |
| CHB | The proportion of ancestry associated to Han Chinese in Beijing, China (CHB) population |

| | |
|---|---|
| CHS | The proportion of ancestry associated to Han Chinese South (CHS) population |
| CLM | The proportion of ancestry associated to Colombian in Medellin, Colombia (CLM) population |
| ESN | The proportion of ancestry associated to Esan in Nigeria (ESN) population |
| FIN | The proportion of ancestry associated to Finnish in Finland (FIN) population |
| GBR | The proportion of ancestry associated to British in England and Scotland (GBR) population |
| GIH | The proportion of ancestry associated to Gujarati Indian in Houston, Texas (GIH) population |
| GWD | The proportion of ancestry associated to Gambian in Western Division, The Gambia (GWD) population |
| IBS | The proportion of ancestry associated to Iberian populations in Spain (IBS) population |
| ITU | The proportion of ancestry associated to Indian Telugu in the United Kingdom (ITU) population |
| JPT | The proportion of ancestry associated to Japanese in Tokyo, Japan (JPT) population |
| KHV | The proportion of ancestry associated to Kinh in Ho Chi Minh City, Vietnam (KHV) population |
| LWK | The proportion of ancestry associated to Luhya in Webuye, Kenya (LWK) population |
| MSL | The proportion of ancestry associated to Mende in Sierra Leone (MSL) population |
| PEL | The proportion of ancestry associated to Peruvians in Lima, Peru (PEL) population |
| PJL | The proportion of ancestry associated to Punjabi in Lahore, Pakistan (PJL) population |
| PUR | The proportion of ancestry associated to Puerto Rican in Puerto Rico (PUR) population |
| STU | The proportion of ancestry associated to Sri Lankan Tamil in the United Kingdom (STU) population |
| TSI | The proportion of ancestry associated to Toscani in Italia (TSI) population |
| YRI | The proportion of ancestry associated to Yoruba in Ibadan, Nigeria (YRI) population |

| PatientID | BEB | CDX | CEU | CHB | CHS | CLM | ESN | FIN | GBR | GIH | GWD | IBS | ITU | JPT | KHV | LWK | MSL | PEL | PJL | PUR | STU | TSI | YRI |
|---|---|---|---|---|---|---|---|---|---|---|---|---|---|---|---|---|---|---|---|---|---|---|---|
| C-01 | 1,00E-05 | 1,00E-05 | 1,00E-05 | 1,00E-05 | 1,00E-05 | 0,026317 | 0,580445 | 1,00E-05 | 1,00E-05 | 1,00E-05 | 1,00E-05 | 1,00E-05 | 0,001372 | 0,013123 | 1,00E-05 | 0,019896 | 0,353802 | 0,004885 | 1,00E-05 | 1,00E-05 | 1,00E-05 | 1,00E-05 | 1,00E-05 |
| C-02 | 1,00E-05 | 1,00E-05 | 0,042911 | 1,00E-05 | 1,00E-05 | 1,00E-05 | 0,572892 | 1,00E-05 | 1,00E-05 | 1,00E-05 | 0,091446 | 1,00E-05 | 0,00335 | 1,00E-05 | 1,00E-05 | 0,080794 | 0,208437 | 1,00E-05 | 1,00E-05 | 1,00E-05 | 1,00E-05 | 1,00E-05 | 1,00E-05 |
| C-04 | 1,00E-05 | 1,00E-05 | 0,088555 | 1,00E-05 | 1,00E-05 | 1,00E-05 | 0,552403 | 1,00E-05 | 1,30E-05 | 1,00E-05 | 0,1135 | 1,40E-05 | 1,00E-05 | 1,00E-05 | 1,00E-05 | 0,078126 | 0,146859 | 1,00E-05 | 1,00E-05 | 1,00E-05 | 0,008166 | 1,00E-05 | 0,012226 |
| C-05 | 1,00E-05 | 1,20E-05 | 1,00E-05 | 1,00E-05 | 0,001402 | 1,00E-05 | 0,64878 | 1,00E-05 | 1,00E-05 | 1,00E-05 | 1,00E-05 | 1,00E-05 | 1,00E-05 | 1,00E-05 | 1,00E-05 | 0,035257 | 0,146825 | 1,00E-05 | 1,00E-05 | 1,00E-05 | 1,00E-05 | 1,00E-05 | 0,167554 |
| C-06 | 1,00E-05 | 1,00E-05 | 1,00E-05 | 1,00E-05 | 1,00E-05 | 1,00E-05 | 0,63998 | 1,00E-05 | 1,00E-05 | 1,00E-05 | 1,00E-05 | 1,00E-05 | 1,00E-05 | 1,00E-05 | 1,00E-05 | 0,05126 | 0,224628 | 1,00E-05 | 1,00E-05 | 0,028608 | 1,00E-05 | 0,055344 | 1,00E-05 |
| C-07 | 1,00E-05 | 1,00E-05 | 1,00E-05 | 1,00E-05 | 1,00E-05 | 1,00E-05 | 0,681075 | 0,000438 | 1,00E-05 | 1,00E-05 | 1,00E-05 | 1,00E-05 | 0,004938 | 1,00E-05 | 1,00E-05 | 1,00E-05 | 0,210849 | 1,00E-05 | 1,00E-05 | 1,00E-05 | 1,00E-05 | 0,10252 | 1,00E-05 |
| C-08 | 1,00E-05 | 1,00E-05 | 1,00E-05 | 1,00E-05 | 1,00E-05 | 1,00E-05 | 0,861324 | 1,00E-05 | 1,00E-05 | 1,00E-05 | 0,010776 | 1,00E-05 | 1,00E-05 | 1,00E-05 | 1,00E-05 | 1,00E-05 | 0,071101 | 1,00E-05 | 1,00E-05 | 1,00E-05 | 1,00E-05 | 1,00E-05 | 0,05661 |
| C-09 | 1,00E-05 | 1,00E-05 | 1,00E-05 | 1,00E-05 | 1,00E-05 | 1,00E-05 | 0,320013 | 0,04902 | 1,00E-05 | 1,00E-05 | 1,00E-05 | 0,015327 | 1,00E-05 | 1,00E-05 | 1,00E-05 | 1,00E-05 | 0,508377 | 0,005036 | 1,00E-05 | 1,00E-05 | 1,00E-05 | 0,102057 | 1,00E-05 |
| C-10 | 0,008336 | 1,00E-05 | 0,032467 | 1,00E-05 | 1,00E-05 | 0,017831 | 0,467218 | 1,00E-05 | 1,00E-05 | 1,00E-05 | 0,072157 | 1,00E-05 | 1,00E-05 | 1,00E-05 | 1,00E-05 | 0,110712 | 0,241031 | 1,00E-05 | 1,00E-05 | 1,00E-05 | 1,00E-05 | 0,050098 | 1,00E-05 |
| C-11 | 1,00E-05 | 1,00E-05 | 0,035247 | 1,00E-05 | 1,00E-05 | 0,033363 | 0,501728 | 0,010167 | 1,00E-05 | 1,00E-05 | 0,063707 | 1,00E-05 | 1,00E-05 | 1,00E-05 | 1,00E-05 | 0,074389 | 0,255326 | 1,00E-05 | 1,00E-05 | 1,00E-05 | 1,00E-05 | 1,00E-05 | 0,025923 |
| C-12 | 1,00E-05 | 1,00E-05 | 1,00E-05 | 1,00E-05 | 1,00E-05 | 1,00E-05 | 0,464192 | 1,00E-05 | 0,051964 | 1,00E-05 | 0,093525 | 1,00E-05 | 0,006709 | 1,00E-05 | 1,00E-05 | 0,148898 | 0,157638 | 1,00E-05 | 1,00E-05 | 1,00E-05 | 1,00E-05 | 1,00E-05 | 0,076915 |
| C-14 | 0,015915 | 1,00E-05 | 1,00E-05 | 1,00E-05 | 1,00E-05 | 1,00E-05 | 0,372438 | 1,00E-05 | 0,223756 | 1,00E-05 | 0,120603 | 1,00E-05 | 1,00E-05 | 1,00E-05 | 1,00E-05 | 0,020055 | 0,126016 | 0,022999 | 1,00E-05 | 0,085219 | 1,00E-05 | 0,012859 | 1,00E-05 |
| C-15 | 0,005071 | 0,005918 | 0,082028 | 1,00E-05 | 1,00E-05 | 1,00E-05 | 0,402035 | 1,00E-05 | 1,00E-05 | 1,00E-05 | 0,057806 | 1,00E-05 | 1,00E-05 | 0,005374 | 1,00E-05 | 0,093701 | 0,347918 | 1,00E-05 | 1,00E-05 | 1,00E-05 | 1,00E-05 | 1,00E-05 | 1,00E-05 |
| C-16 | 1,00E-05 | 1,00E-05 | 1,00E-05 | 1,00E-05 | 1,00E-05 | 1,00E-05 | 0,521653 | 0,05204 | 1,00E-05 | 1,00E-05 | 0,00613 | 1,00E-05 | 1,00E-05 | 1,00E-05 | 1,00E-05 | 0,033485 | 0,236681 | 1,00E-05 | 1,00E-05 | 1,00E-05 | 1,00E-05 | 0,116543 | 0,033308 |
| C-17 | 1,00E-05 | 1,00E-05 | 1,00E-05 | 1,00E-05 | 1,00E-05 | 1,00E-05 | 0,528715 | 1,00E-05 | 0,014867 | 1,00E-05 | 0,077823 | 1,00E-05 | 1,00E-05 | 1,00E-05 | 1,00E-05 | 0,085381 | 0,282647 | 1,00E-05 | 1,00E-05 | 1,00E-05 | 1,00E-05 | 1,00E-05 | 0,010398 |
| C-18 | 1,00E-05 | 1,00E-05 | 0,063869 | 1,00E-05 | 1,00E-05 | 1,00E-05 | 0,465981 | 0,024801 | 0,064117 | 1,00E-05 | 0,014922 | 1,00E-05 | 1,00E-05 | 1,00E-05 | 1,00E-05 | 0,034248 | 0,331902 | 1,00E-05 | 1,00E-05 | 1,00E-05 | 1,00E-05 | 1,00E-05 | 1,00E-05 |
| C-19 | 1,00E-05 | 1,00E-05 | 0,011047 | 1,00E-05 | 1,00E-05 | 0,017767 | 0,477528 | 1,00E-05 | 2,00E-05 | 1,00E-05 | 0,030217 | 0,080444 | 1,00E-05 | 1,00E-05 | 1,00E-05 | 0,049333 | 0,190772 | 0,066981 | 0,024868 | 1,00E-05 | 1,00E-05 | 1,00E-05 | 0,050903 |
| C-20 | 1,00E-05 | 1,00E-05 | 1,00E-05 | 1,00E-05 | 1,00E-05 | 1,00E-05 | 0,440921 | 0,01608 | 0,208436 | 1,00E-05 | 0,050171 | 0,006353 | 1,00E-05 | 1,00E-05 | 0,006545 | 1,00E-05 | 0,267599 | 1,00E-05 | 1,00E-05 | 1,00E-05 | 0,003745 | 1,00E-05 | 1,00E-05 |

| | | | | | | | | | | | | | | | | | | | | | | | |
|---|---|---|---|---|---|---|---|---|---|---|---|---|---|---|---|---|---|---|---|---|---|---|---|
| C-22 | 0,013373 | 1,00E-05 | 0,115953 | 1,00E-05 | 1,00E-05 | 0,107647 | 0,375942 | 0,024712 | 1,00E-05 | 1,00E-05 | 0,011249 | 0,014348 | 1,00E-05 | 1,00E-05 | 1,00E-05 | 0,05162 | 0,284392 | 1,00E-05 | 1,00E-05 | 1,00E-05 | 0,000633 | 1,00E-05 | 1,00E-05 |
| C-23 | 1,00E-05 | 1,00E-05 | 1,00E-05 | 1,00E-05 | 1,00E-05 | 1,00E-05 | 0,731571 | 1,00E-05 | 1,00E-05 | 1,00E-05 | 0,003879 | 1,00E-05 | 1,00E-05 | 1,00E-05 | 1,00E-05 | 0,055456 | 0,208904 | 1,00E-05 | 1,00E-05 | 1,00E-05 | 1,00E-05 | 1,00E-05 | 1,00E-05 |
| C-24 | 1,00E-05 | 1,00E-05 | 1,00E-05 | 1,00E-05 | 0,000488 | 1,00E-05 | 0,630205 | 1,00E-05 | 1,00E-05 | 1,00E-05 | 0,171775 | 1,00E-05 | 1,00E-05 | 1,00E-05 | 1,00E-05 | 0,12383 | 0,032224 | 1,00E-05 | 1,00E-05 | 1,00E-05 | 1,00E-05 | 1,00E-05 | 0,041308 |
| C-25 | 1,00E-05 | 0,005667 | 1,00E-05 | 1,00E-05 | 1,00E-05 | 1,00E-05 | 0,795071 | 1,00E-05 | 1,00E-05 | 1,00E-05 | 0,018535 | 1,00E-05 | 1,00E-05 | 1,00E-05 | 1,00E-05 | 0,062133 | 0,056493 | 1,00E-05 | 1,00E-05 | 1,00E-05 | 1,00E-05 | 1,00E-05 | 0,061931 |
| C-26 | 1,00E-05 | 0,005393 | 0,138959 | 1,00E-05 | 1,00E-05 | 1,00E-05 | 0,496196 | 1,00E-05 | 1,00E-05 | 1,00E-05 | 1,00E-05 | 1,00E-05 | 1,00E-05 | 1,00E-05 | 1,00E-05 | 0,009473 | 0,349799 | 1,00E-05 | 1,00E-05 | 1,00E-05 | 1,00E-05 | 1,00E-05 | 1,00E-05 |
| C-27 | 1,00E-05 | 0,008533 | 1,00E-05 | 1,00E-05 | 1,00E-05 | 0,001729 | 0,696864 | 1,00E-05 | 1,00E-05 | 1,00E-05 | 0,062276 | 1,00E-05 | 4,10E-05 | 1,00E-05 | 1,00E-05 | 0,054029 | 0,103901 | 1,00E-05 | 1,00E-05 | 1,00E-05 | 1,00E-05 | 0,072477 | 1,00E-05 |
| C-28 | 1,00E-05 | 1,00E-05 | 1,00E-05 | 1,00E-05 | 1,00E-05 | 1,00E-05 | 0,297764 | 1,00E-05 | 0,275743 | 1,00E-05 | 0,048417 | 0,103516 | 1,00E-05 | 1,00E-05 | 1,00E-05 | 0,011258 | 0,262348 | 1,00E-05 | 0,000793 | 1,00E-05 | 1,00E-05 | 1,00E-05 | 1,00E-05 |
| C-29 | 1,00E-05 | 1,00E-05 | 0,055208 | 1,00E-05 | 1,00E-05 | 0,007748 | 0,728119 | 1,00E-05 | 1,00E-05 | 1,00E-05 | 0,055491 | 1,00E-05 | 1,00E-05 | 1,00E-05 | 1,00E-05 | 0,079511 | 0,068685 | 1,00E-05 | 1,00E-05 | 1,00E-05 | 1,00E-05 | 1,00E-05 | 0,005077 |
| C-31 | 1,00E-05 | 1,00E-05 | 0,76652 | 1,00E-05 | 1,00E-05 | 1,00E-05 | 1,00E-05 | 1,00E-05 | 1,40E-05 | 1,00E-05 | 0,006507 | 0,152282 | 1,00E-05 | 1,00E-05 | 1,00E-05 | 1,00E-05 | 0,003633 | 1,00E-05 | 0,070874 | 1,00E-05 | 1,00E-05 | 1,00E-05 | 1,00E-05 |
| C-32 | 1,00E-05 | 1,00E-05 | 0,696199 | 1,00E-05 | 1,00E-05 | 1,00E-05 | 1,00E-05 | 0,00581 | 0,109675 | 1,00E-05 | 1,00E-05 | 0,145978 | 1,00E-05 | 1,00E-05 | 1,00E-05 | 1,00E-05 | 0,011099 | 1,00E-05 | 0,03107 | 1,00E-05 | 1,00E-05 | 1,00E-05 | 1,00E-05 |
| C-33 | 1,00E-05 | 1,00E-05 | 1,00E-05 | 1,00E-05 | 1,00E-05 | 1,00E-05 | 1,00E-05 | 1,00E-05 | 1,00E-05 | 1,00E-05 | 0,017217 | 1,00E-05 | 1,00E-05 | 1,00E-05 | 1,00E-05 | 1,00E-05 | 1,00E-05 | 1,00E-05 | 1,00E-05 | 0,031806 | 0,058635 | 0,892152 | 1,00E-05 |
| C-36 | 1,00E-05 | 1,00E-05 | 0,837212 | 1,00E-05 | 1,00E-05 | 1,00E-05 | 1,00E-05 | 1,00E-05 | 1,00E-05 | 1,00E-05 | 0,003461 | 0,112987 | 1,00E-05 | 1,00E-05 | 1,00E-05 | 1,00E-05 | 1,00E-05 | 1,00E-05 | 0,046151 | 1,00E-05 | 1,00E-05 | 1,00E-05 | 1,00E-05 |
| C-37 | 1,00E-05 | 1,00E-05 | 0,18561 | 1,00E-05 | 1,00E-05 | 1,70E-05 | 1,00E-05 | 1,00E-05 | 0,731074 | 1,00E-05 | 0,016301 | 1,00E-05 | 1,00E-05 | 1,00E-05 | 1,00E-05 | 1,00E-05 | 1,00E-05 | 1,00E-05 | 0,038744 | 1,80E-05 | 0,001964 | 0,026123 | 1,00E-05 |
| C-38 | 1,00E-05 | 1,00E-05 | 1,00E-05 | 1,00E-05 | 1,00E-05 | 0,092296 | 0,011002 | 0,005613 | 1,00E-05 | 1,00E-05 | 0,062776 | 0,110108 | 1,00E-05 | 1,00E-05 | 1,00E-05 | 1,00E-05 | 1,00E-05 | 0,048264 | 0,058007 | 0,611784 | 1,00E-05 | 1,00E-05 | 1,00E-05 |
| C-39 | 1,00E-05 | 1,00E-05 | 1,00E-05 | 1,00E-05 | 1,00E-05 | 1,00E-05 | 0,01827 | 1,00E-05 | 1,00E-05 | 1,00E-05 | 0,012338 | 1,00E-05 | 1,00E-05 | 1,00E-05 | 1,00E-05 | 1,00E-05 | 1,00E-05 | 1,00E-05 | 0,081692 | 1,00E-05 | 1,00E-05 | 0,88751 | 1,00E-05 |
| C-41 | 0,028613 | 0,305743 | 1,00E-05 | 1,00E-05 | 0,237858 | 1,00E-05 | 0,005409 | 1,00E-05 | 1,00E-05 | 1,00E-05 | 1,00E-05 | 1,00E-05 | 1,00E-05 | 1,20E-05 | 0,390592 | 1,00E-05 | 1,00E-05 | 1,00E-05 | 0,015223 | 1,00E-05 | 0,016401 | 1,00E-05 | 1,00E-05 |
| C-42 | 1,00E-05 | 1,00E-05 | 1,00E-05 | 0,299826 | 0,275631 | 1,00E-05 | 1,10E-05 | 1,00E-05 | 1,00E-05 | 1,00E-05 | 0,009256 | 1,00E-05 | 1,00E-05 | 0,415096 | 1,00E-05 | 1,00E-05 | 1,00E-05 | 1,00E-05 | 1,00E-05 | 1,00E-05 | 1,00E-05 | 1,00E-05 | 1,00E-05 |
| C-43 | 1,00E-05 | 1,00E-05 | 0,095035 | 1,00E-05 | 1,00E-05 | 0,026324 | 0,522725 | 0,000849 | 1,00E-05 | 1,00E-05 | 0,142148 | 1,00E-05 | 0,009074 | 1,00E-05 | 1,00E-05 | 0,101199 | 0,102497 | 1,00E-05 | 1,00E-05 | 1,00E-05 | 1,00E-05 | 1,00E-05 | 1,00E-05 |
| C-44 | 1,00E-05 | 1,00E-05 | 1,00E-05 | 1,00E-05 | 1,00E-05 | 0,0208 | 1,00E-05 | 1,00E-05 | 1,00E-05 | 1,00E-05 | 0,021369 | 0,02041 | 0,070582 | 1,00E-05 | 1,00E-05 | 1,00E-05 | 1,00E-05 | 1,00E-05 | 1,00E-05 | 0,034272 | 1,00E-05 | 0,832397 | 1,00E-05 |
| C-45 | 1,00E-05 | 1,00E-05 | 1,00E-05 | 1,00E-05 | 1,00E-05 | 0,023647 | 0,383394 | 1,00E-05 | 1,00E-05 | 1,00E-05 | 0,041622 | 1,00E-05 | 1,00E-05 | 1,00E-05 | 1,00E-05 | 0,144802 | 0,39645 | 1,00E-05 | 1,00E-05 | 1,00E-05 | 1,00E-05 | 1,00E-05 | 0,009914 |
| C-46 | 0,01828 | 1,00E-05 | 1,00E-05 | 1,00E-05 | 1,00E-05 | 0,04096 | 0,582331 | 0,002137 | 0,057935 | 1,00E-05 | 1,00E-05 | 0,033429 | 1,00E-05 | 1,00E-05 | 1,00E-05 | 0,043297 | 0,163168 | 0,058324 | 1,00E-05 | 1,00E-05 | 1,00E-05 | 1,00E-05 | 1,00E-05 |
| C-47 | 0,014344 | 1,00E-05 | 1,00E-05 | 1,00E-05 | 1,00E-05 | 0,012227 | 0,332184 | 1,00E-05 | 0,018525 | 1,00E-05 | 0,201399 | 0,000438 | 1,00E-05 | 0,018083 | 6,10E-05 | 0,020314 | 0,090865 | 1,00E-05 | 0,091579 | 1,00E-05 | 0,15016 | 1,00E-05 | 0,049721 |
| C-48 | 0,077441 | 1,00E-05 | 1,00E-05 | 1,00E-05 | 1,00E-05 | 1,00E-05 | 0,396499 | 1,00E-05 | 1,00E-05 | 1,00E-05 | 0,059155 | 1,00E-05 | 1,00E-05 | 1,00E-05 | 0,000207 | 0,007494 | 0,270312 | 1,00E-05 | 0,02777 | 1,00E-05 | 1,00E-05 | 0,044335 | 0,116647 |
| C-49 | 0,02375 | 0,000763 | 1,00E-05 | 1,00E-05 | 1,00E-05 | 0,004019 | 0,271917 | 1,00E-05 | 0,22108 | 1,00E-05 | 0,209354 | 1,00E-05 | 1,00E-05 | 1,00E-05 | 1,00E-05 | 0,036832 | 0,200567 | 0,008861 | 1,00E-05 | 1,00E-05 | 1,00E-05 | 0,022726 | 1,00E-05 |
| C-50 | 1,00E-05 | 1,00E-05 | 0,148852 | 1,00E-05 | 0,003179 | 1,00E-05 | 0,48174 | 1,00E-05 | 1,00E-05 | 1,00E-05 | 0,028536 | 1,00E-05 | 1,00E-05 | 1,00E-05 | 1,00E-05 | 0,12949 | 0,200705 | 0,007339 | 1,00E-05 | 1,00E-05 | 1,00E-05 | 1,00E-05 | 1,00E-05 |
| C-51 | 1,00E-05 | 1,00E-05 | 1,00E-05 | 0,00095 | 1,00E-05 | 1,00E-05 | 0,642594 | 0,004109 | 0,132903 | 1,00E-05 | 0,052185 | 1,00E-05 | 0,002135 | 1,00E-05 | 1,00E-05 | 0,017582 | 0,141529 | 1,00E-05 | 0,002665 | 0,003219 | 1,00E-05 | 1,00E-05 | 1,00E-05 |
| C-52 | 1,00E-05 | 1,00E-05 | 0,933768 | 1,00E-05 | 1,00E-05 | 1,00E-05 | 0,012087 | 1,00E-05 | 1,00E-05 | 1,00E-05 | 1,00E-05 | 1,00E-05 | 1,00E-05 | 1,00E-05 | 1,00E-05 | 1,00E-05 | 1,00E-05 | 1,00E-05 | 0,053945 | 1,00E-05 | 1,00E-05 | 1,00E-05 | 1,00E-05 |
| C-53 | 1,00E-05 | 1,00E-05 | 1,00E-05 | 1,00E-05 | 1,00E-05 | 1,00E-05 | 0,577011 | 1,00E-05 | 0,01071 | 1,00E-05 | 0,124549 | 0,025231 | 1,00E-05 | 1,00E-05 | 1,00E-05 | 1,00E-05 | 1,00E-05 | 1,00E-05 | 1,00E-05 | 0,015803 | 1,00E-05 | 1,00E-05 | 0,246527 |
| C-54 | 1,00E-05 | 1,00E-05 | 1,00E-05 | 1,00E-05 | 1,00E-05 | 1,00E-05 | 1,00E-05 | 1,00E-05 | 1,00E-05 | 1,00E-05 | 0,022817 | 1,00E-05 | 1,00E-05 | 1,00E-05 | 1,00E-05 | 1,00E-05 | 0,000887 | 1,00E-05 | 0,037052 | 0,07161 | 1,00E-05 | 0,867454 | 1,00E-05 |
| C-55 | 0,027551 | 1,00E-05 | 1,00E-05 | 1,00E-05 | 1,00E-05 | 1,00E-05 | 0,231138 | 1,00E-05 | 0,28255 | 1,00E-05 | 0,056569 | 1,00E-05 | 1,00E-05 | 1,00E-05 | 1,00E-05 | 1,00E-05 | 1,00E-05 | 1,00E-05 | 1,00E-05 | 0,402012 | 1,00E-05 | 1,00E-05 | 1,00E-05 |
| C-56 | 1,00E-05 | 1,00E-05 | 1,00E-05 | 1,00E-05 | 1,00E-05 | 1,00E-05 | 1,00E-05 | 1,00E-05 | 1,00E-05 | 1,00E-05 | 0,043168 | 1,00E-05 | 0,061602 | 1,00E-05 | 1,00E-05 | 1,00E-05 | 0,00213 | 1,00E-05 | 1,00E-05 | 0,009466 | 1,00E-05 | 0,883453 | 1,00E-05 |
| C-57 | 0,000743 | 1,00E-05 | 0,06338 | 1,00E-05 | 1,00E-05 | 1,00E-05 | 0,232653 | 1,00E-05 | 1,00E-05 | 1,00E-05 | 0,121327 | 1,00E-05 | 1,00E-05 | 1,00E-05 | 1,00E-05 | 0,143003 | 0,438725 | 1,00E-05 | 1,00E-05 | 1,00E-05 | 1,00E-05 | 1,00E-05 | 1,00E-05 |
| C-58 | 1,00E-05 | 1,00E-05 | 1,00E-05 | 1,00E-05 | 1,00E-05 | 1,00E-05 | 0,457537 | 0,025307 | 1,00E-05 | 1,00E-05 | 1,00E-05 | 0,153283 | 1,00E-05 | 0,00238 | 1,00E-05 | 0,023589 | 0,322616 | 1,00E-05 | 0,013746 | 0,001391 | 1,00E-05 | 1,00E-05 | 1,00E-05 |
| C-59 | 1,00E-05 | 1,00E-05 | 1,00E-05 | 1,00E-05 | 0,007641 | 1,00E-05 | 0,386483 | 3,50E-05 | 0,042571 | 1,00E-05 | 0,244328 | 0,007171 | 1,00E-05 | 1,00E-05 | 1,00E-05 | 0,063544 | 0,2404 | 1,00E-05 | 1,00E-05 | 0,007686 | 1,00E-05 | 1,00E-05 | 1,00E-05 |
| C-60 | 1,00E-05 | 1,00E-05 | 1,10E-05 | 1,00E-05 | 1,00E-05 | 1,00E-05 | 0,471052 | 0,007274 | 0,047008 | 1,00E-05 | 0,007095 | 1,00E-05 | 1,00E-05 | 0,003742 | 0,01279 | 0,04413 | 0,245712 | 0,009867 | 0,003732 | 0,054524 | 1,00E-05 | 1,00E-05 | 0,092964 |
| C-62 | 1,00E-05 | 1,00E-05 | 0,092597 | 1,00E-05 | 1,00E-05 | 0,042923 | 0,46542 | 1,00E-05 | 1,00E-05 | 1,00E-05 | 0,095193 | 1,00E-05 | 1,00E-05 | 1,00E-05 | 1,00E-05 | 0,076058 | 0,199351 | 1,00E-05 | 1,00E-05 | 1,00E-05 | 0,028298 | 1,00E-05 | 1,00E-05 |
| C-63 | 1,00E-05 | 1,00E-05 | 0,202606 | 1,00E-05 | 1,00E-05 | 1,00E-05 | 0,414492 | 1,00E-05 | 1,00E-05 | 1,00E-05 | 0,014366 | 0,127736 | 1,00E-05 | 1,00E-05 | 1,00E-05 | 0,007013 | 0,167056 | 0,011578 | 0,006939 | 0,048075 | 1,00E-05 | 1,00E-05 | 1,00E-05 |
| C-64 | 1,00E-05 | 1,00E-05 | 1,00E-05 | 1,00E-05 | 1,00E-05 | 0,016917 | 0,633657 | 0,014394 | 1,00E-05 | 1,00E-05 | 0,052482 | 1,00E-05 | 1,00E-05 | 1,00E-05 | 1,00E-05 | 0,071579 | 0,173947 | 1,00E-05 | 1,00E-05 | 1,00E-05 | 1,00E-05 | 1,00E-05 | 0,036864 |
| C-65 | 1,00E-05 | 1,00E-05 | 1,00E-05 | 1,00E-05 | 1,00E-05 | 1,00E-05 | 0,32948 | 1,00E-05 | 0,26081 | 1,00E-05 | 0,279421 | 1,00E-05 | 1,00E-05 | 1,00E-05 | 1,00E-05 | 0,061459 | 0,058885 | 1,00E-05 | 1,00E-05 | 1,00E-05 | 0,009776 | 1,00E-05 | 1,00E-05 |
| C-67 | 1,00E-05 | 1,10E-05 | 1,00E-05 | 1,00E-05 | 1,00E-05 | 1,00E-05 | 0,294534 | 0,0187 | 0,083266 | 1,00E-05 | 0,125216 | 0,025312 | 1,00E-05 | 1,00E-05 | 0,010671 | 0,11505 | 0,302954 | 0,002447 | 1,00E-05 | 0,021719 | 1,00E-05 | 1,00E-05 | 1,00E-05 |
| C-68 | 1,00E-05 | 1,00E-05 | 0,031404 | 1,00E-05 | 1,00E-05 | 1,00E-05 | 0,631394 | 1,00E-05 | 1,00E-05 | 1,00E-05 | 0,083665 | 1,00E-05 | 1,00E-05 | 1,00E-05 | 1,00E-05 | 0,054726 | 0,191923 | 1,00E-05 | 1,00E-05 | 1,20E-05 | 0,006715 | 1,00E-05 | 1,00E-05 |

Table S6. Sub-continental admixture proportion for the profiles in the CSHL FFPE cohort

| The columns are: | |
|---|---|
| PatientID | The identifiant of the patient whom sub-continental ancestry has been infered. |
| GBR.CEU | The proportion of ancestry associated to British in England and Scotland (GBR) or to Utah residents with Northern and Western European ancestry (CEU) populations |
| FIN | The proportion of ancestry associated to Finnish in Finland (FIN) population |
| CHS | The proportion of ancestry associated to Han Chinese South (CHS) population |
| PUR.CLM.MXL | The proportion of ancestry associated to Puerto Rican in Puerto Rico (PUR) or to Medellin, Colombia (CLM) or to Mexican Ancestry in Los Angeles, California (MXL) populations |
| CDX | The proportion of ancestry associated to Chinese Dai in Xishuangbanna, China (CDX) population |
| IBS | The proportion of ancestry associated to Iberian populations in Spain (IBS) population |
| PEL.MXL | The proportion of ancestry associated to Peruvians in Lima, Peru (PEL) or to Mexican Ancestry in Los Angeles, California (MXL) populations |
| KHV | The proportion of ancestry associated to Kinh in Ho Chi Minh City, Vietnam (KHV) population |
| GWD | The proportion of ancestry associated to Gambian in Western Division, The Gambia (GWD) population |
| PJL.BEB.STU.ITU | The proportion of ancestry associated to Punjabi in Lahore, Pakistan (PJL) or to Bengali in Bangladesh (BEB) or to Sri Lankan Tamil in the United Kingdom (STU) or to Indian Telugu in the United Kingdom (ITU) populations |
| ESN | The proportion of ancestry associated to Esan in Nigeria (ESN) population |
| MSL | The proportion of ancestry associated to Mende in Sierra Leone (MSL) population |
| YRI | The proportion of ancestry associated to Yoruba in Ibadan, Nigeria (YRI) population |
| CHB | The proportion of ancestry associated to Han Chinese in Beijing, China (CHB) population |
| JPT | The proportion of ancestry associated to Japanese in Tokyo, Japan (JPT) population |
| LWK | The proportion of ancestry associated to Luhya in Webuye, Kenya (LWK) population |
| TSI | The proportion of ancestry associated to Toscani in Italia (TSI) population |
| GIH | The proportion of ancestry associated to Gujarati Indian in Houston, Texas (GIH) population |

| PatientID | GBR.CEU | FIN | CHS | PUR.CLM.MXL | CDX | IBS | PEL.MXL | KHV | GWD | PJL.BEB.STU.ITU | ESN | MSL | YRI | CHB | JPT | LWK | TSI | GIH |
|---|---|---|---|---|---|---|---|---|---|---|---|---|---|---|---|---|---|---|
| DS-R-C1 | 0,098284 | 1,00E-05 | 1,00E-05 | 1,00E-05 | 1,00E-05 | 1,00E-05 | 0,000894 | 1,00E-05 | 1,00E-05 | 0,168089 | 0,728353 | 1,00E-05 | 1,00E-05 | 1,00E-05 | 1,00E-05 | 1,00E-05 | 1,00E-05 | 0,004251 |
| DS-R-C10 | 1,00E-05 | 1,00E-05 | 1,00E-05 | 1,00E-05 | 1,00E-05 | 1,00E-05 | 1,00E-05 | 1,00E-05 | 1,00E-05 | 1,00E-05 | 1,00E-05 | 0,606413 | 0,258518 | 1,00E-05 | 1,00E-05 | 0,109521 | 0,025408 | 1,00E-05 |
| DS-R-C11 | 1,00E-05 | 1,00E-05 | 1,00E-05 | 1,00E-05 | 1,00E-05 | 1,00E-05 | 1,00E-05 | 1,00E-05 | 1,00E-05 | 1,00E-05 | 0,441673 | 0,558167 | 1,00E-05 | 1,00E-05 | 1,00E-05 | 1,00E-05 | 1,00E-05 | 1,00E-05 |
| DS-R-C12 | 0,133463 | 1,00E-05 | 1,00E-05 | 1,00E-05 | 1,00E-05 | 1,00E-05 | 1,00E-05 | 1,00E-05 | 1,00E-05 | 1,00E-05 | 0,866377 | 1,00E-05 | 1,00E-05 | 1,00E-05 | 1,00E-05 | 1,00E-05 | 1,00E-05 | 1,00E-05 |
| DS-R-C13 | 1,00E-05 | 1,00E-05 | 1,70E-05 | 1,00E-05 | 1,00E-05 | 1,00E-05 | 0,019614 | 1,00E-05 | 0,003599 | 1,00E-05 | 0,205959 | 0,402152 | 0,174011 | 1,00E-05 | 1,00E-05 | 0,134923 | 0,059624 | 1,00E-05 |
| DS-R-C14 | 0,031651 | 1,00E-05 | 1,00E-05 | 1,00E-05 | 1,00E-05 | 1,00E-05 | 1,00E-05 | 1,00E-05 | 1,00E-05 | 1,00E-05 | 0,133208 | 0,465634 | 0,260673 | 1,00E-05 | 1,00E-05 | 0,040496 | 0,068219 | 1,00E-05 |
| DS-R-C15 | 1,00E-05 | 1,00E-05 | 1,00E-05 | 1,00E-05 | 1,00E-05 | 1,00E-05 | 1,00E-05 | 1,00E-05 | 1,00E-05 | 1,00E-05 | 0,058452 | 0,458468 | 0,270626 | 0,006469 | 1,00E-05 | 0,11002 | 0,095843 | 1,00E-05 |
| DS-R-C16 | 0,193195 | 1,00E-05 | 1,00E-05 | 1,00E-05 | 1,00E-05 | 1,00E-05 | 0,021937 | 1,00E-05 | 1,00E-05 | 1,00E-05 | 0,17133 | 0,461587 | 1,00E-05 | 1,00E-05 | 1,00E-05 | 0,097789 | 0,054042 | 1,00E-05 |
| DS-R-C17 | 0,043681 | 1,00E-05 | 0,010489 | 0,086827 | 1,00E-05 | 1,00E-05 | 1,00E-05 | 1,00E-05 | 1,00E-05 | 1,00E-05 | 0,587135 | 0,198078 | 1,00E-05 | 1,00E-05 | 1,00E-05 | 0,07367 | 1,00E-05 | 1,00E-05 |
| DS-R-C18 | 0,18701 | 1,00E-05 | 1,00E-05 | 1,00E-05 | 0,004474 | 1,00E-05 | 1,00E-05 | 1,00E-05 | 0,057833 | 1,00E-05 | 0,453439 | 0,097812 | 0,101978 | 1,00E-05 | 1,00E-05 | 0,097345 | 1,00E-05 | 1,00E-05 |
| DS-R-C19 | 1,00E-05 | 1,00E-05 | 1,00E-05 | 1,00E-05 | 1,00E-05 | 1,00E-05 | 1,00E-05 | 1,00E-05 | 1,00E-05 | 1,00E-05 | 0,340351 | 0,470362 | 1,00E-05 | 1,00E-05 | 1,10E-05 | 0,127477 | 0,061669 | 1,00E-05 |

| | | | | | | | | | | | | | | | | | | |
|---|---|---|---|---|---|---|---|---|---|---|---|---|---|---|---|---|---|---|
| DS-R-C2 | 1,00E-05 | 1,00E-05 | 1,00E-05 | 1,00E-05 | 1,00E-05 | 0,99983 | 1,00E-05 | 1,00E-05 | 1,00E-05 | 1,00E-05 | 1,00E-05 | 1,00E-05 | 1,00E-05 | 1,00E-05 | 1,00E-05 | 1,00E-05 | 1,00E-05 | 1,00E-05 |
| DS-R-C20 | 1,00E-05 | 1,00E-05 | 1,00E-05 | 1,00E-05 | 1,00E-05 | 1,00E-05 | 0,004745 | 1,00E-05 | 1,00E-05 | 1,00E-05 | 0,326777 | 0,405421 | 1,00E-05 | 1,00E-05 | 1,00E-05 | 0,195228 | 0,0677 | 1,00E-05 |
| DS-R-C21 | 1,00E-05 | 1,00E-05 | 1,00E-05 | 1,00E-05 | 1,00E-05 | 1,00E-05 | 1,00E-05 | 1,00E-05 | 1,00E-05 | 1,00E-05 | 0,418106 | 0,490011 | 1,00E-05 | 1,00E-05 | 1,00E-05 | 0,062822 | 0,028921 | 1,00E-05 |
| DS-R-C22 | 0,09596 | 1,00E-05 | 1,00E-05 | 1,00E-05 | 1,00E-05 | 1,00E-05 | 1,00E-05 | 1,00E-05 | 1,00E-05 | 1,00E-05 | 0,676502 | 0,227388 | 1,00E-05 | 1,00E-05 | 1,00E-05 | 1,00E-05 | 1,00E-05 | 1,00E-05 |
| DS-R-C23 | 0,075055 | 1,00E-05 | 0,030142 | 0,098897 | 1,00E-05 | 0,097153 | 0,046381 | 1,00E-05 | 1,00E-05 | 1,00E-05 | 0,13932 | 0,302079 | 1,00E-05 | 1,00E-05 | 1,00E-05 | 0,004237 | 0,206647 | 1,00E-05 |
| DS-R-C24 | 1,00E-05 | 1,00E-05 | 1,00E-05 | 1,00E-05 | 1,00E-05 | 1,00E-05 | 1,00E-05 | 0,023812 | 0,138425 | 1,00E-05 | 0,661442 | 0,110707 | 1,00E-05 | 1,00E-05 | 1,00E-05 | 1,00E-05 | 0,065485 | 1,00E-05 |
| DS-R-C25 | 1,00E-05 | 1,00E-05 | 1,00E-05 | 1,00E-05 | 1,00E-05 | 1,00E-05 | 1,00E-05 | 1,00E-05 | 1,00E-05 | 1,00E-05 | 0,33897 | 0,539402 | 1,00E-05 | 1,00E-05 | 1,00E-05 | 0,070651 | 0,050837 | 1,00E-05 |
| DS-R-C26 | 1,00E-05 | 1,00E-05 | 1,00E-05 | 0,125275 | 1,00E-05 | 1,00E-05 | 1,00E-05 | 1,00E-05 | 1,00E-05 | 1,00E-05 | 0,244729 | 0,540841 | 1,00E-05 | 1,00E-05 | 1,00E-05 | 0,089016 | 1,00E-05 | 1,00E-05 |
| DS-R-C27 | 0,098688 | 1,00E-05 | 1,00E-05 | 1,00E-05 | 1,00E-05 | 1,00E-05 | 0,012013 | 1,00E-05 | 1,00E-05 | 0,025294 | 0,182257 | 0,424191 | 0,184071 | 1,00E-05 | 1,00E-05 | 0,073377 | 1,00E-05 | 1,00E-05 |
| DS-R-C28 | 0,135661 | 0,008602 | 1,00E-05 | 0,084309 | 1,00E-05 | 1,00E-05 | 1,00E-05 | 1,00E-05 | 0,05497 | 0,158219 | 0,279715 | 0,053693 | 1,00E-05 | 1,00E-05 | 1,00E-05 | 0,065606 | 0,159134 | 1,00E-05 |
| DS-R-C29 | 0,428989 | 1,00E-05 | 1,00E-05 | 1,00E-05 | 1,00E-05 | 1,00E-05 | 1,00E-05 | 1,00E-05 | 1,00E-05 | 0,011258 | 0,559603 | 1,00E-05 | 1,00E-05 | 1,00E-05 | 1,00E-05 | 1,00E-05 | 1,00E-05 | 1,00E-05 |
| DS-R-C3 | 0,065761 | 1,00E-05 | 1,00E-05 | 1,00E-05 | 1,00E-05 | 1,00E-05 | 1,00E-05 | 1,00E-05 | 0,06955 | 1,00E-05 | 0,276405 | 0,41749 | 1,00E-05 | 1,00E-05 | 1,00E-05 | 0,170665 | 1,00E-05 | 1,00E-05 |
| DS-R-C30 | 1,00E-05 | 1,00E-05 | 1,00E-05 | 1,00E-05 | 1,00E-05 | 1,00E-05 | 1,00E-05 | 1,00E-05 | 1,00E-05 | 1,00E-05 | 1,00E-05 | 0,851172 | 1,00E-05 | 1,00E-05 | 1,00E-05 | 1,00E-05 | 0,148668 | 1,00E-05 |
| DS-R-C31 | 1,00E-05 | 1,00E-05 | 0,011535 | 1,00E-05 | 1,00E-05 | 0,988305 | 1,00E-05 | 1,00E-05 | 1,00E-05 | 1,00E-05 | 1,00E-05 | 1,00E-05 | 1,00E-05 | 1,00E-05 | 1,00E-05 | 1,00E-05 | 1,00E-05 | 1,00E-05 |
| DS-R-C32 | 0,159947 | 1,00E-05 | 1,00E-05 | 1,00E-05 | 1,00E-05 | 1,00E-05 | 0,022694 | 1,00E-05 | 1,00E-05 | 1,00E-05 | 0,176239 | 0,497685 | 1,00E-05 | 1,00E-05 | 1,00E-05 | 0,143305 | 1,00E-05 | 1,00E-05 |
| DS-R-C33 | 1,00E-05 | 1,00E-05 | 1,00E-05 | 1,00E-05 | 1,00E-05 | 0,060236 | 0,022606 | 1,00E-05 | 1,00E-05 | 1,00E-05 | 0,23435 | 0,501241 | 1,00E-05 | 1,00E-05 | 0,008059 | 0,173388 | 1,00E-05 | 1,00E-05 |
| DS-R-C34 | 0,126266 | 1,00E-05 | 0,001463 | 1,00E-05 | 1,00E-05 | 1,00E-05 | 0,01793 | 1,00E-05 | 0,010895 | 1,00E-05 | 0,298406 | 0,399893 | 1,00E-05 | 1,00E-05 | 1,00E-05 | 0,145038 | 1,00E-05 | 1,00E-05 |
| DS-R-C35 | 1,00E-05 | 1,00E-05 | 1,00E-05 | 1,00E-05 | 1,00E-05 | 1,00E-05 | 1,00E-05 | 1,00E-05 | 1,00E-05 | 1,00E-05 | 0,906416 | 1,00E-05 | 1,00E-05 | 1,00E-05 | 1,00E-05 | 1,00E-05 | 0,093424 | 1,00E-05 |
| DS-R-C36 | 1,00E-05 | 0,008834 | 1,00E-05 | 1,00E-05 | 1,00E-05 | 1,00E-05 | 0,006848 | 1,00E-05 | 0,098035 | 1,00E-05 | 0,154409 | 0,554054 | 1,00E-05 | 1,00E-05 | 1,00E-05 | 0,138592 | 0,039119 | 1,00E-05 |
| DS-R-C37 | 1,00E-05 | 1,00E-05 | 1,00E-05 | 1,00E-05 | 1,00E-05 | 0,026475 | 1,00E-05 | 1,00E-05 | 1,00E-05 | 1,00E-05 | 0,794058 | 0,179316 | 1,00E-05 | 1,00E-05 | 1,00E-05 | 1,00E-05 | 1,00E-05 | 1,00E-05 |
| DS-R-C38 | 1,00E-05 | 1,00E-05 | 0,014062 | 1,00E-05 | 1,00E-05 | 0,120123 | 1,00E-05 | 1,00E-05 | 1,00E-05 | 1,00E-05 | 1,00E-05 | 0,505394 | 0,360281 | 1,00E-05 | 1,00E-05 | 1,00E-05 | 1,00E-05 | 1,00E-05 |
| DS-R-C39 | 1,00E-05 | 1,00E-05 | 1,00E-05 | 1,00E-05 | 1,00E-05 | 1,00E-05 | 1,00E-05 | 1,00E-05 | 1,00E-05 | 1,00E-05 | 0,389745 | 0,610095 | 1,00E-05 | 1,00E-05 | 1,00E-05 | 1,00E-05 | 1,00E-05 | 1,00E-05 |
| DS-R-C4 | 0,045816 | 1,00E-05 | 1,00E-05 | 1,00E-05 | 1,00E-05 | 1,00E-05 | 0,075788 | 1,00E-05 | 1,00E-05 | 1,00E-05 | 0,339749 | 0,514354 | 1,00E-05 | 1,00E-05 | 1,00E-05 | 0,024164 | 1,00E-05 | 1,00E-05 |
| DS-R-C40 | 0,089485 | 1,00E-05 | 1,00E-05 | 1,00E-05 | 1,00E-05 | 1,00E-05 | 0,005573 | 1,00E-05 | 1,00E-05 | 1,00E-05 | 0,105246 | 0,503122 | 0,08597 | 1,00E-05 | 1,00E-05 | 0,143818 | 0,066676 | 1,00E-05 |
| DS-R-C41 | 1,00E-05 | 1,00E-05 | 1,00E-05 | 1,00E-05 | 1,00E-05 | 0,993321 | 1,00E-05 | 1,00E-05 | 1,00E-05 | 0,006519 | 1,00E-05 | 1,00E-05 | 1,00E-05 | 1,00E-05 | 1,00E-05 | 1,00E-05 | 1,00E-05 | 1,00E-05 |
| DS-R-C42 | 0,063632 | 1,00E-05 | 1,00E-05 | 1,00E-05 | 1,00E-05 | 1,00E-05 | 0,011275 | 1,00E-05 | 1,00E-05 | 0,064476 | 0,267904 | 0,592584 | 1,00E-05 | 1,00E-05 | 1,00E-05 | 1,00E-05 | 1,00E-05 | 1,00E-05 |
| DS-R-C43 | 1,00E-05 | 0,053702 | 1,00E-05 | 1,00E-05 | 1,00E-05 | 1,00E-05 | 1,00E-05 | 1,00E-05 | 1,00E-05 | 1,00E-05 | 0,239627 | 0,682429 | 0,024103 | 1,00E-05 | 1,00E-05 | 1,00E-05 | 1,00E-05 | 1,00E-05 |
| DS-R-C44 | 1,00E-05 | 1,00E-05 | 1,00E-05 | 1,00E-05 | 1,00E-05 | 0,225971 | 0,005791 | 0,007522 | 0,104118 | 1,00E-05 | 0,656468 | 1,00E-05 | 1,00E-05 | 1,00E-05 | 1,00E-05 | 1,00E-05 | 1,00E-05 | 1,00E-05 |
| DS-R-C45 | 0,186855 | 1,00E-05 | 1,00E-05 | 1,00E-05 | 1,00E-05 | 1,00E-05 | 1,00E-05 | 1,00E-05 | 0,031871 | 1,00E-05 | 0,170432 | 0,305679 | 0,243589 | 1,00E-05 | 1,00E-05 | 0,061455 | 1,00E-05 | 1,00E-05 |
| DS-R-C46 | 1,00E-05 | 1,00E-05 | 1,00E-05 | 1,00E-05 | 1,00E-05 | 0,073892 | 1,00E-05 | 1,00E-05 | 1,00E-05 | 1,00E-05 | 0,621537 | 0,136469 | 1,00E-05 | 1,00E-05 | 1,00E-05 | 0,167962 | 1,00E-05 | 1,00E-05 |
| DS-R-C5 | 0,042337 | 1,00E-05 | 1,00E-05 | 1,00E-05 | 1,00E-05 | 1,00E-05 | 1,00E-05 | 1,00E-05 | 0,036624 | 1,00E-05 | 0,186928 | 0,31126 | 0,235112 | 1,00E-05 | 1,00E-05 | 0,13524 | 0,052389 | 1,00E-05 |
| DS-R-C6 | 0,109764 | 1,00E-05 | 1,00E-05 | 1,00E-05 | 0,000408 | 1,00E-05 | 0,013779 | 1,00E-05 | 1,00E-05 | 1,00E-05 | 0,357699 | 0,51822 | 1,00E-05 | 1,00E-05 | 1,00E-05 | 1,00E-05 | 1,00E-05 | 1,00E-05 |
| DS-R-C7 | 1,00E-05 | 1,00E-05 | 1,00E-05 | 1,00E-05 | 1,00E-05 | 1,00E-05 | 0,013014 | 1,00E-05 | 1,00E-05 | 1,00E-05 | 0,138891 | 0,369651 | 0,280556 | 1,00E-05 | 1,00E-05 | 0,012691 | 0,185077 | 1,00E-05 |
| DS-R-C8 | 1,00E-05 | 1,00E-05 | 1,00E-05 | 1,00E-05 | 1,00E-05 | 0,112296 | 0,083383 | 1,00E-05 | 1,00E-05 | 0,081506 | 0,234657 | 0,411887 | 1,00E-05 | 1,00E-05 | 1,00E-05 | 0,076149 | 1,00E-05 | 1,00E-05 |
| DS-R-C9 | 0,115295 | 1,00E-05 | 1,00E-05 | 1,00E-05 | 1,00E-05 | 0,021496 | 1,00E-05 | 0,011007 | 0,136575 | 1,00E-05 | 0,117037 | 0,062252 | 0,463394 | 1,00E-05 | 1,00E-05 | 0,072844 | 1,00E-05 | 1,00E-05 |
| NH0000513 | 0,975828 | 1,00E-05 | 1,00E-05 | 1,00E-05 | 1,00E-05 | 1,00E-05 | 1,00E-05 | 1,00E-05 | 1,00E-05 | 0,021877 | 1,00E-05 | 0,002145 | 1,00E-05 | 1,00E-05 | 1,00E-05 | 1,00E-05 | 1,00E-05 | 1,00E-05 |
| NH0000580 | 1,00E-05 | 1,00E-05 | 1,00E-05 | 1,00E-05 | 1,00E-05 | 1,00E-05 | 1,00E-05 | 1,00E-05 | 1,00E-05 | 0,084467 | 1,00E-05 | 1,00E-05 | 1,00E-05 | 1,00E-05 | 1,00E-05 | 1,00E-05 | 0,915373 | 1,00E-05 |
| NH0001224 | 1,00E-05 | 1,00E-05 | 1,00E-05 | 1,00E-05 | 1,00E-05 | 0,555606 | 1,00E-05 | 1,00E-05 | 1,00E-05 | 0,035747 | 1,00E-05 | 1,00E-05 | 1,00E-05 | 1,00E-05 | 1,00E-05 | 1,00E-05 | 0,408496 | 1,00E-05 |
| NH0001391 | 1,00E-05 | 1,00E-05 | 1,00E-05 | 1,00E-05 | 0,00065 | 1,00E-05 | 1,00E-05 | 1,00E-05 | 1,00E-05 | 0,067577 | 1,00E-05 | 1,00E-05 | 1,00E-05 | 1,00E-05 | 1,00E-05 | 1,00E-05 | 0,931623 | 1,00E-05 |
| NH0001540 | 1,00E-05 | 1,00E-05 | 1,00E-05 | 1,00E-05 | 1,00E-05 | 1,00E-05 | 1,00E-05 | 1,00E-05 | 1,00E-05 | 0,057242 | 1,00E-05 | 1,00E-05 | 1,00E-05 | 1,00E-05 | 1,00E-05 | 1,00E-05 | 0,942598 | 1,00E-05 |
| NH0001541 | 1,00E-05 | 1,00E-05 | 1,00E-05 | 1,00E-05 | 1,00E-05 | 1,00E-05 | 1,00E-05 | 1,00E-05 | 1,00E-05 | 0,17861 | 1,00E-05 | 1,00E-05 | 1,00E-05 | 1,00E-05 | 1,00E-05 | 1,00E-05 | 0,82123 | 1,00E-05 |
| NH0001542 | 0,93866 | 1,00E-05 | 1,00E-05 | 1,00E-05 | 1,00E-05 | 1,00E-05 | 1,00E-05 | 1,00E-05 | 0,010044 | 0,051146 | 1,00E-05 | 1,00E-05 | 1,00E-05 | 1,00E-05 | 1,00E-05 | 1,00E-05 | 1,00E-05 | 1,00E-05 |
| NH0001543 | 0,118097 | 1,00E-05 | 1,00E-05 | 0,038084 | 1,00E-05 | 1,00E-05 | 1,00E-05 | 1,00E-05 | 0,087903 | 0,003128 | 0,208612 | 0,294271 | 0,243118 | 1,00E-05 | 1,00E-05 | 0,006687 | 1,00E-05 | 1,00E-05 |

| | | | | | | | | | | | | | | | | | | |
|---|---|---|---|---|---|---|---|---|---|---|---|---|---|---|---|---|---|---|
| NH0001544 | 1,00E-05 | 1,00E-05 | 1,00E-05 | 1,00E-05 | 1,00E-05 | 1,00E-05 | 1,00E-05 | 1,00E-05 | 1,00E-05 | 1,00E-05 | 0,345519 | 0,395204 | 0,139431 | 1,00E-05 | 1,00E-05 | 0,072949 | 0,046766 | 1,00E-05 |
| NH0001545 | 1,00E-05 | 1,00E-05 | 0,000364 | 1,00E-05 | 1,00E-05 | 1,00E-05 | 0,00753 | 1,00E-05 | 1,00E-05 | 1,00E-05 | 0,299213 | 0,412099 | 0,02433 | 1,00E-05 | 1,00E-05 | 0,179038 | 0,077317 | 1,00E-05 |
| NH0001669 | 0,055651 | 1,00E-05 | 1,00E-05 | 1,00E-05 | 1,00E-05 | 1,00E-05 | 0,014651 | 1,00E-05 | 0,108715 | 1,00E-05 | 0,108422 | 0,416955 | 0,206626 | 1,00E-05 | 1,00E-05 | 0,08887 | 1,00E-05 | 1,00E-05 |
| NH0001670 | 1,00E-05 | 1,00E-05 | 1,00E-05 | 1,00E-05 | 1,00E-05 | 1,00E-05 | 1,00E-05 | 1,00E-05 | 1,00E-05 | 1,00E-05 | 0,954302 | 1,00E-05 | 1,00E-05 | 1,00E-05 | 1,00E-05 | 0,045538 | 1,00E-05 | 1,00E-05 |
| NH0001671 | 0,117823 | 0,00426 | 1,00E-05 | 1,00E-05 | 1,00E-05 | 0,157732 | 0,028458 | 1,00E-05 | 0,075407 | 1,00E-05 | 0,414356 | 0,000179 | 0,164456 | 1,00E-05 | 1,00E-05 | 0,037238 | 1,00E-05 | 1,00E-05 |
| NH0001672 | 1,00E-05 | 1,10E-05 | 1,00E-05 | 1,00E-05 | 1,00E-05 | 1,00E-05 | 1,00E-05 | 1,00E-05 | 0,133188 | 1,00E-05 | 1,00E-05 | 0,121032 | 0,705656 | 1,00E-05 | 1,00E-05 | 1,00E-05 | 0,039983 | 1,00E-05 |
| NH0001673 | 0,084978 | 0,010089 | 1,00E-05 | 1,00E-05 | 1,00E-05 | 1,00E-05 | 1,00E-05 | 1,00E-05 | 1,00E-05 | 1,00E-05 | 0,271861 | 0,415412 | 1,00E-05 | 1,00E-05 | 1,00E-05 | 0,123834 | 0,093706 | 1,00E-05 |
| NH0001674 | 0,153646 | 1,00E-05 | 1,00E-05 | 0,102241 | 1,00E-05 | 1,00E-05 | 1,00E-05 | 1,00E-05 | 0,0761 | 0,013417 | 0,159349 | 0,080163 | 0,355181 | 1,00E-05 | 1,00E-05 | 0,059803 | 1,00E-05 | 1,00E-05 |
| NH0001675 | 0,941246 | 1,00E-05 | 1,00E-05 | 1,00E-05 | 1,00E-05 | 1,00E-05 | 1,00E-05 | 1,00E-05 | 1,00E-05 | 0,058594 | 1,00E-05 | 1,00E-05 | 1,00E-05 | 1,00E-05 | 1,00E-05 | 1,00E-05 | 1,00E-05 | 1,00E-05 |
| NH0001676 | 0,956731 | 1,00E-05 | 1,00E-05 | 1,00E-05 | 1,00E-05 | 1,00E-05 | 1,00E-05 | 1,00E-05 | 0,043109 | 1,00E-05 | 1,00E-05 | 1,00E-05 | 1,00E-05 | 1,00E-05 | 1,00E-05 | 1,00E-05 | 1,00E-05 | 1,00E-05 |
| NH0001679 | 1,00E-05 | 1,00E-05 | 1,00E-05 | 1,00E-05 | 1,00E-05 | 0,604788 | 1,00E-05 | 1,00E-05 | 1,00E-05 | 0,040446 | 1,00E-05 | 1,00E-05 | 1,00E-05 | 1,00E-05 | 1,00E-05 | 1,00E-05 | 0,354616 | 1,00E-05 |
| NH0001680 | 1,00E-05 | 1,00E-05 | 1,00E-05 | 1,00E-05 | 1,00E-05 | 0,679907 | 1,00E-05 | 1,00E-05 | 1,00E-05 | 0,004325 | 1,00E-05 | 1,00E-05 | 1,00E-05 | 1,00E-05 | 1,00E-05 | 1,00E-05 | 0,315619 | 1,00E-05 |
| NH0001681 | 1,00E-05 | 1,00E-05 | 1,00E-05 | 1,00E-05 | 1,00E-05 | 0,617783 | 1,00E-05 | 1,00E-05 | 1,00E-05 | 0,016565 | 1,00E-05 | 1,00E-05 | 1,00E-05 | 1,00E-05 | 1,00E-05 | 1,00E-05 | 0,365502 | 1,00E-05 |
| NH0001682 | 1,00E-05 | 1,00E-05 | 1,00E-05 | 1,00E-05 | 1,00E-05 | 0,558818 | 0,009404 | 1,00E-05 | 1,00E-05 | 0,018399 | 1,00E-05 | 1,00E-05 | 1,00E-05 | 1,00E-05 | 1,00E-05 | 1,00E-05 | 0,413238 | 1,00E-05 |
| NH0001719 | 1,00E-05 | 1,00E-05 | 1,00E-05 | 1,00E-05 | 1,00E-05 | 1,00E-05 | 1,00E-05 | 1,00E-05 | 1,00E-05 | 0,060266 | 1,00E-05 | 1,00E-05 | 1,00E-05 | 1,00E-05 | 1,00E-05 | 1,00E-05 | 0,939574 | 1,00E-05 |
| NH0001798 | 1,00E-05 | 1,00E-05 | 1,00E-05 | 1,00E-05 | 1,00E-05 | 1,00E-05 | 1,00E-05 | 1,00E-05 | 0,024559 | 0,079939 | 1,00E-05 | 1,00E-05 | 1,00E-05 | 1,00E-05 | 1,00E-05 | 1,00E-05 | 0,895352 | 1,00E-05 |
| NH0001799_set6 | 1,00E-05 | 1,00E-05 | 1,00E-05 | 1,00E-05 | 1,00E-05 | 1,00E-05 | 1,00E-05 | 1,00E-05 | 1,00E-05 | 0,061958 | 1,00E-05 | 1,00E-05 | 1,00E-05 | 1,00E-05 | 1,00E-05 | 1,00E-05 | 0,937882 | 1,00E-05 |
| NH0001800_set6 | 1,00E-05 | 1,00E-05 | 1,00E-05 | 1,00E-05 | 1,00E-05 | 1,00E-05 | 1,00E-05 | 1,00E-05 | 1,00E-05 | 0,041043 | 1,00E-05 | 1,00E-05 | 1,00E-05 | 1,00E-05 | 1,00E-05 | 1,00E-05 | 0,958797 | 1,00E-05 |
| NH0001801 | 1,00E-05 | 1,00E-05 | 1,00E-05 | 1,00E-05 | 1,00E-05 | 0,540731 | 1,00E-05 | 1,00E-05 | 1,00E-05 | 1,00E-05 | 1,00E-05 | 1,00E-05 | 1,00E-05 | 1,00E-05 | 1,00E-05 | 1,00E-05 | 0,459109 | 1,00E-05 |
| NH0001802 | 1,00E-05 | 1,00E-05 | 1,00E-05 | 1,00E-05 | 1,00E-05 | 1,00E-05 | 1,00E-05 | 1,00E-05 | 1,00E-05 | 0,04751 | 1,00E-05 | 0,002988 | 1,00E-05 | 1,00E-05 | 1,00E-05 | 1,00E-05 | 0,949351 | 1,00E-05 |
| NH0001803 | 0,522254 | 0,359393 | 1,00E-05 | 1,00E-05 | 1,00E-05 | 1,00E-05 | 0,004514 | 1,00E-05 | 1,00E-05 | 0,05493 | 1,00E-05 | 1,00E-05 | 1,00E-05 | 1,00E-05 | 1,00E-05 | 1,00E-05 | 0,058779 | 1,00E-05 |
| NH0001804 | 1,00E-05 | 1,00E-05 | 1,00E-05 | 1,00E-05 | 1,00E-05 | 0,514756 | 1,00E-05 | 1,00E-05 | 1,00E-05 | 0,096192 | 1,00E-05 | 1,00E-05 | 1,00E-05 | 1,00E-05 | 1,00E-05 | 1,00E-05 | 0,388901 | 1,00E-05 |
| NH0001805 | 0,94854 | 1,00E-05 | 1,00E-05 | 1,00E-05 | 1,00E-05 | 1,00E-05 | 0,003469 | 1,00E-05 | 1,00E-05 | 0,047841 | 1,00E-05 | 1,00E-05 | 1,00E-05 | 1,00E-05 | 1,00E-05 | 1,00E-05 | 1,00E-05 | 1,00E-05 |
| NH0001806 | 0,981067 | 1,00E-05 | 1,00E-05 | 1,00E-05 | 1,00E-05 | 1,00E-05 | 1,00E-05 | 1,00E-05 | 1,00E-05 | 0,018773 | 1,00E-05 | 1,00E-05 | 1,00E-05 | 1,00E-05 | 1,00E-05 | 1,00E-05 | 1,00E-05 | 1,00E-05 |
| NH0001807 | 1,00E-05 | 1,00E-05 | 1,00E-05 | 1,00E-05 | 1,00E-05 | 1,00E-05 | 1,00E-05 | 1,00E-05 | 1,00E-05 | 0,031576 | 1,00E-05 | 1,00E-05 | 1,00E-05 | 1,00E-05 | 1,00E-05 | 1,00E-05 | 0,968264 | 1,00E-05 |
| NH0001808 | 0,380511 | 0,027019 | 1,00E-05 | 0,035569 | 1,00E-05 | 1,00E-05 | 1,00E-05 | 1,00E-05 | 0,004396 | 1,00E-05 | 0,125455 | 0,108227 | 0,194676 | 1,00E-05 | 1,00E-05 | 0,035196 | 0,088861 | 1,00E-05 |
| NH0001867 | 1,00E-05 | 1,00E-05 | 1,00E-05 | 1,00E-05 | 1,00E-05 | 0,636797 | 1,00E-05 | 1,00E-05 | 1,00E-05 | 0,029269 | 1,00E-05 | 0,022338 | 1,00E-05 | 1,00E-05 | 1,00E-05 | 1,00E-05 | 0,311455 | 1,00E-05 |
| NH0001868 | 1,00E-05 | 0,059727 | 1,00E-05 | 1,00E-05 | 1,00E-05 | 0,664988 | 1,00E-05 | 1,00E-05 | 1,00E-05 | 0,075444 | 1,00E-05 | 1,00E-05 | 1,00E-05 | 1,00E-05 | 1,00E-05 | 1,00E-05 | 0,199702 | 1,00E-05 |
| NH0001869 | 0,954089 | 1,00E-05 | 1,00E-05 | 1,00E-05 | 1,00E-05 | 1,00E-05 | 0,011522 | 1,00E-05 | 1,00E-05 | 0,034238 | 1,00E-05 | 1,00E-05 | 1,00E-05 | 1,00E-05 | 1,00E-05 | 1,00E-05 | 1,00E-05 | 1,00E-05 |
| NH0001870 | 0,862389 | 0,073928 | 1,00E-05 | 1,00E-05 | 1,00E-05 | 1,00E-05 | 1,00E-05 | 1,00E-05 | 1,00E-05 | 0,063533 | 1,00E-05 | 1,00E-05 | 1,00E-05 | 1,00E-05 | 1,00E-05 | 1,00E-05 | 1,00E-05 | 1,00E-05 |
| NH0001871 | 1,00E-05 | 1,30E-05 | 1,00E-05 | 1,00E-05 | 1,00E-05 | 0,650926 | 1,00E-05 | 1,00E-05 | 1,00E-05 | 0,01721 | 1,00E-05 | 1,00E-05 | 1,00E-05 | 1,00E-05 | 1,00E-05 | 1,00E-05 | 0,33171 | 1,00E-05 |
| NH0001872 | 1,00E-05 | 1,00E-05 | 1,00E-05 | 1,00E-05 | 1,00E-05 | 1,00E-05 | 1,00E-05 | 1,00E-05 | 1,00E-05 | 0,151859 | 1,00E-05 | 1,00E-05 | 1,00E-05 | 1,00E-05 | 1,00E-05 | 1,00E-05 | 0,847981 | 1,00E-05 |
| NH0001873 | 0,952163 | 1,00E-05 | 1,00E-05 | 1,00E-05 | 1,00E-05 | 1,00E-05 | 1,00E-05 | 1,00E-05 | 1,00E-05 | 0,047677 | 1,00E-05 | 1,00E-05 | 1,00E-05 | 1,00E-05 | 1,00E-05 | 1,00E-05 | 1,00E-05 | 1,00E-05 |
| NH0001874 | 0,168302 | 1,00E-05 | 1,00E-05 | 1,00E-05 | 1,00E-05 | 0,569061 | 1,00E-05 | 1,00E-05 | 1,00E-05 | 0,004883 | 1,00E-05 | 1,00E-05 | 1,00E-05 | 1,00E-05 | 1,00E-05 | 1,00E-05 | 0,257613 | 1,00E-05 |
| NH0001875 | 0,735649 | 1,00E-05 | 1,00E-05 | 1,00E-05 | 1,00E-05 | 0,201892 | 1,00E-05 | 1,00E-05 | 1,00E-05 | 1,00E-05 | 0,062309 | 1,00E-05 | 1,00E-05 | 1,00E-05 | 1,00E-05 | 1,00E-05 | 1,00E-05 | 1,00E-05 |
| NH0001876 | 0,092113 | 0,148215 | 1,00E-05 | 1,00E-05 | 1,00E-05 | 0,514547 | 1,00E-05 | 1,00E-05 | 1,00E-05 | 0,000159 | 1,00E-05 | 1,00E-05 | 1,00E-05 | 1,00E-05 | 1,00E-05 | 1,00E-05 | 0,244835 | 1,00E-05 |
| NH0001877 | 1,00E-05 | 1,00E-05 | 1,00E-05 | 1,00E-05 | 1,00E-05 | 1,00E-05 | 1,00E-05 | 1,00E-05 | 1,00E-05 | 0,022438 | 1,00E-05 | 1,00E-05 | 1,00E-05 | 1,00E-05 | 1,00E-05 | 1,00E-05 | 0,977402 | 1,00E-05 |
| NH0001878 | 0,983942 | 1,00E-05 | 1,00E-05 | 1,00E-05 | 1,00E-05 | 1,00E-05 | 0,004617 | 1,00E-05 | 1,00E-05 | 0,011292 | 1,00E-05 | 1,00E-05 | 1,00E-05 | 1,00E-05 | 1,00E-05 | 1,00E-05 | 1,00E-05 | 1,00E-05 |
| NH0001879 | 0,880082 | 0,076453 | 1,00E-05 | 1,00E-05 | 1,00E-05 | 1,00E-05 | 1,00E-05 | 1,00E-05 | 1,00E-05 | 0,043315 | 1,00E-05 | 1,00E-05 | 1,00E-05 | 1,00E-05 | 1,00E-05 | 1,00E-05 | 1,00E-05 | 1,00E-05 |
| NH0001880 | 0,691429 | 0,119243 | 1,00E-05 | 1,00E-05 | 1,00E-05 | 0,178346 | 1,00E-05 | 1,00E-05 | 1,00E-05 | 0,010841 | 1,00E-05 | 1,00E-05 | 1,00E-05 | 1,00E-05 | 1,00E-05 | 1,00E-05 | 1,00E-05 | 1,00E-05 |
| NH0001881 | 1,00E-05 | 1,00E-05 | 1,00E-05 | 1,00E-05 | 1,00E-05 | 1,00E-05 | 1,00E-05 | 1,00E-05 | 1,00E-05 | 0,07367 | 1,00E-05 | 1,00E-05 | 1,00E-05 | 1,00E-05 | 1,00E-05 | 1,00E-05 | 0,92617 | 1,00E-05 |
| NH0001882 | 0,941001 | 1,00E-05 | 1,00E-05 | 1,00E-05 | 1,00E-05 | 1,00E-05 | 1,00E-05 | 1,00E-05 | 1,00E-05 | 0,058839 | 1,00E-05 | 1,00E-05 | 1,00E-05 | 1,00E-05 | 1,00E-05 | 1,00E-05 | 1,00E-05 | 1,00E-05 |
| NH0001883 | 0,332382 | 1,00E-05 | 1,00E-05 | 1,00E-05 | 1,00E-05 | 0,649583 | 1,00E-05 | 1,00E-05 | 1,00E-05 | 0,017885 | 1,00E-05 | 1,00E-05 | 1,00E-05 | 1,00E-05 | 1,00E-05 | 1,00E-05 | 1,00E-05 | 1,00E-05 |

| | | | | | | | | | | | | | | | | | | |
|---|---|---|---|---|---|---|---|---|---|---|---|---|---|---|---|---|---|---|
| NH0001884 | 1,00E-05 | 0,147068 | 1,00E-05 | 1,00E-05 | 1,00E-05 | 0,683886 | 0,168896 | 1,00E-05 | 1,00E-05 | 1,00E-05 | 1,00E-05 | 1,00E-05 | 1,00E-05 | 1,00E-05 | 1,00E-05 | 1,00E-05 | 1,00E-05 | 1,00E-05 |
| NH0001885 | 1,00E-05 | 1,10E-05 | 1,00E-05 | 1,00E-05 | 1,00E-05 | 1,00E-05 | 1,00E-05 | 1,00E-05 | 1,00E-05 | 0,042338 | 1,00E-05 | 1,00E-05 | 1,00E-05 | 1,00E-05 | 1,00E-05 | 1,00E-05 | 0,957501 | 1,00E-05 |
| NH0001886 | 0,165422 | 0,473265 | 1,00E-05 | 1,00E-05 | 1,00E-05 | 0,142608 | 1,00E-05 | 1,00E-05 | 1,00E-05 | 0,014124 | 1,00E-05 | 1,00E-05 | 1,00E-05 | 1,10E-05 | 1,00E-05 | 1,00E-05 | 0,20445 | 1,00E-05 |
| NH0001887 | 1,00E-05 | 1,00E-05 | 1,00E-05 | 1,00E-05 | 1,00E-05 | 1,00E-05 | 1,00E-05 | 1,00E-05 | 1,00E-05 | 0,242095 | 1,00E-05 | 1,00E-05 | 1,00E-05 | 1,00E-05 | 1,00E-05 | 1,00E-05 | 0,757745 | 1,00E-05 |
| NH0001888 | 0,114011 | 1,00E-05 | 1,00E-05 | 1,00E-05 | 1,00E-05 | 1,00E-05 | 1,00E-05 | 1,00E-05 | 1,00E-05 | 1,00E-05 | 0,602118 | 0,200832 | 1,00E-05 | 0,002572 | 1,00E-05 | 0,080337 | 1,00E-05 | 1,00E-05 |
| NH0001889 | 0,222837 | 1,00E-05 | 1,00E-05 | 1,00E-05 | 1,00E-05 | 1,00E-05 | 1,00E-05 | 1,00E-05 | 0,050716 | 1,00E-05 | 0,40036 | 0,198194 | 1,00E-05 | 1,00E-05 | 1,00E-05 | 0,039739 | 0,088034 | 1,00E-05 |
| NH0002026 | 1,00E-05 | 1,00E-05 | 1,00E-05 | 1,00E-05 | 1,00E-05 | 1,00E-05 | 1,00E-05 | 1,00E-05 | 1,00E-05 | 0,046766 | 1,00E-05 | 0,043131 | 1,00E-05 | 1,00E-05 | 1,00E-05 | 0,079959 | 0,830004 | 1,00E-05 |
| NH0002027 | 1,00E-05 | 1,00E-05 | 1,00E-05 | 1,00E-05 | 1,00E-05 | 1,00E-05 | 1,00E-05 | 1,00E-05 | 1,00E-05 | 0,048223 | 0,00451 | 1,30E-05 | 1,00E-05 | 1,00E-05 | 1,00E-05 | 1,00E-05 | 0,947115 | 1,00E-05 |

Table S7. The up and down-regulated GSEA results for the HALLMARK geneset analysis

| The output from GSEA: | |
|---|---|
| NAME | Gene set name |
| SIZE | Number of genes in the gene set after filtering out those genes not in the expression dataset |
| ES | Enrichment score for the gene set; that is, the degree to which this gene set is overrepresented at the top or bottom of the ranked list of genes in the expression dataset. |
| NES | Normalized enrichment score; that is, the enrichment score for the gene set after it has been normalized across analyzed gene sets. |
| NOM p-val | Nominal p value; that is, the statistical significance of the enrichment score. The nominal p value is not adjusted for gene set size or multiple hypothesis testing; therefore, it is of limited use in comparing gene sets. |
| FDR q-val | False discovery rate; that is, the estimated probability that the normalized enrichment score represents a false positive finding. |
| FWER p-val | Familywise-error rate; that is, a more conservatively estimated probability that the normalized enrichment score represents a false positive finding. |
| RANK AT MAX | The position in the ranked list at which the maximum enrichment score occurred. |
| LEADING EDGE | Displays the three statistics used to define the leading edge subset: |
| | Tags - The percentage of gene hits before (for positive ES) or after (for negative ES) the peak in the running enrichment score. |
| | List - The percentage of genes in the ranked gene list before (for positive ES) or after (for negative ES) the peak in the running enrichment score. |
| | Signal- The enrichment signal strength that combines the two previous statisticspeak in the running enrichment score. |

| **NAME** | **SIZE** | **ES** | **NES** | **NOM p-val** | **FDR q-val** | **FWER p-val** | **RANK AT MAX** | **LEADING EDGE** |
|---|---|---|---|---|---|---|---|---|
| HALLMARK_EPITHELIAL_MESENCHYMAL_TRANSITION | 196 | 0,5657118 | 2,2042103 | 0 | 0 | 0 | 3768 | tags=57%, list=20%, signal=70% |
| HALLMARK_KRAS_SIGNALING_UP | 186 | 0,52319187 | 1,9985508 | 0 | 0 | 0 | 3670 | tags=40%, list=20%, signal=49% |

| | | | | | | | | |
|---|---|---|---|---|---|---|---|---|
| HALLMARK_INFLAMMATORY_RESPONSE | 189 | 0,49264094 | 1,9070023 | 0 | 9,71E-04 | 0,002 | 4052 | tags=44%, list=22%, signal=56% |
| HALLMARK_ALLOGRAFT_REJECTION | 186 | 0,48069036 | 1,8378925 | 0 | 0,0020914811 | 0,006 | 3887 | tags=36%, list=21%, signal=45% |
| HALLMARK_IL6_JAK_STAT3_SIGNALING | 83 | 0,49087122 | 1,7134684 | 0 | 0,007683711 | 0,027 | 3026 | tags=31%, list=16%, signal=37% |
| HALLMARK_FATTY_ACID_METABOLISM | 146 | 0,4468113 | 1,677772 | 0 | 0,011050713 | 0,048 | 5028 | tags=42%, list=27%, signal=57% |
| HALLMARK_COAGULATION | 111 | 0,4547866 | 1,636534 | 0,0014836795 | 0,013567286 | 0,069 | 4377 | tags=44%, list=23%, signal=57% |
| HALLMARK_G2M_CHECKPOINT | 197 | 0,41649923 | 1,6051984 | 0 | 0,015658991 | 0,09 | 6037 | tags=43%, list=32%, signal=62% |
| HALLMARK_MYOGENESIS | 164 | 0,4166427 | 1,5802467 | 0,0027210885 | 0,020961603 | 0,132 | 2968 | tags=30%, list=16%, signal=36% |
| HALLMARK_APICAL_JUNCTION | 180 | 0,40678105 | 1,555372 | 0,0013227513 | 0,023637386 | 0,162 | 3960 | tags=36%, list=21%, signal=45% |
| HALLMARK_ESTROGEN_RESPONSE_LATE | 187 | 0,40013033 | 1,5390147 | 0,0026350461 | 0,024929244 | 0,188 | 2139 | tags=23%, list=11%, signal=26% |
| HALLMARK_ADIPOGENESIS | 189 | 0,38413745 | 1,479664 | 0,0027662518 | 0,043514997 | 0,32 | 5636 | tags=42%, list=30%, signal=59% |
| HALLMARK_APICAL_SURFACE | 39 | 0,48144186 | 1,4448133 | 0,043902438 | 0,059792474 | 0,427 | 4104 | tags=49%, list=22%, signal=62% |
| HALLMARK_IL2_STAT5_SIGNALING | 194 | 0,37415728 | 1,4410989 | 0,012 | 0,057225436 | 0,441 | 4588 | tags=36%, list=25%, signal=47% |
| HALLMARK_E2F_TARGETS | 200 | 0,37300754 | 1,4387197 | 0,008097166 | 0,054292697 | 0,446 | 7107 | tags=55%, list=38%, signal=87% |
| HALLMARK_COMPLEMENT | 187 | 0,37258694 | 1,423348 | 0,01055409 | 0,060118183 | 0,499 | 4753 | tags=39%, list=25%, signal=51% |
| HALLMARK_XENOBIOTIC_METABOLISM | 175 | 0,37179902 | 1,4159665 | 0,010723861 | 0,061437514 | 0,53 | 3978 | tags=31%, list=21%, signal=39% |
| HALLMARK_ANGIOGENESIS | 32 | 0,46729648 | 1,3944752 | 0,07020548 | 0,07105753 | 0,603 | 3563 | tags=47%, list=19%, signal=58% |
| HALLMARK_OXIDATIVE_PHOSPHORYLATION | 199 | 0,3637588 | 1,3901007 | 0,009020618 | 0,069485374 | 0,612 | 6968 | tags=53%, list=37%, signal=84% |
| HALLMARK_MTORC1_SIGNALING | 200 | 0,35507572 | 1,3778205 | 0,012987013 | 0,07370265 | 0,656 | 6159 | tags=47%, list=33%, signal=69% |
| HALLMARK_KRAS_SIGNALING_DN | 128 | 0,37496015 | 1,3739074 | 0,027417028 | 0,07259083 | 0,671 | 2847 | tags=31%, list=15%, signal=37% |

| | | | | | | | | |
|---|---|---|---|---|---|---|---|---|
| HALLMARK_SPERMATOGENESIS | 88 | 0,39072073 | 1,3514171 | 0,046715327 | 0,08658947 | 0,754 | 3542 | tags=35%, list=19%, signal=43% |
| HALLMARK_APOPTOSIS | 158 | 0,35755834 | 1,3312511 | 0,03504043 | 0,099347614 | 0,807 | 5248 | tags=41%, list=28%, signal=57% |
| HALLMARK_BILE_ACID_METABOLISM | 95 | 0,3775138 | 1,3277541 | 0,05671642 | 0,098765016 | 0,824 | 3627 | tags=32%, list=19%, signal=39% |
| HALLMARK_ANDROGEN_RESPONSE | 99 | 0,36703584 | 1,2920651 | 0,06321839 | 0,13021466 | 0,905 | 5993 | tags=45%, list=32%, signal=67% |
| HALLMARK_MITOTIC_SPINDLE | 199 | 0,32331094 | 1,2461026 | 0,07853403 | 0,17949541 | 0,958 | 3841 | tags=24%, list=21%, signal=30% |
| HALLMARK_ESTROGEN_RESPONSE_EARLY | 194 | 0,31714898 | 1,2219646 | 0,09746329 | 0,21160994 | 0,984 | 1811 | tags=18%, list=10%, signal=19% |
| HALLMARK_HEDGEHOG_SIGNALING | 33 | 0,42404172 | 1,2207664 | 0,18600954 | 0,20615052 | 0,984 | 1590 | tags=21%, list=8%, signal=23% |
| HALLMARK_REACTIVE_OXYGEN_SPECIES_PATHWAY | 48 | 0,38178673 | 1,2042456 | 0,18320611 | 0,22439261 | 0,988 | 6114 | tags=48%, list=33%, signal=71% |
| HALLMARK_PEROXISOME | 96 | 0,33815864 | 1,1996043 | 0,16288953 | 0,22431763 | 0,989 | 6198 | tags=48%, list=33%, signal=71% |
| HALLMARK_UV_RESPONSE_DN | 144 | 0,30986002 | 1,1601787 | 0,18156424 | 0,28535232 | 0,999 | 5394 | tags=38%, list=29%, signal=53% |
| HALLMARK_PANCREAS_BETA_CELLS | 23 | 0,43644524 | 1,1601703 | 0,26504064 | 0,27643505 | 0,999 | 2239 | tags=30%, list=12%, signal=35% |
| HALLMARK_GLYCOLYSIS | 189 | 0,30080524 | 1,154568 | 0,16 | 0,27830198 | 1 | 5915 | tags=42%, list=32%, signal=61% |
| HALLMARK_INTERFERON_GAMMA_RESPONSE | 196 | 0,2879839 | 1,1174133 | 0,20933333 | 0,34389973 | 1 | 5025 | tags=33%, list=27%, signal=44% |
| HALLMARK_PI3K_AKT_MTOR_SIGNALING | 99 | 0,28586403 | 1,0229418 | 0,41202345 | 0,5698468 | 1 | 5533 | tags=35%, list=30%, signal=50% |
| HALLMARK_TNFA_SIGNALING_VIA_NFKB | 198 | 0,2652445 | 1,0224531 | 0,4152203 | 0,555234 | 1 | 4891 | tags=34%, list=26%, signal=45% |
| HALLMARK_HYPOXIA | 187 | 0,26416913 | 1,006997 | 0,4419525 | 0,58020717 | 1 | 3536 | tags=24%, list=19%, signal=29% |
| HALLMARK_NOTCH_SIGNALING | 32 | 0,34510008 | 1,0039699 | 0,45333335 | 0,5733785 | 1 | 4722 | tags=28%, list=25%, signal=38% |
| HALLMARK_UV_RESPONSE_UP | 151 | 0,25813383 | 0,9740819 | 0,5301543 | 0,6336612 | 1 | 4652 | tags=27%, list=25%, signal=36% |
| HALLMARK_CHOLESTEROL_HOMEOSTASIS | 73 | 0,2782744 | 0,94666255 | 0,5538462 | 0,6919574 | 1 | 3886 | tags=30%, list=21%, signal=38% |

| | | | | | | | | |
|---|---|---|---|---|---|---|---|---|
| HALLMARK_HEME_METABOLISM | 180 | 0,24732459 | 0,9426974 | 0,58616185 | 0,68591934 | 1 | 4777 | tags=27%, list=26%, signal=36% |
| HALLMARK_MYC_TARGETS_V1 | 200 | 0,21425353 | 0,8339013 | 0,85286105 | 0,93404424 | 1 | 8364 | tags=52%, list=45%, signal=92% |
| HALLMARK_DNA_REPAIR | 148 | 0,21909395 | 0,8178948 | 0,85983825 | 0,9451982 | 1 | 7301 | tags=42%, list=39%, signal=68% |
| HALLMARK_P53_PATHWAY | 197 | 0,19590086 | 0,7551609 | 0,9683377 | 1 | 1 | 5567 | tags=33%, list=30%, signal=46% |
| HALLMARK_TGF_BETA_SIGNALING | 52 | 0,18666379 | 0,59722227 | 0,9906687 | 1 | 1 | 6930 | tags=42%, list=37%, signal=67% |
| HALLMARK_MYC_TARGETS_V2 | 58 | 0,17514795 | 0,56836146 | 0,9921507 | 1 | 1 | 7686 | tags=45%, list=41%, signal=76% |
| HALLMARK_UNFOLDED_PROTEIN_RESPONSE | 112 | 0,15662047 | 0,55339175 | 1 | 1 | 1 | 7157 | tags=39%, list=38%, signal=63% |
| HALLMARK_PROTEIN_SECRETION | 95 | 0,14244103 | 0,5014526 | 1 | 0,9996381 | 1 | 5218 | tags=23%, list=28%, signal=32% |
| HALLMARK_WNT_BETA_CATENIN_SIGNALING | 39 | -0,38446742 | -1,2964368 | 0,098870054 | 0,10283078 | 0,522 | 4125 | tags=36%, list=22%, signal=46% |
| HALLMARK_INTERFERON_ALPHA_RESPONSE | 97 | -0,26346487 | -1,0494032 | 0,30375427 | 0,32908717 | 0,99 | 2140 | tags=15%, list=11%, signal=17% |

Table S8. Microbiome relative abundance at the genus-level

| Genera | Tumor | Non-tumor | Tumor Right | Tumor Left | Non Tumor Right | Non Tumor Left |
|---|---|---|---|---|---|---|
| *Streptococcus* | 0.100 | 0.092 | 0.149 | 0.041 | 0.124 | 0.052 |
| *Prevotella* | 0.057 | 0.070 | 0.024 | 0.098 | 0.068 | 0.078 |
| *Bacteroides* | 0.049 | 0.063 | 0.048 | 0.051 | 0.059 | 0.068 |
| *Escherichia-Shigella* | 0.047 | 0.076 | 0.045 | 0.051 | 0.088 | 0.060 |
| *Pseudomonas* | 0.038 | 0.029 | 0.058 | 0.014 | 0.033 | 0.024 |
| *Fusobacterium* | 0.035 | 0.004 | 0.062 | 0.003 | 0.003 | 0.006 |
| *Lactobacillus* | 0.034 | 0.009 | 0.019 | 0.051 | 0.013 | 0.005 |
| *Leptotrichia* | 0.031 | 0.006 | 0.044 | 0.016 | 0.000 | 0.012 |
| *Haemophilus* | 0.029 | 0.003 | 0.027 | 0.033 | 0.004 | 0.003 |
| *Shuttleworthia* | 0.029 | 0.014 | 0.034 | 0.023 | 0.021 | 0.006 |
| *Enterococcus* | 0.027 | 0.042 | 0.044 | 0.006 | 0.075 | 0.002 |
| *Faecalibacterium* | 0.026 | 0.029 | 0.010 | 0.047 | 0.035 | 0.023 |
| *Selenomonas* | 0.023 | 0.001 | 0.042 | 0.001 | 0.001 | 0.002 |
| *Gemella* | 0.023 | 0.007 | 0.013 | 0.035 | 0.006 | 0.010 |
| *Dialister* | 0.019 | 0.022 | 0.014 | 0.027 | 0.028 | 0.015 |
| *Bifidobacterium* | 0.017 | 0.052 | 0.005 | 0.031 | 0.016 | 0.096 |
| *Parabacteroides* | 0.015 | 0.007 | 0.002 | 0.032 | 0.008 | 0.005 |
| *Treponema* | 0.015 | 0.002 | 0.019 | 0.010 | 0.000 | 0.005 |

| | | | | | | |
|---|---|---|---|---|---|---|
| *Gardnerella* | 0.015 | 0.009 | 0.012 | 0.018 | 0.008 | 0.010 |
| *Klebsiella* | 0.015 | 0.019 | 0.024 | 0.003 | 0.031 | 0.005 |
| *Campylobacter* | 0.015 | 0.003 | 0.012 | 0.018 | 0.004 | 0.002 |
| *Akkermansia* | 0.014 | 0.007 | 0.001 | 0.030 | 0.007 | 0.006 |
| *Roseburia* | 0.012 | 0.003 | 0.020 | 0.002 | 0.002 | 0.004 |
| *Staphylococcus* | 0.011 | 0.004 | 0.018 | 0.003 | 0.003 | 0.005 |
| *Ruminococcus* | 0.010 | 0.010 | 0.005 | 0.017 | 0.016 | 0.002 |
| *Megasphaera* | 0.010 | 0.008 | 0.001 | 0.020 | 0.006 | 0.011 |
| *Holdemanella* | 0.008 | 0.008 | 0.004 | 0.012 | 0.012 | 0.003 |
| *Lachnoclostridium* | 0.007 | 0.007 | 0.003 | 0.012 | 0.008 | 0.006 |
| *Porphyromonas* | 0.006 | 0.010 | 0.004 | 0.009 | 0.012 | 0.008 |
| *Blautia* | 0.006 | 0.015 | 0.001 | 0.011 | 0.005 | 0.026 |
| *Granulicatella* | 0.005 | 0.006 | 0.003 | 0.008 | 0.004 | 0.009 |
| *Agathobacter* | 0.005 | 0.010 | 0.002 | 0.008 | 0.009 | 0.011 |
| *Subdoligranulum* | 0.004 | 0.011 | 0.002 | 0.007 | 0.009 | 0.013 |
| *Helicobacter* | 0.003 | 0.013 | 0.004 | 0.001 | 0.023 | 0.001 |
| *Anaeroplasma* | 0.002 | 0.015 | 0.004 | 0.001 | 0.000 | 0.034 |
| *Limosilactobacillus* | 0.001 | 0.001 | 0.001 | 0.001 | 0.000 | 0.003 |
| *Comamonas* | 0.001 | 0.007 | 0.002 | 0.000 | 0.003 | 0.012 |
| *Bacillus* | 0.001 | 0.003 | 0.000 | 0.002 | 0.002 | 0.005 |
| *Serratia* | 0.000 | 0.007 | 0.000 | 0.000 | 0.009 | 0.005 |
| *Salmonella* | 0.000 | 0.000 | 0.000 | 0.001 | 0.001 | 0.000 |
| *Oceanobacillus* | 0.000 | 0.001 | 0.000 | 0.000 | 0.000 | 0.001 |
| Others | 0.232 | 0.293 | 0.220 | 0.247 | 0.246 | 0.345 |

Table S9. Tumor species enrichment by ancestry

| Species | OR_AFR | AdjP_AFR | OR_EUR | AdjP_EUR | Sig_AFR | Sig_EUR | Sig_Category | log2_OR_AFR | log2_OR_EUR |
|---|---|---|---|---|---|---|---|---|---|
| **Aliarcobacter butzleri** | 27.51621806 | 4.04E-07 | 17.27586805 | 0.605162401 | TRUE | FALSE | Significant AFR only | 4.782210289 | 4.110686298 |
| **Aliarcobacter cryaerophilus** | Inf | 1.45E-06 | 5.485860934 | 1 | TRUE | FALSE | Significant AFR only | Inf | 2.45571805 |
| **Amedibacterium intestinale** | 4.417586529 | 0.035643216 | 3.718713027 | 1 | TRUE | FALSE | Significant AFR only | 2.143258394 | 1.894803419 |
| **Anaerococcus obesiensis** | 5.174769724 | 0.037838684 | 1.6685911 | 1 | TRUE | FALSE | Significant AFR only | 2.371494664 | 0.738630455 |
| **Aspergillus luchuensis** | 57.32389206 | 4.95E-07 | 3.610815266 | 1 | TRUE | FALSE | Significant AFR only | 5.841064661 | 1.852324612 |
| **Bacteroides coprosuis** | 5.711240889 | 0.047465448 | 2.234095803 | 1 | TRUE | FALSE | Significant AFR only | 2.513804236 | 1.159691053 |
| **Bacteroides helcogenes** | 5.724182602 | 0.022853574 | 1.521345782 | 1 | TRUE | FALSE | Significant AFR only | 2.517069695 | 0.605348096 |
| **Bacteroides heparinolyticus** | 5.174769724 | 0.037838684 | 2.234095803 | 1 | TRUE | FALSE | Significant AFR only | 2.371494664 | 1.159691053 |
| **Bifidobacterium breve** | 5.711240889 | 0.047465448 | 0.447608379 | 1 | TRUE | FALSE | Significant AFR only | 2.513804236 | -1.159691053 |
| **Blautia wexlerae** | 5.724182602 | 0.022853574 | 1 | 1 | TRUE | FALSE | Significant AFR only | 2.517069695 | 0 |
| **Brachyspira pilosicoli** | 8.703152729 | 0.000594823 | 4.949396405 | 1 | TRUE | FALSE | Significant AFR only | 3.121538114 | 2.307252595 |
| **Butyricimonas faecalis** | 3.799109852 | 0.047465448 | 1 | 1 | TRUE | FALSE | Significant AFR only | 1.925661428 | 0 |
| **Campylobacter concisus** | 30.71233169 | 2.11E-07 | 17.27586805 | 0.605162401 | TRUE | FALSE | Significant AFR only | 4.940746141 | 4.110686298 |
| **Campylobacter jejuni** | 5.977354684 | 0.009496572 | Inf | 0.808630031 | TRUE | FALSE | Significant AFR only | 2.579507152 | Inf |

| | | | | | | | | | |
|---|---|---|---|---|---|---|---|---|---|
| **Campylobacter ureolyticus** | 9.488561446 | 0.002264161 | 5.430472516 | 1 | TRUE | FALSE | Significant AFR only | 3.246189378 | 2.441077735 |
| **Candida albicans** | 11.57386941 | 3.08E-05 | 11.63669634 | 1 | TRUE | FALSE | Significant AFR only | 3.532799366 | 3.54060963 |
| **Candida dubliniensis** | 34.45967576 | 1.09E-07 | 17.27586805 | 0.605162401 | TRUE | FALSE | Significant AFR only | 5.106837222 | 4.110686298 |
| **Citrobacter freundii** | 9.444677656 | 0.00016938 | 3.610815266 | 1 | TRUE | FALSE | Significant AFR only | 3.239501559 | 1.852324612 |
| **Clostridium argentinense** | 28.14001346 | 2E-06 | 27.32632049 | 0.436008573 | TRUE | FALSE | Significant AFR only | 4.814551114 | 4.772219308 |
| **Clostridium baratii** | 24.75774648 | 7.59E-07 | 11.63669634 | 1 | TRUE | FALSE | Significant AFR only | 4.629808097 | 3.54060963 |
| **Clostridium butyricum** | Inf | 1.85E-05 | Inf | 1 | TRUE | FALSE | Significant AFR only | Inf | Inf |
| **Clostridium cadaveris** | 11.46455832 | 0.000558862 | 2.234095803 | 1 | TRUE | FALSE | Significant AFR only | 3.51910887 | 1.159691053 |
| **Clostridium neonatale** | 20.16839404 | 4.04E-07 | 8.153062604 | 1 | TRUE | FALSE | Significant AFR only | 4.334024305 | 3.027342093 |
| **Clostridium tetani** | 18.36070331 | 1.07E-05 | 5.430472516 | 1 | TRUE | FALSE | Significant AFR only | 4.198549417 | 2.441077735 |
| **Coprobacter secundus** | 9.75174988 | 0.010179119 | 1.521345782 | 1 | TRUE | FALSE | Significant AFR only | 3.285661123 | 0.605348096 |
| **Cryptosporidium parvum** | 10.53242705 | 0.000133743 | 3.718713027 | 1 | TRUE | FALSE | Significant AFR only | 3.396766018 | 1.894803419 |
| **Enterobacter cloacae** | 8.406447906 | 0.001692405 | 1 | 1 | TRUE | FALSE | Significant AFR only | 3.071496327 | 0 |
| **Enterobacter hormaechei** | 7.781277087 | 0.000769098 | 8.018207377 | 1 | TRUE | FALSE | Significant AFR only | 2.960006954 | 3.003279731 |
| **Escherichia albertii** | 7.650853927 | 0.003417727 | 3.718713027 | 1 | TRUE | FALSE | Significant AFR only | 2.935620779 | 1.894803419 |
| **Escherichia fergusonii** | 7.05880222 | 0.016324413 | 0.680427209 | 1 | TRUE | FALSE | Significant AFR only | 2.819423399 | -0.55548726<br>2 |
| **Finegoldia magna** | 6.726820575 | 0.001161248 | 1 | 1 | TRUE | FALSE | Significant AFR only | 2.749924778 | 0 |
| **Fusobacterium canifelinum** | 6.568186846 | 0.004730763 | 1.469664921 | 1 | TRUE | FALSE | Significant AFR only | 2.715495168 | 0.555487262 |
| **Fusobacterium hwasookii** | 14.38874962 | 5.27E-06 | 3.718713027 | 1 | TRUE | FALSE | Significant AFR only | 3.846869323 | 1.894803419 |
| **Fusobacterium necrophorum** | 5.339593898 | 0.011810108 | 5.430472516 | 1 | TRUE | FALSE | Significant AFR only | 2.416730022 | 2.441077735 |
| **Fusobacterium pseudoperiodonticum** | 15.9991855 | 0.000282757 | Inf | 1 | TRUE | FALSE | Significant AFR only | 3.999926556 | Inf |
| **Fusobacterium sp. oral taxon 203** | 4.471449285 | 0.047465448 | 2.158165565 | 1 | TRUE | FALSE | Significant AFR only | 2.160742513 | 1.109805546 |
| **Fusobacterium ulcerans** | 28.14001346 | 2E-06 | Inf | 0.047392236 | TRUE | TRUE | Significant Both | 4.814551114 | Inf |
| **Klebsiella michiganensis** | 4.934218786 | 0.030057817 | 1.469664921 | 1 | TRUE | FALSE | Significant AFR only | 2.302821687 | 0.555487262 |
| **Klebsiella quasipneumoniae** | 5.912474868 | 0.0038523 | 1 | 1 | TRUE | FALSE | Significant AFR only | 2.563762146 | 0 |
| **Lachnoanaerobaculum umeaense** | 4.471449285 | 0.047465448 | 3.274232944 | 1 | TRUE | FALSE | Significant AFR only | 2.160742513 | 1.711156966 |
| **Lacrimispora sphenoides** | 9.75174988 | 0.010179119 | 1.469664921 | 1 | TRUE | FALSE | Significant AFR only | 3.285661123 | 0.555487262 |
| **Lactococcus lactis** | 14.54855572 | 0.0006318 | 3.610815266 | 1 | TRUE | FALSE | Significant AFR only | 3.862804034 | 1.852324612 |
| **Leptotrichia hofstadii** | 5.174769724 | 0.037838684 | 3.610815266 | 1 | TRUE | FALSE | Significant AFR only | 2.371494664 | 1.852324612 |
| **Leptotrichia hongkongensis** | 7.05880222 | 0.016324413 | 3.610815266 | 1 | TRUE | FALSE | Significant AFR only | 2.819423399 | 1.852324612 |
| **Leptotrichia shahii** | 7.821539299 | 0.034675712 | Inf | 1 | TRUE | FALSE | Significant AFR only | 2.967452562 | Inf |
| **Leptotrichia sp. oral taxon 212** | 4.417586529 | 0.035643216 | 11.63669634 | 1 | TRUE | FALSE | Significant AFR only | 2.143258394 | 3.54060963 |
| **Leptotrichia sp. oral taxon 218** | 7.821539299 | 0.034675712 | Inf | 1 | TRUE | FALSE | Significant AFR only | 2.967452562 | Inf |
| **Leptotrichia sp. oral taxon 847** | 12.49915313 | 0.035643216 | Inf | 1 | TRUE | FALSE | Significant AFR only | 3.643758444 | Inf |
| **Leptotrichia trevisanii** | 7.781277087 | 0.000769098 | Inf | 0.094784472 | TRUE | FALSE | Significant AFR only | 2.960006954 | Inf |

| | | | | | | | | | |
|---|---|---|---|---|---|---|---|---|---|
| **Leptotrichia wadei** | 7.767960717 | 0.000281396 | 8.153062604 | 1 | TRUE | FALSE | Significant AFR only | 2.957535905 | 3.027342093 |
| **Ligilactobacillus salivarius** | 9.982457853 | 4.74E-05 | 17.27586805 | 0.605162401 | TRUE | FALSE | Significant AFR only | 3.319395076 | 4.110686298 |
| **Limosilactobacillus reuteri** | 6.44128922 | 0.003312935 | 5.485860934 | 1 | TRUE | FALSE | Significant AFR only | 2.687349472 | 2.45571805 |
| **Listeria monocytogenes** | 9.568381587 | 0.000279208 | Inf | 0.479188166 | TRUE | FALSE | Significant AFR only | 3.258274925 | Inf |
| **Monoglobus pectinilyticus** | 7.922224596 | 0.00121112 | 1 | 1 | TRUE | FALSE | Significant AFR only | 2.985905602 | 0 |
| **Morganella morganii** | 8.725417919 | 0.00020608 | 2.536920682 | 1 | TRUE | FALSE | Significant AFR only | 3.125224233 | 1.343078413 |
| **Paeniclostridium sordellii** | 14.17643254 | 0.020065981 | Inf | 1 | TRUE | FALSE | Significant AFR only | 3.825422623 | Inf |
| **Parabacteroides goldsteinii** | 4.187716298 | 0.020824216 | 0.680427209 | 1 | TRUE | FALSE | Significant AFR only | 2.066163708 | -0.555487262 |
| **Paraclostridium bifermentans** | 28.14001346 | 2E-06 | Inf | 0.479188166 | TRUE | FALSE | Significant AFR only | 4.814551114 | Inf |
| **Parvimonas micra** | 21.19032077 | 3.01E-05 | Inf | 1 | TRUE | FALSE | Significant AFR only | 4.405333522 | Inf |
| **Peptostreptococcus sp. CBA3647** | 5.711240889 | 0.047465448 | 1.6685911 | 1 | TRUE | FALSE | Significant AFR only | 2.513804236 | 0.738630455 |
| **Prevotella melaninogenica** | 3.61596395 | 0.037838684 | 2.536920682 | 1 | TRUE | FALSE | Significant AFR only | 1.854380295 | 1.343078413 |
| **Proteus mirabilis** | 7.650853927 | 0.003417727 | 5.485860934 | 1 | TRUE | FALSE | Significant AFR only | 2.935620779 | 2.45571805 |
| **Proteus terrae** | 4.489719192 | 0.012007777 | 0.657312763 | 1 | TRUE | FALSE | Significant AFR only | 2.166625215 | -0.605348096 |
| **Proteus vulgaris** | 11.13336883 | 0.000191818 | 3.610815266 | 1 | TRUE | FALSE | Significant AFR only | 3.476818297 | 1.852324612 |
| **Providencia rettgeri** | Inf | 7.27E-07 | Inf | 0.808630031 | TRUE | FALSE | Significant AFR only | Inf | Inf |
| **Romboutsia ilealis** | 7.076886749 | 0.001567927 | 5.430472516 | 1 | TRUE | FALSE | Significant AFR only | 2.823114832 | 2.441077735 |
| **Romboutsia sp. CE17** | 11.57386941 | 3.08E-05 | 8.153062604 | 1 | TRUE | FALSE | Significant AFR only | 3.532799366 | 3.027342093 |
| **Salmonella enterica** | Inf | 0.029931986 | 0 | 1 | TRUE | FALSE | Significant AFR only | Inf | #NAME? |
| **Selenomonas timonae** | 7.821539299 | 0.034675712 | 1 | 1 | TRUE | FALSE | Significant AFR only | 2.967452562 | 0 |
| **Serratia marcescens** | 9.231862527 | 0.000838556 | 1.6685911 | 1 | TRUE | FALSE | Significant AFR only | 3.206621741 | 0.738630455 |
| **Staphylococcus epidermidis** | 22.08240758 | 0.001220975 | Inf | 1 | TRUE | FALSE | Significant AFR only | 4.464825569 | Inf |
| **Staphylococcus hominis** | 7.767960717 | 0.000281396 | 2.536920682 | 1 | TRUE | FALSE | Significant AFR only | 2.957535905 | 1.343078413 |
| **Streptococcus cristatus** | 7.808131201 | 0.009072533 | 2.162188194 | 1 | TRUE | FALSE | Significant AFR only | 2.964977295 | 1.112492099 |
| **Streptococcus equi** | 9.75174988 | 0.010179119 | 3.718713027 | 1 | TRUE | FALSE | Significant AFR only | 3.285661123 | 1.894803419 |
| **Streptococcus milleri** | 4.934218786 | 0.030057817 | 1 | 1 | TRUE | FALSE | Significant AFR only | 2.302821687 | 0 |
| **Streptococcus pneumoniae** | 4.068957962 | 0.022280044 | 1.469664921 | 1 | TRUE | FALSE | Significant AFR only | 2.024659375 | 0.555487262 |
| **Streptococcus pyogenes** | 21.19032077 | 3.01E-05 | 3.718713027 | 1 | TRUE | FALSE | Significant AFR only | 4.405333522 | 1.894803419 |
| **Streptococcus suis** | 9.444677656 | 0.00016938 | 3.274232944 | 1 | TRUE | FALSE | Significant AFR only | 3.239501559 | 1.711156966 |
| **Streptococcus thermophilus** | 4.899132487 | 0.014349572 | 1 | 1 | TRUE | FALSE | Significant AFR only | 2.292526307 | 0 |
| **Thomasclavelia spiroformis** | 7.650853927 | 0.003417727 | 2.234095803 | 1 | TRUE | FALSE | Significant AFR only | 2.935620779 | 1.159691053 |
| **Treponema denticola** | 4.894741043 | 0.006624424 | 8.018207377 | 1 | TRUE | FALSE | Significant AFR only | 2.291232536 | 3.003279731 |
| **Trypanosoma brucei** | 29.67925321 | 0.000136565 | Inf | 1 | TRUE | FALSE | Significant AFR only | 4.891382886 | Inf |
| **Yersinia enterocolitica** | 6.131074535 | 0.001477116 | Inf | 0.479188166 | TRUE | FALSE | Significant AFR only | 2.616139943 | Inf |

Table S10. Species–cluster assignments from tumor and matched non-tumor WGS

| Species name | Cluster |
|---|---|
| Bifidobacterium breve | Cluster 5 |
| Thomasclavelia ramosa | Cluster 5 |
| Adlercreutzia equolifaciens | Cluster 5 |
| Gordonibacter pamelaeae | Cluster 5 |
| Phocaeicola coprocola | Cluster 5 |
| Adlercreutzia hattorii | Cluster 5 |
| Blautia liquoris | Cluster 5 |
| Clostridium sp. SY8519 | Cluster 5 |
| Bifidobacterium dentium | Cluster 5 |
| Sutterella wadsworthensis | Cluster 5 |
| Eubacterium callanderi | Cluster 5 |
| Phascolarctobacterium sp. Marseille−Q4147 | Cluster 5 |
| Emergencia timonensis | Cluster 5 |
| Escherichia coli | Cluster 5 |
| Anaerostipes caccae | Cluster 5 |
| Eubacterium maltosivorans | Cluster 5 |
| Lacrimispora saccharolytica | Cluster 5 |
| Acutalibacter muris | Cluster 5 |
| Eubacterium sp. c−25 | Cluster 5 |
| Alistipes sp. dk3624 | Cluster 5 |
| Desulfovibrio fairfieldensis | Cluster 5 |
| Acidaminococcus intestini | Cluster 5 |
| Desulfovibrio piger | Cluster 5 |
| Phascolarctobacterium succinatutens | Cluster 5 |
| Bacteroides sp. BFG−257 | Cluster 5 |
| Bifidobacterium adolescentis | Cluster 5 |
| Bifidobacterium pseudocatenulatum | Cluster 5 |
| Actinomyces odontolytica | Cluster 4 |
| Eikenella corrodens | Cluster 4 |
| Streptococcus oralis | Cluster 4 |
| Streptococcus agalactiae | Cluster 4 |
| Streptococcus pasteurianus | Cluster 4 |
| Fusobacterium hwasookii | Cluster 4 |
| Fusobacterium pseudoperiodonticum | Cluster 4 |
| Prevotella sp. oral taxon 299 | Cluster 4 |
| Filifactor alocis | Cluster 4 |

| | |
|---|---|
| Mogibacterium pumilum | Cluster 4 |
| Fretibacterium fastidiosum | Cluster 4 |
| Prevotella ruminicola | Cluster 4 |
| Prevotella sp. E13−17 | Cluster 4 |
| Murdochiella vaginalis | Cluster 4 |
| Tannerella serpentiformis | Cluster 4 |
| Gemella morbillorum | Cluster 4 |
| Streptococcus anginosus | Cluster 4 |
| Prevotella histicola | Cluster 4 |
| Streptococcus constellatus | Cluster 4 |
| Fusobacterium sp. oral taxon 203 | Cluster 4 |
| Mogibacterium diversum | Cluster 4 |
| Fusobacterium canifelinum | Cluster 4 |
| Parvimonas micra | Cluster 4 |
| Prevotella bryantii | Cluster 4 |
| Prevotella sp. E2−28 | Cluster 4 |
| Prevotella sp. E9−3 | Cluster 4 |
| Pseudoprevotella muciniphila | Cluster 4 |
| Prevotella fusca | Cluster 4 |
| Prevotella scopos | Cluster 4 |
| Prevotella sp. E15−22 | Cluster 4 |
| Prevotella sp. oral taxon 475 | Cluster 4 |
| Porphyromonas endodontalis | Cluster 4 |
| Tannerella forsythia | Cluster 4 |
| Bacteroides sp. PHL 2737 | Cluster 3 |
| Prevotella copri | Cluster 3 |
| Bacteroides sp. ZJ−18 | Cluster 3 |
| Parabacteroides faecis | Cluster 3 |
| Bacteroides caecimuris | Cluster 3 |
| Bacteroides faecium | Cluster 3 |
| Lachnoclostridium [Clostridium] scindens | Cluster 3 |
| Bacteroides intestinalis | Cluster 3 |
| Enterocloster asparagiformis | Cluster 3 |
| Enterocloster bolteae | Cluster 3 |
| Butyricimonas virosa | Cluster 3 |
| Alistipes senegalensis | Cluster 3 |
| Alistipes indistinctus | Cluster 3 |
| Phocaeicola salanitronis | Cluster 3 |
| Flintibacter sp. KGMB00164 | Cluster 3 |
| Flavonifractor plautii | Cluster 3 |
| Parabacteroides merdae | Cluster 3 |
| Bacteroides sp. HF−162 | Cluster 3 |

| | |
|---|---|
| Bacteroides nordii | Cluster 3 |
| Alistipes onderdonkii | Cluster 3 |
| Bacteroides sp. A1C1 | Cluster 3 |
| Bacteroides sp. CACC 737 | Cluster 3 |
| Vescimonas coprocola | Cluster 3 |
| Vescimonas fastidiosa | Cluster 3 |
| Oscillibacter hominis | Cluster 3 |
| Dysosmobacter sp. Marseille−Q4140 | Cluster 3 |
| Pusillibacter faecalis | Cluster 3 |
| Bacteroides stercoris | Cluster 3 |
| Phocaeicola vulgatus | Cluster 3 |
| Bacteroides uniformis | Cluster 3 |
| Phocaeicola dorei | Cluster 3 |
| Ruthenibacterium lactatiformans | Cluster 3 |
| Dysosmobacter welbionis | Cluster 3 |
| Intestinimonas butyriciproducens | Cluster 3 |
| Alistipes megaguti | Cluster 3 |
| Butyricimonas faecalis | Cluster 3 |
| Phocaeicola coprophilus | Cluster 3 |
| Alistipes shahii | Cluster 3 |
| Alistipes finegoldii | Cluster 3 |
| Odoribacter splanchnicus | Cluster 3 |
| Bacteroides cellulosilyticus | Cluster 3 |
| Bacteroides humanifaecis | Cluster 3 |
| Parabacteroides johnsonii | Cluster 3 |
| Bacteroides caccae | Cluster 3 |
| Bacteroides eggerthii | Cluster 3 |
| Paraprevotella xylaniphila | Cluster 3 |
| Bacteroides helcogenes | Cluster 3 |
| Barnesiella viscericola | Cluster 3 |
| Bacteroides sp. M10 | Cluster 3 |
| Bacteroides luhongzhouii | Cluster 3 |
| Bacteroides sp. D2 | Cluster 3 |
| Enterocloster clostridioformis | Cluster 3 |
| Hungatella hathewayi | Cluster 3 |
| Bacteroides sp. DH3716P | Cluster 3 |
| Bacteroides faecis | Cluster 3 |
| Bacteroides zhangwenhongii | Cluster 3 |
| Parabacteroides distasonis | Cluster 3 |
| Bacteroides thetaiotaomicron | Cluster 3 |
| Bacteroides ovatus | Cluster 3 |

| | |
|---|---|
| Bacteroides xylanisolvens | Cluster 3 |
| Bacteroides fragilis | Cluster 3 |
| Bacteroides salyersiae | Cluster 3 |
| Lachnoclostridium sp. YL32 | Cluster 3 |
| Ruminococcus champanellensis | Cluster 3 |
| Parabacteroides goldsteinii | Cluster 3 |
| Parabacteroides sp. CT06 | Cluster 3 |
| Alistipes ihumii | Cluster 3 |
| Christensenella sp. Marseille−P3954 | Cluster 3 |
| Christensenella minuta | Cluster 3 |
| Desulfovibrio desulfuricans | Cluster 3 |
| Collinsella aerofaciens | Cluster 3 |
| Coprococcus comes | Cluster 3 |
| Clostridium sp. M62/1 | Cluster 3 |
| Subdoligranulum variabile | Cluster 3 |
| Blautia obeum | Cluster 3 |
| Faecalibacterium prausnitzii | Cluster 3 |
| Coprococcus catus | Cluster 3 |
| Roseburia hominis | Cluster 3 |
| Roseburia intestinalis | Cluster 3 |
| Simiaoa sunii | Cluster 3 |
| Mediterraneibacter [Ruminococcus] torques | Cluster 3 |
| Dorea formicigenerans | Cluster 3 |
| Dorea longicatena | Cluster 3 |
| Faecalibacterium sp. I4−1−79 | Cluster 3 |
| Faecalibacterium sp. I4−3−84 | Cluster 3 |
| Faecalibacterium sp. I2−3−92 | Cluster 3 |
| Faecalibacterium sp. IP−3−29 | Cluster 3 |
| Faecalitalea cylindroides | Cluster 3 |
| Faecalibacterium sp. IP−1−18 | Cluster 3 |
| Faecalibacterium duncaniae | Cluster 3 |
| Faecalibacterium sp. I3−3−89 | Cluster 3 |
| Eggerthella lenta | Cluster 3 |
| Mediterraneibacter [Ruminococcus] gnavus | Cluster 3 |
| Faecalicatena sp. Marseille−Q4148 | Cluster 3 |
| Thomasclavelia [Clostridium] innocuum | Cluster 3 |
| Anaerotruncus colihominis | Cluster 3 |
| Massilistercora timonensis | Cluster 3 |

| | |
|---|---|
| Alistipes communis | Cluster 3 |
| Alistipes dispar | Cluster 3 |
| Faecalibacterium sp. HTF−F | Cluster 3 |
| Faecalibacterium sp. I3−3−33 | Cluster 3 |
| Blautia wexlerae | Cluster 3 |
| Blautia sp. SC05B48 | Cluster 3 |
| Mediterraneibacter [Ruminococcus] lactaris | Cluster 3 |
| Butyrivibrio crossotus | Cluster 3 |
| Anaerostipes hadrus | Cluster 3 |
| Anaerobutyricum hallii | Cluster 3 |
| Lachnospira eligens | Cluster 3 |
| Lachnoclostridium phocaeense | Cluster 3 |
| Akkermansia muciniphila | Cluster 3 |
| Solibaculum mannosilyticum | Cluster 3 |
| Roseburia sp. NSJ−69 | Cluster 3 |
| Marvinbryantia formatexigens | Cluster 3 |
| Wansuia hejianensis | Cluster 3 |
| Blautia argi | Cluster 3 |
| Coprococcus eutactus | Cluster 3 |
| Longicatena caecimuris | Cluster 3 |
| Wujia chipingensis | Cluster 3 |
| Blautia sp. NBRC 113351 | Cluster 3 |
| Lachnoclostridium [Clostridium] hylemonae | Cluster 3 |
| Blautia hansenii | Cluster 3 |
| Blautia producta | Cluster 3 |
| Bifidobacterium longum | Cluster 3 |
| Coprococcus sp. ART55/1 | Cluster 3 |
| Ruminococcus bicirculans | Cluster 3 |
| Eubacterium hominis | Cluster 3 |
| Catenibacterium mitsuokai | Cluster 3 |
| Eubacterium sp. MSJ−33 | Cluster 3 |
| Shigella flexneri | Cluster 3 |
| Blautia pseudococcoides | Cluster 3 |
| Sellimonas intestinalis | Cluster 3 |
| Phascolarctobacterium faecium | Cluster 3 |
| Qiania dongpingensis | Cluster 3 |
| Bifidobacterium bifidum | Cluster 3 |
| Eubacterium ventriosum | Cluster 3 |
| Prevotella multiformis | Cluster 2 |
| Fusobacterium nucleatum | Cluster 2 |
| Prevotella veroralis | Cluster 2 |

| | |
|---|---|
| Prevotella melaninogenica | Cluster 2 |
| Hoylesella enoeca | Cluster 2 |
| Prevotella denticola | Cluster 2 |
| Prevotella oris | Cluster 2 |
| Selenomonas sputigena | Cluster 2 |
| Prevotella sp. Rep29 | Cluster 2 |
| Pyramidobacter piscolens | Cluster 2 |
| Hoylesella buccalis | Cluster 2 |
| Porphyromonas somerae | Cluster 2 |
| Dialister succinatiphilus | Cluster 2 |
| Bacteroides heparinolyticus | Cluster 2 |
| Bacteroides zoogleoformans | Cluster 2 |
| Dialister pneumosintes | Cluster 2 |
| Porphyromonas asaccharolytica | Cluster 2 |
| Prevotella nigrescens | Cluster 2 |
| Prevotella corporis | Cluster 2 |
| Prevotella dentalis | Cluster 2 |
| Prevotella jejuni | Cluster 2 |
| Porphyromonas gingivalis | Cluster 2 |
| Prevotella intermedia | Cluster 2 |
| Fusobacterium periodonticum | Cluster 1 |
| Selenomonas sp. oral taxon 126 | Cluster 1 |
| Treponema socranskii | Cluster 1 |
| Treponema sp. Marseille−Q4132 | Cluster 1 |
| Streptococcus sp. NSJ−72 | Cluster 1 |
| Streptococcus intermedius | Cluster 1 |
| Streptococcus milleri | Cluster 1 |
| Prevotella herbatica | Cluster 1 |
| Gemella sp. oral taxon 928 | Cluster 1 |
| Treponema vincentii | Cluster 1 |
| Fusobacterium necrophorum | Cluster 1 |
| Selenomonas sp. oral taxon 920 | Cluster 1 |
| Selenomonas timonae | Cluster 1 |
| Fusobacterium gonidiaformans | Cluster 1 |
| Selenomonas sp. oral taxon 136 | Cluster 1 |
| Campylobacter showae | Cluster 1 |
| Campylobacter rectus | Cluster 1 |
| Lachnoanaerobaculum umeaense | Cluster 1 |

| | |
|---|---|
| Leptotrichia sp. oral taxon 212 | Cluster 1 |
| Leptotrichia sp. oral taxon 218 | Cluster 1 |
| Leptotrichia sp. oral taxon 847 | Cluster 1 |
| Leptotrichia shahii | Cluster 1 |
| Leptotrichia sp. oral taxon 498 | Cluster 1 |
| Megasphaera massiliensis | Cluster 1 |
| Streptococcus mutans | Cluster 1 |
| Granulicatella adiacens | Cluster 1 |
| Streptococcus pneumoniae | Cluster 1 |
| Treponema denticola | Cluster 1 |
| Streptococcus equi | Cluster 1 |
| Streptococcus pyogenes | Cluster 1 |
| Cruoricaptor ignavus | Cluster 1 |
| Ezakiella coagulans | Cluster 1 |
| Granulicatella elegans | Cluster 1 |
| Peptostreptococcus sp. CBA3647 | Cluster 1 |
| Campylobacter gracilis | Cluster 1 |
| Muribaculum gordoncarteri | Cluster 1 |
| Anaerotignum propionicum | Cluster 1 |
| Duncaniella dubosii | Cluster 1 |
| Muribaculum intestinale | Cluster 1 |
| Prevotella sp. E13−3 | Cluster 1 |
| Prevotella sp. E2−25 | Cluster 1 |
| Dialister hominis | Cluster 1 |
| Selenomonas sp. oral taxon 478 | Cluster 1 |
| Leptotrichia trevisanii | Cluster 1 |
| Leptotrichia hofstadii | Cluster 1 |
| Leptotrichia wadei | Cluster 1 |
| Leptotrichia buccalis | Cluster 1 |
| Leptotrichia hongkongensis | Cluster 1 |
| Faecalibacillus intestinalis | Cluster 1 |
| Haemophilus parainfluenzae | Cluster 1 |
| Bifidobacterium catenulatum | Cluster 1 |
| Streptococcus sp. HSISM1 | Cluster 1 |
| Streptococcus sp. LPB0220 | Cluster 1 |
| Streptococcus parasanguinis | Cluster 1 |
| Streptococcus salivarius | Cluster 1 |
| Carjivirus communis | Cluster 1 |
| Coprobacter secundus | Cluster 1 |

| | |
|---|---|
| Desulfovibrio sp. G11 | Cluster 1 |
| Ruminococcus gauvreauii | Cluster 1 |
| Escherichia fergusonii | Cluster 1 |
| Shigella sonnei | Cluster 1 |
| Shigella boydii | Cluster 1 |
| Shigella dysenteriae | Cluster 1 |
| Pseudobutyrivibrio xylanivorans | Cluster 1 |
| Pseudomonas aeruginosa | Cluster 1 |
| Campylobacter ureolyticus | Cluster 1 |
| Anaerococcus obesiensis | Cluster 1 |
| Anaerococcus vaginalis | Cluster 1 |
| Gemella haemolysans | Cluster 1 |
| Pseudomonas fluorescens | Cluster 1 |
| Schaalia turicensis | Cluster 1 |
| Finegoldia magna | Cluster 1 |
| Fusobacterium ulcerans | Cluster 1 |
| Bacteroides coprosuis | Cluster 1 |
| Peptoniphilus sp. SAHP1 | Cluster 1 |
| Roseolovirus Human betaherpesvirus 6B | Cluster 1 |
| Roseolovirus Human betaherpesvirus 7 | Cluster 1 |
| Stenotrophomonas maltophilia | Cluster 1 |
| Sphingomonas sanguinis | Cluster 1 |
| Sphingomonas sp. LK11 | Cluster 1 |
| Ligilactobacillus salivarius | Cluster 1 |
| Clostridium neonatale | Cluster 1 |
| Clostridium tetani | Cluster 1 |
| Clostridium sporogenes | Cluster 1 |
| Paeniclostridium sordellii | Cluster 1 |
| Paraclostridium bifermentans | Cluster 1 |
| Serratia marcescens | Cluster 1 |
| Lactobacillus johnsonii | Cluster 1 |
| Morganella morganii | Cluster 1 |
| Bacillus mycoides | Cluster 1 |
| Providencia rettgeri | Cluster 1 |
| Aliarcobacter butzleri | Cluster 1 |
| Brachyspira pilosicoli | Cluster 1 |
| Staphylococcus hominis | Cluster 1 |
| Bacillus thuringiensis | Cluster 1 |
| Aliarcobacter cryaerophilus | Cluster 1 |
| Staphylococcus aureus | Cluster 1 |
| Yersinia enterocolitica | Cluster 1 |

| | |
|---|---|
| Clostridium beijerinckii | Cluster 1 |
| Clostridium botulinum | Cluster 1 |
| Clostridium baratii | Cluster 1 |
| Clostridium argentinense | Cluster 1 |
| Helicobacter pylori | Cluster 1 |
| Pseudomonas putida | Cluster 1 |
| Proteus terrae | Cluster 1 |
| Bacillus paranthracis | Cluster 1 |
| Bacillus subtilis | Cluster 1 |
| Listeria monocytogenes | Cluster 1 |
| Enterococcus faecalis | Cluster 1 |
| Clostridium sp. MD294 | Cluster 1 |
| Salmonella enterica | Cluster 1 |
| Clostridium sp. LQ25 | Cluster 1 |
| Romboutsia sp. CE17 | Cluster 1 |
| Campylobacter jejuni | Cluster 1 |
| Clostridium perfringens | Cluster 1 |
| Hathewaya histolytica | Cluster 1 |
| Stenotrophomonas sp. SBJS02 | Cluster 1 |
| Bacillus amyloliquefaciens | Cluster 1 |
| Clostridium cadaveris | Cluster 1 |
| Proteus mirabilis | Cluster 1 |
| Candida albicans | Cluster 1 |
| Limosilactobacillus reuteri | Cluster 1 |
| Lactococcus lactis | Cluster 1 |
| Terrisporobacter hibernicus | Cluster 1 |
| Bacillus tropicus | Cluster 1 |
| Staphylococcus epidermidis | Cluster 1 |
| Clostridium butyricum | Cluster 1 |
| Candida dubliniensis | Cluster 1 |
| Clostridium sp. DL−VIII | Cluster 1 |
| Aspergillus luchuensis | Cluster 1 |
| Trypanosoma brucei | Cluster 1 |
| Proteus vulgaris | Cluster 1 |
| Cryptosporidium parvum | Cluster 1 |
| Trichomonas vaginalis | Cluster 1 |
| Neisseria mucosa | Cluster 1 |
| Streptococcus lutetiensis | Cluster 1 |
| Veillonella nakazawae | Cluster 1 |
| Rothia mucilaginosa | Cluster 1 |
| Streptococcus thermophilus | Cluster 1 |
| Streptococcus gallolyticus | Cluster 1 |
| Haemophilus influenzae | Cluster 1 |

| | |
|---|---|
| Neisseria elongata | Cluster 1 |
| Campylobacter concisus | Cluster 1 |
| Gemella sanguinis | Cluster 1 |
| Streptococcus cristatus | Cluster 1 |
| Streptococcus suis | Cluster 1 |
| Veillonella parvula | Cluster 1 |
| Streptococcus vestibularis | Cluster 1 |
| Veillonella sp. S12025−13 | Cluster 1 |
| Veillonella atypica | Cluster 1 |
| Veillonella dispar | Cluster 1 |
| Veillonella rogosae | Cluster 1 |
| Streptococcus mitis | Cluster 1 |
| Streptococcus sanguinis | Cluster 1 |
| Streptococcus australis | Cluster 1 |
| Streptococcus gordonii | Cluster 1 |
| Methanobrevibacter smithii | Cluster 1 |
| Klebsiella oxytoca | Cluster 1 |
| Klebsiella pneumoniae | Cluster 1 |
| Escherichia albertii | Cluster 1 |
| Escherichia marmotae | Cluster 1 |
| Klebsiella variicola | Cluster 1 |
| Lacrimispora sphenoides | Cluster 1 |
| Butyrivibrio fibrisolvens | Cluster 1 |
| Clostridium sp. C1 | Cluster 1 |
| Ruminococcus albus | Cluster 1 |
| Amedibacterium intestinale | Cluster 1 |
| Lacrimispora xylanolytica | Cluster 1 |
| Citrobacter freundii | Cluster 1 |
| Citrobacter braakii | Cluster 1 |
| Enterobacter hormaechei | Cluster 1 |
| Klebsiella quasipneumoniae | Cluster 1 |
| Klebsiella grimontii | Cluster 1 |
| Klebsiella michiganensis | Cluster 1 |
| Romboutsia ilealis | Cluster 1 |
| Enterobacter cloacae | Cluster 1 |
| Monoglobus pectinilyticus | Cluster 1 |
| Helicobacter typhlonius | Cluster 1 |
| Thomasclavelia spiroformis | Cluster 1 |
| Pasteurella multocida | Cluster 1 |
| Clostridioides difficile | Cluster 1 |
| Ruminococcus bovis | Cluster 1 |
| Enterococcus faecium | Cluster 1 |

| | |
|---|---|
| Bifidobacterium pseudolongum | Cluster 1 |
| Eubacterium limosum | Cluster 1 |

Table S11. Regions of recurring large copy number events in the NYGC AFRg patients with MSS tumors.

| The columns are: | |
|---|---|
| Chromosome | The name of the chromosome on which the region is located. |
| Starting position | The starting position of the region on the chromosome. |
| Ending position | The ending position of the region on the chromosome. |
| Type | The type of event that has been identified in the region (Amplification or Deletion). |
| Cytoband | The cytoband associated to the region. |
| Identifier | The unique identifier for the region (only used in the analyses). |
| COSMIC genes | The genes from COSMIC that are present in the region; empty when no gene present |

| Chromosome | Starting position | Ending position | Type | Cytoband | Identifier | COSMIC genes |
|---|---|---|---|---|---|---|
| 1 | 1777362 | 119796153 | Deletion | p36.33-p12 | c1 | ARHGEF10L,ARID1A,ATP1A1,BCL10,CAMTA1,CASP9,CDKN2C,CSF3R,EPS15,FUBP1,JAK1,JUN,LCK,MDS2,MPL,MTOR,MUTYH,MYCL,NRAS,PAX7,PIK3R3,PRDM16,PRDM2,RBM15,RPL22,RPL5,SDHB,SFPQ,SKI,SPEN,STIL,TAL1,TENT5C,THRAP3,TRIM33 |
| 1 | 97788182 | 115432598 | Deletion | p21.3-p13.2 | c2 | NRAS,RBM15,TRIM33 |
| 1 | 144548471 | 181091482 | Amplification | q21.1-q25.3 | c3 | ABL2,ARNT,BCL9,DDR2,FCGR2B,FCRL4,LMNA,MLLT11,MUC1,NTRK1,PBX1,PDE4DIP,PRCC,PRRX1,S100A7,SDHC,SETDB1,TPM3 |
| 1 | 201558793 | 236334938 | Amplification | q32.1-q42.3 | c4 | BTG2,ELF3,ELK4,MDM4,SLC45A3 |
| 1 | 239438247 | 248916508 | Amplification | q43-q44 | c5 | FH,RGS7 |
| 2 | 4188676 | 19623618 | Amplification | p25.3-p24.1 | c6 | MYCN |
| 2 | 49210512 | 89225084 | Amplification | p16.3-p11.2 | c7 | BCL11A,CTNNA2,DCTN1,PCBP1,REL,XPO1 |
| 2 | 143745876 | 242109969 | Amplification | q22.3-q37.3 | c8 | ACKR3,ACSL3,ACVR1,ACVR2A,ATIC,BARD1,CASP8,CD28,CREB1,CUL3,ERBB4,FEV,HOXD11,HOXD13,IDH1,ITGAV,NFE2L2,PAX3,PMS1,SF3B1 |
| 3 | 12690 | 50136751 | Deletion | p26.3-p21.31 | c9 | CCR4,CTNNB1,FANCD2,FBLN2,MLH1,MYD88,NCKIPSD,PPARG,RAF1,RHOA,SETD2,SRGAP3,TGFBR2,VHL,XPC |
| 3 | 8821239 | 198098659 | Deletion | p25.3-q29 | c10 | ATR,BAP1,BCL6,CACNA1D,CBLB,CCR4,CNBP,CTNNB1,EIF4A2,EPHA3,ETV5,FANCD2,FBLN2,FHIT,FOXL2,FOXP1,GATA2,GMPS,GSK3B,IGF2BP2,LPP,MAP3K13,MB21D2,MECOM,MITF,MLF1,MLH1,MYD88,NCKIPSD,PBRM1,PIK3CA,PIK3CB,POLQ,PPARG,RAF1,RHOA,ROBO2,RPN1,SETD2,SOX2,SRGAP3,STAG1,TBL1XR1,TFG,TFRC,TGFBR2,TP63,VHL,WWTR1,XPC |
| 3 | 71123221 | 174528173 | Amplification | p13-q26.31 | c11 | ATR,CBLB,CNBP,EPHA3,FOXL2,FOXP1,GATA2,GMPS,GSK3B,MECOM,MLF1,PIK3CB,POLQ,ROBO2,RPN1,STAG1,TFG,WWTR1 |
| 3 | 175699585 | 198098659 | Amplification | q26.31-q29 | c12 | BCL6,EIF4A2,ETV5,IGF2BP2,LPP,MAP3K13,MB21D2,PIK3CA,SOX2,TBL1XR1,TFRC,TP63 |
| 4 | 72156 | 9155672 | Deletion | p16.3-p16.1 | c13 | FGFR3,NSD2 |
| 4 | 9404218 | 165426755 | Deletion | p16.1-q32.3 | c14 | AFF1,CHIC2,FAT4,FBXW7,FIP1L1,IL2,KDR,KIT,LEF1,N4BP2,PDGFRA,PHOX2B,PTPN13,RAP1GDS1,RHOH,SLC34A2,TEC,TET2 |
| 4 | 74912784 | 86849309 | Deletion | q13.3-q21.3 | c15 | PTPN13 |
| 4 | 91054710 | 156085194 | Deletion | q22.1-q32.1 | c16 | FAT4,FBXW7,IL2,LEF1,RAP1GDS1,TET2 |

| | | | | | | |
|---|---|---|---|---|---|---|
| 4 | 168555439 | 190035460 | Deletion | q32.3-q35.2 | c17 | CASP3,FAT1 |
| 5 | 44922 | 685633 | Amplification | p15.33 | c18 | SDHA |
| 5 | 789923 | 818075 | Amplification | p15.33 | c19 | |
| 5 | 8328588 | 46431572 | Amplification | p15.31-p11 | c20 | CDH10,CTNND2,DROSHA,GOLPH3,IL7R,LIFR,RICTOR,SUB1 |
| 5 | 29966143 | 46431572 | Amplification | p13.3-p11 | c21 | DROSHA,GOLPH3,IL7R,LIFR,RICTOR,SUB1 |
| 5 | 60074483 | 167232830 | Deletion | q12.1-q34 | c22 | ACSL6,AFF4,APC,ARHGAP26,CD74,CSF1R,CTNNA1,EBF1,ITK,PDGFRB,PIK3R1,PWWP2A,RAD50,RASA1 |
| 5 | 60327558 | 84753632 | Deletion | q12.1-q14.3 | c23 | PIK3R1 |
| 5 | 89489567 | 157366098 | Amplification | q14.3-q33.3 | c24 | ACSL6,AFF4,APC,ARHGAP26,CD74,CSF1R,CTNNA1,ITK,PDGFRB,RAD50 |
| 5 | 110481848 | 181170660 | Deletion | q22.1-q35.3 | c25 | ACSL6,AFF4,APC,ARHGAP26,CD74,CSF1R,CTNNA1,EBF1,FGFR4,FLT4,ITK,NPM1,NSD1,PDGFRB,PWWP2A,RAD50,TLX3 |
| 6 | 149609 | 2198407 | Amplification | p25.3 | c26 | IRF4 |
| 6 | 149609 | 32591881 | Deletion | p25.3-p21.32 | c27 | DEK,IRF4 |
| 6 | 14805982 | 32683103 | Amplification | p23-p21.32 | c28 | DEK |
| 6 | 82995298 | 149682891 | Amplification | q14.1-q25.1 | c29 | BCLAF1,CCNC,ECT2L,EPHA7,FOXO3,GOPC,LATS1,MYB,PRDM1,ROS1,RSPO3,SGK1,TNFAIP3 |
| 6 | 85862632 | 161995799 | Deletion | q14.3-q26 | c30 | ARID1B,BCLAF1,CCNC,ECT2L,EPHA7,ESR1,EZR,FOXO3,GOPC,LATS1,MYB,PRDM1,ROS1,RSPO3,SGK1,TNFAIP3 |
| 6 | 103387583 | 144612154 | Deletion | q16.3-q24.2 | c31 | BCLAF1,ECT2L,FOXO3,GOPC,MYB,PRDM1,ROS1,RSPO3,SGK1,TNFAIP3 |
| 6 | 162430247 | 162627482 | Deletion | q26 | c32 | |
| 6 | 162759854 | 170606038 | Deletion | q26-q27 | c33 | AFDN,QKI |
| 7 | 50257 | 49530353 | Amplification | p22.3-p12.2 | c34 | CARD11,ETV1,FKBP9,HNRNPA2B1,HOXA11,HOXA13,HOXA9,JAZF1,MACC1,PMS2,RAC1,SFRP4,TNRC18 |
| 7 | 50257 | 23819598 | Amplification | p22.3-p15.3 | c35 | CARD11,ETV1,MACC1,PMS2,RAC1,TNRC18 |
| 7 | 32827111 | 86203811 | Amplification | p14.3-q21.11 | c36 | EGFR,ELN,FKBP9,GTF2I,HGF,HIP1,IKZF1,SBDS,SFRP4,ZNF479 |
| 7 | 68563372 | 142465064 | Amplification | q11.22-q34 | c37 | AKAP9,BRAF,CDK6,CREB3L2,CUX1,ELN,GRM3,GTF2I,HGF,HIP1,KIAA1549,MET,POT1,SMO,SND1,TRIM24,TRRAP |
| 7 | 125561858 | 159325753 | Amplification | q31.33-q36.3 | c38 | BRAF,CREB3L2,EZH2,FAM131B,KIAA1549,KMT2C,MNX1,SMO,SND1,TRIM24 |
| 8 | 211532 | 4139291 | Deletion | p23.3-p23.2 | c39 | |
| 8 | 4141150 | 4145947 | Deletion | p23.2 | c40 | |
| 8 | 4147499 | 24627540 | Deletion | p23.2-p21.2 | c41 | PCM1 |
| 8 | 16304937 | 32631169 | Deletion | p22-p12 | c42 | LEPROTL1,NRG1,PCM1,WRN |
| 8 | 47920008 | 94383580 | Amplification | q11.21-q22.1 | c43 | CDH17,CHCHD7,CNBD1,HEY1,LYN,NBN,NCOA2,PLAG1,PREX2,RUNX1T1,TCEA1 |
| 8 | 98460437 | 125079578 | Amplification | q22.2-q24.13 | c44 | COX6C,CSMD3,EIF3E,EXT1,PABPC1,RAD21,RSPO2,UBR5 |
| 8 | 114692312 | 139882707 | Amplification | q23.3-q24.3 | c45 | EXT1,FAM135B,MYC,NDRG1,RAD21 |
| 8 | 125641790 | 145074901 | Amplification | q24.13-q24.3 | c46 | FAM135B,MYC,NDRG1,PLEC,RECQL4 |
| 9 | 57539 | 138181722 | Amplification | p24.3-q34.3 | c47 | ABL1,BRD3,CD274,CDKN2A,CNTRL,FANCC,FANCG,FNBP1,GNAQ,JAK2,KLF4,MLLT3,NFIB,NOTCH1,NR4A3,NTRK2,NUP214,PAX5,PDCD1LG2,PPP6C,PSIP1,PTCH1,PTPRD,RALGDS,RXRA,SET,SYK,TAL2,TNC,TSC1,WNK2,XPA |
| 9 | 15525056 | 138181722 | Deletion | p22.3-q34.3 | c48 | ABL1,BRD3,CDKN2A,CNTRL,FANCC,FANCG,FNBP1,GNAQ,KLF4,MLLT3,NOTCH1,NR4A3,NTRK2,NUP214,PAX5,PPP6C,PTCH1,RALGDS,RXRA,SET,SYK,TAL2,TNC,TSC1,WNK2,XPA |
| 10 | 48486 | 28549058 | Deletion | p15.3-p12.1 | c49 | ABI1,GATA3,KLF6,LARP4B,MLLT10 |
| 10 | 51787782 | 119432039 | Deletion | q21.1-q26.11 | c50 | BMPR1A,CCDC6,CPEB3,CYP2C8,FAS,NFKB2,NT5C2,NUTM2B,NUTM2D,PRF1,SHTN1,SUFU,TCF7L2,TET1,TLX1,VTI1A |
| 10 | 61850051 | 100602358 | Deletion | q21.2-q24.31 | c51 | BMPR1A,CPEB3,CYP2C8,FAS,NUTM2B,NUTM2D,PRF1,TET1 |
| 10 | 122385624 | 133382473 | Deletion | q26.13-q26.3 | c52 | MGMT |
| 11 | 199813 | 135075414 | Amplification | p15.5-q25 | c53 | ARHGEF12,ATM,BCL9L,BIRC3,CBL,CCND1,CLP1,CREB3L1,CTNND1,DDB2,DDX10,DDX6,EED,EXT2,FADD,FANCF,FAT3,FEN1,FLI1,FOXR1,KCNJ5,KMT2A,LMO1,LMO2,MALAT1,MAML2,MEN1, |

| | | | | | | |
|---|---|---|---|---|---|---|
| | | | | | | MYOD1,NUMA1,NUP98,PAFAH1B2,PICALM,POU2AF1,RRAS2,SDHAF2,SDHD,WT1,YAP1,ZBTB16 |
| 11 | 199813 | 115428709 | Deletion | p15.5-q23.3 | c54 | ATM,BIRC3,CCND1,CLP1,CREB3L1,CTNND1,DDB2,DDX10,EED,EXT2,FADD,FANCF,FAT3,FEN1,LMO1,LMO2,MALAT1,MAML2,MEN1,MYOD1,NUMA1,NUP98,PICALM,POU2AF1,RRAS2,SDHAF2,SDHD,WT1,YAP1,ZBTB16 |
| 12 | 79927 | 18911792 | Amplification | p13.33-p12.3 | c55 | CCND2,CDKN1B,CHD4,ERC1,ETV6,KDM5A,PTPN6,ZNF384 |
| 12 | 79927 | 34155198 | Deletion | p13.33-p11.1 | c56 | CCND2,CDKN1B,CHD4,ERC1,ETNK1,ETV6,KDM5A,KRAS,PPFIBP1,PTPN6,ZNF384 |
| 12 | 24629874 | 31859532 | Amplification | p12.1-p11.21 | c57 | KRAS,PPFIBP1 |
| 12 | 77362842 | 132868266 | Deletion | q21.2-q24.33 | c58 | ALDH2,BCL7A,BTG1,CHST11,CLIP1,HNF1A,NCOR2,POLE,PTPN11,SETD1B,SH2B3,TBX3,USP44,ZCCHC8 |
| 13 | 18606106 | 25133455 | Amplification | q11-q12.13 | c59 | LATS2,ZMYM2 |
| 13 | 18606106 | 57482593 | Amplification | q11-q21.1 | c60 | BRCA2,CDX2,CYSLTR2,FLT3,FOXO1,LATS2,LCP1,LHFPL6,NBEA,RB1,ZMYM2 |
| 13 | 34444165 | 100095312 | Amplification | q13.2-q32.3 | c61 | CYSLTR2,FOXO1,GPC5,LCP1,LHFPL6,NBEA,RB1,SOX21 |
| 13 | 86193668 | 114343518 | Amplification | q31.1-q34 | c62 | ERCC5,GPC5,SOX21 |
| 14 | 20954507 | 106002195 | Deletion | q11.2-q32.33 | c63 | AKT1,ARHGAP5,BAZ1A,BCL11B,DICER1,FOXA1,GOLGA5,GPHN,HIF1A,HSP90AA1,KTN1,MAX,NIN,NKX2-1,PRKD1,RAD51B,SIX1,TCL1A,TRIP11,TSHR |
| 14 | 22524578 | 45335206 | Deletion | q11.2-q21.2 | c64 | ARHGAP5,BAZ1A,FOXA1,NKX2-1,PRKD1 |
| 14 | 68620345 | 104719996 | Deletion | q24.1-q32.33 | c65 | BCL11B,DICER1,GOLGA5,HSP90AA1,RAD51B,TCL1A,TRIP11,TSHR |
| 15 | 24415432 | 101182262 | Deletion | q11.2-q26.3 | c66 | BLM,BUB1B,CHD2,CRTC3,FES,HMGN2P46,IDH2,KNL1,KNSTRN,MAP2K1,MYO5A,NR2F2,NTRK3,PML,POLG,RAD51,SMAD3,TCF12,USP8 |
| 15 | 41402545 | 101182262 | Deletion | q15.1-q26.3 | c67 | BLM,CHD2,CRTC3,FES,HMGN2P46,IDH2,MAP2K1,MYO5A,NR2F2,NTRK3,PML,POLG,SMAD3,TCF12,USP8 |
| 16 | 34170 | 5291928 | Amplification | p13.3 | c68 | AXIN1,CREBBP,NTHL1,TRAF7,TSC2 |
| 16 | 6715686 | 6805936 | Deletion | p13.3 | c69 | |
| 16 | 7187954 | 26081195 | Amplification | p13.3-p12.1 | c70 | CIITA,ERCC4,GRIN2A,PALB2,PRKCB,RMI2,SNX29,SOCS1,TNFRSF17 |
| 16 | 46426366 | 65756095 | Amplification | q11.2-q21 | c71 | CDH11,CYLD,HERPUD1 |
| 16 | 59452205 | 77753008 | Amplification | q21-q23.1 | c72 | CBFB,CDH1,CDH11,CTCF,RFWD3,ZFHX3 |
| 17 | 151035 | 19684290 | Deletion | p13.3-p11.2 | c73 | FLCN,GAS7,MAP2K4,NCOR1,PER1,RABEP1,TP53,USP6 |
| 17 | 10131098 | 20477140 | Deletion | p13.1-p11.2 | c74 | FLCN,GAS7,MAP2K4,NCOR1,SPECC1 |
| 17 | 21298238 | 21310470 | Deletion | p11.2 | c75 | |
| 17 | 21312847 | 22054380 | Deletion | p11.2 | c76 | |
| 17 | 34301045 | 83204151 | Deletion | q12-q25.3 | c77 | ASPSCR1,AXIN2,BRCA1,BRIP1,CANT1,CCR7,CD79B,CDK12,CLTC,COL1A1,DDX5,ERBB2,ETV4,HLF,IKZF3,KAT7,LASP1,MSI2,PPM1D,PRKAR1A,RAD51C,RAD51D,RARA,RNF213,RNF43,SMARCE1,SPOP,SRSF2,STAT3,STAT5B |
| 17 | 54152817 | 83204151 | Amplification | q22-q25.3 | c78 | ASPSCR1,AXIN2,BRIP1,CANT1,CD79B,CLTC,DDX5,HLF,MSI2,PPM1D,PRKAR1A,RAD51C,RNF213,RNF43,SRSF2 |
| 18 | 64775 | 80256731 | Deletion | p11.32-q23 | c79 | BCL2,DCC,KDSR,MALT1,ROCK1,SETBP1,SMAD2,SMAD4,SS18,ZNF521 |
| 18 | 9373624 | 22390150 | Deletion | p11.22-q11.2 | c80 | ROCK1 |
| 18 | 44752406 | 80256731 | Deletion | q12.3-q23 | c81 | BCL2,DCC,KDSR,MALT1,SETBP1,SMAD2,SMAD4 |
| 18 | 54853449 | 63724733 | Deletion | q21.2-q21.33 | c82 | BCL2,KDSR,MALT1 |
| 18 | 66590871 | 80256731 | Deletion | q22.1-q23 | c83 | |
| 19 | 12384348 | 12434494 | Amplification | p13.2 | c84 | |
| 19 | 24320784 | 58533199 | Amplification | p11-q13.43 | c85 | AKT2,ARHGAP35,BAX,BCL2L12,BCL3,CBLC,CCNE1,CD79A,CEBPA,CEP89,CIC,ERCC2,KLK2,POLD1,PPP2R1A,ZNF331 |
| 19 | 41636104 | 58533199 | Deletion | q13.2-q13.43 | c86 | ARHGAP35,BAX,BCL2L12,BCL3,CBLC,CD79A,CIC,ERCC2,KLK2,POLD1,PPP2R1A,ZNF331 |
| 20 | 68303 | 14129062 | Amplification | p13-p12.1 | c87 | SIRPA |
| 20 | 2661389 | 14746740 | Deletion | p13-p12.1 | c88 | |

| | | | | | | |
|---|---|---|---|---|---|---|
| 20 | 14830831 | 15126785 | Deletion | p12.1 | c89 | |
| 20 | 15255441 | 21643019 | Amplification | p12.1-p11.22 | c90 | CRNKL1 |
| 20 | 16478416 | 28932393 | Amplification | p12.1-q11.1 | c91 | CRNKL1 |
| 20 | 17342388 | 24024025 | Deletion | p12.1-p11.21 | c92 | CRNKL1 |
| 20 | 30458472 | 30990520 | Amplification | q11.21 | c93 | |
| 20 | 30994015 | 32152631 | Amplification | q11.21 | c94 | |
| 20 | 32346265 | 41714702 | Amplification | q11.21-q12 | c95 | ASXL1,MAFB,PLCG1,TOP1 |
| 20 | 35343541 | 64283878 | Amplification | q11.22-q13.33 | c96 | GNAS,MAFB,NFATC2,PLCG1,PTK6,PTPRT,SALL4,SDC4,SS18L1,TOP1 |
| 20 | 44139643 | 46105671 | Amplification | q13.12 | c97 | SDC4 |
| 20 | 46934360 | 62893458 | Amplification | q13.12-q13.33 | c98 | GNAS,NFATC2,SALL4,SS18L1 |
| 20 | 62894671 | 64283878 | Amplification | q13.33 | c99 | PTK6 |
| 21 | 13224688 | 46681423 | Deletion | q11.2-q22.3 | c100 | ERG,OLIG2,RUNX1,TMPRSS2,U2AF1 |
| 21 | 13224688 | 20585151 | Deletion | q11.2-q21.1 | c101 | |
| 22 | 15513541 | 50757736 | Deletion | q11.1-q13.33 | c102 | APOBEC3B,BCR,CHEK2,CLTCL1,DGCR8,EP300,EWSR1,ISX,LZTR1,MAPK1,MN1,MRTFA,MYH9,NF2,PATZ1,PDGFB,ZNRF3 |
| 22 | 38767851 | 50757736 | Deletion | q13.1-q13.33 | c103 | APOBEC3B,EP300,MRTFA,PDGFB |

Table S12. Difference, in the median expression, between samples having an event vs samples without event, in the regions of recurring large copy number events in the NYGC AFRg patients with MSS tumors.

| Core identifier | AMP - Median expression | NEUTRAL - Median expression | DEL - Median expression | p-value | Adjusted p-value | Chromosome | Starting position | Ending position | Core type |
|---|---|---|---|---|---|---|---|---|---|
| c1 | NA | 0,170741468 | -0,28229027 | 0 | 0 | 1 | 1777362 | 119796153 | Deletion |
| c96 | 0,349268556 | -0,484046497 | NA | 0 | 0 | 20 | 35343541 | 64283878 | Amplification |
| c47 | 0,462696494 | -0,075933289 | NA | 4,21E-221 | 1,32E-219 | 9 | 57539 | 138181722 | Amplification |
| c34 | 0,419918892 | -0,219649638 | NA | 2,77E-214 | 6,51E-213 | 7 | 50257 | 49530353 | Amplification |
| c95 | 0,413318792 | -0,538563372 | NA | 1,16E-199 | 2,18E-198 | 20 | 32346265 | 41714702 | Amplification |
| c73 | NA | 0,274838572 | -0,382118319 | 1,57E-192 | 2,46E-191 | 17 | 151035 | 19684290 | Deletion |
| c63 | NA | 0,148562187 | -0,336344974 | 8,70E-188 | 1,17E-186 | 14 | 20954507 | 106002195 | Deletion |
| c66 | NA | 0,165703878 | -0,268192085 | 1,49E-165 | 1,75E-164 | 15 | 24415432 | 101182262 | Deletion |
| c102 | NA | 0,164892461 | -0,279376748 | 6,68E-151 | 6,97E-150 | 22 | 15513541 | 50757736 | Deletion |
| c22 | NA | 0,112473283 | -0,354857389 | 8,41E-150 | 7,91E-149 | 5 | 60074483 | 167232830 | Deletion |
| c25 | NA | 0,129126926 | -0,382963445 | 3,85E-147 | 3,29E-146 | 5 | 110481848 | 181170660 | Deletion |
| c67 | NA | 0,175530202 | -0,23797926 | 3,82E-146 | 2,99E-145 | 15 | 41402545 | 101182262 | Deletion |
| c65 | NA | 0,234545125 | -0,325683121 | 5,34E-136 | 3,86E-135 | 14 | 68620345 | 104719996 | Deletion |
| c98 | 0,32706797 | -0,478478953 | NA | 2,73E-135 | 1,83E-134 | 20 | 46934360 | 62893458 | Amplification |
| c60 | 0,395413877 | -0,168866164 | NA | 3,94E-133 | 2,47E-132 | 13 | 18606106 | 57482593 | Amplification |
| c54 | NA | 0,066919088 | -0,317517312 | 2,16E-126 | 1,27E-125 | 11 | 199813 | 115428709 | Deletion |
| c37 | 0,349527692 | -0,086712977 | NA | 5,14E-124 | 2,84E-123 | 7 | 68563372 | 142465064 | Amplification |
| c79 | NA | 0,214050869 | -0,310539774 | 8,79E-118 | 4,59E-117 | 18 | 64775 | 80256731 | Deletion |
| c81 | NA | 0,408114182 | -0,358378846 | 3,38E-113 | 1,67E-112 | 18 | 44752406 | 80256731 | Deletion |
| c53 | 0,428754667 | -0,02452846 | NA | 4,23E-113 | 1,99E-112 | 11 | 199813 | 135075414 | Amplification |
| c42 | NA | 0,322556208 | -0,46339358 | 5,99E-109 | 2,68E-108 | 8 | 16304937 | 32631169 | Deletion |

| | | | | | | | | | |
|---|---|---|---|---|---|---|---|---|---|
| c35 | 0,415369041 | -0,244545909 | NA | 4,51E-105 | 1,93E-104 | 7 | 50257 | 23819598 | Amplification |
| c3 | 0,273278563 | -0,096382626 | NA | 1,16E-92 | 4,74E-92 | 1 | 144548471 | 181091482 | Amplification |
| c36 | 0,34859083 | -0,136447582 | NA | 1,89E-91 | 7,40E-91 | 7 | 32827111 | 86203811 | Amplification |
| c87 | 0,51542588 | -0,244675991 | NA | 1,70E-89 | 6,40E-89 | 20 | 68303 | 14129062 | Amplification |
| c77 | NA | 0,09258274 | -0,234273488 | 4,88E-88 | 1,76E-87 | 17 | 34301045 | 83204151 | Deletion |
| c103 | NA | 0,218483998 | -0,334480428 | 5,78E-87 | 2,01E-86 | 22 | 38767851 | 50757736 | Deletion |
| c16 | NA | 0,18691766 | -0,32170658 | 3,34E-86 | 1,12E-85 | 4 | 91054710 | 156085194 | Deletion |
| c72 | 0,468279322 | -0,122871146 | NA | 5,57E-86 | 1,81E-85 | 16 | 59452205 | 77753008 | Amplification |
| c61 | 0,374816991 | -0,127805235 | NA | 4,01E-80 | 1,26E-79 | 13 | 34444165 | 100095312 | Amplification |
| c30 | NA | 0,097442209 | -0,415366429 | 8,07E-75 | 2,45E-74 | 6 | 85862632 | 161995799 | Deletion |
| c74 | NA | 0,269636687 | -0,377736969 | 3,64E-74 | 1,07E-73 | 17 | 10131098 | 20477140 | Deletion |
| c38 | 0,362271164 | -0,094737084 | NA | 8,42E-74 | 2,40E-73 | 7 | 125561858 | 159325753 | Amplification |
| c97 | 0,339409815 | -0,546874707 | NA | 1,72E-72 | 4,76E-72 | 20 | 44139643 | 46105671 | Amplification |
| c48 | NA | 0,063818723 | -0,302362697 | 8,92E-69 | 2,40E-68 | 9 | 15525056 | 138181722 | Deletion |
| c43 | 0,371725696 | -0,158823052 | NA | 1,28E-68 | 3,33E-68 | 8 | 47920008 | 94383580 | Amplification |
| c31 | NA | 0,155518252 | -0,420944292 | 1,36E-66 | 3,44E-66 | 6 | 103387583 | 144612154 | Deletion |
| c14 | NA | 0,098523441 | -0,226526794 | 1,49E-65 | 3,70E-65 | 4 | 9404218 | 165426755 | Deletion |
| c64 | NA | 0,19665736 | -0,344252356 | 3,43E-64 | 8,27E-64 | 14 | 22524578 | 45335206 | Deletion |
| c4 | 0,278694417 | -0,082706631 | NA | 2,70E-62 | 6,34E-62 | 1 | 201558793 | 236334938 | Amplification |
| c44 | 0,387707836 | -0,21687471 | NA | 9,43E-62 | 2,16E-61 | 8 | 98460437 | 125079578 | Amplification |
| c58 | NA | 0,082602058 | -0,370836467 | 2,98E-61 | 6,68E-61 | 12 | 77362842 | 132868266 | Deletion |
| c100 | NA | 0,143552356 | -0,408159867 | 8,44E-59 | 1,85E-58 | 21 | 13224688 | 46681423 | Deletion |
| c46 | 0,313088456 | -0,209843495 | NA | 4,94E-58 | 1,05E-57 | 8 | 125641790 | 145074901 | Amplification |
| c68 | 0,469800868 | -0,060893364 | NA | 6,91E-58 | 1,44E-57 | 16 | 34170 | 5291928 | Amplification |
| c91 | 0,41466648 | -0,253480004 | NA | 6,35E-53 | 1,30E-52 | 20 | 16478416 | 28932393 | Amplification |
| c41 | NA | 0,318880132 | -0,36536635 | 2,17E-51 | 4,34E-51 | 8 | 4147499 | 24627540 | Deletion |
| c80 | NA | 0,390993564 | -0,463179535 | 2,26E-49 | 4,43E-49 | 18 | 9373624 | 22390150 | Deletion |
| c24 | 0,352892498 | -0,020002385 | NA | 1,46E-45 | 2,79E-45 | 5 | 89489567 | 157366098 | Amplification |
| c23 | NA | 0,118847565 | -0,305394942 | 5,92E-45 | 1,11E-44 | 5 | 60327558 | 84753632 | Deletion |
| c83 | NA | 0,486821991 | -0,430162331 | 1,41E-44 | 2,59E-44 | 18 | 66590871 | 80256731 | Deletion |
| c15 | NA | 0,235814841 | -0,35653625 | 3,31E-42 | 5,99E-42 | 4 | 74912784 | 86849309 | Deletion |
| c62 | 0,392135752 | -0,150183207 | NA | 5,05E-42 | 8,96E-42 | 13 | 86193668 | 114343518 | Amplification |
| c99 | 0,267707402 | -0,448515429 | NA | 1,03E-41 | 1,79E-41 | 20 | 62894671 | 64283878 | Amplification |
| c45 | 0,347300662 | -0,171012246 | NA | 9,06E-40 | 1,55E-39 | 8 | 114692312 | 139882707 | Amplification |
| c51 | NA | 0,118352518 | -0,175063083 | 7,74E-39 | 1,30E-38 | 10 | 61850051 | 100602358 | Deletion |
| c85 | 0,258921346 | 0,007210972 | NA | 1,91E-37 | 3,15E-37 | 19 | 24320784 | 58533199 | Amplification |
| c13 | NA | 0,157264415 | -0,347688008 | 3,67E-36 | 5,95E-36 | 4 | 72156 | 9155672 | Deletion |
| c82 | NA | 0,465067112 | -0,3469677 | 5,69E-36 | 9,07E-36 | 18 | 54853449 | 63724733 | Deletion |
| c90 | 0,454763733 | -0,284398563 | NA | 1,01E-34 | 1,58E-34 | 20 | 15255441 | 21643019 | Amplification |
| c17 | NA | 0,208714307 | -0,395797962 | 1,14E-34 | 1,75E-34 | 4 | 168555439 | 190035460 | Deletion |
| c27 | NA | 0,068299762 | -0,479484927 | 2,51E-33 | 3,81E-33 | 6 | 149609 | 32591881 | Deletion |
| c2 | NA | 0,177146382 | -0,190144803 | 1,35E-30 | 2,01E-30 | 1 | 97788182 | 115432598 | Deletion |
| c50 | NA | 0,094882486 | -0,133882378 | 1,32E-29 | 1,94E-29 | 10 | 51787782 | 119432039 | Deletion |

| | | | | | | | | | |
|---|---|---|---|---|---|---|---|---|---|
| c78 | 0,333745999 | -0,004382898 | NA | 3,19E-29 | 4,61E-29 | 17 | 54152817 | 83204151 | Amplification |
| c20 | 0,400598726 | -0,064908784 | NA | 1,03E-26 | 1,47E-26 | 5 | 8328588 | 46431572 | Amplification |
| c70 | 0,36559571 | -0,065572858 | NA | 1,19E-25 | 1,67E-25 | 16 | 7187954 | 26081195 | Amplification |
| c49 | NA | 0,114208532 | -0,214108017 | 1,57E-23 | 2,17E-23 | 10 | 48486 | 28549058 | Deletion |
| c88 | NA | 0,128629571 | -0,46978723 | 1,17E-22 | 1,59E-22 | 20 | 2661389 | 14746740 | Deletion |
| c55 | 0,281309909 | -0,036278256 | NA | 2,16E-22 | 2,90E-22 | 12 | 79927 | 18911792 | Amplification |
| c59 | 0,320158119 | -0,168239199 | NA | 3,40E-21 | 4,50E-21 | 13 | 18606106 | 25133455 | Amplification |
| c8 | 0,214979071 | 0,012125331 | NA | 1,45E-17 | 1,89E-17 | 2 | 143745876 | 242109969 | Amplification |
| c21 | 0,357779865 | -0,074053201 | NA | 7,65E-17 | 9,85E-17 | 5 | 29966143 | 46431572 | Amplification |
| c94 | 0,286172479 | -0,426572805 | NA | 2,91E-16 | 3,69E-16 | 20 | 30994015 | 32152631 | Amplification |
| c86 | NA | 0,071987839 | -0,178230025 | 2,02E-15 | 2,54E-15 | 19 | 41636104 | 58533199 | Deletion |
| c10 | NA | 0,03904168 | -0,064364897 | 2,17E-14 | 2,68E-14 | 3 | 8821239 | 198098659 | Deletion |
| c92 | NA | 0,160771841 | -0,479301684 | 3,11E-14 | 3,79E-14 | 20 | 17342388 | 24024025 | Deletion |
| c28 | 0,263982954 | -0,03373831 | NA | 9,75E-14 | 1,18E-13 | 6 | 14805982 | 32683103 | Amplification |
| c29 | 0,165743906 | -0,01981643 | NA | 1,53E-12 | 1,82E-12 | 6 | 82995298 | 149682891 | Amplification |
| c5 | 0,353206883 | -0,124172068 | NA | 2,62E-12 | 3,08E-12 | 1 | 239438247 | 248916508 | Amplification |
| c56 | NA | 0,04895631 | -0,164340586 | 9,43E-11 | 1,09E-10 | 12 | 79927 | 34155198 | Deletion |
| c57 | 0,383152806 | -0,071249115 | NA | 1,70E-09 | 1,95E-09 | 12 | 24629874 | 31859532 | Amplification |
| c9 | NA | 0,055871001 | -0,065001291 | 4,07E-09 | 4,61E-09 | 3 | 12690 | 50136751 | Deletion |
| c52 | NA | 0,089512395 | -0,138344702 | 1,13E-06 | 1,27E-06 | 10 | 122385624 | 133382473 | Deletion |
| c12 | 0,170240307 | -0,013088849 | NA | 1,40E-06 | 1,55E-06 | 3 | 175699585 | 198098659 | Amplification |
| c11 | 0,093765798 | -0,00032202 | NA | 2,77E-05 | 3,03E-05 | 3 | 71123221 | 174528173 | Amplification |
| c71 | 0,170018083 | -0,025816171 | NA | 3,44E-05 | 3,71E-05 | 16 | 46426366 | 65756095 | Amplification |
| c39 | NA | 0,180572352 | -0,364363217 | 6,89E-05 | 7,36E-05 | 8 | 211532 | 4139291 | Deletion |
| c33 | NA | 0,070107969 | -0,174022825 | 0,004783859 | 0,005052615 | 6 | 162759854 | 170606038 | Deletion |
| c7 | 0,077776228 | 0,002090947 | NA | 0,005847852 | 0,006107757 | 2 | 49210512 | 89225084 | Amplification |
| c6 | 0,111358568 | -0,000460505 | NA | 0,032866072 | 0,033949569 | 2 | 4188676 | 19623618 | Amplification |
| c18 | 0,130456948 | -0,08742408 | NA | 0,033681087 | 0,034413285 | 5 | 44922 | 685633 | Amplification |
| c101 | NA | 0,036537436 | -0,026769932 | 0,115943952 | 0,117190661 | 21 | 13224688 | 20585151 | Deletion |
| c26 | 0,035297597 | -0,054413078 | NA | 0,503335303 | 0,503335303 | 6 | 149609 | 2198407 | Amplification |

### Table S13. Noncoding variant enrichment

| Region | region_name | num_mutations | length | effective_length | bg_type | bg_prob | region_pval | WAP | WAP_pval | hotspot_pvals | Fisher_pval | Fisher_FDR | num_samples | position_counts | mutation_counts | AFR samples |
|---|---|---|---|---|---|---|---|---|---|---|---|---|---|---|---|---|
| **chr3:50599047-50602727** | enhancer | 3 | 3681 | 121473 | global | 6.5E-06 | 0.0418 | 2 | 1.7E-05 | 1.1E-09 | 1.11E-09 | 8.38E-07 | 3 | 50602063_1;50602069_1;50602071_1 | 50602063_C_A_1;50602069_C_A_1;50602071_C_A_1 | C-26T;C-49T;C-67T |

| | | | | | | | | | | | | | | | | |
|---|---|---|---|---|---|---|---|---|---|---|---|---|---|---|---|---|
| **chr16:73791294-73794667** | enhancer | 6 | 3374 | 111342 | global | 5E-06 | 1.7E-05 | 0.99 | 0.0209 | NA | 5.66E-06 | 0.00214 | 6 | 73791449_1;73791651_1;73792863_1;73792864_1;73793690_2 | 73791449_G_A_1;73791651_C_A_1;73792863_CCCT_C_1;73792864_CCT_C_1;73793690_A_AT_2 | C-05T;C-20T;C-48T;C-49T;C-59T;C-68T |
| **chr5:140796059-140797518** | CpG_island | 4 | 1460 | 48180 | global | 4.4E-06 | 6.09E-05 | 0.32 | 0.0751 | NA | 6.08E-05 | 0.0153 | 4 | 140796626_1;140796635_1;140796894_1;140797471_1 | 140796626_C_T_1;140796635_C_T_1;140796894_C_T_1;140797471_C_T_1 | C-09T;C-18T;C-45T;C-68T |
| **chr12:63541700-63541806** | non_allelic_homologous | 2 | 107 | 3531 | global | 5.6E-06 | 0.00019 | nan | nan | NA | 0.00019 | 0.0329 | 2 | 63541743_1;63541771_1 | 63541743_C_T_1;63541771_A_G_1 | C-45T;C-67T |
| **chrX:856168-874963** | enhancer | 13 | 18796 | 620268 | global | 4.7E-06 | 5.44E-05 | 2.4E-19 | 0.335 | NA | 0.000217 | 0.0329 | 11 | 857560_1;857911_1;858814_1;858870_2;859584_1;861173_1;862715_1;864291_1;866897_1;867265_1;872113_1;873539_1 | 857560_C_T_1;857911_G_T_1;858814_AAAG_A_1;858870_AT_A_2;859584_ATTCTTTCTTTCTTTTTCTTTCT_A_1;861173_C_T_1;862715_G_A_1;864291_T_TGGGGGGGGGGG_1;866897_G_A_1;867265_G_C_1;872113_C_T_1;873539_TTTAAA_T_1 | C-05T;C-06T;C-09T;C-11T;C-12T;C-16T;C-22T;C-27T;C-48T;C-64T;C-65T |
| **chr16:87808-152854** | locus_control_region | 27 | 65047 | 2146551 | global | 7.2E-06 | 0.852 | 2.9 | 1.3E-05 | 3E-05 | 0.000293 | 0.037 | 10 | 87878_1;96225_1;108662_1;108969_1;109865_1;112635_1;117695_1;118978_1;118980_1;121266_1;123486_1;123784_1;123832_1;129126_1;129661_1;132110_1;132155_1;132156_1;132417_1;132855_1;132991_1;133082_1;133418_1;133420_1;137993_1;144519_1;147105_1 | 108662_TG_T_1;108969_CT_C_1;109865_G_T_1;112635_G_A_1;117695_G_A_1;118978_AG_A_1;118980_C_A_1;121266_C_A_1;123486_G_A_1;123784_A_C_1;123832_AG_A_1;129126_T_A_1;129661_C_A_1;132110_G_A_1;132155_T_G_1;132156_G_A_1;132417_G_C_1;132855_G_C_1;132991_G_C_1;133082_G_C_1;133418_G_A_1;133420_TG_T_1;137993_AT_A_1;144519_G_A_1;147105_AT_A_1;87878_T_A_1;96225_T_C_1 | C-02T;C-08T;C-09T;C-12T;C-22T;C-26T;C-27T;C-47T;C-58T;C-65T |

| | | | | | | | | | | | | | | | |
|---|---|---|---|---|---|---|---|---|---|---|---|---|---|---|---|
| **chr5:25077654-25083685** | non-LTR_retrotransposon:L1PA3 | 7 | 6032 | 199056 | global | 5.3E-06 | 6.41E-05 | 2.9E-117 | 0.657 | NA | 0.000466 | 0.0504 | 7 | 25079406_1;25080005_1;25080144_1;25080546_1;25081829_1;25082053_1;25083368_1 | 25079406_C_A_1;25080005_G_A_1;25080144_T_G_1;25080546_CT_C_1;25081829_C_A_1;25082053_T_A_1;25083368_G_A_1 | C-07T;C-11T;C-16T;C-18T;C-22T;C-26T;C-67T |

| Region | region_name | num_mutations | length | effective_length | bg_type | bg_prob | region_pval | WAP | WAP_pval | hotspot_pvals | Fisher_pval | Fisher_FDR | num_samples | position_counts | mutation_counts | EUR samples |
|---|---|---|---|---|---|---|---|---|---|---|---|---|---|---|---|---|
| **chr5:140842622-140843862** | CpG_island | 2 | 1241 | 7446 | global | 4.9E-06 | 0.000535 | nan | nan | NA | 0.000535 | 0.0451 | 2 | 140842770_1;140842965_1 | 140842770_C_T_1;140842965_C_T_1 | C-44T;C-52T |
| **chr21:24715428-24717804** | meiotic | 3 | 2377 | 14262 | global | 1.1E-05 | 0.000322 | 2.8E-30 | 0.17 | NA | 0.000591 | 0.0451 | 3 | 24715863_1;24716459_1;24716529_1 | 24715863_G_GAA_1;24716459_C_T_1;24716529_T_C_1 | C-36T;C-37T;C-52T |
| **chr21:24715300-24717804** | meiotic | 3 | 2505 | 15030 | global | 1.1E-05 | 0.000375 | 2.8E-30 | 0.153 | NA | 0.000619 | 0.0451 | 3 | 24715863_1;24716459_1;24716529_1 | 24715863_G_GAA_1;24716459_C_T_1;24716529_T_C_1 | C-36T;C-37T;C-52T |
| **chr2:89027458-89027466** | IGKV3-11_leader_sequence | 1 | 9 | 54 | global | 1.5E-05 | 0.000803 | nan | nan | NA | 0.000803 | 0.0451 | 1 | 89027458_1 | 89027458_C_T_1 | C-37T |
| **chr9:136252418-136252430** | nucleotide_motif | 1 | 13 | 78 | global | 1.4E-05 | 0.00106 | nan | nan | NA | 0.00106 | 0.0451 | 1 | 136252424_1 | 136252424_T_G_1 | C-36T |
| **chr1:119507314-119507337** | enhancer | 1 | 24 | 144 | global | 1E-05 | 0.00146 | nan | nan | NA | 0.00146 | 0.0451 | 1 | 119507323_1 | 119507323_CA_C_1 | C-33T |
