## Supplemental Notes for "Integrative Genomic, Transcriptomic, and Microbiome Profiles of Colon Cancer by Ancestry Provide Insights into Molecular Distinctions"

**Note S1. Additional P-1000 cohort characteristics for Table 1.**

The median age for the cohort was 68, though age did vary across genetic ancestries. The median age was 67 for African ancestry patients compared to a median of 79 for European ancestry patients (Table 1. Wilcoxon rank sum test p-value = 0.002). Similarly, tumor histology (Table 1. Fisher's exact test p-value = <0.001) and tumor grade (Table 1. Fisher's exact test p-value = 0.025) differed across genetic ancestries. European ancestry patients had higher-grade tumors and a higher proportion of mucinous adenocarcinoma histologies. The majority of AFRg tumors were either stage II (52%) or stage III (31%). For this study, aside from the samples described in the main text, four DNA sequencing samples (tumor-non-normal pairs) were excluded for tumor QC issues, indicating possible normal overgrowth, and an additional 4 samples (tumor-non-normal pairs) were excluded due to characterization as rectal cancer. These data will also be made available as a public resource for further study.

**Note S2. MSI status determination**

MSI status was determined both by pathology, using immunohistochemistry (IHC) staining, and by genomic metrics including germline and somatic MMR gene mutation assessment, tumor mutation burden, and, for the P-1000 cohort, microsatellite repeat length comparison between tumor and non-tumor using MSIsensor(Niu *et al.*, 2014) and MANTIS(Kautto *et al.*, 2017). There are cases in which microsatellite stability determination was made in light of conflicting or heterogeneous information across different sources, and in these cases, genomic evidence was prioritized.  Details of each metric and samples with conflicts are noted (see Table S12).

**Note S3. Similarity coefficients between copy-number profiles**

Similarity between the profiles of the P-1000 patients (AFRg and EURg patients) was assessed using the Szymkiewicz–Simpson coefficient on the copy-number amplifications and deletions separately (see Figures S14A-B). For both amplification and deletion analyses, the clustering generated a stratification by CMS subtyping rather than by ancestry. For the amplifications, the profiles are not randomly distributed between the two main clusters when examining CMS subtyping (only subtypes CMS1 to 4 are retained, Fisher test p-value = 0.0002648)  (see Figure S14A). The CMS1 and CMS4 subtypes are respectively enriched in cluster 2 and cluster 1 (one CMS subtype vs. the sum of the other subtypes, Fisher test p-values of 0.0001351 and 0.03549, respectively). For the deletions, the profiles are also not randomly distributed between the two main clusters when examining CMS subtyping (only CMS1 to 4 subtypes retained, Fisher test p-value = 0.02104) (see Figure S14B). This time, only the CMS1 subtyping is enriched in cluster 1 (CMS1 vs. the sum of the other subtypes, Fisher test p-value = 0.01018). Additionally, all profiles except one from cluster 2 in the amplification analysis are found in cluster 1 of the deletion analysis (see Figure S4C). Both of those clusters are CMS1 subtype-enriched.

### Note S4. Human Leukocyte Antigen (HLA) loss of heterozygosity (LOH)

We have also investigated the HLA loss of heterozygosity (LOH) in the tumor by comparing the typing between the WGS tumor and the matching adjacent non-tumor samples. The HLA LOH reduces the diversity of HLA alleles available to present antigens and may facilitate immune evasion by limiting the recognition of tumor neoantigens that lead to an immune response (McGranahan *et al.*, 2017),(McGranahan *et al.*, 2017). In the P-1000 cohort, only a limited number of patients show HLA LOH (see Figure S15A). Additionally, no specific pattern associated with cancer subtyping could be discerned for the P-1000 AFRg cohort. The prevalence of HLA class I haplotype loss in the P-1000 cohort was lower than the 40% CRC patients identified in the cohort by Maleno et al.(Maleno *et al.*, 2004) and the 15% CRC patients in Montesion et al.(Montesion *et al.*, 2021). However, in a different cohort, Sewastianik et al. (2025)(Sewastianik *et al.*, 2025) found that 90% of cancer cases, including those with CRC and lung adenocarcinoma, are genomically intact at the HLA class I locus.

### Note S5. Human Leukocyte Antigen (HLA) allele diversity in the P-1000 AFRg cohort

We have investigated the HLA allele diversity in the P-1000 AFRg cohort. For this purpose, both HLA class I and class II genes have been typed using the WGS adjacent non-tumor samples. We identified the most frequent alleles for three HLA class I (A, B, and C) and two class II (DQB1 and DRB1) genes in the P-1000 AFRg cohort. We then compared their ranking with the National Marrow Donor Program (NMDP) registry haplotype frequencies for African Americans(Gragert *et al.*, 2013) (see Figure S15B-F). For all compared genes, the P-1000 AFRg HLA allele diversity recapitulates the diversity seen in the African American population. More specifically, for all analyzed genes, the three most frequent alleles from the NMDP registry consistently ranked in the top 5 in the P-1000 AFRg cohort. No rare alleles (NMDP ranking > 45) were seen in the P-1000 AFRg cohort. Those results may not be generalizable due to the limitations of the sample size.

### Note S6. Standard methods from NYGC

This process is completed by first genotyping the markers from 1,964 unrelated 1KG individuals with GATK pileup(McKenna *et al.*, 2010) (RRID:SCR_001876). Due to known high admixture, individuals assigned to Mexican Ancestry from Los Angeles USA (MXL), African Caribbean in Barbados (ACB), and African Ancestry in Southwest US (ASW) populations were excluded. At this step, only the SNP markers with a minimum minor allele frequency (MAF) of 0.01 overall and 0.05 in at least one 1KG superpopulation were retained. The final SNP markers dataset, for the reference population set, was generated by linkage disequilibrium (LD) pruning using PLINK(Alexander, Novembre and Lange, 2009) (RRID:SCR_001757) v1.9 with a window size of 500kb, a step size of 250kb and a ‘r2’ threshold of 0.2. The proportional breakdown of each P-1000 sample into 5 continental and 23 subcontinental populations were obtained.

Gragert, L. *et al.* (2013) ‘Six-locus high resolution HLA haplotype frequencies derived from mixed-resolution DNA typing for the entire US donor registry’, *Human immunology*, 74(10), pp.

1313–1320.

Kautto, E.A. *et al.* (2017) ‘Performance evaluation for rapid detection of pan-cancer microsatellite instability with MANTIS’, *Oncotarget*, 8(5), pp. 7452–7463.

Maleno, I. *et al.* (2004) ‘Distribution of HLA class I altered phenotypes in colorectal carcinomas: high frequency of HLA haplotype loss associated with loss of heterozygosity in chromosome region 6p21’, *Immunogenetics*, 56(4), pp. 244–253.

McGranahan, N. *et al.* (2017) ‘Allele-Specific HLA Loss and Immune Escape in Lung Cancer Evolution’, *Cell*, 171(6), pp. 1259–1271.e11.

Montesion, M. *et al.* (2021) ‘Somatic HLA Class I Loss Is a Widespread Mechanism of Immune Evasion Which Refines the Use of Tumor Mutational Burden as a Biomarker of Checkpoint Inhibitor Response’, *Cancer discovery*, 11(2), pp. 282–292.

Niu, B. *et al.* (2014) ‘MSIsensor: microsatellite instability detection using paired tumor-normal sequence data’, *Bioinformatics (Oxford, England)*, 30(7), pp. 1015–1016.

Sewastianik, T. *et al.* (2025) ‘Abstract 3745: Allele-specific HLA LOH frequencies and survival outcomes in cancer: a real-world analysis’, *Cancer research*, 85(8_Supplement_1), pp. 3745–3745.
